# Multi-ancestry genome-wide association study improves resolution of genes, pathways and pleiotropy for lung function and chronic obstructive pulmonary disease

**DOI:** 10.1101/2022.05.11.22274314

**Authors:** Nick Shrine, Abril G Izquierdo, Jing Chen, Richard Packer, Robert J Hall, Anna L Guyatt, Chiara Batini, Rebecca J Thompson, Chandan Pavuluri, Vidhi Malik, Brian D Hobbs, Matthew Moll, Wonji Kim, Ruth Tal-Singer, Per Bakke, Katherine A Fawcett, Catherine John, Kayesha Coley, Noemi Nicole Piga, Alfred Pozarickij, Kuang Lin, Iona Y Millwood, Zhengming Chen, Liming Li, Sara RA Wielscher, Lies Lahousse, Guy Brusselle, Andre G Uitterlinden, Ani Manichaikul, Elizabeth C Oelsner, Stephen S Rich, R. Graham Barr, Shona M Kerr, Veronique Vitart, Michael R Brown, Matthias Wielscher, Medea Imboden, Ayoung Jeong, Traci M Bartz, Sina A Gharib, Claudia Flexeder, Stefan Karrasch, Christian Gieger, Annette Peters, Beate Stubbe, Xiaowei Hu, Victor E Ortega, Deborah A Meyers, Eugene R Bleecker, Stacey B Gabriel, Namrata Gupta, Albert Vernon Smith, Jian’an Luan, Jing-Hua Zhao, Ailin F Hansen, Arnulf Langhammer, Cristen Willer, Laxmi Bhatta, David Porteous, Blair H Smith, Archie Campbell, Tamar Sofer, Jiwon Lee, Martha L Daviglus, Bing Yu, Elise Lim, Hanfei Xu, George T O’Connor, Gaurav Thareja, Omar M E Albagha, Hamdi Mbarek, Karsten Suhre, Raquel Granell, Tariq O Faquih, Pieter S Hiemstra, Annelies M Slats, Benjamin H Mullin, Jennie Hui, Alan James, John Beilby, Karina Patasova, Pirro Hysi, Jukka T Koskela, Annah B Wyss, Jianping Jin, Sinjini Sikdar, Mikyeong Lee, Sebastian May-Wilson, Nicola Pirastu, Katherine A Kentistou, Peter K Joshi, Paul RHJ Timmers, Alexander T Williams, Robert C Free, Xueyang Wang, John L Morrison, Frank D Gilliland, Zhanghua Chen, Carol A Wang, Rachel E Foong, Sarah E Harris, Adele Taylor, Paul Redmond, James P Cook, Anubha Mahajan, Lars Lind, Teemu Palviainen, Terho Lehtimäki, Olli T Raitakari, Jaakko Kaprio, Taina Rantanen, Kirsi H Pietiläinen, Simon R Cox, Craig E Pennell, Graham L Hall, W. James Gauderman, Chris Brightling, James F Wilson, Tuula Vasankari, Tarja Laitinen, Veikko Salomaa, Dennis O Mook-Kanamori, Nicholas J Timpson, Eleftheria Zeggini, Josée Dupuis, Caroline Hayward, Ben Brumpton, Claudia Langenberg, Stefan Weiss, Georg Homuth, Carsten Oliver Schmidt, Nicole Probst-Hensch, Marjo-Riitta Jarvelin, Alanna C Morrison, Ozren Polasek, Igor Rudan, Joo-Hyeon Lee, Ian Sayers, Emma L Rawlins, Frank Dudbridge, Edwin K Silverman, David P Strachan, Robin G Walters, Andrew P Morris, Stephanie J London, Michael H Cho, Louise V Wain, Ian P Hall, Martin D Tobin

## Abstract

Lung function impairment underlies chronic obstructive pulmonary disease (COPD) and predicts mortality. In the largest multi-ancestry GWAS meta-analysis of lung function to date, comprising 580,869 participants, 1020 independent association signals identified 559 genes supported by ≥2 criteria from a systematic variant-to-gene mapping framework. These genes were enriched in 29 pathways. Individual variants showed heterogeneity across ancestries, age and smoking groups, and collectively as a genetic risk score (GRS) showed strong association with COPD across ancestry groups. We undertook phenome-wide association studies (PheWAS) for selected associated variants, and trait and pathway-specific GRS to infer possible consequences of intervening in pathways underlying lung function. We highlight new putative causal variants, genes, proteins and pathways, including those targeted by existing drugs. These findings bring us closer to understanding the mechanisms underlying lung function and COPD, and should inform functional genomics experiments and potentially future COPD therapies.

## Introduction

Lung function - even within the normal range - predicts mortality and is a key diagnostic criterion for COPD^1^, which has the highest prevalence of respiratory diseases globally^2^ and lacks disease-modifying treatments. Whilst smoking and environmental risk factors for COPD are well known, and genetic susceptibility (heritability) is recognised, the molecular pathways underlying COPD are incompletely understood. In common with many other complex traits, there has been under-representation of diverse ancestries in genome-wide association studies (GWAS)^3^ of lung function^4-6^. Multi-ancestry studies improve the power and fine-mapping resolution of GWAS, and ultimately the prospects for prediction, prevention, diagnosis and treatment in diverse populations^3,4,7^.

Understanding of genes, proteins and pathways involved in diseases and disease-related traits underpins modern drug development. A high yield of genetic association signals, improved signal resolution and integration with functional evidence are all required to confidently identify causal genes and the variants and pathways that impact gene function and regulation. Although datasets and *in-silico* tools to connect GWAS signals to causal genes are improving, the findings from different datasets and tools have lacked consensus^8,9^, highlighting a need for frameworks to integrate functional evidence types and to compare findings^10^.

Aggregation of genetic variants associated with lung function into a genetic risk score (GRS) provides a tool for COPD prediction^5^. When a GRS comprises a sufficient number of variants, partitioning the GRS according to the biological pathways the variants influence could provide a tool to explore their aggregated consequences across a wide range of traits through phenome-wide association studies (PheWAS). Just as PheWAS of individual genetic variants can predict the consequences of perturbation of specific protein targets, informing assessment of drug efficacy, drug safety and drug repurposing opportunities^11^, PheWAS of pathway-partitioned GRS could inform the understanding of consequences of perturbing specific pathways.

Through the largest global assembly of lung function genomics studies to date we: (i) undertook a multi-ancestry meta-analysis of GWAS of lung function traits in 580,869 individuals to detect novel signals, improve fine-mapping and estimate the extent of heterogeneity in allelic effects attributable to ancestry; (ii) tested whether lung function signals were age-dependent or smoking-dependent, and assessed their relationship to height; (iii) investigated cell type and functional specificity of lung function association signals; (iv) fine-mapped signals through annotation-informed credible sets, integrating functional data such as respiratory cell-specific chromatin accessibility signatures; (v) applied a consensus-based framework to systematically investigate and identify putative causal genes, integrating eight locus-based or similarity-based criteria; (vi) developed and applied a GRS for the ratio of forced expiratory volume in 1 second to forced vital capacity (FEV_1_/FVC) in different ancestries in UK Biobank and in COPD case-control studies; (vii) applied PheWAS to individual variants, GRS for each lung function trait, and GRS partitioned by pathway. Through these approaches we aimed to detect novel lung function signals and novel putative causal genes, and provide new insights into the mechanistic pathways underlying lung function, some of which may be amenable to drug therapy.

## Results

We undertook genome-wide association analyses of forced expired volume in 1 second (FEV_1_), forced vital capacity (FVC), FEV_1_/FVC, and peak expiratory flow rate (PEF) from 49 cohorts (**Methods, Supplementary Table 1, Supplementary Table 2**). Our sample of up to 580,869 participants comprised individuals of African (AFR: N=8,590), American/Hispanic (AMR: N=14,668), East Asian (EAS: N=85,279), South Asian (SAS: N=10,093) and European ancestry (EUR: N=462,239, **Supplementary Figure 1a, 1b**). Adjustments were made for age, age^2^, sex, and height in association testing within cohorts, and we accounted for population structure and, where appropriate, relatedness (**Methods** and **Supplementary Tables 2-3**). Genomic control was applied to each cohort before meta-analysis using the linkage disequilibrium (LD) score intercept^12^. After filtering and meta-analysis across multi-ancestry cohorts, 66.8M variants were available genome-wide for signal selection in each of 4 lung function traits, with genomic inflation factors λ of 1.025, 1.022, 0.984 and 0.996 for FEV_1_, FVC, FEV_1_/FVC and PEF respectively (**Supplementary Figures 2-3**).

### 1020 signals for lung function

After excluding 8 signals associated with smoking behaviour (**Supplementary Note**), and combining signals that colocalised across multiple traits, we identified 1020 distinct signals for lung function using a stringent threshold of P<5×10^−9^ (ref.^13^, Figure 1a). Of these, 713 are novel with respect to the signals and studies described in the **Supplementary Note**. These 1020 signals show a pattern of increasing effect size as allele frequency decreases, in keeping with other complex traits^14^ (**Supplementary Figure 6**), and explain 33.0% of FEV_1_/FVC heritability (21.3% for FEV_1_, 17.3% for FVC, 21.4% for PEF, **Methods**).

**Figure 1:**
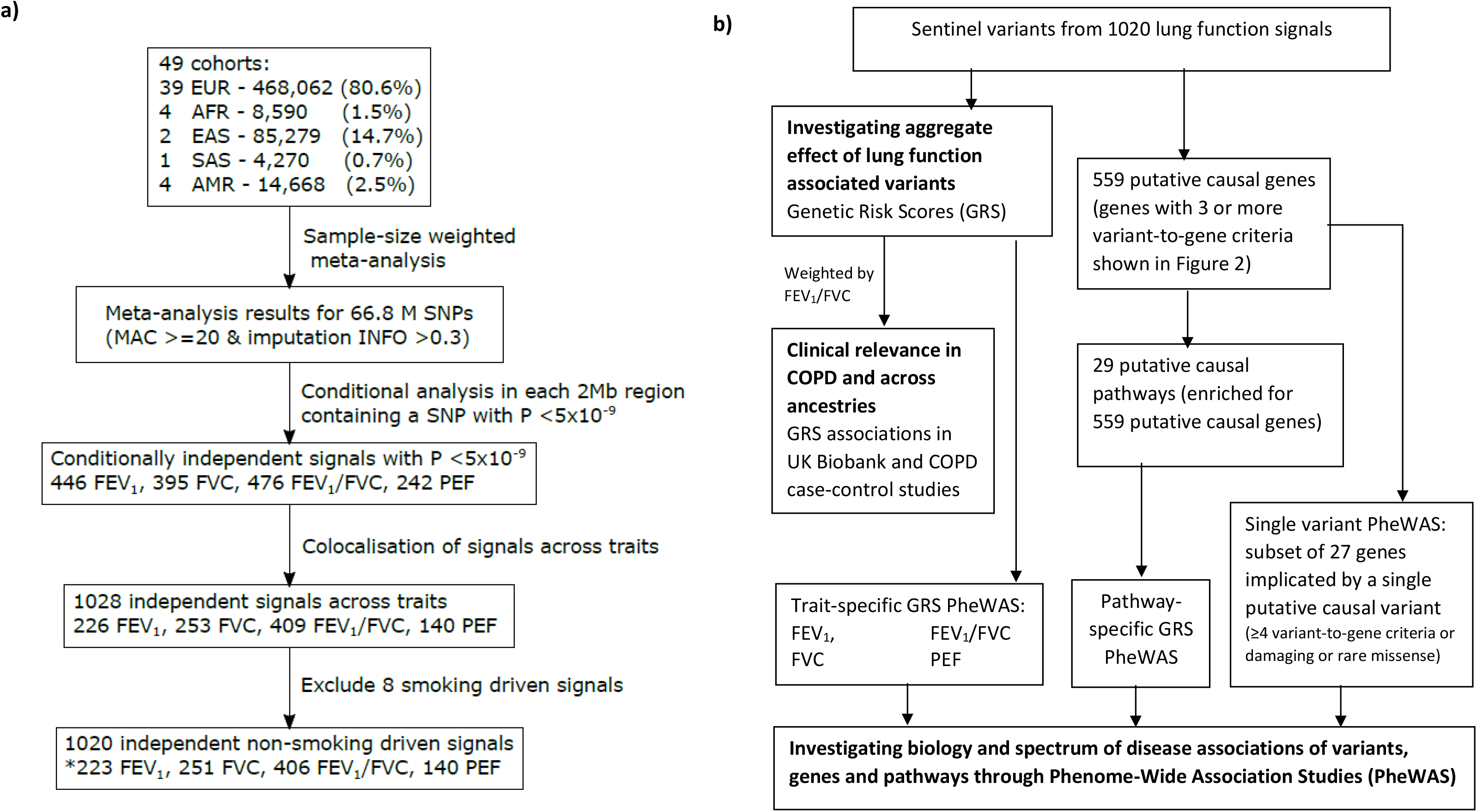
Overview of (a) Discovery meta-analysis; Ancestry abbreviations: EUR – European, AFR – African, EAS – East Asian, SAS – South Asian, AMR – Admixed American/Hispanic. *For signals present in more than one trait, the signal is only counted once (for the most significant trait); **(b) pathway analyses, genetic risk score (GRS) analyses and phenome-wide association studies (PheWAS)**

To facilitate fine-mapping, we included larger and more diverse populations than previous lung function GWAS. We performed multi-ancestry meta-regression with MR-MEGA^7^, which incorporates axes of genetic ancestry as covariates to model heterogeneity (**Methods**). We then incorporated functional annotation for chromatin accessibility and transcription factor binding sites in respiratory-relevant cells and tissues, and enriched genomic annotations^15^ to weight prior causal probabilities of association for putative causal variants (**Methods**). Overall reductions in credible set size and higher maximum posterior probabilities of association for the most likely causal variant in the credible set were evident after multi-ancestry meta-regression was employed and after functional annotation was incorporated (**Supplementary Figure 7**). Following fine-mapping, 438 (43%) signals had a single putative causal variant (posterior probability >50%) and the median credible set size was 9 variants (**Supplementary Note**).

We assessed heterogeneity of variant associations attributable to ancestry utilising MR-MEGA. Of the 960 signals represented in ≥7 cohorts, 109 signals showed ancestry-correlated heterogeneity (P_Het_<0.05, Figure 4 **Supplementary Table 8**), more than expected by chance (binominal test, P = 3.93×10^−15^). Among these, five signals (rs9393688, rs28574670 (*LTBP4*), rs7183859 (*THSD4*), rs59985551 (*EFEMP1*), rs78101726 (*MECOM*)) showed significant ancestry-correlated heterogeneity (Bonferroni correction for 960 signals tested, P_Het_< 5.21×10^−5^, **Supplementary Figures 5(a-e)**). The intronic variant rs7183859 in *THSD4*, which we previously implicated in lung function^16^, showed larger effect size estimates in non-EUR ancestries, and in particular African ancestries (**P**_**HET**_**=**3.33×10^−5^, **Supplementary Figure 5c**).

We tested for differences in the estimated effect sizes of our 1,020 signals between children and adults, as well as between ever-smokers and never-smokers, in European individuals (**Methods**; Bonferroni P thresholds 5.14×10^−5^ and 4.9×10^−5^ for age-dependent and smoking-dependent effect analyses respectively). Of the 972 signals compared between children and adults, effect size estimates were correlated (r from 0.46 for FEV_1_/FVC to 0.62 for FEV_1_), although 136 signals showed nominal evidence (P <0.05) of age-dependent effects (more than expected by chance, binomial P = 1.14×10^−26^). Four signals showed age-dependent effects (P <5.14×10^−5^): rs7977418 (*CCDC91*), rs11722554 (*CYTL1*), rs6806825 (*MECOM*) and rs11079718 (*MAPT*) (**Supplementary Table 9, Supplementary Figure 9**). We observed 69 out of 1020 signals with nominal evidence (P <0.05) of smoking-dependent effects, more than expected by chance (binomial P= 0.0079). The intronic SNP rs7733410 in *HTR4*, a signal we previously reported for lung function^16^, showed a 76.2% larger effect on FEV_1_ in ever than never-smokers (P=4.09×10^−5^, **Supplementary Table 10**). As height is a determinant of lung growth, we compared height and lung function associations and tested the impact of additional height adjustments for sentinel SNPs. We found no correlation between estimated effect sizes for height and lung function of the 1020 sentinels (**Supplementary Figure 10**), and the addition of height^2^ and height^3^ covariates had little impact on effect size estimates (**Supplementary Figure 11**).

### Cell-type and functional specificity

We assessed whether our association signals were enriched for regulatory or functional features in specific cell types. Using stratified LD-score regression^17^ we found enrichment of all histone marks we tested (H3K27ac, H3K9ac, H3K4me3, H3K4me1) in lung and smooth muscle containing cell lines (**Supplementary Table 17)**. Using GARFIELD^18^ we assessed enrichment of our signals for DNasel hypersensitivity sites (DHS) and chromatin accessibility peaks, showing enrichment in a wide variety of cell types, including higher enrichment in foetal and adult lung and blood for FEV_1_, FEV_1_/FVC, and PEF and fibroblast enrichment for FVC (**Supplementary Figure 8a**). Our signals were enriched for transcription factor footprints in foetal lung for FEV_1_, FEV_1_/FVC, and PEF, for footprints in skin for FVC, and also in blood for PEF (**Supplementary Figure 8b**). Genic annotation enrichment patterns were similar across all traits, with enrichment mainly in exonic, 3’ UTR and 5’ UTR regions (**Supplementary Figure 8c**). For all traits we saw enrichment for transcription start sites (TSS), weak enhancers, enhancers and promoter flanks, with cell types for weak enhancer enrichment including endothelial cells for FEV_1_, FEV_1_/FVC, and PEF (**Supplementary Figure 8d**). For transcription factor binding sites, we observed a similar enrichment pattern across all the lung function traits with the largest fold-enrichment in endothelial cells (**Supplementary Figure 8e**). We used ATAC-seq data for the above fine-mapping and also to describe enrichment of our signals in specific cell types. Our signals were enriched in ATAC-seq peaks (**Supplementary Note**) in matrix fibroblast 1 for FVC, matrix fibroblast 2 for FEV_1_, myofibroblast for FEV_1_, FEV_1_/FVC, and PEF, and alveolar type 1 cells in FEV_1_/FVC and genic annotations showed enrichment of exon variants for FEV_1_, FEV_1_/FVC, and 3’ UTR variants for FEV_1_ and FVC. We also found enrichment of transcription factor binding sites in lung across all phenotypes and in bronchus for FEV_1_/FVC (**Supplementary Table 13**).

### Identification of putative causal genes and causal variants

To systematically investigate and identify putative causal genes, we integrated orthogonal evidence, using eight locus-based or similarity-based criteria (**Supplementary Note**): (i) the nearest gene to the sentinel SNP; (ii) colocalisation of GWAS signal and eQTL or (iii) pQTL signals in relevant tissues (**Methods)**; (iv) rare variant association in whole exome sequencing in UK Biobank; (v) proximity to a gene for a Mendelian disease with a respiratory phenotype (+/-500kb); (vi) proximity to a human ortholog of a mouse knockout gene with a respiratory phenotype (+/-500kb); (vii) an annotation-informed credible set^15^ containing a missense/deleterious/damaging variant with a posterior probability of association >50% and; (viii) the gene with the highest polygenic priority score (PoPS), a method based on the assumption that causal genes on different chromosomes share similar functional characteristics^9^. We identified 559 putative causal genes satisfying at least two criteria, of which 135 were supported by at least three criteria (Figure 1b, Figure 2, Figure 3). Among 20 genes supported by 4 or more criteria (**Supplementary Table 14**), six previously implicated genes (*TGFB2, NPNT, LTBP4, TNS1, SMAD3, AP3B1*) ^5,16,19-21^ were supported by additional criteria compared with the original reports. Fourteen of the 20 genes supported by 4 or more criteria have not been confidently implicated in lung function previously (*CYTL1, HMCN1, GATA5, ADAMTS10, IGHMBP2, SCMH1, GLI3, ABCA3, TIM1, CFH, FGFR1, LRBA, CLDN18, IGF2BP2*). These are involved in smooth muscle function (*FGFR1, GATA5, STIM1*), tissue organisation (*ADAMTS10*), alveolar and epithelial function (*ABCA3, CLDN18*), and inflammation and immune response to infection (*CFH, CYTL1, HMCN1, LRBA, STIM1*).

**Figure 2:**
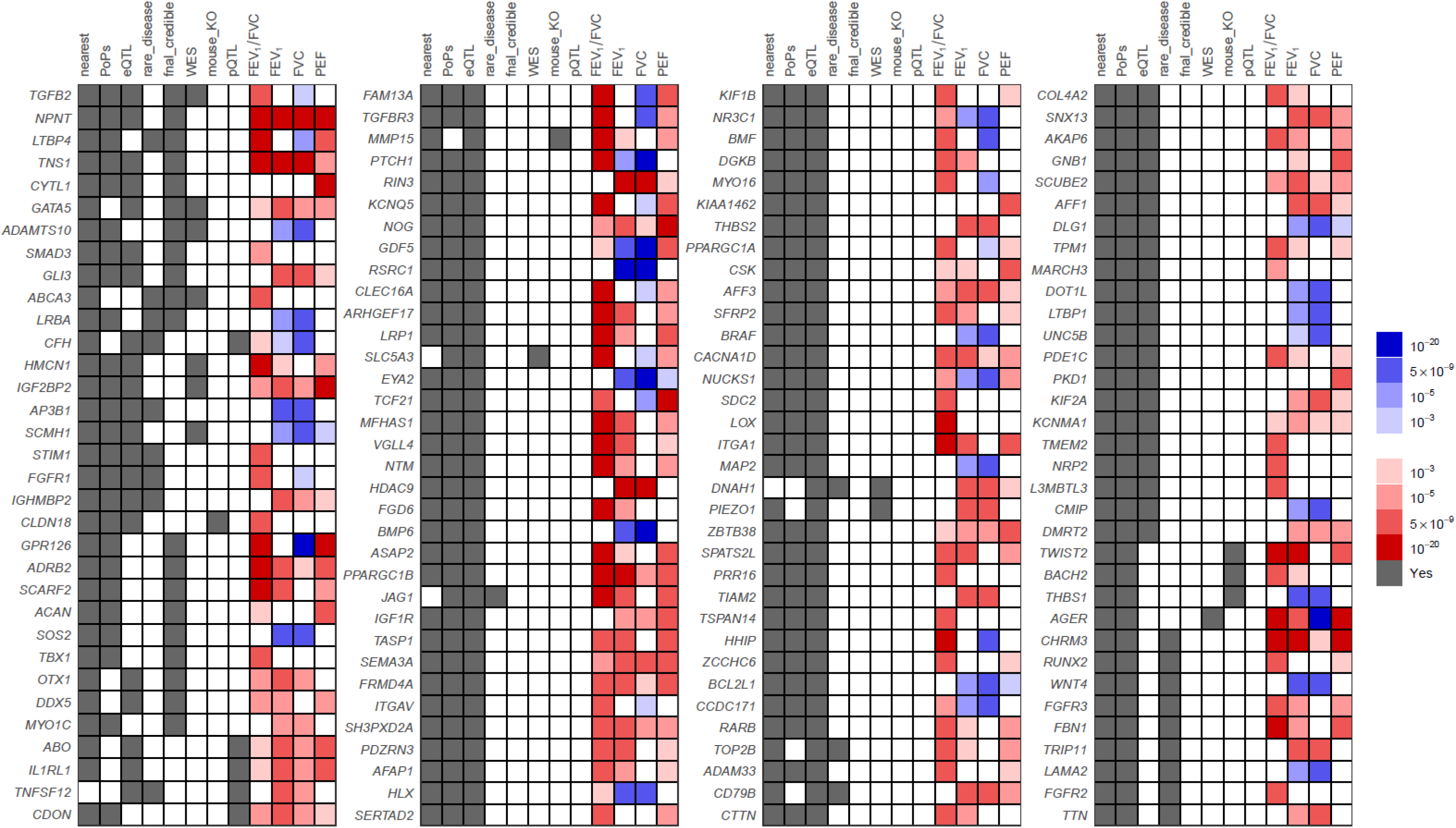
135 genes prioritised with 3 or more variant-to-gene criteria. The grey in the first 8 columns indicates that at least 1 variant implicates the gene as causal via the evidence for that column. The last 4 columns indicates the level of association of the most significant variant implicating the gene as causal with respect to the FEV_1_/FVC decreasing allele: the same direction of effect as the FEV_1_/FVC decreasing allele has red shades, the opposite direction has blue shades.

**Figure 3:**
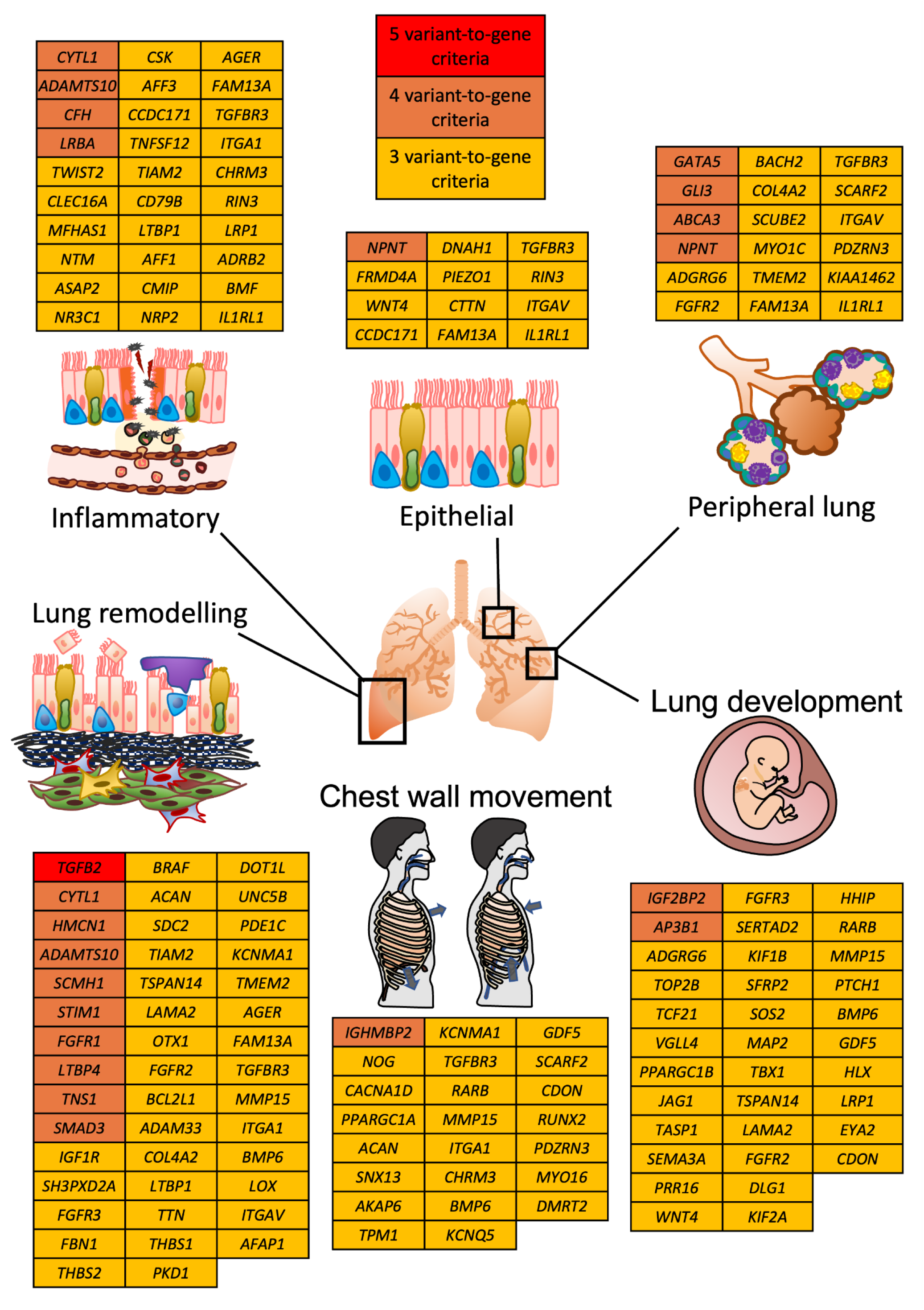
Summary of lung function biology. Genes implicated by 3 or more variant-to-gene criteria are displayed on the basis of their putative lung function role (**Methods**). A summary of all prioritised genes is shown in **Supplementary Table 14** (Prioritised genes).

In order to supplement understanding of the biological pathways and range of clinical phenotypes that lung function associated variants influence, we undertook PheWAS of selected individual variants. We selected 27 putative causal genes implicated by ≥4 criteria (20 genes), or implicated by a single putative causal missense variant that was deleterious (5 genes: *ACAN, ADGRG6, SCARF2, CACNA1S, HIST1H2BE*) or rare (2 genes: *SOS2, ADRB2*, **Supplementary Table 15**). We interpreted the PheWAS findings (shown in full in **Supplementary Figure 14** and **Supplementary Table 23**) alongside literature reviews (**Supplementary Table 32)** for each of these 27 genes; examples are highlighted in the three paragraphs below.

The putative causal deleterious missense variant in *ABCA3* associated with reduced FEV_1_/FVC, rs149989682 (A allele, frequency 0.6%), has been reported to cause paediatric interstitial lung disease^22^. ABCA3, expressed in alveolar type II cells and localised to lamellar bodies, is involved in surfactant phospholipid metabolism and several *ABCA3* mutations cause severe neonatal surfactant deficiency^23^. The putative causal deleterious missense variant rs200383755 C allele (frequency 0.6%) in *GATA5*, associated with lower FEV_1_, was associated with increased risk of asthma, higher blood pressure and reduced risk of benign prostatic hyperplasia in our PheWAS. *GATA5* associations have not been previously noted in GWAS of asthma, although Gata5-deficient mice show airway hyperresponsiveness^24^ (**Supplementary Figure 14j**). GATA5 is a transcription factor involved in smooth muscle cell diversity, expressed in bronchial smooth muscle, and highly expressed in bladder and prostate; a previous benign prostatic hyperplasia GWAS reported a *GATA5* signal ^25 24^. *CLDN18* was implicated by 4 criteria, including a mouse knockout with abnormal pulmonary alveolar epithelium morphology^26^. Through calcium-independent cell-adhesion, *CLDN18* influences epithelial barrier function through tight junction-specific obliteration of the intercellular space^27^, and its splice variant CLDN18.1 is predominantly expressed in the lung^28^. Reduced *CLDN18* expression has been reported in asthma^29^. However, our PheWAS showed no association with asthma susceptibility or other traits (CLDN18_rs182770 in **Supplementary Table 23**). *LRBA* was also implicated by 4 criteria. Mutations resulting in LRBA deficiency cause common variable immunodeficiency-8 with autoimmunity, which can include cough, respiratory infections, bronchiectasis, or interstitial lung disease ^30 31^. Putative causal *LRBA* variant rs2290846 (posterior probability 56.3%) is a tolerated missense variant which showed pleiotropic associations with 31 associated traits (FDR<1%) in our PheWAS (**Supplementary Figure 14o, Supplementary Table 23**). The rs2290846 G allele, associated with lower FVC and lower FEV_1_, was associated with lower neutrophils, lower risks of cholelithiasis and cholecystitis^32^, and lower diverticular disease risk.

*FGFR1*, encoding Fibroblast Growth Factor Receptor 1, has roles in lung development and regeneration^33^, and loss-of-function *FGFR1* mutations cause hypogonadotropic hypogonadism^34^. Notably, in our PheWAS, the T allele of rs881299, associated with lower FEV_1_/FVC and higher FVC, is strongly associated with higher testosterone (particularly in males) and higher sex hormone binding globulin (SHBG), lower BMI, lower alanine transaminase and urate levels (**Supplementary Figure 14z, Supplementary Table 23**). Missense variant rs72681869 in *SOS2* also showed association with SHBG in our PheWAS. In both sexes the C allele of rs72681869, associated with higher FVC and higher FEV_1_, was associated with lower SHBG, higher alanine aminotransferase (ALT) and aspartate aminotransferase (AST), higher fat mass, HbA1c and higher systolic and diastolic blood pressure, higher urate and creatinine, and in males lower testosterone, and reduced inguinal hernia risk (**Supplementary Figure 14ac-ae)**. Mutations in *SOS2* have been reported in Noonan Syndrome. The A allele of rs7514261 implicating *CFH*, associated with lower FVC, was strongly associated with reduced risk of macular degeneration^35^ and also with raised albumin in our PheWAS (**Supplementary Figure 14h**).

*CACNA1S* is one of several genes prioritised encoding calcium voltage-gated channel subunits in skeletal muscle (*CACNA1S, CACNA1D*, and *CACNA2D3* supported by ≥2 criteria; *CACNA1C* was supported by PoPS). Mutations in *CACNA1S* have been reported to cause hypokalemic periodic paralysis^36^ and malignant hyperthermia^37^. *CACNA1S* is strongly expressed in skeletal muscle, but at much lower levels in airway smooth muscle. The common *CACNA1S* missense variant, rs3850625 (A allele, frequency EUR 11.8%, SAS 21.4%) was associated with lower FVC, lower FEV_1_, and in the PheWAS, with lower whole body fat-free mass, reduced hand grip strength, and lower aspartate aminotransferase and creatinine levels (**Supplementary Figure 14f**). CACNA1S *and* CACNA1D are targeted by dihydropyridine calcium channel blockers, which have been reported to produce small improvements in lung function in asthma^38^. The low frequency missense variant rs1800888 in *ADRB2* (T, 1.49% EUR), associated with lower FEV_1_ and lower FEV_1_/FVC, showed strongest association in the PheWAS with increased eosinophil count.

### Druggable targets

Using the Drug Gene Interaction Database (DGIDB), we surveyed 559 genes supported by ≥2 criteria. We found 292 drugs indicated by ChEMBL interactions mapping to 55 genes (**Supplementary Table 16**), including *ITGA2*, encoding Integrin Subunit Alpha 2. The reduced expression of ITGA2 in lung tissue with the C allele of rs12522114 mimics vatelizumab-induced ITGA2 inhibition; this allele is associated with higher FEV_1_ and FEV_1_/FVC, indicating a potential to repurpose vatelizumab, which increases T regulatory cell populations^39^, for COPD.

### Pathway analysis

Using ConsensusPathDB^40^, we tested whether specific biological pathways were enriched for the 559 causal genes supported by 2 or more criteria, highlighting multiple pathways consistent with developmental pathways, tissue integrity and remodelling (**Supplementary Table 25**). These include pathways not previously implicated in pathway enrichment analyses for lung function such as PI3K-Akt signalling, integrin pathways, endochondral ossification, calcium signalling, hypertrophic cardiomyopathy, and dilated cardiomyopathy, as well as those previously implicated via individual genes^5^ such as TNF signalling, actin cytoskeleton, AGE-RAGE signalling, Hedgehog signalling and cancers. We also show strengthened enrichment by newly identified genes in pathways we previously described, such as extracellular matrix organisation (34 new genes), elastic fibre formation (8 genes), and TGF-Core (4 new genes). Consistent with our ConsensusPathDB findings, Ingenuity Pathway Analysis (https://digitalinsights.qiagen.com/IPA)^41^ highlighted enrichment of cardiac hypertrophy signalling and osteoarthritis pathways, and additionally implicated pulmonary and hepatic fibrosis signalling pathways, axonal guidance and PTEN signalling, and upstream regulators TGFB1 and IGF1 (**Supplementary Table 26**).

### Multi-ancestry genetic risk score associations with FEV_1_/FVC and COPD

We built multi-ancestry and ancestry-specific genetic risk scores weighted by FEV_1_/FVC effect sizes and tested for association with FEV_1_/FVC and COPD (GOLD stage 2-4) using independent testing datasets in different ancestry groups in UK Biobank (**Methods**). Our new GRS noticeably improved the predictive power for quantitative lung function and COPD compared with our previous GRS based only on European ancestry samples^5^ (Figure 5a and 5b, **Supplementary Table 18**) and the multi-ancestry GRS outperformed the ancestry-specific GRS in all ancestry groups in UK Biobank. We then tested the association of the multi-ancestry GRS with COPD susceptibility in five independent COPD case-control studies (**Supplementary Table 19, Methods**). Improved association results were observed across all the five European ancestry studies compared with previous GRS^5^ (Figure 5c, **Supplementary Table 20**). The odds ratio for COPD per standard deviation of the weighted GRS was 1.63 (95% CI: [1.56, 1.71], P=7.1×10^−93^) in the meta-analysis of these EUR studies compared to 1.55 (95% CI: [1.48, 1.62], P=2.9×10^−75^) using the previous GRS^5^. In SPIROMICS African ancestry individuals, results were comparable to UK Biobank African ancestry individuals, but of a lower magnitude in the COPDGene African ancestry population (Figure 5c).

**Figure 4:**
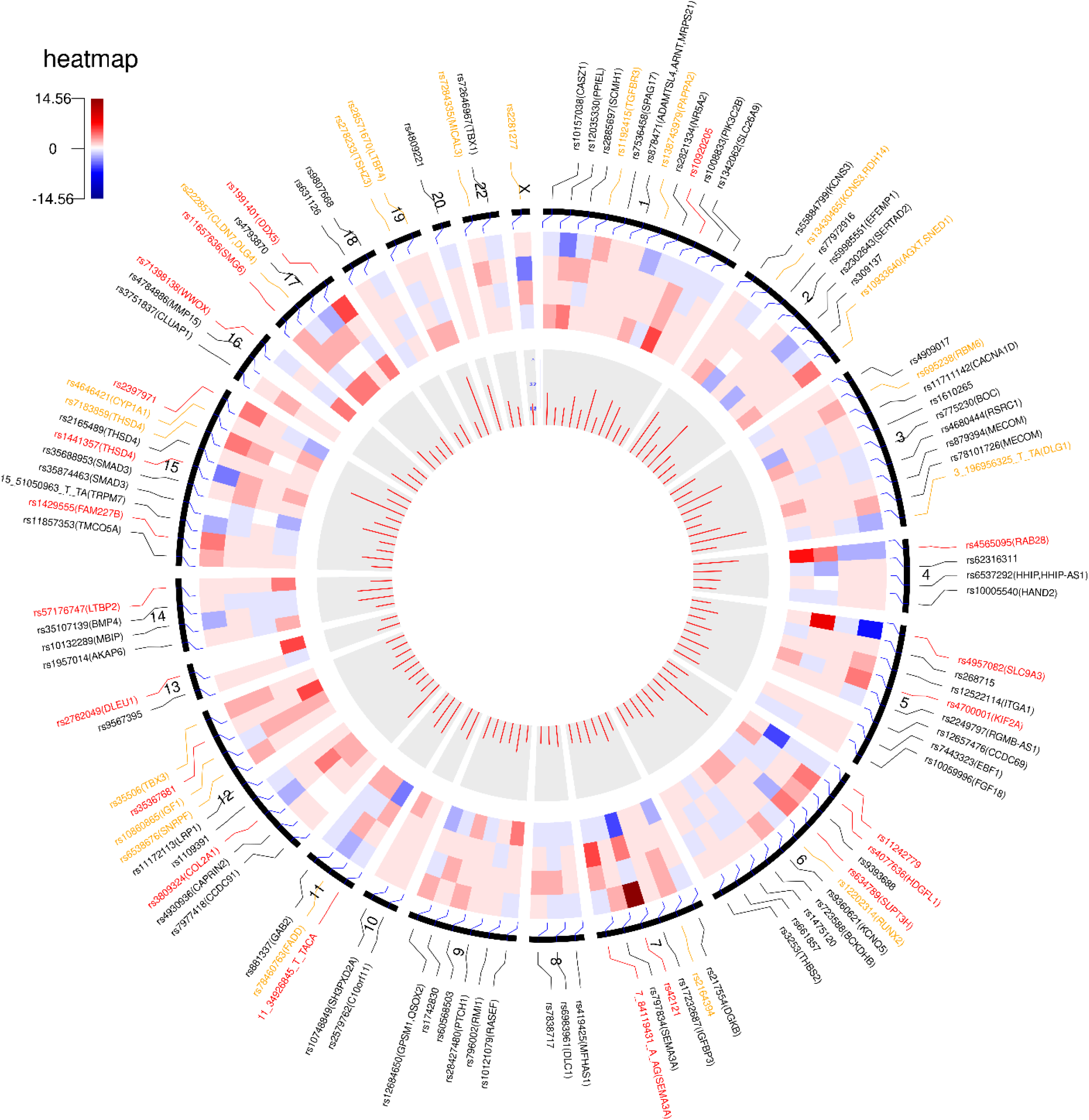
Summary of the 109 signals with nominal significant evidence for heterogeneity correlated to ancestry. The signals (represented by rs ID and the corresponding prioritised gene(s) which was(were) supported by the largest number of evidence resources for this signal and have at least two evidence resources across signals) are mapped to the corresponding chromosomes (outer segment). Signals attaining at least nominal evidence of association in non-EUR populations and with effect size at least 2(3) times larger than that in EUR populations are shown in orange (red). Circular tracks from inside to outside: (1). Bar plot shows the significance of heterogeneity correlated to ancestry by MR.MEGA (-log(p value)); (2). Heatmap shows the relative effect size (i.e. β_non-EUR_/β_EUR_) estimated in non-EUR cohorts in ancestry-specific meta-analysis compared with EUR (from inside to outside: SAS, EAS, AMR and AFR).

**Figure 5:**
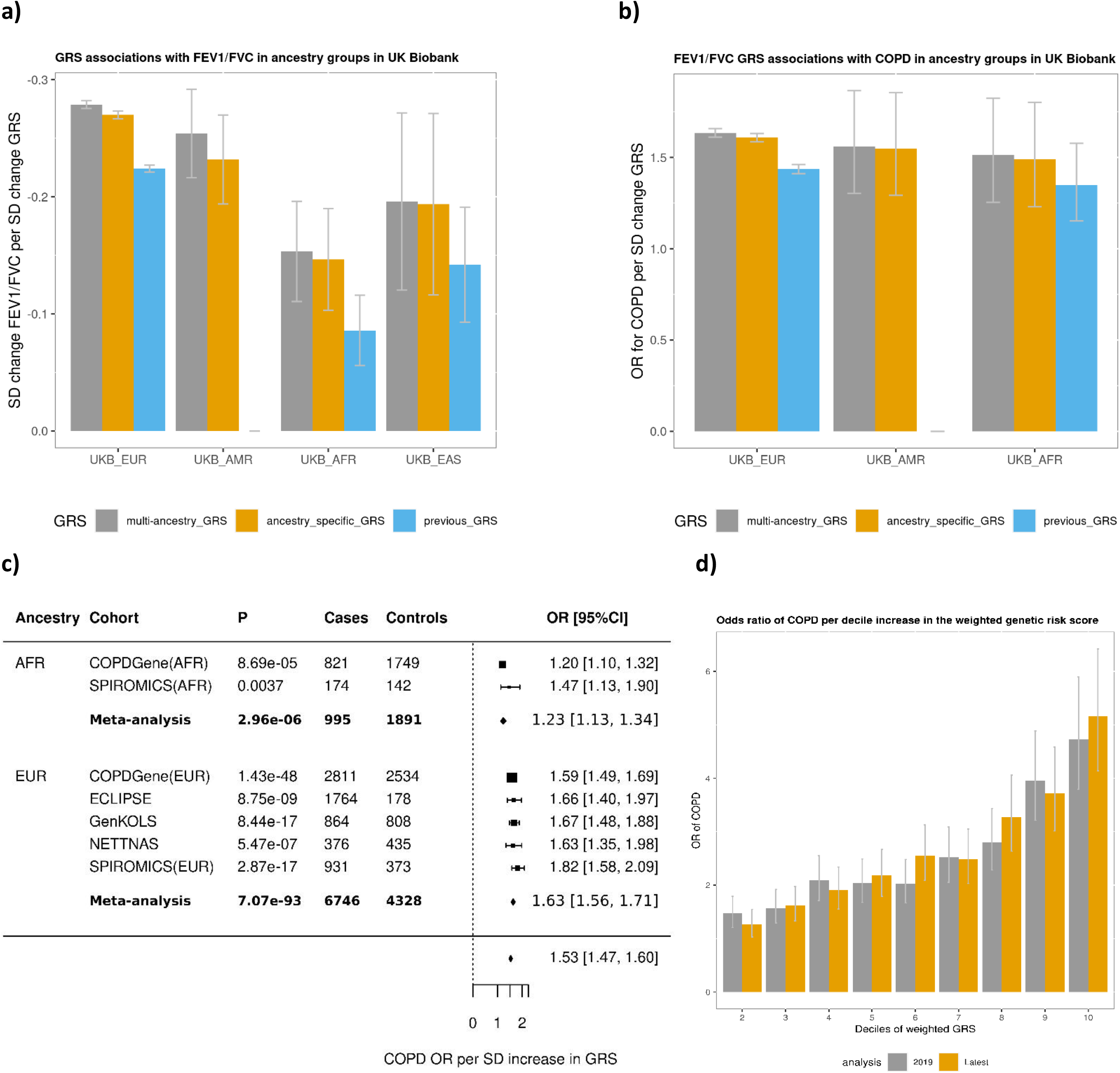
GRS performance: **a)** Prediction performance of 3 GRS across ancestry groups for FEV_1_/FVC shown as standard deviation (SD) change in FEV_1_/FVC per SD increase in GRS in the individuals of UK Biobank ancestry groups (whiskers represent 95% confidence intervals); **b)** Prediction performance of 3 GRS for COPD shown as COPD odds ratio per SD increase in GRS; **c)** OR for COPD per S.D. change in GRS in COPD case-control studies; **d)** decile analysis.

To aid clinical interpretation, we divided individuals in each of the five European ancestry COPD case-control studies into ten deciles according to their values of the multi-ancestry GRS. The odds ratio for COPD in members of the highest GRS decile compared to the lowest GRS decile was 5.16 (95% CI: [4.14, 6.42], P=1.0×10^−48^, **Supplementary Table 21**). The odds ratio for COPD showed steeper increase as the GRS decile increases using the multi-ancestry GRS compared with the previous GRS (Figure 5d).

### Phenome-wide associations of trait-specific genetic risk scores

To study the aggregate effects of genetic variants associated with each specific lung function trait on a wide range of diseases and disease-relevant traits, we created genetic risk scores (GRS) for each of, one for each trait FEV_1_, FVC, FEV_1_/FVC and PEF, and used each of these GRS in PheWAS. To construct each GRS, we included all sentinel variants associated with the trait (P <5×10^−9^), using the weights estimated from the multi-ancestry meta-regression (**Methods**), for a total of 425, 372, 442 and 194 variants in each trait-specific GRS respectively.

GRS constructed from the four lung function traits showed distinct patterns of associations with a range of respiratory and non-respiratory phenotypes in our PheWAS (**Figure 6**). A GRS for lower FEV_1_ was most strongly associated with increased risk of asthma and COPD, as well as family history of chronic bronchitis/emphysema, lower hand grip strength, increased fat mass, increased HbA1c and type 2 diabetes risk, and elevated C-reactive protein (CRP). Additionally, associations were seen with increased asthma exacerbations and lower age of onset for COPD (**Figure 6a**). The GRS for lower FEV_1_/FVC was associated with key respiratory phenotypes: increased risk of COPD and asthma, increased family history of chronic bronchitis/emphysema, increased emphysema risk, and increased risk of respiratory insufficiency or respiratory failure, younger age of onset for COPD but a slightly lower risk of COPD exacerbations (**Figure 6b**). In contrast, the GRS for lower FVC was strongly associated with many traits – among the strongest associations were with high CRP, increased fat mass, raised HbA1c and type 2 diabetes, raised systolic blood pressure, lower hand grip strength and raised alanine aminotransferase, as well as showing increased risk of clinical codes for asthma and COPD (**Figure 6c**). Whilst the GRS for lower FEV_1_/FVC was associated with increased standing height and sitting height, the GRS for lower FEV_1_ and FVC were associated with increased standing height but reduced sitting height. Broadly similar phenome-wide associations were seen for the PEF GRS as for the FEV_1_ GRS (**Figure 6d**).

**Figure 6:**
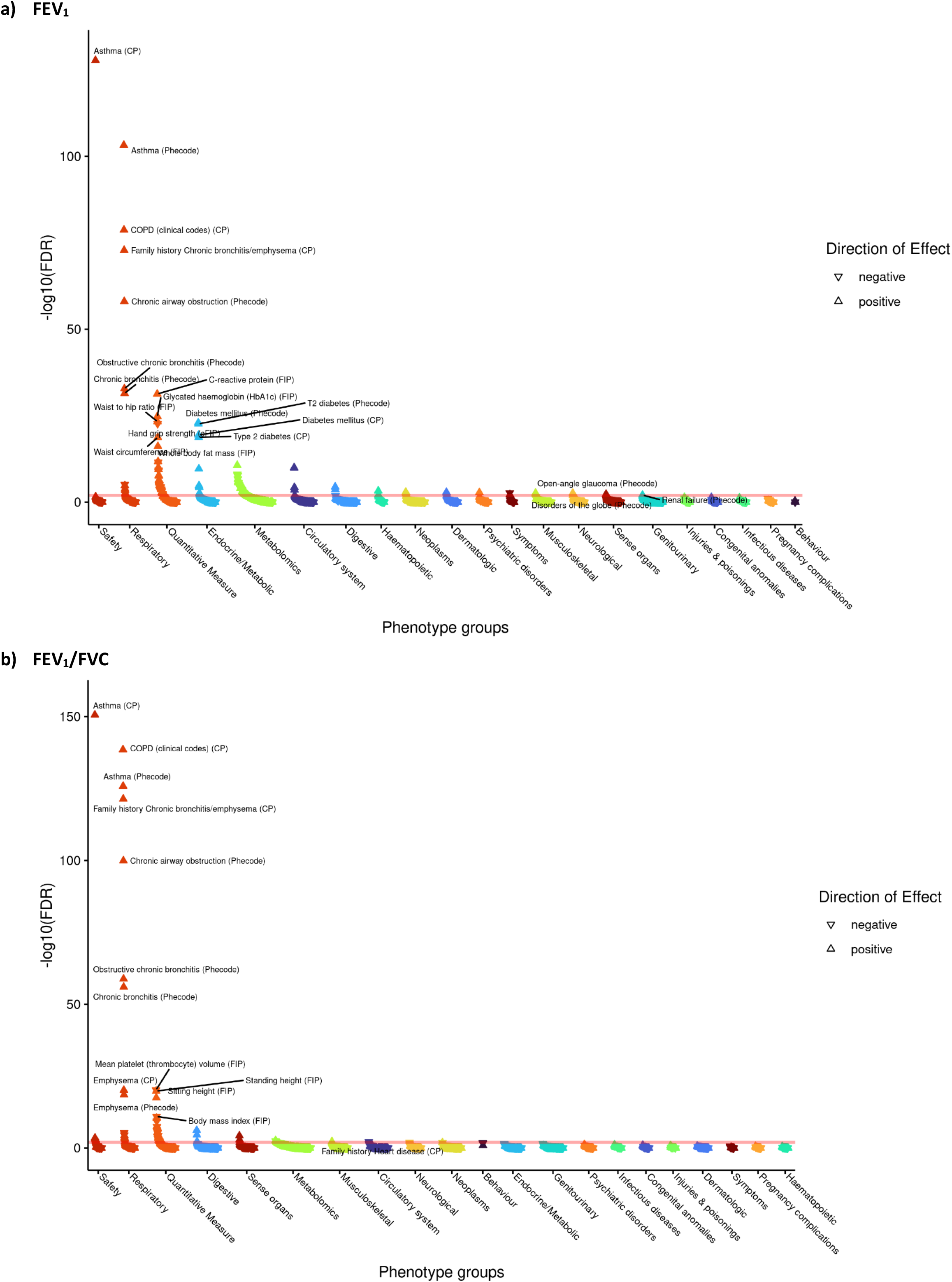
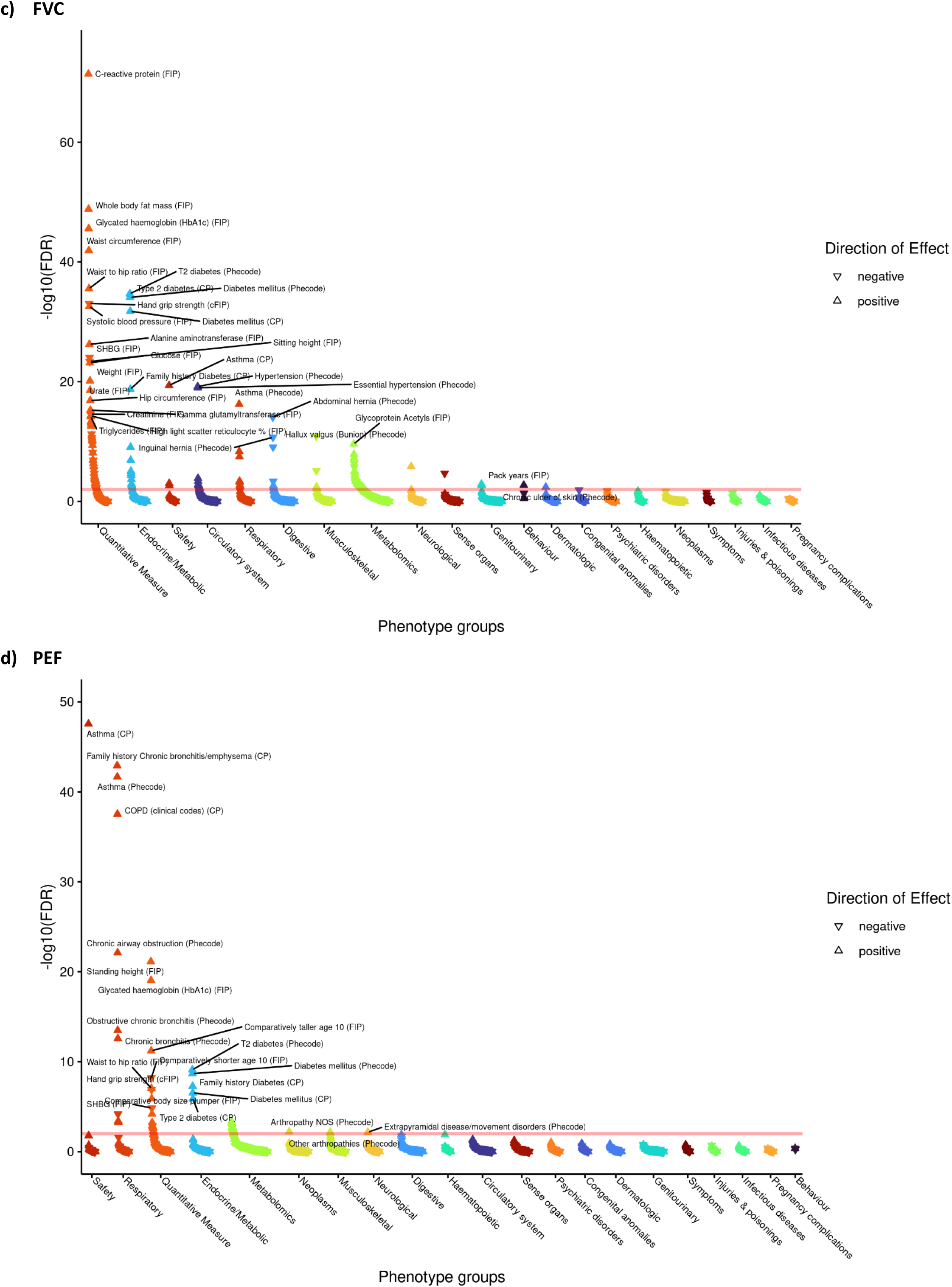
PheWAS of lung function trait GRS: **a)** FEV_1_; **b)** FEV_1_/FVC; **c)** FVC; **d)** PEF

### Phenome-wide associations of genetic risk scores partitioned by pathway

Finally, we hypothesised that partitioning our lung function GRS into pathway-specific GRSs according to the biological pathways the variants influence could inform understanding of mechanisms underlying lung function and COPD, and the likely consequences of perturbing specific pathways. Informed by the above prioritisation of putative causal genes and classification of these genes by pathway (**Pathway analysis**, above), we undertook PheWAS for FEV_1_/FVC GRSs partitioned by each of the 29 pathways enriched (FDR <10^−5^) for the 559 genes implicated by ≥2 criteria. In each case, we weighted the GRSs using the FEV_1_/FVC multi-ancestry meta-regression results (**Methods**). Partitioning GRSs in this way highlighted marked differences in patterns of phenome-wide associations (full results in **Supplementary Figures 15a-w** and **Supplementary Table 27**). We highlight four examples in **Figure 7**; whilst all four pathway-specific GRSs illustrated showed association with COPD clinical codes and with a family history of chronic bronchitis/emphysema, associations with other traits varied. The GRS for lower FEV_1_/FVC specific to elastic fibre formation was associated with increased risk of inguinal, abdominal, diaphragmatic and femoral hernia, diverticulosis, arthropathies, hallux valgus and genital prolapse, but reduced risk of carpal tunnel syndrome, as well as reduced BMI, and increased asthma risk (**Figure 7a**). In contrast, the GRS for lower FEV_1_/FVC specific to PI3K-Akt signalling was associated with increased asthma risk, lower IGF-1, liver enzymes (ALT, AST, gamma glutamyltransferase (GGT)) and lower lymphocyte count, raised eosinophils, lower fat free mass and BMI and reduced diabetes risk (**Figure 7b**). The GRS for lower FEV_1_/FVC specific to the hypertrophic cardiomyopathy pathway was associated with reduced liver enzymes (ALT, GGT), lower apolipoprotein B and lower LDL, lower IGF-1 and lower mean platelet volume (**Figure 7c**). The GRS associations for lower FEV_1_/FVC partitioned to signal transduction were specific to respiratory traits, including asthma and emphysema (**Figure 7d**). Variable height associations were evident: the GRS for lower FEV_1_/FVC showed association with increased height when partitioned to elastic fibre formation or hypertrophic cardiomyopathy (**Figure 7a,c**), reduced height when partitioned to ESC pluripotency (**Supplementary Figure 15g**), and no height association with height when partitioned to PI3K-Akt signalling or signal transduction (**Figure 7b,d**).

**Figure 7:**
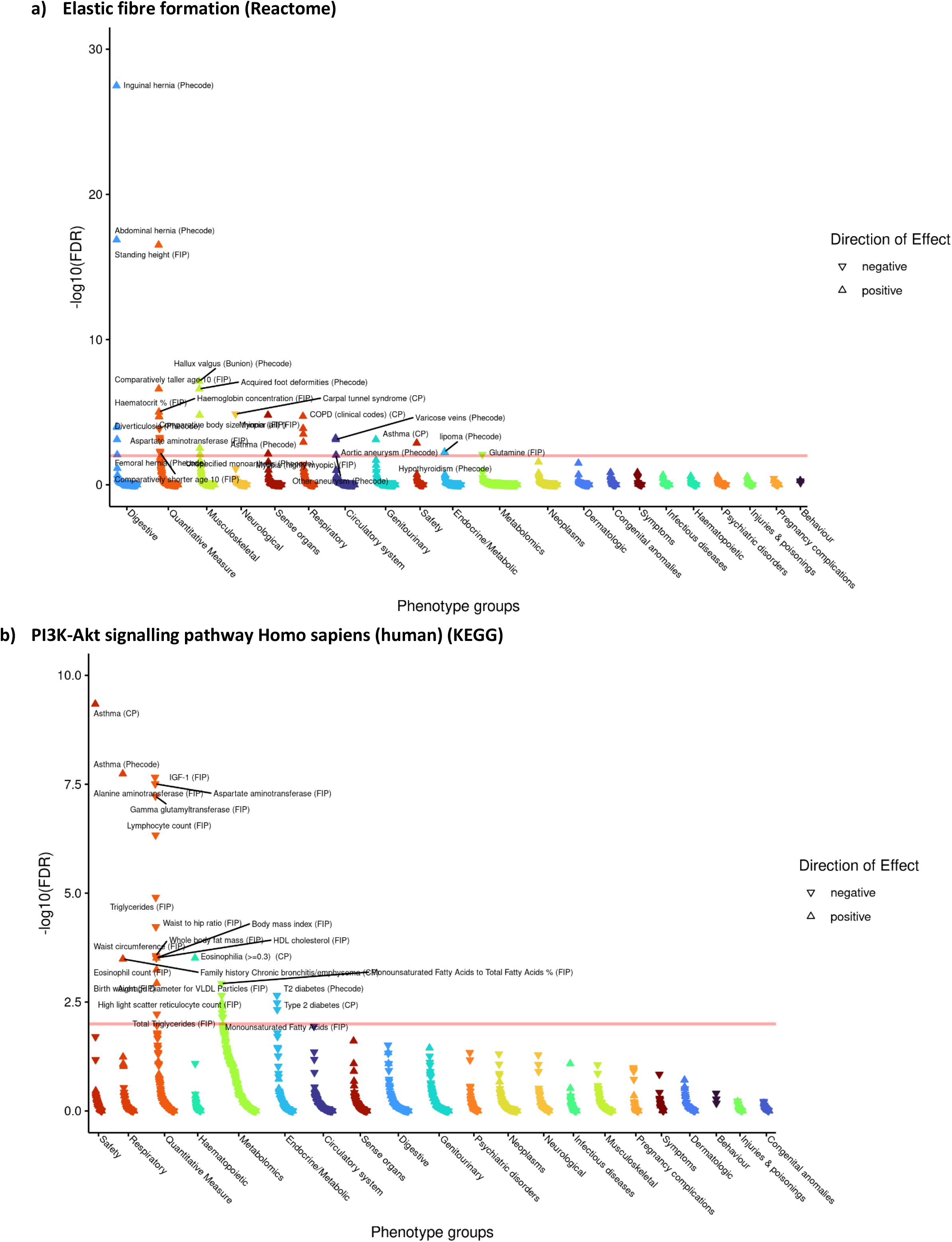
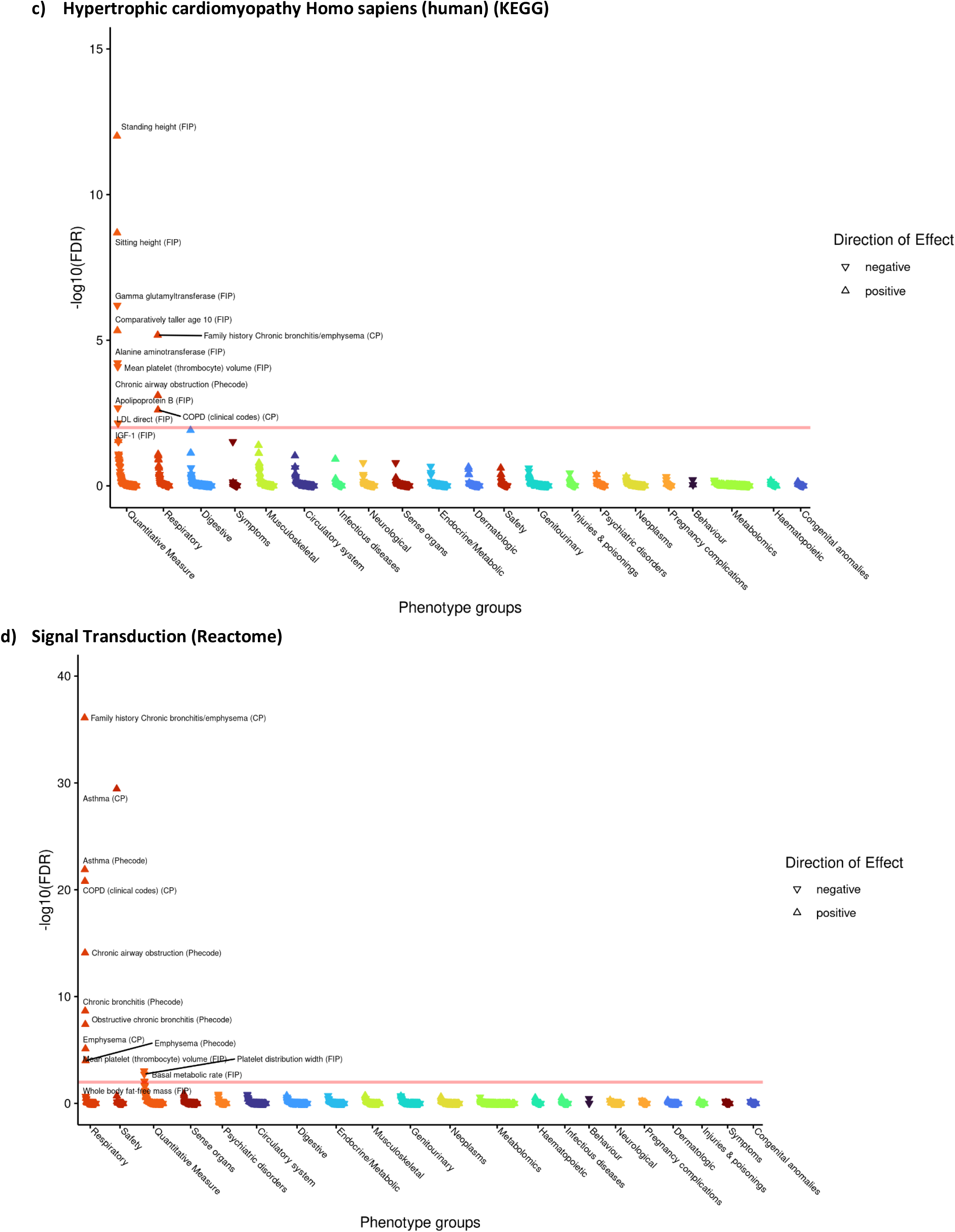
PheWAS for FEV_1_/FVC weighted GRS partitioned by: **a)** Elastic fibre formation (Reactome); **b)** PI3K-Akt signalling pathway Homo sapiens (human) (KEGG); **c)** Hypertrophic cardiomyopathy Homo sapiens (human) (KEGG); **d)** Signal Transduction (Reactome).

We hypothesised that individuals may have high GRS for one or more pathways and low GRS for other pathways. Comparing individuals’ GRS across pairs of pathways for each of 29 pathways (**Supplementary Figure 13b**) and in detail for the elastic fibre, PI3K-Akt signalling, hypertrophic cardiomyopathy and signal transduction pathways (**Supplementary Figure 13c**) show how GRS profiles may be concordant or discordant across pathways, which could have implications for choice of therapies.

## Discussion

Our study represents the largest and most ancestrally diverse GWAS of lung function to date and the most comprehensive initiative to relate lung function and COPD associated variants to functional annotations, cell types, genes and pathways. It is also the first to investigate possible phenotypic consequences of intervening in relevant pathways through PheWAS studies, utilising pathway-partitioned GRS.

The 1020 signals identified to date were enriched in functionally active regions in alveolar type 1 cells, fibroblasts and myofibroblasts, bronchial epithelial cells, adult and fetal lung. We showed effect heterogeneity attributable to ancestry for 109 signals (including *LTBP4, THSD4, EFEMP1, MECOM*), between ever-smokers and never-smokers (*HTR4*), and differences in effects between adults and children (*CCDC91, CYTL1, MECOM, MAPT*). We mapped lung function signals to 559 genes putatively inferred as causal based on meeting at least two independent criteria. Exemplar genes supported by ≥4 criteria or by deleterious or rare putative causal missense variants implicated surfactant phospholipid metabolism, smooth muscle function, epithelial morphology and barrier function, innate immunity, calcium signalling, adrenoceptor signalling, lung development and regeneration. Among the pathways enriched for the putative causal genes, were PI3K-Akt signalling, integrin pathways, endochondral ossification, calcium signalling, hypertrophic cardiomyopathy, and dilated cardiomyopathy that have not been previously implicated in lung function using GWAS approaches.

Combined as a genetic risk score weighted by FEV_1_/FVC effect size, the 1020 variants strongly predicted COPD in UK Biobank and in COPD case-control studies, with a more than five-fold change in risk between highest and lowest GRS deciles, illustrating the clinical relevance of our findings. This GRS more strongly predicted FEV_1_/FVC and COPD across all ancestries than a previously constructed risk score^5^. Partitioning this lung function GRS by the pathways defined by specific variants, informed by detailed, systematic variant-to-gene mapping and pathway analyses and using our new Deep-PheWAS platform^42^, illustrated unique patterns of phenotype associations for each pathway GRS. These patterns of PheWAS findings are relevant to the potential efficacy and potential side-effects of intervening in these pathways. As a proof-of-concept, the GRS associated with lower FEV_1_/FVC specific to PI3K-Akt signalling was associated with increased risk of COPD but a lower risk of diabetes; PI3K inhibition impairs glucose uptake in muscle and increases hepatic gluconeogenesis, contributing to glucose intolerance and diabetes^43^. The PheWAS and druggability analyses we conducted have potential to identify drug repurposing opportunities such as vatelizumab for COPD.

The patterns of pleiotropy we show through PheWAS for individual variants, for trait-specific GRS and pathway-partitioned GRS may help to explain variants and pathways that increase susceptibility to more than one disease, and thereby predispose to particular patterns of multimorbidity. For example, the elastic fibre pathway GRS was associated with increased risk of muscular (e.g. herniae) and musculoskeletal conditions related to connective tissue laxity. Our findings also help to further elucidate the complex relationship between height, body mass index or obesity, and lung function, and their genetic determinants^5,44^. We saw no overall correlation between the magnitude of lung function and height associations, and relationships differed between GRS for different lung function traits, and even between sitting and standing height for the same trait. The pathway-partitioned GRS indicate that the relationship between genetic variants, height and lung function traits depends on the pathways through which the variants act.

Our discovery effort was enabled by the largest worldwide collaboration to bring together multi-ancestry populations with curated lung function and genomic data, and to map these signals to putative causal genes. The last comprehensive attempt to map lung function associated variants to genes identified 107 putative causal genes, mostly through eQTLs only, and only eight genes were then implicated by ≥2 criteria^5^. In contrast, we implicated 559 causal genes meeting at least two criteria, through drawing upon new data and methodologies, such as single cell epigenome data, rare variant associations identified in sequencing data in UK Biobank and similarity-based approach PoPs^9^. Nevertheless, our study has limitations. Sample sizes for lung function genomics studies in all non-European ancestry groups fall far short of those in European ancestries, particularly in African ancestry populations^4^. Indeed, non-European ancestries are under-represented generally in genomic studies^3^, constraining both genome-wide and especially phenome-wide approaches in these populations. Correcting this will require substantial global investment in studies with suitably phenotyped and genotyped individuals, coupled with appropriate models of community participation and workforce development. Improved sample sizes across all ancestries would improve power for discovery in multi-ancestry meta-analyses and for ancestry-specific studies^44^, and for fine-mapping these genetic associations.

Strategies for in-silico mapping of association signals to causal genes are constantly evolving and difficult to evaluate until a reference set of fully functionally characterised lung function-associated variants and causal genes is developed. The framework we used to map signals to genes parallels one recently adopted^10^, showing the consensus between approaches in implicating each putative causal gene. We recognise that the in-silico evidence we used cannot firmly demonstrate causality, and confirmation of mechanism will require functional genomics experiments such as gene editing in suitable organoids with appropriate readouts. Our evidence will be of utility for prioritising such experiments. An additional limitation is that classifications of pathways may be imperfect; we used multiple pathway classifications as it is unclear which is superior across all component pathways, and we present the pathway-partitioned PheWAS results as a resource to others.

In summary, our multi-ancestry study highlights new putative causal variants, genes and pathways, some of which are targeted by existing drug compounds. These findings bring us closer to understanding mechanisms underlying lung function and COPD and will inform functional genomics experiments to confirm mechanisms and consequently guide the development of therapies for impaired lung function and COPD.

## Data Availability

All data produced in the present study are available upon reasonable request to the authors

