## Supplementary material for "Multi-ancestry genome-wide association study improves resolution of genes, pathways and pleiotropy for lung function and chronic obstructive pulmonary disease": Online Methods

#### GWAS in each cohort

All GWAS included appropriate adjustments for covariates age, sex, height, smoking and ancestry principal components and relatedness was accounted for using appropriate association software (BOLT-LMM or SAIGE). All phenotypes were rank inverse-normal transformed after adjustment. Quality control of the imputation in each cohort and the returned association summary statistics was performed by the central analysis team (**Supplementary Note**). We assigned each cohort to one of the five 1000 Genomes super-populations European (EUR), African (AFR), Admixed American (AMR), East Asian (EAS) or South Asian (SAS) based on self-reported ancestry apart from UK Biobank (57.4% of total sample size) where we used ADMIXTURE v1.3.0[1] to determine ancestry (**Supplementary Note, Supplementary Table 4**). MR-MEGA requires effect estimates from cohorts to be meta-analysed to be on the same scale hence we also required lung function association results from each cohort using untransformed phenotypes.

#### Meta-analysis

Before meta-analysis, association statistics in each cohort were adjusted by the LD score regression intercept calculated in each cohort to adjust for any residual confounding (**Supplementary Table 5**); the appropriate ancestry-specific LD reference was used for each cohort (10,000 UK Biobank samples for European cohorts and 1000 genomes project samples for African, Admixed American and South and East Asian cohorts). Before meta-analysis variants were excluded from each study with imputation INFO <0.5 or minor-allele count (MAC) <3. As transformed effect estimates were not on comparable scales we meta-analysed across cohorts using sample-size weighted, Z-score meta-analysis with METAL (released v2018-08-28)[2]. No genomic control was applied post-meta-analysis. After meta-analysis variants with MAC <20 were excluded.

#### Signal selection and conditional analysis

We chose a genome-wide significance threshold of $P < 5\times10^{-9}$ as recommended from sequencing studies[3]. We selected 2 Mb regions centred on the most significant variant for all regions containing a variant with $P < 5\times10^{-9}$. Regions within 500 kb of each other were merged for conditional analysis. Stepwise conditional analysis was run in each region in each cohort using GCTA v1.93.2beta[4] using an ancestry-specific LD reference for each cohort (**Supplementary Note**) and then conditional results were meta-analysed across cohorts and any new conditionally independent signals with $P < 5\times10^{-9}$ were added to our list of signals. moloc v0.1.0[5] was used to colocalise signals across the 4 lung function traits to obtain a set of distinct signals which were then colocalised with previously reported signals to obtain a set of novel lung function signals (**Supplementary Note**).

#### Exclusion of smoking signals from follow up

We checked our sentinels for association with smoking quantitative traits “Age of Initiation" (N=262,990) and "Cigarettes Per Day" (N=263,954), and binary traits "Smoking Cessation" (N cases=139,453; N controls=407,766) and "Smoking Initiation” (N cases=557,337; N controls=674,754) in the GWAS and Sequencing Consortium of Alcohol and Nicotine use (GSCAN) consortium[6] (proxies with $r^2$ >0.8 were checked for sentinels not present in GSCAN). We excluded 8 lung function signals from further analysis that we determined to be primarily driven by smoking behaviour (**Supplementary Table 7**) by the following criteria: (i) $P < 4.86\times10^{-5}$ (Bonferroni corrected 5% threshold for 1028 signals) for association with any smoking trait; (ii) the same “risk” allele that increases smoking exposure behaviour and decreases lung function.

#### Heritability estimate

We calculated the proportion of variance explained by the sentinels reported for each trait using the formula:

$$\frac{\sum_{i=1}^{n} 2f_i(1-f_i)\beta_i^2}{V}$$

where n is the number of variants, $f_i$ and $\beta_i$ are the frequency and effect estimate of the i'th variant from the UK Biobank European ancestry untransformed results, and V is the phenotypic variance (always 1 as our phenotypes were inverse-normal transformed). We assumed a heritability of 40%[7,8] to estimate the proportion of additive polygenic variance.

### Ancestry-adjusted trans-ethnic meta-analysis with MR-MEGA

To improve fine-mapping resolution using LD differences between ancestries and to estimate heterogeneity of variant associations attributable to ancestry, we undertook multi-ancestry meta-regression with MR-MEGA v0.2[9], which incorporates axes of genetic ancestry as covariates. MR-MEGA uses multidimensional scaling (MDS) of allele frequencies across cohorts to derive principal axes of genetic variation to use for ancestry adjustment (**Supplementary Note**). The location of the cohorts on the first two MDS-derived principal components, plotted in **Supplementary Figure 4**, show clustering in accordance with the assigned ancestry groups. We used 4 principal components for ancestry adjustment, as this captured most of the variance in ancestry and meant that we could meta-analyse the subset of 960 signals with results from at least 7 cohorts (MR-MEGA requires at least the number of principal components + 3 cohorts). MR-MEGA implements genomic control at study level, therefore, no further genomic control was applied. We ran MR-MEGA at each locus containing one or more signals; in the loci with multiple signals we ran MR-MEGA multiple times each time conditioning on all except 1 signal at the locus to obtain the conditional results for each signal. For each sentinel, we obtained an estimated ancestry-associated (P-value_ancestry_het) and residual (P-value_residual_het) heterogeneity. Additionally, MR-MEGA reports the log of Bayes factor, which can be used for the construction of credible sets.

### Effects in children

We meta-analysed associations for the top signals (972 out of 1020 available in at least one children cohort) with the lung function traits in three European-ancestry children's cohorts: USC children's study, ALSPAC and Raine Study (Age 13-15) (i.e. $FEV_1$, FVC and $FEV_1$/FVC signals were meta-analysed from the three cohorts but PEF signals were meta-analysed from ALSPAC and Raine Study). To investigate the age-dependent effects of genetic variants on lung function, we compared the effect sizes estimated from untransformed phenotypes in children of the top signals with adults in the European-specific meta-analysis (N = 35 cohorts for $FEV_1$ and FVC; 34 for $FEV_1$/FVC; 16 for PEF) by Welch's t test. Bonferroni significance threshold for 972 tests was applied ($P < 5.14\times10^{-5}$).

### Cell-type and functional specificity

#### Stratified LD score regression

We tested for enrichment of regulatory features at variants overlapping four histone marks (H3K27ac, H3K9ac, H3K4me3, H3K4me1) that are specific to lung, fetal lung and smooth muscle containing cell lines (colon and stomach) using stratified LD-score regression[10]. In this study, we only considered European specific meta-analysis with 39 cohorts for FVC, $FEV_1$ and $FEV_1$/FVC (17 cohorts for PEF). For the analysis of cell-type-specific annotations, we assessed statistical significance at the 0.05 level after Bonferroni correction for 48 hypotheses tested. Since these annotations are not independent, a Bonferroni correction is conservative. We also report results with false discovery rate (FDR) <0.05 using the Benjamini-Hochberg method.

#### Regulatory and functional enrichment using GARFIELD

We tested enrichment of SNPs at different types of functional annotation regions (DNase I hypersensitivity hotspots (DHS), open chromatin peaks, transcription factor footprints and FAIRE, histone modifications, chromatin segmentation states, genic annotations and transcription factor binding sites) using GARFIELD[11]. We used the European ancestry meta-analysis with 17 European cohorts for PEF and 39 European cohorts for FVC, $FEV_1$ and $FEV_1$/FVC. We applied GARFIELD to DHS annotation in 424 cell lines and primary cell types from ENCODE and Roadmap Epigenomics and derived enrichment estimates at trait-genotype association P-value thresholds at $P < 5\times10^{-5}$ and $P < 5\times10^{-9}$.

#### Enrichment of annotations in respiratory relevant cell types and tissues

We curated annotations from the following assays of respiratory relevant cells and tissues (i) Single-cell genome ATAC-seq data[12] from 19 cell types: *Myofibroblast, Pericyte, Ciliated, T_cell, Club, Capillary_endothelial_1/2, Basal,*

*Matrix_fibroblast_1/2, Arterial_endothelial, Pulmonary_neuroendocrine, Natural_killer_cell, Macrophage, B_cell, Erythrocyte, Lymphatic_endothelial, Alveolar Type 1/2* (downloaded from https://www.lungepigenome.org/)*;* (ii) ATAC-seq data for 5 human primary lung-cell types implicated in COPD pathobiology[13]; cell types: *large and small airway epithelial cells, alveolar type II, pneumocytes and lung fibroblasts* (downloaded from http://www.copdconsortium.org/); (iii) tissue-specific transcription factor binding sites from DNase-seq footprinting of 589 human transcription factors in lung and bronchus[14]. We tested for cell and tissue specific enrichment of these annotations at our lung function signals using fGWAS[15] (**Supplementary Note**).

### Identification of putative causal genes and variants

#### eQTL/pQTL colocalisation

Three eQTL resources were used for colocalisation of lung function signals with gene expression signals: (i) GTEx V8 (downloaded from https://www.gtexportal.org/ July 2020, tissues: *Stomach, Small Intestine Terminal Ileum, Lung, Esophagus Muscularis, Esophagus Gastroesophageal Junction, Colon Transverse, Colon Sigmoid, Artery Tibial, Artery Coronary, Artery Aorta*); (ii) eQTLgen[16] blood eQTLs; (iii) UBC lung eQTL[17]. Two blood pQTL resources were used to colocalise with associations with protein levels: (i) INTERVAL pQTL[18] and (ii) SCALLOP pQTL (http://scallop-consortium.com/). eQTL and pQTL colocalisation was tested with coloc_susie method[19] (**Supplementary Note**).

#### Rare variants from exome sequencing

We checked for rare (MAF <1%) exonic associations near (±500 kb) our lung function sentinels using both single-variant and gene-based collapsing tests from (i) single variant and gene-based associations in 281,104 UK Biobank exomes from the AstraZeneca PheWAS Portal[20] (https://azphewas.com/), (ii) single variant and gene-based tests of loss-of-function and missense variants in 454,787 UK Biobank participants[21], (iii) gene-based tests on whole-exome imputation in 500,000 UK Biobank participants[22]. We used a threshold of $P < 5 \times 10^{-6}$ for both single-variant and gene-based tests.

#### Nearby mendelian respiratory disease genes

We selected rare mendelian-disease genes from ORPHANET (https://www.orpha.net/) within ±500kb of a lung function sentinel that were associated with respiratory terms matching regular expression: *respir, lung, pulm, asthma, COPD, pneum, eosin, immunodef, cili, autoimm, leukopenia, neutropenia, Alagille syndrome.* We implicated the gene if it has a corresponding respiratory term match in the disease name or if it appears frequently in human phenotype ontology (HPO) terms for that disease.

#### Nearby mouse knockout orthologs with a respiratory phenotype

We selected human orthologs of mouse knockout genes with phenotypes in the “respiratory” category, as listed in the International Mouse Phenotyping consortium (https://www.mousephenotype.org/) within ±500kb of a lung function sentinel.

#### Polygenic priority score

We used a similarity-based method to calculate a gene-level Polygenic Priority Score (PoPS)[23] based on the assumption that causal genes share functional characteristics. PoPS assumes that if the associations enriched in genes share functional characteristics with a gene nearby a lung function signal, then that gene is more likely to be causal. The full set of gene features used in the analysis included 57,543 total features – 40,546 derived from gene expression data, 8,718 extracted from a protein-protein interaction network, and 8,479 based on pathway membership. In this study, we prioritized genes for all autosomal lung function signals within a 500kb (±250kb) window of the sentinel and reported the top prioritised genes in the region. For the signals that did not have prioritized genes within the 500kb window, we looked for prioritized genes using a 1Mb (±500kb) window (**Supplementary Note**).

#### Annotation-informed credible sets

We used the enriched annotations in respiratory relevant cell types and tissues and enriched genic annotations to create annotation-informed 95% credible sets with fGWAS, based on the MR-MEGA ancestry-adjusted meta-regression results (**Supplementary Note**). We implicated a putative causal missense variant if it accounted for >50% of the posterior probability in the credible set and annotated these with Ensembl Variant Effect Predictor[24], to check for a deleterious effect by the SIFT, PolyPhen or CADD metrics.

### Allocation of genes prioritised with 3 or more variant-to-gene to lung function biology categories

We allocated prioritised genes with 3 or more criteria to different lung function roles (epithelial, inflammatory, peripheral lung (including alveolus and endothelial),lung remodelling (including connective tissue), chest wall movement and lung development – Figure 3) based on literature review, including GeneCards (https://www.genecards.org) and PubMed (https://pubmed.ncbi.nlm.nih.gov). Eighteen of the genes were difficult to assign to a specific category on this basis, mainly because they were involved in generic processes such as transcriptional control in a wide variety of cell types, these are not shown in Figure 3 but are included in **Supplementary Table 14**.

### Summary of lung function biology

To provide an illustration of the biology of genes relating to lung function, we undertook a literature review using GeneCards (https://www.genecards.org) and PubMed (https://pubmed.ncbi.nlm.nih.gov) for genes implicated by 3 or more variant-to-gene criteria. We then allocated genes to one or more of the following: epithelial, inflammatory, peripheral lung (including alveolus and endothelial), lung remodelling (including connective tissue), chest wall movement, and lung development. Genes involved in generic processes such as transcriptional control in a wide variety of cell types were not assigned to the above categories and were omitted from the figure.

### Interaction with smoking

Association testing for lung function traits ($FEV_1$, FVC, $FEV_1$/FVC and PEF) was calculated separately in ever and never smokers subgroups and meta-analysed across European ancestry cohorts. We included untransformed phenotypes from which we had individual ever and never smoking summary statistics (N=28 cohorts), comprising 206,162 ever smokers and 229,046 never smokers. A z-test was used to compare genetic effect between ever and never smokers untransformed association results:

$$z = \frac{\beta_1 - \beta_2}{\sqrt{se_1^2 + se_1^2}}$$

We considered as significant evidence of interaction any signal with a P <$4.9 \times 10^{-5}$ (5% Bonferroni corrected for 1,020 signals tested).

### Genetic risk score

We selected four ancestry groups in UK Biobank as test data sets (SAS was excluded from GRS analyses because UKB_SAS was the only cohort in the multi-ancestry analysis for South Asian): UKB_EUR, UKB_AMR, UKB_EAS, UKB_AFR. All the other cohorts except UKB_SAS and QBB were used as discovery datasets.

We repeated the multi-ancestry meta-regression (MR-MEGA), after excluding the four test GWAS, incorporating the same four axes of genetic variation as covariates to account for ancestry. Autosomal signals identified in the signal selection process for each lung function trait and reported in the target ancestry population were included in downstream analysis for each ancestry. For ancestry *j* (*j*=EUR, AMR, EAS or AFR), we estimated ancestry-specific predicted allelic effects for $i^{th}$ SNP to be used as weights in the multi-ancestry GRS by

$$\hat{b}_{ij} = \alpha_{0i} + \sum_{k=1}^{4} \alpha_{ki}\bar{x}_{kj}$$

where $\bar{x}_{kj}$ is the averaged position of discovery studies with ancestry *j* on the $k^{th}$ axis of genetic variation from multi-ancestry meta-regression, and $\alpha_{0i}$ and $\alpha_{ki}$ denote the intercept and effect of the $k^{th}$ axis of genetic variation for the $i^{th}$ SNP from the multi-ancestry meta-regression.

We ran each of the ancestry-specific fixed-effect meta-analyses after excluding the test GWAS from the ancestry group using METAL with inverse-variance weighting method. For comparison, SNPs used as weights in multi-ancestry GRS were selected to build ancestry-specific GRS for each ancestry.

### Testing genetic risk score in independent COPD case-control cohorts

We tested association of multi-ancestry GRS with COPD susceptibility in five European-ancestry COPD case-control studies: COPDGene (Non-Hispanic White) (2,811 cases, 2,534 controls), ECLIPSE (1,764 cases, 178 controls), GenKOLS (864 cases, 808 controls), NETT/NAS (376 cases, 435 controls) and SPIROMICS (Non-Hispanic White) (931 cases, 373 control) (**Supplementary Table 19**). We also tested the association in two African-ancestry COPD case-control studies: COPDGene (African-American) (821 cases, 1,749 controls) and SPIROMICS (African-American) (174 cases, 142 controls) (**Supplementary Table 19**). Associations were tested using logistic regression models, adjusted for age, $age^2$, sex, height and principal components. In each COPD case-control study, we divided individuals into deciles according to their values of the weighted GRS. For each decile, logistic models were fitted to compare the risk of COPD for members of the decile compared to those with lowest decile (i.e. those with lowest genetic risk). Results were meta-analysed by ancestry-specific study groups using the fixed effect model.

### PheWAS

We used Deep-PheWAS[25], which addresses both phenotype matrix generation and efficient association testing while incorporating the following developments that are not yet available in current platforms and online resources: (i) clinically-curated composite phenotypes for selected health conditions that integrate different data types (including primary and secondary care data) to study phenotypes not well captured by current classification trees; (ii) Integrationi of quantitative phenotypes from primary care data, such as pathology records and clinical measures; (iii) clinically-curated phenotype selection for traits that are extremely highly correlated; (iv) genetic risk scores. The platform includes 2421 phenotypes in UK Biobank, with a subset of 2243 recommended for association testing: some phenotypes that are generated are used solely in the definition of other phenotypes and do not have utility by themselves. We removed the four measures of lung function and added seven phenotypes defined in-house (P4002-6 see **Supplementary Table 22**) to give 2246 as our final maximum number of phenotypes for association (**Supplementary Table 22**). Deep-PheWAS then filters these, requiring a minimum case number; we chose to keep the default settings of 50 -case minimum for binary phenotypes and a 100-case minimum for quantitative phenotypes. After limiting to European ancestry and filtering for case numbers this left 1909 phenotypes for association analysis. No additional phenotypes were removed when removing pairs related up to 2$^{nd}$ degree (KING kinship coefficient >=0.0884).

There are five types of phenotypes within Deep-PheWAS categorised according to the data and methods used to create them. Composite phenotypes are made using linked hospital and primary care data including in some cases primary care prescription data, alongside any of the UK Biobank field-IDs including self-reported non-cancer diagnosis and self-reported operations. Phecodes are defined using only linked hospital data (https://phewascatalog.org/phecodes_icd10). Formula phenotypes combine available data using bespoke R code per phenotype rather than using the inbuilt functions of phenotype development available in Deep-PheWAS. Added phenotypes are lists of cases and controls that have been added to the PheWAS and not developed by the Deep-PheWAS phenotype matrix generation pipeline. More complete definitions for all none-added phenotypes can be found in the Deep-PheWAS description[25].

#### Single variant PheWAS

We ran 27 single-variant PheWAS across 1909 traits (**Supplementary Table 22**) in UK Biobank in up to 430,402 unrelated European individuals. For 20 genes with 4 or more lines of evidence for being causal (**Supplementary Table 14**) we selected for each gene the variant with the most significant P value that implicates the gene. A further 7 variants were included in single-variant PheWAS that were putatively causal (accounted for >50% posterior probability in the credible set and had a deleterious annotation; **Supplementary Table 15**) but within a gene that was implicated by fewer than 4 lines of evidence. We also included 4 variants implicating the gene *CACNA1D*, one of several genes prioritised encoding calcium voltage-gated channel subunits and the genes *MECOM*, *CCDC91* and *MAPT* where a different effect was observed in adults and children. The single variant PheWAS was aligned to the lung function trait decreasing allele. Where we noted associations with testosterone and sex hormone binding globulin, we also undertook sex-stratified PheWAS.

#### Association with trait-specific genetic risk score

We created 4 genetic risk scores (GRS) for UK Biobank European samples, one for each trait $FEV_1$, FVC, $FEV_1$/FVC and PEF including all conditionally independent sentinel variants for the trait that were associated with $P < 5\times10^{-9}$, giving 425, 372, 442 and 194 variants in each trait-specific GRS respectively. Each of the 4 GRS were weighted by the effect sizes from the multi-ancestry meta-regression for the relevant trait and then checked for association with 1909 traits in the PheWAS.

### Association with pathway-specific genetic risk score

We selected 29 pathways that were enriched at FDR <$10^{-5}$ for our 559 genes implicated by 2 or more lines of evidence (**Supplementary Table 25**). We created a weighted GRS (weights estimated from multi-ancestry meta-regression for $FEV_1$/FVC) for each of the 29 pathways by including for each gene in the pathway, as for the single-variant PheWAS above, the variant with the most significant P value for the trait that implicates the gene in our variant-to-gene mapping (**Supplementary Table 25)**. Each of the 29 GRS were then checked for association with 1909 traits in the PheWAS.
