## Supplementary Note for "Multi-ancestry genome-wide association study improves resolution of genes, pathways and pleiotropy for lung function and chronic obstructive pulmonary disease"

### Extended acknowledgements and funding information

#### The following authors report specific personal funding from the following grants:

**A. Guyatt**: Wellcome Trust Institutional Strategic Support Fund (204801/Z/16/Z), BHF Accelerator Award (AA/18/3/34220).

**C. John**: Medical Research Council Clinical Research Training Fellowship (MR/P00167X/1).

**A. T. Williams**: BBSRC CASE studentship with GSK.

**L. V. Wain**: GSK/British Lung Foundation Chair in Respiratory Research.

**M. D. Tobin**: Wellcome Trust Investigator Award (WT202849/Z/16/Z).

**M.D. Tobin and L. V. Wain**: MRC (MR/N011317/1).

**M.D. Tobin and I. P. Hall** hold NIHR Senior Investigator Awards.

**I. Sayers:** MRC (G1000861)

**A. Morris:** Wellcome Trust (WT098017 & WT064890)

**C. Batini** was supported by a UKRI Innovation Fellowship at Health Data Research UK (MR/S003762/1).

The research was supported by BREATHE — The Health Data Research Hub for Respiratory Health (MC_PC_19004).

The research was partially supported by the NIHR Leicester Biomedical Research Centre and the NIHR Nottingham Biomedical Research Centre; views expressed are those of the author(s) and not necessarily those of the NHS, the NIHR or the Department of Health. The funders had no role in the design of the Mendelian randomization analyses.

This research was funded in whole, or in part, by the Wellcome Trust.

For the purpose of open access, the author has applied a CC BY public copyright licence to any Author Accepted Manuscript version arising from this submission.

#### Cohort and study-group specific acknowledgements and funding:

**ALHS:** The Agricultural Lung Health Study (ALHS) was supported by the Intramural Research Program of the National Institutes of Health, National Institute of Environmental Health Sciences (Z01-ES102385, Z01-ES049030, Z01-ES043012, and for ABW, contract no. HHSN273201600003I) and the National Cancer Institute (Z01-CP010119). ALHS also supported in part by American Recovery and Reinvestment Act (ARRA) funds through NIEHS contract number N01-ES-55546.ALHS thank the numerous study staff at Social & Scientific Systems, Inc. who played a role in the data collection, and acknowledge Dr. Nathan Gaddis, RTI, International for assistance performing genotype QC and HRC imputation for ALHS.

**B58C:** Acknowledge use of phenotype and genotype data from the British 1958 Birth Cohort DNA collection, funded by the Medical Research Council grant G0000934 and the Wellcome Trust grant 068545/Z/02. Genotyping for the B58C-WTCCC subset was funded by the Wellcome Trust grant 076113/B/04/Z. The B58C-T1DGC genotyping utilized resources provided by the Type 1 Diabetes Genetics Consortium, a collaborative clinical study sponsored by the National Institute of Diabetes and Digestive and Kidney Diseases (NIDDK), National Institute of Allergy and Infectious Diseases (NIAID), National Human Genome Research Institute (NHGRI), National Institute of Child Health and Human Development (NICHD), and Juvenile Diabetes Research Foundation International (JDRF) and supported by U01 DK062418. B58C-T1DGC GWAS data were deposited by the Diabetes and Inflammation Laboratory, Cambridge Institute for Medical Research (CIMR), University of Cambridge, which is funded by Juvenile Diabetes Research Foundation International, the Wellcome Trust and the National Institute for Health and Care Research Cambridge Biomedical Research Centre; the CIMR is in receipt of a Wellcome Trust Strategic Award (079895). The B58C-GABRIEL genotyping was supported by a contract from the European Commission Framework Programme 6 (018996) and grants from the French Ministry of Research.

**BHS:** National Health and Medical Research Council (NHMRC), Ideas Grant 2003629 (funding to Benjamin H. Mullin). Department of Health Western Australia, Merit Award 1186046 (funding to Benjamin H. Mullin). The Busselton Health Study is supported by the Government of Western Australia (Office of Science and Department of Health), the City of Busselton and private donations to the Busselton Population Medical Research Institute. Generous support for the 1994/5 follow-up study was also received from Healthway, Western Australia, and support from The Great Wine Estates of the Margaret River region of Western Australia. We also thank the WA Country Health Service and acknowledges the numerous Busselton community volunteers who assisted with data collection and the study participants from the Shire of Busselton.

**COPDGene and ECLIPSE (COPD case-control studies):** received support from the NIH, grant numbers: R01HL149861 (funding to Cho), R01HL135142 (funding to Cho, Hobbs), R01HL137927 (funding to Cho, Silverman), R01HL089856 (funding to Cho, Silverman), R01HL147148 (funding to Cho, Silverman), K08HL136928 (funding to Hobbs), P01HL114501 (funding to Silverman) and P01HL132825 (funding to Silverman, Cho). The **COPDGene** project (NCT00608764) was supported by Award Number U01 HL089897 and Award Number U01 HL089856 from the National Heart, Lung, and Blood Institute. The content is solely the responsibility of the authors and does not necessarily represent the official views of the National Heart, Lung, and Blood Institute or the National Institutes of Health. The COPDGene® project is also supported by the COPD Foundation through contributions made to an Industry Advisory Board that has included AstraZeneca, Bayer Pharmaceuticals, Boehringer Ingelheim, Genentech, GlaxoSmithKline, Novartis, Pfizer, and Sunovion. The **ECLIPSE** study (NCT00292552; GSK code SCO104960) and GenKOLS were funded by GlaxoSmithKline. The COPDGene, ECLIPSE, GenKOLS, and NETT/NAS studies would like to acknowledge Pooja Srinivasa and Chandan Pavuluri for their programming support.

**CHS**: This CHS research was supported by NHLBI contracts HHSN268201200036C, HHSN268200800007C, HHSN268201800001C, N01HC55222, N01HC85079, N01HC85080, N01HC85081, N01HC85082, N01HC85083, N01HC85086, 75N92021D00006; and NHLBI grants U01HL080295, R01HL085251, R01HL087652, R01HL105756, R01HL103612, R01HL120393, and U01HL130114 with additional contribution from the National Institute of Neurological Disorders and Stroke (NINDS). Additional support was provided through R01AG023629 from the National Institute on Aging (NIA). A full list of principal CHS investigators and institutions can be found at CHS-NHLBI.org. The provision of genotyping data was supported in part by the National Center for Advancing Translational Sciences, CTSI grant UL1TR001881, and the National Institute of Diabetes and Digestive and Kidney Disease Diabetes Research Center (DRC) grant DK063491 to the Southern California Diabetes Endocrinology Research Center. The content is solely the responsibility of the authors and does not necessarily represent the official views of the National Institutes of Health. **CROATIA-Korcula/Split/Vis:** MRC Human Genetics Unit core funding, the Ministry of Science, Education and Sport in the Republic of Croatia (216-1080315-0302) and the Croatian Science Foundation (grant 8875) (funding to I. Rudan, C. Hayward, S.M. Kerr, O. Polasek, V. Vitart, and J. Marten).

**CKB:** The CKB baseline survey and the first re-survey were supported by the Kadoorie Charitable Foundation in Hong Kong. Long-term follow-up was supported by the Wellcome Trust (212946/Z/18/Z, 202922/Z/16/Z, 104085/Z/14/Z, 088158/Z/09/Z), the National Key Research and Development Program of China (2016YFC0900500, 2016YFC0900501, 2016YFC0900504, 2016YFC1303904), and the National Natural Science Foundation of China (91843302). DNA extraction and genotyping was funded by GlaxoSmithKline and the UK Medical Research Council (MC-PC-13049, MC-PC-14135). The project is supported by core funding from the UK Medical Research Council (MC_UU_00017/1,MC_UU_12026/2, MC_U137686851), Cancer Research UK (C16077/A29186; C500/A16896), and the British Heart Foundation (CH/1996001/9454) to the Clinical Trial Service Unit and Epidemiological Studies Unit and to the MRC Population Health Research Unit at Oxford University. CKB gratefully acknowledges the participants in the study, the members of the survey teams in each of the 10 regional centres, and the project development and management teams based at Beijing, Oxford and the 10 regional centres.

**EPIC-Norfolk:** received support from the MRC, grant numbers: MR/N003284/1, MC-UU_12015/1, MC_UU_00006/1, MC_PC_13048 and C864/A14136, and the Cancer Research UK, grant number C864/A14136. EPIC acknowledge all study participants that have been part of the project.

**EXCEED:** Recruitment was supported by the EXtended Cohort for E-health, Environment and DNA (EXCEED) study. EXCEED is supported by the University of Leicester, the National Institute for Health and Care Research Leicester Respiratory Biomedical Research Centre, the Wellcome Trust (WT 202849), and Cohort Access fees from studies funded by the Medical Research Council (MRC), Biotechnology and Biological Sciences Research Council, National Institute for Health and Care Research, the UK Space Agency, and GlaxoSmithKline. It was previously supported by Medical Research Council grant G0902313. EXCEED is supported by BREATHE - The Health Data Research Hub for Respiratory Health (UKRI_PC_19004) in partnership with SAIL Databank. BREATHE is funded through the UK Research and Innovation Industrial Strategy Challenge Fund and delivered through Health Data Research UK.

**FHS:** The Framingham Heart Study is conducted and supported by the National Heart, Lung, and Blood Institute (NHLBI) in collaboration with Boston University (Contract No. N01-HC-25195, HHSN268201500001I and 75N92019D00031).

**Generation Scotland:** Received support from Chief Scientist Office of the Scottish Government Health Directorate (grant: CZD/16/6), the Scottish Funding Council (grant: HR03006), the Medical Research Council UK, and the Wellcome Trust Strategic Award 'STratifying Resilience and Depression Longitudinally' (STRADL) (grant: 104036/Z/14/Z). Genotyping of the GS:SFHS samples was carried out by the Edinburgh Clinical Research Facility, University of Edinburgh. Acknowledge all the families who took part in the Generation Scotland: Scottish Family Health Study, the general practitioners and Scottish School of Primary Care for their help in recruiting them, and the whole Generation Scotland team, which includes academic researchers, IT staff, laboratory technicians, statisticians and research managers.

**H2000:** Finnish Foundation for Cardiovascular Research (funding to Veikko Salomaa).

**HCHS/SOL:** The Hispanic Community Health Study/Study of Latinos is a collaborative study supported by contracts from the National Heart, Lung, and Blood Institute (NHLBI) to the University of North Carolina (HHSN268201300001I / N01-HC-65233), University of Miami (HHSN268201300004I / N01-HC-65234), Albert Einstein College of Medicine (HHSN268201300002I / N01-HC-65235), University of Illinois at Chicago (HHSN268201300003I / N01- HC-65236 Northwestern Univ), and San Diego State University (HHSN268201300005I / N01-HC-65237). The following Institutes/Centers/Offices have contributed to the HCHS/SOL through a transfer of funds to the NHLBI: National Institute on Minority Health and Health Disparities, National Institute on Deafness and Other Communication Disorders, National Institute of Dental and Craniofacial Research, National Institute of Diabetes and Digestive and Kidney Diseases, National Institute of Neurological Disorders and Stroke, NIH Institution-Office of Dietary Supplements. The Genetic Analysis Center at the University of Washington was supported by NHLBI and NIDCR contracts (HHSN268201300005C AM03 and MOD03).

**HUNT:** The Trøndelag Health Study (HUNT) is a collaboration between HUNT Research Centre (Faculty of Medicine and Health Sciences, NTNU, Norwegian University of Science and Technology), Trøndelag County Council, Central Norway Regional Health Authority, and the Norwegian Institute of Public Health. The genotyping in HUNT was financed by the National Institutes of Health and Care Research; University of Michigan; the Research Council of Norway; the Liaison Committee for Education, Research and Innovation in Central Norway; and the Joint Research Committee between St Olavs hospital and the Faculty of Medicine and Health Sciences, NTNU. The genetic investigations of the HUNT Study is a collaboration between researchers from the K.G. Jebsen Center for Genetic Epidemiology, NTNU and the University of Michigan Medical School and the University of Michigan School of Public Health. The K.G. Jebsen Center for Genetic Epidemiology is financed by Stiftelsen Kristian Gerhard Jebsen; Faculty of Medicine and Health Sciences, NTNU, Norway.

**KORA F4 and KORA S3:** The KORA study was initiated and financed by the Helmholtz Zentrum München – German Research Center for Environmental Health, which is funded by the German Federal Ministry of Education and Research (BMBF) and by the State of Bavaria. Furthermore, KORA research was supported within the Munich Center of Health Sciences (MC-Health), Ludwig-Maximilians-Universität, as part of LMUinnovativ and by the Competence Network Asthma and COPD (ASCONET), network COSYCONET (subproject 2, BMBF FKZ 01GI0882) funded by the German Federal Ministry of Education and Research (BMBF). This work was further supported by the German Center for Lung Research (DZL), grant numbers 82DZL083B2 and 82DZLI05A2.

**LBC1936:** Phenotype collection in the Lothian Birth Cohort 1936 was supported by Age UK (The Disconnected Mind project) and the University of Edinburgh. Genotyping was funded by the Biotechnology and Biological Sciences Research Council (BBSRC -BB/F019394/1).

**MESA:** MESA and the MESA SHARe project are conducted and supported by the National Heart, Lung, and Blood Institute (NHLBI) in collaboration with MESA investigators. The MESA Lung Study is supported by R01-HL077612 and R01-HL093081. Support for MESA is provided by contracts 75N92020D00001, HHSN268201500003I, N01-HC-95159, 75N92020D00005, N01-HC-95160, 75N92020D00002, N01-HC-95161, 75N92020D00003, N01-HC-95162, 75N92020D00006, N01-HC-95163, 75N92020D00004, N01-HC-95164, 75N92020D00007, N01-HC-95165, N01-HC-95166, N01-HC-95167, N01-HC-95168, N01-HC-95169, UL1-TR-000040, UL1-TR-001079, UL1-TR-001420, UL1-TR-001881, and DK063491. Funding for SHARe genotyping was provided by NHLBI Contract N02-HL-64278. MESA Family is conducted and supported by the National Heart, Lung, and Blood Institute (NHLBI) in collaboration with MESA investigators. Support is provided by grants and contracts R01HL071051, R01HL071205, R01HL071250, R01HL071251, R01HL071258, R01HL071259, by the National Center for Research Resources, Grant UL1RR033176, and the National Center for Advancing Translational Sciences, Grant UL1TR001881. This publication was developed under Science to Achieve Results (STAR) research assistance agreements, No. RD831697 (MESA Air), and RD-83830001 (MESA Air Next Stage), awarded by the U.S Environmental protection Agency. It has not been formally reviewed by the EPA. The views expressed in this document are solely those of the authors and the EPA does not endorse any products or commercial services mentioned in this publication. Genotyping was performed at Affymetrix (Santa Clara, California, USA) and the Broad Institute of Harvard and MIT (Boston, Massachusetts, USA) using the Affymetrix Genome-Wide Human SNP Array 6.0. MESA is also supported by the NHLBI, grant: R01-HL-131565 (funding to Ani Manichaikul).

**NEO**: Received support from the Netherlands Cardiovascular Research Initiative, grant number: CVON2014-02 ENERGISE (funding to Dennis Mook-Kanamori),VELUX Stiftung [grant number 1156] (funding to Dennis Mook-Kanamori), and the Dutch Science Organization, grant number: ZonMW-VENI grant 916.14.023 (funding to Dennis Mook-Kanamori). The authors of the NEO study thank all individuals who participated in the Netherlands Epidemiology in Obesity study, all participating general practitioners for inviting eligible participants and all research nurses for collection of the data. We thank the NEO study group, Pat van Beelen, Petra Noordijk and Ingeborg de Jonge for the coordination, lab and data management of the NEO study. The genotyping in the NEO study was supported by the Centre National de Génotypage (Paris, France), headed by Jean-Francois Deleuze. The NEO study is supported by the participating Departments, the Division and the Board of Directors of the Leiden University Medical Center, and by the Leiden University, Research Profile Area Vascular and Regenerative Medicine. Tariq Faquih was supported by the King Abdullah Scholarship Program and King Faisal Specialist Hospital & Research Centre (No. 1012879283).

**NFBC1966 and NFBC1986:** NFBC1966 and 1986 have received financial support from the Academy of Finland (project grants 104781, 120315, 129269, 1114194, 24300796, Center of Excellence in Complex Disease Genetics and SALVE), University Hospital Oulu, Biocenter, University of Oulu, Finland (75617), NIHM (MH063706, Smalley and Jarvelin), Juselius Foundation, NHLBI grant 5R01HL087679-02 through the STAMPEED program (1RL1MH083268-01), NIH/NIMH (5R01MH63706:02), the European Commission (EURO-BLCS, Framework 5 award QLG1-CT-2000-01643), ENGAGE project and grant agreement HEALTH-F4-2007-201413, EU FP7 EurHEALTHAgeing -277849, the Medical Research Council, UK (G0500539, G0600705, G1002319, PrevMetSyn/SALVE) and the MRC, Centenary Early Career Award. The program is currently being funded by the H2020 DynaHEALTH action (grant agreement 633595), EU H2020-HCO-2004 iHEALTH Action (grant agreement 643774), EU H2020-PHC-2014 ALEC Action (grant agreement No. 633212), EU H2020-SC1-2016-2017 LifeCycle Action (grant agreement No 733206), EU H2020-MSCA-ITN-2016 CAPICE Action (grant agreement 721567), Academy of Finland EGEA-project (285547) and MRC Grant nro MR/M013138/1.

**ORCADES:** Supported by the Chief Scientist Office of the Scottish Government (CZB/4/276, CZB/4/710), the Royal Society, the MRC Human Genetics Unit, Arthritis Research UK and the European Union framework program 6 EUROSPAN project (contract no. LSHG-CT-2006-018947) (funding to James F. Wilson). ORCADES DNA extractions were performed at the Wellcome Trust Clinical Research Facility in Edinburgh. ORCADES acknowledge the people of Orkney for sharing their data and samples.

**PIVUS:** Received support from Versus Arthritis, grant number: 21754 (funding to Andrew Morris), the Wellcome Trust under awards WT098017, WT064890, WT090532, Uppsala University and Uppsala University Hospital, the Swedish Research Council and the Swedish Heart-Lung Foundation. The PIVUS investigators express their deepest gratitude to the study participants.

**QBB:** Received funding from Qatar Foundation (QF) by the Biomedical Research Program at Weill Cornell Medicine in Qatar, grant: NPRP11C-0115-180010 (funding to Karsten Suhre and Omar Albagha) Qatar biobank and Qatar Genome Program are both Research, Development & Innovation entities within Qatar Foundation for Education, Science and Community Development.

**Raine Study**: Funded by National Health and Medical Research Council (NHMRC) of Australia, grants: 572613, 403981 and 1059711, and the Canadian Institutes of Health Research – CIHR (MOP-82893). The authors are grateful to the Raine Study participants and their families and we thank the Raine Study and Lions Eye Institute research staff for cohort coordination and data collection. The authors gratefully acknowledge the NHMRC for their long term funding to the study over the last 30 years and also the following institutes for providing funding for Core Management of the Raine Study: The University of Western Australia (UWA) , Curtin University, the Raine Medical Research Foundation, Telethon Kids Institute, Women's and Infant's Research Foundation, Murdoch University, The University of Notre Dame (Australia), and Edith Cowan University. The authors gratefully acknowledge the assistance of the Western Australian DNA Bank (National Health and Medical Research Council of Australia National Enabling

Facility). This work was supported by resources provided by the Pawsey Supercomputing Centre with funding from the Australian Government and Government of Western Australia.

**Rotterdam:** Funded by FWO, grant: 3G037618 (funding to Sara Wijnant). The Rotterdam Study is funded by Erasmus Medical Center and Erasmus University, Rotterdam, Netherlands Organization for the Health Research and Development (ZonMw), the Research Institute for Diseases in the Elderly (RIDE), the Ministry of Education, Culture and Science, the Ministry for Health, Welfare and Sports, the European Commission (DG XII), and the Municipality of Rotterdam. The generation and management of genetic data for the Rotterdam Study (RS I, RS II, RS III) was executed by the Human Genotyping Facility of the Genetic Laboratory of the Department of Internal Medicine, Erasmus MC, Rotterdam, The Netherlands, with the support of Netherlands Organisation of Scientific Research NWO Investments (nr. 175.010.2005.011, 911-03-012), the Genetic Laboratory of the Department of Internal Medicine, Erasmus MC, the Research Institute for Diseases in the Elderly (014-93-015; RIDE2), the Netherlands Genomics Initiative (NGI)/Netherlands Organisation for Scientific Research (NWO) Netherlands Consortium for Healthy Aging (NCHA), project nr. 050-060-810. The generation and management of lung disease and spirometric data was supported by FWO project G035014N.

**SAPALDIA:** Swiss National Science Foundation (33CS30-148470/1&2, 33CSCO-134276/1, SPIROMICS 33CSCO-108796, 324730_135673, 3247BO-104283, 3247BO-104288, 3247BO-104284, 3247-065896, 3100-059302, 3200-052720, 3200-042532, 4026-028099, PMPDP3_129021/1, PMPDP3_141671/1), the Federal Office for the Environment, the Federal Office of Public Health, the Federal Office of Roads and Transport, the canton's government of Aargau, Basel-Stadt, Basel-Land, Geneva, Luzern, Ticino, Valais, and Zürich, the Swiss Lung League, the canton's Lung League of Basel Stadt/ Basel Landschaft, Geneva, Ticino, Valais, Graubünden and Zurich, Stiftung ehemals Bündner Heilstätten, SUVA, Freiwillige Akademische Gesellschaft, UBS Wealth Foundation, Talecris Biotherapeutics GmbH, Abbott Diagnostics, European Commission 018996 (GABRIEL) and the Wellcome Trust (WT 084703MA). The SAPALDIA study could not have been done without the help of the study participants, technical and administrative support and the medical teams and field workers at the local study sites. Local fieldworkers : Aarau: S Brun, G Giger, M Sperisen, M Stahel, Basel: C Bürli, C Dahler, N Oertli, I Harreh, F Karrer, G Novicic, N Wyttenbacher, Davos: A Saner, P Senn, R Winzeler, Geneva: F Bonfils, B Blicharz, C Landolt, J Rochat, Lugano: S Boccia, E Gehrig, MT Mandia, G Solari, B Viscardi, Montana: AP Bieri, C Darioly, M Maire, Payerne: F Ding, P Danieli A Vonnez, Wald: D Bodmer, E Hochstrasser, R Kunz, C Meier, J Rakic, U Schafroth, A Walder.

**SHIP**: Received support from the Federal Ministry of Education and Research, grants: 01ZZ9603, 01ZZ0103, 01ZZ0403 and 03ZIK012, and the German Research Foundation, grant: GR 1912/5-1.

**SPIROMICS**: Supported by contracts from the NIH/NHLBI (HHSN268200900013C, HHSN268200900014C, HHSN268200900015C, HHSN268200900016C, HHSN268200900017C, HHSN268200900018C, HHSN268200900019C, HHSN268200900020C), grants from the NIH/NHLBI (U01 HL137880 and U24 HL141762), and supplemented by contributions made through the Foundation for the NIH and the COPD Foundation from AstraZeneca/MedImmune; Bayer; Bellerophon Therapeutics; Boehringer-Ingelheim Pharmaceuticals, Inc.; Chiesi Farmaceutici S.p.A.; Forest Research Institute, Inc.; GlaxoSmithKline; Grifols Therapeutics, Inc.; Ikaria, Inc.; Novartis Pharmaceuticals Corporation; Nycomed GmbH; ProterixBio; Regeneron Pharmaceuticals, Inc.; Sanofi; Sunovion; Takeda Pharmaceutical Company; and Theravance Biopharma and Mylan. The NHLBI also supported SPIROMICS, grants: R01-HL-131565 (funding to Xiaowei Hu and Ani Manichaikul) and R01-HL-153248 (funding to Ani Manichaikul). The authors thank the SPIROMICS participants and participating physicians, investigators and staff for making this research possible. More information about the study and how to access SPIROMICS data is available at www.spiromics.org. The authors would like to acknowledge the University of North Carolina at Chapel Hill BioSpecimen Processing Facility for sample processing, storage, and sample disbursements (http://bsp.web.unc.edu/). We would like to acknowledge the following current and former investigators of the SPIROMICS sites and reading centers: Neil E Alexis, MD; Wayne H Anderson, PhD; Mehrdad Arjomandi, MD; Igor Barjaktarevic, MD, PhD; R Graham Barr, MD, DrPH; Patricia Basta, PhD; Lori A Bateman, MSc; Surya P Bhatt, MD; Eugene R Bleecker, MD; Richard C Boucher, MD; Russell P Bowler, MD, PhD; Stephanie A Christenson, MD; Alejandro P Comellas, MD; Christopher B Cooper, MD, PhD; David J Couper, PhD; Gerard J Criner, MD; Ronald G Crystal, MD; Jeffrey L Curtis, MD; Claire M Doerschuk, MD; Mark T Dransfield, MD; Brad Drummond, MD; Christine M Freeman, PhD; Craig Galban, PhD; MeiLan K Han, MD, MS; Nadia N Hansel, MD, MPH; Annette T Hastie, PhD; Eric A Hoffman, PhD; Yvonne Huang, MD; Robert J Kaner, MD; Richard E Kanner, MD; Eric C Kleerup, MD; Jerry A Krishnan, MD, PhD; Lisa M LaVange, PhD; Stephen C Lazarus, MD; Fernando J Martinez, MD, MS; Deborah A Meyers, PhD; Wendy C Moore, MD; John D Newell Jr, MD; Robert Paine, III, MD; Laura Paulin, MD, MHS; Stephen P Peters, MD, PhD; Cheryl Pirozzi, MD; Nirupama Putcha, MD, MHS; Elizabeth C Oelsner, MD, MPH; Wanda K O'Neal, PhD; Victor E Ortega, MD, PhD; Sanjeev Raman, MBBS, MD; Stephen I. Rennard, MD; Donald P Tashkin, MD; J Michael Wells, MD; Robert A

Wise, MD; and Prescott G Woodruff, MD, MPH. The project officers from the Lung Division of the National Heart, Lung, and Blood Institute were Lisa Postow, PhD, and Lisa Viviano, BSN. Variant discovery and genotype calling was performed jointly, across TOPMed studies, for all samples in a given freeze using the GotCloud pipeline. This procedure results in a single, multi-study genotype call set. A support vector machine quality filter for variant sites was trained using a large set of site-specific quality metrics and known variants from arrays and the 1000 Genomes Project as positive controls and variants with Mendelian inconsistencies in multiple families as negative controls (see online documentation80 for more details). After removing all sites with a minor allele count less than 2, the genotypes with a minimal depth of more than 10× were phased using Eagle 2.481. Sample-level quality control included checks for pedigree errors, discrepancies between self-reported and genetic sex, and concordance with previous genotyping array data. Any errors detected were addressed before dbGaP submission. Details regarding Quality control on the TOPmed WGS was performed using SVM-based filter. Data acquisition, processing and quality control vary among the TOPMed data freezes. Freeze-specific methods are described on the TOPMed website (https://www.nhlbiwgs.org/data-sets).

**YFS:** The Young Finns Study has been financially supported by the Academy of Finland: grants 286284, 134309 (Eye), 126925, 121584, 124282, 129378 (Salve), 117787 (Gendi), and 41071 (Skidi); the Social Insurance Institution of Finland; Competitive State Research Financing of the Expert Responsibility area of Kuopio, Tampere and Turku University Hospitals (grant X51001); Juho Vainio Foundation; Paavo Nurmi Foundation; Finnish Foundation for Cardiovascular Research ; Finnish Cultural Foundation; Tampere Tuberculosis Foundation; Emil Aaltonen Foundation; Yrjö Jahnsson Foundation; Signe and Ane Gyllenberg Foundation; Diabetes Research Foundation of Finnish Diabetes Association; and EU Horizon 2020 (grant 755320 for TAXINOMISIS).

**TOPMed:** Molecular data for the Trans-Omics in Precision Medicine (TOPMed) program was supported by the National Heart, Lung and Blood Institute (NHLBI). Genome Sequencing for "NHLBI TOPMed: SubPopulations and InteRmediate Outcome Measures In COPD Study (SPIROMICS)" (phs001927) was performed at Broad Genomics (HHSN268201600034I). Core support including centralized genomic read mapping and genotype calling, along with variant quality metrics and filtering were provided by the TOPMed Informatics Research Center (3R01HL-117626-02S1; contract HHSN268201800002I). Core support including phenotype harmonization, data management, sample-identity QC, and general program coordination were provided by the TOPMed Data Coordinating Center (R01HL-120393; U01HL-120393; contract HHSN268201800001I). We gratefully acknowledge the studies and participants who provided biological samples and data for TOPMed.

**TwinsUK**: Received support from Fight for Sight, grant: Ref 5037/5038 (funding to Karina Patasova).

**UKHLS:** The UK Household Longitudinal Study was funded by grants from the Economic and Social Research Council (ES/H029745/1) and the Wellcome Trust (WT098051). These data are from Understanding Society: The UK Household Longitudinal Study, which is led by the Institute for Social and Economic Research at the University of Essex and funded by the Economic and Social Research Council. The data were collected by NatCen and the genome wide scan data were analysed by the Wellcome Trust Sanger Institute. The Understanding Society DAC have an application system for genetics data and all use of the data should be approved by them. The application form is at: https://www.understandingsociety.ac.uk/about/health/data. Full list of authors: Michaela Benzeval(1), Jonathan Burton(1), Nicholas Buck(1), Annette Jäckle(1), Meena Kumari(1), Heather Laurie(1), Peter Lynn(1), Stephen Pudney(1), Birgitta Rabe(1), Dieter Wolke(2): 1) Institute for Social and Economic Research; 2) University of Warwick.

**VIKING:** Supported by a MRC Human Genetics Unit quinquennial programme grant “QTL in Health and Disease”. VIKING DNA extractions and genotyping were performed at the Edinburgh Clinical Research Facility, University of Edinburgh.

**UBC lung eQTL resource:** The authors acknowledge Ma’en Obeidat of the University of British Columbia, Canada and Yohan Bossé and the eQTL committee at Laval University, Canada for facilitating access to the UBC lung eQTL data set.

**INTERVAL pQTL resource:** Participants in the INTERVAL randomised controlled trial were recruited with the active collaboration of NHS Blood and Transplant England (www.nhsbt.nhs.uk), which has supported field work and other elements of the trial. DNA extraction and genotyping was co-funded by the National Institute for Health Research (NIHR), the NIHR BioResource (http://bioresource.nihr.ac.uk) and the NIHR [Cambridge Biomedical Research Centre at the Cambridge University Hospitals NHS Foundation Trust] [*]. The INTERVAL study was funded by NHSBT (11-01-GEN). The academic coordinating centre for INTERVAL was supported by core funding from: NIHR Blood and Transplant Research Unit in Donor Health and Genomics (NIHR BTRU-2014-10024), UK Medical Research Council (MR/L003120/1), British Heart Foundation (SP/09/002; RG/13/13/30194; RG/18/13/33946) and the NIHR [Cambridge Biomedical Research Centre at the Cambridge University Hospitals NHS Foundation Trust] [*]. Proteomic assays were funded by the academic coordinating centre for INTERVAL and MRL, Merck & Co., Inc. A complete list of the investigators and contributors to the INTERVAL trial is provided in reference[1]. The academic coordinating centre would like to thank blood donor centre staff and blood donors for participating in the INTERVAL trial.

This work was supported by Health Data Research UK, which is funded by the UK Medical Research Council, Engineering and Physical Sciences Research Council, Economic and Social Research Council, Department of Health and Social Care (England), Chief Scientist Office of the Scottish Government Health and Social Care Directorates, Health and Social Care Research and Development Division (Welsh Government), Public Health Agency (Northern Ireland), British Heart Foundation and Wellcome.

*The views expressed are those of the authors and not necessarily those of the NHS, the NIHR or the Department of Health and Social Care.

#### Declaration of Interests

M. D. Tobin and L. V. Wain receive funding from GSK for collaborative research projects outside of the submitted work.

I. P. Hall has funded research collaborations with GSK, Boehringer Ingelheim and Orion.

M. H. Cho has received grant funding from GSK and Bayer, and speaking or consulting fees from AstraZeneca, Illumina, and Genentech.

I. Sayers has funded research collaborations with GSK, Boehringer Ingelheim and Orion outside of the submitted work.

### Cohort details

#### UK Biobank

The UK Biobank genetic and phenotypic data were obtained under UK Biobank Application 648[2].

#### SpiroMeta consortium

This section provides study descriptions for the cohorts contributing to the SpiroMeta consortium. All participants provided written informed consent and studies were approved by local Research Ethics Committees and/or Institutional Review boards.

**ALSPAC:** The Avon Longitudinal Study of Parents and Children is a prospective birth cohort study that recruited pregnant women from 1990 to 1992 in the South West of England[3,4]. Data was collected since the early pregnancy of these women and their partners. Children offspring to these mothers have been followed-up regularly through clinical assessments and questionnaires. Our study used spirometry data from these children aged between 8 and 9.

Pregnant women resident in Avon, UK with expected dates of delivery 1st April 1991 to 31st December 1992 were invited to take part in the study. The initial number of pregnancies enrolled is 14,541 (for these at least one questionnaire has been returned or a "Children in Focus" clinic had been attended by 19/07/99). Of these initial pregnancies, there was a total of 14,676 foetuses, resulting in 14,062 live births and 13,988 children who were alive at 1 year of age.

When the oldest children were approximately 7 years of age, an attempt was made to bolster the initial sample with eligible cases who had failed to join the study originally. As a result, when considering variables collected from the age of seven onwards (and potentially abstracted from obstetric notes) there are data available for more than the 14,541 pregnancies mentioned above. The number of new pregnancies not in the initial sample (known as Phase I enrolment) that are currently represented on the built files and reflecting enrolment status at the age of 24 is 913 (456, 262 and 195 recruited during Phases II, III and IV respectively), resulting in an additional 913 children being enrolled. The phases of enrolment are described in more detail in the cohort profile paper and its update. The total sample size for analyses using any data collected after the age of seven is therefore 15,454 pregnancies, resulting in 15,589 foetuses. Of these 14,901 were alive at 1 year of age.

A 10% sample of the ALSPAC cohort, known as the Children in Focus (CiF) group, attended clinics at the University of Bristol at various time intervals between 4 to 61 months of age. The CiF group were chosen at random from the last 6 months of ALSPAC births (1432 families attended at least one clinic). Excluded were those mothers who had moved out of the area or were lost to follow-up, and those partaking in another study of infant development in Avon.

Please note that the study website contains details of all the data that is available through a fully searchable data dictionary and variable search tool" and reference the following webpage:

http://www.bristol.ac.uk/alspac/researchers/our-data/

Ethical approval for the study was obtained from the ALSPAC Ethics and Law Committee and the Local Research Ethics Committees. Informed consent for the use of data collected via questionnaires and clinics was obtained from participants following the recommendations of the ALSPAC Ethics and Law Committee at the time. The study was approved by the IRB at National Institute of Environmental Health Sciences (NIEHS).

We are extremely grateful to all the families who took part in this study, the midwives for their help in recruiting them, and the whole ALSPAC team, which includes interviewers, computer and laboratory technicians, clerical workers, research scientists, volunteers, managers, receptionists and nurses.

Supported by the MRC Programme Grant MR/S025340/1 and the Wellcome Trust Strategic Award (108,818/15/Z). The UK Medical Research Council and Wellcome (Grant ref: 217065/Z/19/Z) and the University of Bristol provide core support for ALSPAC. This publication is the work of the authors and Nick Timpson and Raquel Granell will serve as guarantors for the contents of this paper. A comprehensive list of grants funding is available on the ALSPAC website (http://www.bristol.ac.uk/alspac/external/documents/grant-acknowledgements.pdf . GWAS data was generated by Sample Logistics and Genotyping Facilities at Wellcome Sanger Institute and LabCorp (Laboratory Corporation of America) using support from 23andMe.

**FinnTwin:** The Finnish Twin Cohort (FTC) sample originates from two subsamples. The first one is the Finnish Twin Study of Ageing (FITSA) sub-study. Participants were recruited from the older Finnish Twin Cohort for a clinical study of functional limitations in older women. Clinical assessment was conducted in 2000-2001 at the University of

Jyväskylä. Lung function was measured in the standing position using an electronic spirometer (Medikro 202, Kuopio, Finland). A blood sample was taken for DNA extraction and genotyping. For details, see [5]. The second sample comes from the TwinFat study, which sampled twin pairs discordant and concordant for obesity. Potential twin pairs were identified from the questionnaire responses to weight and height in FTC. Pairs with comorbidities were excluded. As part of an extensive clinical assessment, lung function was measured by spiroergometry [6].

The study was approved by the local ethics committees (University Hospitals of Helsinki, Turku, Tampere, Kuopio and Oulu) and was conducted following the guidelines of the Declaration of Helsinki. All participants gave their written informed consent.

**HUNT:** The Trøndelag Health Study (the HUNT Study) is a population-based cohort study of the adult population in Trøndelag County, Norway. The study has been running in the northern part of the county, Nord-Trøndelag, since 1984 and is designed to cover a broad range of health-related topics through repeated surveys with questionnaires, interviews, clinical examinations, laboratory measurements and storage of biological samples[7]. In addition blood samples from HUNT2, 3 and 4 are genotyped using Illumina HumanCoreExome arrays including 600.000 single nucleotide polymorphisms (SNPs); with further imputation using a combined Haplotype Reference Consortium and locally sequenced reference panel into ~25 million SNPs for each participant[8]. The HUNT Study data can be linked to a wide range of local and national health registries by means of the unique identification number allocated to all Norwegian residents. All current residents ≥20 years of age in Nord-Trøndelag have been invited to each survey wave: the HUNT1 (1984-86, n= 77000, 89.4%), HUNT2 (1995-97, n=65 000 participants, 69.5%), HUNT3 (2006-08, 51 000 participants, 54.1%) and HUNT4 (2017-19, n=56 000,54%) surveys. Totally 11-15 000 participants were included in spirometry in HUNT2,3 and 4, these were either randomly (10%) selected or due to symptoms or diagnosis of obstructive lung disease. The Trøndelag Health Study (HUNT) is a collaboration between HUNT Research Centre (Faculty of Medicine and Health Sciences, Norwegian University of Science and Technology NTNU), Trøndelag County Council, Central Norway Regional Health Authority, and the Norwegian Institute of Public Health.

Regional Committees for Medical and Health Research Ethics (REK 2015/616) gave ethical approval for this work.

**Raine Study:** The Raine Study is a prospective pregnancy cohort of 2900 mothers recruited between 1989-1991 (https://www.rainestudy.org.au/) [9]. Recruitment took place at Western Australia's major perinatal centre, King Edward Memorial Hospital, and nearby private practices. Women who had sufficient English language skills, an expectation to deliver at King Edward Memorial Hospital, and an intention to reside in Western Australia to allow for future follow-up of their child were eligible for the study. The primary carers (Gen1) completed questionnaires regarding their respective study child, and the children (Gen2) had physical examinations at ages 1, 2, 3, 5, 8, 10, 14, 17, 18, 20, and 22. Spirometry was undertaken at the 5, 14 and 22 year follow-ups, with data from the 14-year follow-up used in this study. Ethics approval for the original pregnancy cohort and subsequent follow-ups were granted by the Human Research Ethics Committee of King Edward Memorial Hospital, Princess Margaret Hospital, the University of Western Australia, and the Health Department of Western Australia. Parents, guardians and young adult participants provided written informed consent either before enrolment or at data collection at each follow-up.

Ethics approval for the original pregnancy cohort and subsequent follow-ups were granted by the Human Research Ethics Committee of King Edward Memorial Hospital, Princess Margaret Hospital, the University of Western Australia, and the Health Department of Western Australia. Parents, guardians and young adult participants provided written informed consent either before enrolment or at data collection at each follow-up.

**Twins UK:** The TwinsUK adult twin registry comprises 14,010 twins from the general population [ref]. Twins were seen at the clinical facilities of the Department of Twin Research and Genetic Epidemiology, King's College London, where comprehensive phenotyping assessments, including lung function, were performed. The zygosity was ascertained from a standardised questionnaire that was completed by twins.

Twins provided informed written consent and the study was approved by St. Thomas' Hospital Research Ethics Committee (REC Ref: EC04/015).

Details of the **British 1958 Birth Cohort** biomedical follow-up have been previously reported[10] Spirometry at age 44–45 years was done in the standing position without nose clips, using a Vitalograph handheld spirometer as previously described[11]. In the analysis, all readings with a best-test variation greater than 10% were excluded.

The South East Research Ethics Committee of the (UK) National Health Service gave ethical approval for the British 1958 birth cohort (B58C) component of this study (REC references 01/1/044 and 06/MRE01/28).

The **Busselton Health Study** (BHS) is a longitudinal survey of the town of Busselton in the south-western region of Western Australia that began in 1966. In 1994/1995 a cross-sectional community follow-up study was undertaken where blood was taken for DNA extraction. A sample of 1,168 European-ancestry individuals were genotyped using the Illumina 610-Quad BeadChip (BHS1), and subsequent genotyping was carried out on an independent group of 3,428 European-ancestry individuals using Illumina 660W-Quad (BHS2). Spirometric measures of forced expired volume in one second ($FEV_1$) and forced vital capacity (FVC) were assessed.

Human Ethics committee of The University of Western Australia gave ethical approval for this work.

The **CROATIA** study was initiated to investigate the use of isolated rather than urban populations for the identification of genes associated with medically-relevant quantitative traits. Three cohorts have been recruited as part of the CROATIA study: **CROATIA-Vis[12], CROATIA-Korcula[13]** and **CROATIA-Split[14].** CROATIA-Vis was the first to be collected when 1,008 Croatians aged 18-93 recruited from the villages of Komiza and Vis on the Dalmatian island of Vis. Recruitment occurred from 2003 to 2004 with participants donating blood for DNA extraction and biochemical measurements as well as undergoing some anthropometric measurements and physiological tests to measure traits such as height, weight and blood pressure, and finally completing several questionnaires relating to general health, medical history, diet and lifestyle. CROATIA-Korcula was recruited from 2007 to 2008 from the town of Korcula and the villages of Lumbarda, Zrnovo and Racisce on the island of Korcula, Croatia with 969 adults aged 18-98 agreeing to participate. This study followed the same recruitment procedures as CROATIA-Vis and the same samples and tests were collected with a few additions to reflect the research interests and expertise in Edinburgh. Volunteers were recruited to be part of the CROATIA-Split cohort in 2009-2010 from the Dalmatian mainland city of Split. This is the main ferry port to the islands and is the second largest city in Croatia and the largest along the Dalmatian coast. 1,012 adults aged 18-85 were recruited using the same methodology and with the same samples collected as in CROATIA-Korcula.

Ethical approval was obtained from The Ethical board of the University of Split, School of Medicine, Croatia gave ethical approval for this work.

**European Prospective Investigation of Cancer (EPIC)-Norfolk** is an ongoing UK-based prospective cohort and part of the Europe-wide multi-centre EPIC study. Details of the study design were described previously[15]. Briefly, 25,639 men and women aged 40-79 in eastern England were recruited through general practice registers and underwent baseline assessment between 1993 and 1997. Participants were further invited to the follow-up assessment (1998 to 2000), and were followed up by 2009 for incident outcomes and by 2013 for mortality.

The Norfolk (UK) Research Ethics Committee gave ethical approval for this work.

The **Generation Scotland: Scottish Family Health Study** is a collaboration between the Scottish Universities and the NHS, funded by the Chief Scientist Office of the Scottish Government. GS:SFHS is a family-based genetic epidemiology cohort with DNA, other biological samples (serum, urine and cryopreserved whole blood) and socio-demographic and clinical data from ~24,000 volunteers, aged 18-98 years, in ~7,000 family groups. Participants were recruited across Scotland, with some family members from further afield, from 2006-2011. Most (87%) participants were born in Scotland and 96% in the UK or Ireland. The cohort profile has been published[16].

Ethical approval for the GS:SFHS study was received from the Tayside Committee on Medical Research Ethics (on behalf of the National Health Service), ref 05/S1401/89.

Generation Scotland obtained Research Tissue Bank approval from the East of Scotland Research Ethics Service (on behalf of NHS Scotland), ref 20/ES/0021. All participants gave written informed consent. Generation Scotland is a collaboration between the University Medical Schools and National Health Service in Aberdeen, Dundee, Edinburgh and Glasgow (UK).

The DNA archive established from the **Health 2000** Survey Cohort was used. Details of this study population and phenotyping procedures have been previously reported[17]. Genome-wide genotyping was available for 2124 individuals selected from the Health 2000 cohort as metabolic syndrome cases and their matched controls[18]. Spirometry was done in the standing position without nose clips, using a Vitalograph 2150 spirometer. In the analysis, the maximum permissible difference between the two highest $FEV_1$ and FVC values was 10%.

The Ethics Committee of the National Public Health Institute gave ethical approval for the Health 2000 project in Sept. 29, 1999. A later amendment has been approved by the Ethics Committee of the Hospital District of Helsinki and Uusimaa at the National Public Health Institute in May 31, 2000.

The KORA studies (Cooperative Health Research in the Region of Augsburg) are a series of independent population based studies from the general population living in the region of Augsburg, Southern Germany[19,20].

The **KORA S3** study including 4,856 individuals was conducted in 1994/95. Spirometry was measured during a follow up in 1997/98 for all participants younger than 60 years who did not smoke or use inhalers one hour before the test. All spirometric tests were performed strictly adhering to the ECRHS protocol[21,22] using Biomedin Spirometers (Biomedin srl, Padova, Italy). Tests were accounted valid if at least two technically satisfactory manoeuvres could be obtained throughout a maximum of nine trials. $FEV_1$ and FVC were defined as the maximum value within all valid manoeuvers. For KORA S3 participants without spirometry measurements in 1997/98 we used measurements from the KORA-Age time point conducted in 2008/09. KORA Age contains subjects from all KORA studies born until 1943 (aged 65–90 years)[23]. Spirometry was measured in 935 randomly selected participants. Conditions including the examiner were the same as in KORA F4 (see below) except that inhalation of salbutamol was not performed due to the high number of contraindications anticipated in this aged population.

The **KORA F4** study including 3,080 individuals was conducted from 2006-2008 as a follow-up study to KORA S4 (1999-2001). Lung function tests were performed in a random subsample of subjects born between 1946 and 1965 (age range 41–63 years). Spirometry was performed in line with the ATS/ERS recommendations[24] using a pneumotachograph-type spirometer (Masterscreen PC, CardinalHealth, Würzburg, Germany) before and after inhalation of 200µg salbutamol. The present study is based on maximum values of $FEV_1$ and FVC measured before bronchodilation. The spirometer was calibrated daily using a calibration pump (CardinalHealth, Würzburg, Germany), and additionally, an internal control was used to ensure constant instrumental conditions. For KORA F4 participants without spirometry measurements in 2006-2008, we used measurements from the KORA-Age time point conducted in 2008/09. KORA Age contains subjects from all KORA studies born until 1943 (aged 65-90 years)[23]. Spirometry was measured in 935 randomly selected participants. Conditions including the examiner were the same as in 2008/09 except that inhalation of salbutamol was not performed due to the high number of contraindications anticipated in this aged population.

The ethics committee of the Bavarian Chamber of Physicians (Munich, Germany) gave ethical approval for this work.

The **Lothian Birth Cohort 1936** consists of 1,091 relatively healthy individuals assessed on cognitive and medical traits at about 70 years of age. They were all born in 1936 and most took part in the Scottish Mental Survey of 1947. At baseline the sample of 548 men and 543 women had a mean age 69.6 years (s.d. = 0.8). They were all Caucasian, community-dwelling, and almost all lived in the Lothian region (Edinburgh city and surrounding area) of Scotland. A full description of participant recruitment and testing can be found elsewhere[25]. Genotyping was performed at the Wellcome Trust Clinical Research Facility, Edinburgh. Quality control measures were applied and 1,005 participants remained. Lung function assessing peak expiratory flow rate, forced expiratory volume in 1 second, and forced vital capacity (each the best of three), using a Micro Medical Spirometer was assessed, sitting down without nose clips, at age 70 years. The accuracy of the spirometer is ±3% (to ATS recommendations Standardisation of Spirometry 1994 update for flows and volumes).

Ethics permission for the Lothian Birth Cohort 1936 (LBC1936) was obtained from the Multi-Centre Research Ethics Committee for Scotland (MREC/01/0/56) and the Lothian Research Ethics Committee (LREC/2003/2/29).

The **Northern Finland Birth Cohort 1966 (NFBC1966)** is a prospective follow-up study of children from the two northernmost provinces of Finland born in 1966.[26]All individuals still living in northern Finland or the Helsinki area (n = 8,463) were contacted and invited for clinical examination. A total of 6007 participants attended the clinical examination at the participants' age of 31 years. DNA was extracted from blood samples given at the clinical examination (5,753 samples available)[27]. The subset with DNA is representative of the original cohort in terms of major environmental and social factors. Informed consent was obtained from all subjects. After performing standard sample QC we included 5,402 NFBC1966 participants that were genotyped on an Illumina HumanCNV370DUO Analysis BeadChip. 329,401 variants were included in the imputation scaffold. Variants were imputed to the HRC reference r1.1 2016 on the Michigan Imputation Server. Prior to analysis we excluded variants monomorphic in this dataset. In NFBC1966, we used a Vitalograph P-model spirometer (Vitalograph Ltd., Buckingham, UK), with a volumetric accuracy of ±2% or ±50 mL whichever was greater. The spirometer was calibrated regularly using a 1-Litre precision syringe. The spirometric manoeuvre was performed three times but was repeated if the coefficient of variation between two maximal readings was >4%.

The **Northern Finland Birth Cohort 1986 (NFBC1986)** consists of 99% of all children, who were born in the provinces of Oulu and Lapland in Northern Finland between 1 July 1985 and 30 June 1986. 9,203 live-born individuals entered the study[28]. At the age of 16, the subjects living in the original target area or in the capital area (n=9,215) were invited to participate in a follow-up study including a clinical examination. 7,344 participants attend the study in year 2001/2002, of which 5,654 completed the postal questionnaire, the clinical examination and provided a blood sample[29]. DNA was extracted from all 5,654 blood samples. An informed consent for the use of the data including

DNA was obtained from all subjects. After performing standard sample QC we included 3,743 NFBC1986 participants that were genotyped on an Illumina Human Omni Express Exome 8v1.2 BeadChip. 889,119 variants were included in the imputation scaffold. Variants were imputed to the HRC reference r1.1 2016 on the Michigan Imputation Server. For Spirometry measurements, we used a Vitalograph Gold Standard (Model 2150) (Vitalograph Ltd., Buckingham, UK). The machines were calibrated every day the medical examination took place. The spirometric manoeuvre was performed in an upright sitting position while wearing a nose clip. At least three acceptable manoeuvres were performed. Acceptable manoeuvers did not exceed a difference between two maximal FEV 1 and FVC values of 4 %. The results were recorded with a 0.05 litre accuracy.

The **Orkney Complex Disease Study** (ORCADES) is an ongoing family-based, cross-sectional study in the isolated Scottish archipelago of Orkney. Spirometry was performed in the sitting position without nose clips, using a Spida handheld spirometer. Measurements were repeated once and the better reading was used for analysis.

All ORCADES participants gave written informed consent and the study was approved by Research Ethics Committees in Orkney, Aberdeen (North of Scotland REC), and South East Scotland REC, NHS Lothian (reference: 12/SS/0151).

The **Prospective Investigation of the Vasculature in Uppsala Seniors** (PIVUS)[30] is a population-based study of cardiovascular health in the elderly. Mailed invitations were sent to subjects who lived in Uppsala, Sweden, within 2 months after their 70th birthday. The subjects were randomly selected from the community register. A total of 1,016 men and women participated in the baseline investigation (participation rate, 50.1%). Spirometry was performed in 901 subjects at baseline in accordance with American Thoracic Society recommendations (α spirometer; Vitalograph Ltd; Buckingham, UK). The best value from three recordings was used. Genotyping of all samples was undertaken using the Illumina Omni Express and CardioMetabochip. Genotypes were called using GENCALL. A total of 738,879 SNPs passed quality control (thresholds: call rate < 0.95, and call rate < 0.99 for MAF<5%; HWE $P$ < 10-6). SNPs with MAF<1% were removed from the imputation scaffold. Imputation was performed using IMPUTE up to haplotypes from the Haplotype Reference Consortium.

The Ethics Committee of the University of Uppsala approved the study, and the participants gave their informed consent.

The **SAPALDIA** cohort is a population-based multi-center study in eight geographic areas representing the range of environmental, meteorological and socio-demographic conditions in Switzerland[31]. It was initiated in 1991 (SAPALDIA 1) with a follow-up assessment in 2002 (SAPALDIA 2) and 2010 (SAPALDIA3). This study has specifically been designed to investigate longitudinally lung function, respiratory and cardiovascular health; to study and identify the associations of these health indicators with individual long term exposure to air pollution, other toxic inhalants, life style and molecular factors.

All procedures in this cohort study were conducted in accordance with the World Medical Association's Declaration of Helsinki and Declaration of Taipei. Written informed consent was obtained from the study participants prior to health examination and blood sample collection. The study protocols of this multi-centric long-term study with baseline and follow-up assessments were approved by Swiss national overarching ethics committees and by regional cantonal ethics committees for each time point of data collection. The SAPALDIA cohort study was approved by the ethics committee of the medical faculty of the University of Lausanne, Switzerland, for the baseline examination (SAPALDIA1) in 1989; and by the Supra-regional Ethics Committee for Clinical Research (UREK approval N° 123/00) of the Swiss Academy of Medical Sciences for the second examination (SAPALDIA2) in September 2001; and given the multi-centric design of the long-term cohort, by multiple cantonal ethics committees for the third examination (SAPALDIA3) in 2009 (ethics committee of the Department of Health and Social Affairs of Aargau, approval N° 2009/056, ethics committee of both Basel, approval N° 219/09, cantonal ethics committee of Zurich, approval N° 52/09, departmental ethics committee for Internal Medicine and Community Medicine of Geneva, approval N° 09-174, cantonal commission for Medical Ethics of Valais, approval N°033/09, cantonal Commission for Ethics in Human Research of Vaud, approval N° 200/09, cantonal Ethics Committee of Ticino, approval N° CE2276).

The **Study of Health In Pomerania (SHIP)**[32] is a cross-sectional and prospective longitudinal population-based cohort study in Western Pomerania assessing the prevalence and incidence of common diseases and their risk factors. SHIP encompasses the two independent cohorts **SHIP** and **SHIP-TREND**. A total of 4,308 participants were recruited between 1997 and 2001 in the SHIP cohort. Between 2008 and 2012 a total of 4,420 participants were recruited in the SHIP-TREND cohort. Individuals were invited to the SHIP study centre for a computer-assisted personal interviews and extensive physical examinations.

The examinations for **SHIP** were conducted using a body plethysmograph equipped with a pneumotachograph (VIASYS Healthcare, JAEGER, Hoechberg, Germany) which meets the American Thoracic Society (ATS) criteria.[33] The volume signal of the equipment was calibrated with a 3.0 litre syringe connected to the pneumotachograph in accordance with the manufacturer´s recommendations and at least once on each day´s testing. Barometric pressure, temperature and relative humidity were registered every morning. Calibration of reference gas and volume was examined under ATS-conditions (Ambient Temperature Pressure) and the integrated volumes were BTPS (Body Temperature Pressure Saturated) corrected[33,34]. Lung function variables were measured continuously throughout the baseline breathing and the forced manoeuvres using a VIASYS HEALTHCARE system (MasterScreen Body/Diff.). Spirometry flow volume loops were conducted in accordance with ATS recommendations[34] in a sitting position and with wearing nose clips. The participants performed at least three forced expiratory lung function manoeuvres in order to obtain a minimum of two acceptable and reproducible values.[35] Immediate on-screen error codes indicating the major acceptability (including start, duration and end of test) and reproducibility criteria supported the attempt for standardised procedures. The procedure was continuously monitored by a physician. The best results for FVC, FEV1, peak expiratory flow (PEF) and expiratory flow at 75%, 50%, 25% of FVC (MEF 75, MEF 50, MEF 25) were taken. The ratio of FEV1 to FVC was calculated from the largest FEV1 and FVC.

In terms of the pulmonary items the computer-assisted interview in **SHIP-TREND** was nearly identical to that of the SHIP. Of the 4.420 subjects who have been investigated in the study, 2.678 (60.6 %) of the subjects have undergone spirometry, body plethysmography, and measurements of diffusing capacity (CO and NO), IOS and respiratory muscle strength. In SHIP-TREND, the following additional methods that are of particular interest in terms of lung health and comorbidities have been applied: polysomnography, analysis of volatile compounds in the exhaled breath, and whole-body MRI. The following devices have been used for the pulmonary investigations in SHIP-TREND: a MasterScreen for body plethysmography, diffusing capacity measurements (single breath) and measurements of respiratory muscle strength (Viasys Healthcare, Hoechberg, Germany), an ABL 500 and later an ABL 80 for blood gas analyses (Radiometer, Copenhagen, Denmark), a MasterScreen PFT Pro CO-NO-Diffusion (CareFusion, Hoechberg, Germany), a MasterSreen IOS for Impuls-Oscillometry (CareFusion, Hoechberg, Germany), and a MicroCO carbon monoxide monitor (CareFusion, Hoechberg, Germany).

This work was reviewd by the Ethics Committee of the Faculty of Medicine at the Ernst-Moritz-Arndt-University of Greifswald. A trial plan adapted to the results of this consultation was received by the office of the Ethics Committee on 02nd of September 2009. The majority of the Ethics Committee found that there were no ethical or legal concerns about continuing the project as described or about including project extension. Therefore, the Ethics Committee grants approval for the proposed study. Reference number: BB 39/08a.

The **United Kingdom Household Longitudinal Study (UKHLS)**, also known as Understanding Society (https://www.understandingsociety.ac.uk) is a longitudinal panel survey of 40,000 UK households (England, Scotland, Wales and Northern Ireland) representative of the UK population. Participants are surveyed annually since 2009 and contribute information relating to their socioeconomic circumstances, attitudes, and behaviours via a computer assisted interview. The study includes phenotypical data for a representative sample of participants for a wide range of social and economic indicators as well as a biological sample collection encompassing biometric, physiological, biochemical, and haematological measurements and self-reported medical history and medication use.

For a subset of individuals who took part in a nurse health assessment, blood samples were taken and genomic DNA extracted. Of these, 10,484 samples were genotyped at the Wellcome Trust Sanger Institute using the Illumina Infinium HumanCoreExome-12 v1.0BeadChip.

Lung function measures in samples from England and Wales were conducted with the NDD Easy On-PC spirometer (NDD Medical Technologies, Zurich, Switzerland). Participants were excluded in the following cases: pregnancy, having had abdominal or chest surgery (past 3 weeks), admitted to the hospital with a heart complaint (in the past 6 weeks), having had recent eye surgery (past 4 weeks), or in case of having a tracheostomy. Subjects were asked to perform up to 8 blows that ideally lasted at least 6 seconds, uninterrupted by coughing, glottis closure, laughing or leakage of air. Upon completion, the measurements were rated either acceptable or unacceptable by the NDD Easy On-PC software.

The United Kingdom Household Longitudinal Study has been approved by the University of Essex Ethics Committee and informed consent was obtained from every participant.

The Viking Health Study - Shetland (**VIKING**) is a family-based, cross-sectional study that seeks to identify genetic factors influencing cardiovascular and other disease risk in the population isolate of the Shetland Isles in northern Scotland. Genetic diversity in this population is decreased compared to Mainland Scotland, consistent with the high levels of endogamy historically. Participants were recruited between 2013 and 2015, each having at least three grandparents from Shetland. Fasting blood samples were collected and over 300 health-related phenotypes and environmental exposures were measured in each individual.

All participants gave informed consent and the study was approved by the South East Scotland Research Ethics Committee.

The **Young Finns Study** (YFS) is a population-based follow up-study started in 1980[36]. The main aim of the YFS is to determine the contribution made by childhood lifestyle, biological and psychological measures to the risk of cardiovascular diseases in adulthood. In 1980, over 3,500 children and adolescents all around Finland participated in the baseline study. The follow-up studies have been conducted mainly with 3-year intervals. The latest 30-year follow-up study was conducted in 2010-2011 (ages 33-49 years) with 2,063 participants.

The Ethics Committee, Hospital District of Southwest Finland gave favourable ethical opinion of the protocol. All participants gave their written informed consent.

**The Extended Cohort for E-health, Environment and DNA (EXCEED)** was set up to develop understanding of the genetic, environmental and lifestyle-related causes of health and disease[37]. Participants were recruited primarily through local general practices in Leicester City, Leicestershire and Rutland. Baseline data collection included a lifestyle questionnaire, anthropometry measurements, and for approximately half of the participants, spirometry.

The original EXCEED study was approved by the Leicester Central Research Ethics Committee (Ref: 13/EM/0226). Substantial amendments have been approved by the same Research Ethics Committee for the collection of new data relating to the COVID-19 pandemic, including the COVID-19 questionnaires and antibody testing.

#### CHARGE consortium

**ALHS:** The Agricultural Lung Health Study (ALHS) is a case-control study of current asthma among farmers and their spouses nested within the prospective Agricultural Health Study (AHS data releases P3REL201209.00, PIREL201209.00 and AHSREL201304.00) [38 39 40]. Pulmonary function was measured by trained staff during in-home visits with an EasyOne® Spirometer (NDD Medical Technologies, Chelmsford, MA) based on American Thoracic Society guidelines. Genotyping based on the UK Biobank Axiom Array (Axiom_UKB_WCSG) by Affymetrix Axiom Genotyping Services (Affymetrix, Inc., Santa Clara, CA) was completed using DNA extracted from blood (96%) or saliva (4%) collected from ALHS participants during the home visit. Variants with missing rate>5% or Hardy-Weinberg $p<1x10^{-6}$ (if MAF>5%) were excluded. Participants with missing call rate>5%, IBS distance>0.9, or sex discrepancies were excluded. The current analysis includes 1,044 asthma cases and 1,804 non-asthma cases of European ancestry with genotyping data, pulmonary function measures and complete covariate information. Imputation was performed using the HRC release 1.1 (http://www.haplotype-reference-consortium.org/site) with the University of Michigan imputation server (https://imputationserver.sph.umich.edu/index.html). Linear regression was performed separately in cases and non-cases using RVTESTS (https://genome.sph.umich.edu/wiki/Rvtests).

Informed consent was obtained from all participants and the study was approved by the IRB at National Institute of Environmental Health Sciences (NIEHS).

**ARIC:** The ARIC study is a population-based prospective cohort study of cardiovascular disease sponsored by the National Heart, Lung, and Blood Institute (NHLBI). ARIC included 15,792 individuals, predominantly European American and African American, aged 45-64 years at baseline (1987-89), chosen by probability sampling from four US communities. Cohort members completed three additional triennial follow-up examinations, a fifth exam in 2011-2013, a sixth exam in 2016-2017, and a seventh exam in 2018-2019. The ARIC study has been described in detail previously (PMC8667593; Wright JD, Folsom AR, Coresh J, et al. The ARIC (Atherosclerosis Risk In Communities) Study: JACC Focus Seminar 3/8. J Am Coll Cardiol. 2021 Jun 15;77(23):2939-2959).

The ARIC study has been approved by Institutional Review Boards (IRB) at all participating institutions: University of North Carolina at Chapel Hill IRB, Johns Hopkins University IRB, University of Minnesota IRB, and University of Mississippi Medical Center IRB. Study participants provided written informed consent at all study visits.

**COPDGene & ECLIPSE (Boston):** The Mass General Brigham Institutional Review Board gave ethical approval for this work.

**CHS:** The Cardiovascular Health Study (CHS) is a population-based cohort study of risk factors for coronary heart disease and stroke in adults ≥65 years conducted across four field centers [41]. The original predominantly European ancestry cohort of 5,201 persons was recruited in 1989-1990 from random samples of the Medicare eligibility lists;

subsequently, an additional predominantly African-American cohort of 687 persons was enrolled in 1992-1993 for a total sample of 5,888. Blood samples were drawn from all participants at their baseline examination and DNA was subsequently extracted from available samples. Genotyping was performed at the General Clinical Research Center's Phenotyping/Genotyping Laboratory at Cedars-Sinai among CHS participants who consented to genetic testing and had DNA available using the Illumina 370CNV BeadChip system (for European ancestry participants, in 2007) or the Illumina HumanOmni1-Quad_v1 BeadChip system (for African-American participants, in 2010). The current analysis uses spirometry values obtained in 1989-1990 for the original cohort and 1993-1994 for the second cohort, which was completed in accordance with ATS recommendations.

Ethics committees/IRBs of Wake Forest University; University of California, Davis; Johns Hopkins University; University of Pittsburgh; University of Vermont; and University of Washington gave ethical approval for this work. Individuals in the present analysis had available DNA and gave informed consent including consent to use of genetic information for the study of cardiovascular disease.

**FHS:** The Framingham Heart Study is a U.S. family-based cohort that was established in 1948, in Framingham, MA (the Original cohort). Beginning in 1971, the Offspring cohort is comprised of children of the original cohort and spouses of these children. 4,095 adults were enrolled in the Gen3 cohort given at least one of their parents were in the Offspring cohort, starting in 2002. All cohorts continue under active surveillance for cardiovascular events. Spirometry from the multiple examinations with acceptable pulmonary function data for each sample was used, eligible examinations included the 13$^{th}$, 16$^{th}$, 17$^{th}$ and 19$^{th}$ examinations for the Original cohort, the 3$^{rd}$, 5$^{th}$, 6$^{th}$ and 7$^{th}$ examinations for the Offspring cohort, and the first examination for the Gen3 cohort. FHS participants were genotyped using the Affymetrix GeneChip Human Mapping 500K Array Set, which consisted of Nsp and Sty arrays, and another Affymetrix 50K gene centric array. Imputation was performed using Michigan Imputation Server based on the HRC reference panel release 1.1 April 2016 (HRC r1.1). This study included samples with spirometry and genotype data available.

All participants provided written informed consent and this study was approved by the Institutional Review Board of the Boston University Medical Campus.

**HCHS-SOL:** The Hispanic Community Health Study/Study of Latinos (dbGaP accession phs000810) is a community-based longitudinal cohort study of 16,415 self-identified Hispanic/Latino persons aged 18–74 years and selected from households in predefined census-block groups across four US field centers (in Chicago, Miami, the Bronx, and San Diego). The census-block groups were chosen to provide diversity among cohort participants with regard to socioeconomic status and national origin or background [42 43]. The HCHS/SOL cohort includes participants who self-identified as having a Hispanic/Latino background; the largest groups are Central American (n = 1,730), Cuban (n = 2,348), Dominican (n = 1,460), Mexican (n = 6,471), Puerto Rican (n = 2,728), and South American (n = 1,068). The HCHS/SOL baseline clinical examination occurred between 2008 and 2011 and included comprehensive biological, behavioral, and sociodemographic assessments. Spirometry was conducted in accordance with American Thoracic Society/European Respiratory Society guidelines [44] using a dry-rolling sealed spirometer, as previously described [45]. The study was approved by the Institutional Review Boards at each participating institution and written informed consent was obtained from all participants.

This study was approved by the institutional review boards (IRBs) at each field center, where all participants gave written informed consent, and by the Non-Biomedical IRB at the University of North Carolina at Chapel Hill, to the HCHS/SOL Data Coordinating Center. All IRBs approving the study are: Non-Biomedical IRB at the University of North Carolina at Chapel Hill. Chapel Hill, NC; Einstein IRB at the Albert Einstein College of Medicine of Yeshiva University. Bronx, NY; IRB at Office for the Protection of Research Subjects (OPRS), University of Illinois at Chicago. Chicago, IL; Human Subject Research Office, University of Miami. Miami, FL; Institutional Review Board of San Diego State University. San Diego, CA.

**MESA:** MESA is a longitudinal study of subclinical cardiovascular disease and risk factors that predict progression to clinically overt cardiovascular disease or progression of the subclinical disease [46]. Between 2000 and 2002, MESA recruited 6,814 men and women 45 to 84 years of age from Forsyth County, North Carolina; New York City; Baltimore; St. Paul, Minnesota; Chicago; and Los Angeles. Exclusion criteria were clinical cardiovascular disease, weight exceeding 136 kg (300 lb.), pregnancy, and impediment to long-term participation. The MESA Lung Study performed spirometry following the 2005 ATS/ERS guidelines in a subset of the MESA Study, as previously described [47]. For the current cross-sectional analysis, we used spirometry data obtained from MESA participants at Exam 4. Analyses were stratified by race/ethnic group for Non-Hispanic White, non-Hispanic African American, and Hispanic participants.

All participants provided informed consent and the protocols of MESA were approved by the IRBs of collaborating institutions and the National Heart, Lung and Blood Institute.

**NEO:** The NEO was designed for extensive phenotyping to investigate pathways that lead to obesity-related diseases. The NEO study is a population-based, prospective cohort study that includes 6,671 individuals aged 45–65 years, with an oversampling of individuals with overweight or obesity. At baseline, information on demography, lifestyle, and medical history have been collected by questionnaires. In addition, samples of 24-h urine, fasting and postprandial blood plasma and serum, and DNA were collected. Genotyping was performed using the Illumina HumanCoreExome chip, which was subsequently imputed to the 1000 genome reference panel. Participants underwent an extensive physical examination, including anthropometry, electrocardiography, spirometry, and measurement of the carotid artery intima-media thickness by ultrasonography. In random subsamples of participants, magnetic resonance imaging of abdominal fat, pulse wave velocity of the aorta, heart, and brain, magnetic resonance spectroscopy of the liver, indirect calorimetry, dual energy X-ray absorptiometry, or accelerometry measurements were performed. The collection of data started in September 2008 and completed at the end of September 2012. Participants are currently being followed for the incidence of obesity-related diseases and mortality.

The Medical Ethics committee of the Leiden University Medical Centre approved The Netherlands Epidemiology of Obesity (NEO) study under protocol P08.109. The NEO study is also registered at clinicaltrials.gov under number NL21981.058.08 / P08.109. All participants gave written informed consent.

**Rotterdam:** The Rotterdam Study is a population-based cohort study that started in 1990 in Rotterdam, the Netherlands, comprising almost 15 000 participants aged ≥45 years [48]. The Rotterdam Study aims to assess the occurrence of, and risk factors for, chronic diseases in the elderly. Every 4-5 years, participants undergo a home interview and clinical examinations at the research center. DNA was extracted from whole peripheral blood (stored in ethylenediamine tetraacetic acid (EDTA) tubes) by standardised salting out methods. Data processing was performed in the Genetic Laboratory of the Dept of Internal Medicine, Erasmus University Medical Centre, Rotterdam. Smoking status (never, former, current) and pack-years (years smoked multiplied by daily number of smoked cigarettes divided by 20) were assessed by interview.

The Rotterdam Study has been approved by the Medical Ethics Committee of the Erasmus MC (registration number MEC 02.1015) and by the Dutch Ministry of Health, Welfare and Sport (Population Screening Act WBO, license number 1071272-159521-PG). All participants provided written informed consent to participate in the study and to have their information obtained from treating physicians.

**SPIROMICS:** The SPIROMICS study was approved by the Institutional Review Boards at each of the cooperating institutions: Columbia University IRB 2 (AAAE9315), University of Iowa IRB-01 (201308719), Johns Hopkins IRB-5 (NA_00035701), UCLA Medical IRB 1 (MIRB1) (10001740/18-000403), University of Michigan IRBMED B1 Board (HUM00036346/ HUM00141222), National Jewish Health IRB (HS2678), UCSF IRB Parnassus Panel (10-03169), Temple University IRB A2 (21416), U of Alabama at Birmingham IRB #2 (120906004), University of Illinois IRB #3 (2013-0939), University of Utah IRB Panel Review Board 5 (00027298/ 00108836), Wake Forest University IRB #5 (00012805/00048727), UNC Non-Biomedical IRB (10-0048), UCLA Medical IRB 1 (MIRB1) (18-000458), and University of Iowa IRB-01 (201003733). All participants provided written informed consent. The research conformed to the principles of the Helsinki Declaration.

#### China Kadoorie Biobank (CKB)

China Kadoorie Biobank (CKB, https://www.ckbiobank.org) is a population-based prospective cohort of approximately 513,000 adults aged 30-79 recruited in 2004-8 from 10 geographically defined regions of China. Questionnaire and physical measurement data and biological samples were collected at baseline and at periodic re-surveys of 25,000 randomly-selected surviving participants. All participants are followed for cause-specific mortality and morbidity and for any hospital admission, through linkages to death and disease registries and to health insurance databases. Local, national and international ethics approval was obtained and all participants provided written informed consent. Genotyping was conducted on custom Affymetrix Axiom® arrays, with 100,706 unique samples and 511,885 variants passing QC. Genotypes were phased using SHAPEIT3 r882 and imputed into the 1000 Genomes Phase 3 reference with IMPUTE4 v4.r265. GWAS was conducted stratified by both sex and the 10 recruitment regions, using BOLT-LMM v2.3.4 with age and age2 as covariates. Subjects were excluded from analysis if they were classed during physical examination as "poor" for either subject's cooperation or data reliability. Due to systematic QC issues for the spirometry data from 2 recruitment regions (Qingdao and Haikou), analyses were restricted to 83,715 participants from the remaining 8 regions.

**Members of the China Kadoorie Biobank Collaborative Group**

**International Steering Committee:** Junshi Chen, Zhengming Chen (PI), Robert Clarke, Rory Collins, Yu Guo, Liming Li (PI), Chen Wang, Jun Lv, Richard Peto, Robin Walters.

**International Co-ordinating Centre, Oxford:** Daniel Avery, Derrick Bennett, Ruth Boxall, Ka Hung Chan, Yumei Chang, Yiping Chen, Zhengming Chen, Johnathan Clarke; Robert Clarke, Huaidong Du, Zammy Fairhurst-Hunter, Hannah Fry, Simon Gilbert, Alex Hacker, Mike Hill, Michael Holmes, Pek Kei Im, Andri Iona, Maria Kakkoura, Christiana Kartsonaki, Rene Kerosi, Kuang Lin, Mohsen Mazidi, Iona Millwood, Qunhua Nie, Alfred Pozarickij, Paul Ryder, Saredo Said, Sam Sansome, Dan Schmidt, Paul Sherliker, Rajani Sohoni, Becky Stevens, Iain Turnbull, Robin Walters, Lin Wang, Neil Wright, Ling Yang, Xiaoming Yang, Pang Yao.

**National Co-ordinating Centre, Beijing:** Yu Guo, Xiao Han, Can Hou, Chun Li, Chao Liu, Jun Lv, Pei Pei, Canqing Yu.

**Regional Co-ordinating Centres:**

**Gansu:** Gansu Provincial CDC – Caixia Dong, Pengfei Ge, Xiaolan Ren. Maiji CDC – Zhongxiao Li, Enke Mao, Tao Wang, Hui Zhang, Xi Zhang. **Haikou:** Hainan Provincial CDC – Jinyan Chen, Ximin Hu, Xiaohuan Wang. Meilan CDC – Zhendong Guo, Huimei Li, Yilei Li, Min Weng, Shukuan Wu. **Harbin:** Heilongjiang Provincial CDC – Shichun Yan, Mingyuan Zou, Xue Zhou. Nangang CDC – Ziyan Guo, Quan Kang, Yanjie Li, Bo Yu, Qinai Xu. **Henan:** Henan Provincial CDC – Liang Chang, Lei Fan, Shixian Feng, Ding Zhang, Gang Zhou. Huixian CDC – Yulian Gao, Tianyou He, Pan He, Chen Hu, Huarong Sun, Xukui Zhang. **Hunan:** Hunan Provincial CDC – Biyun Chen, Zhongxi Fu, Yuelong Huang, Huilin Liu, Qiaohua Xu, Li Yin. Liuyang CDC – Huajun Long, Xin Xu, Hao Zhang, Libo Zhang. **Liuzhou:** Guangxi Provincial CDC – Naying Chen, Duo Liu, Zhenzhu Tang. Liuzhou CDC – Ningyu Chen, Qilian Jiang, Jian Lan, Mingqiang Li, Yun Liu, Fanwen Meng, Jinhuai Meng, Rong Pan, Yulu Qin, Ping Wang, Sisi Wang, Liuping Wei, Liyuan Zhou. **Qingdao:** Qingdao CDC – Liang Cheng, Ranran Du, Ruqin Gao, Feifei Li, Shanpeng Li, Yongmei Liu, Feng Ning, Zengchang Pang, Xiaohui Sun, Xiaocao Tian, Shaojie Wang, Yaoming Zhai, Hua Zhang, Licang CDC – Wei Hou, Silu Lv, Junzheng Wang. **Sichuan:** Sichuan Provincial CDC – Xiaofang Chen, Xianping Wu, Ningmei Zhang, Weiwei Zhou. Pengzhou CDC – Xiaofang Chen, Jianguo Li, Jiaqiu Liu, Guojin Luo, Qiang Sun, Xunfu Zhong. **Suzhou:** Jiangsu Provincial CDC – Jian Su, Ran Tao, Ming Wu, Jie Yang, Jinyi Zhou, Yonglin Zhou. Suzhou CDC – Yihe Hu, Yujie Hua, Jianrong Jin Fang Liu, Jingchao Liu, Yan Lu, Liangcai Ma, Aiyu Tang, Jun Zhang. **Zhejiang**: Zhejiang Provincial CDC – Weiwei Gong, Ruying Hu, Hao Wang, Meng Wang, Min Yu. Tongxiang CDC – Lingli Chen, Qijun Gu, Dongxia Pan, Chunmei Wang, Kaixu Xie, Xiaoyi Zhang.

All participants provided written informed consent at each survey visit, allowing access to their medical records and long-term storage of biosamples for future unspecified medical research purposes, without any feedback of results to the individuals concerned. Ethical approval was obtained from the Oxford Tropical Research Ethics Committee, the Ethical Review Committees of the Chinese Centre for Disease Control and Prevention, Chinese Academy of Medical Sciences, and the Institutional Review Board (IRB) at Peking University. The Chinese Ministry of Health approved the study at the start in 2004 (including export of plasma samples to Oxford), and also approved electronic linkage to health insurance records in 2011. Raw genotyping data were exported from China to the Oxford CKB International Coordinating Centre under Data Export Approvals 2014-13 and 2015-39 from the Office of Chinese Human Genetic Resource Administration.

#### Qatar Biobank (QBB)

The Qatar genome program (QGP) is a population based study established in 2012, designed to perform whole genome sequencing of the Qatar Biobank (QBB) participants with the aim to gain insights into the population structure and the genetic architecture of clinically relevant phenotypes in the Middle Eastern Qatari population. The present study is based on whole genome sequence data from 6218 participants of QBB and further replication in 7768 subjects from the second batch of QBB data[49-51].

All participants gave informed consent. Institutional Review Board approval was obtained from the Hamad Medical Corporation Ethics Committee.

### Extended Methods

#### Assignment of UK Biobank ancestry groups

Previously k-means clustering was used on the first 2 principal components to assign UK Biobank samples to ancestry groups[52]. For this multi-ancestry study we required more tightly defined ancestry clustering with the possibility of those samples lying between clusters to be excluded instead of forcing them to belong to one cluster or another as with k-means clustering. We used ADMIXTURE v1.3.0[53] to determine ancestry using supervised mode whereby samples from the 1000 Genomes project[54] are used as the known ancestry reference and the admixture for UK Biobank samples is estimated in terms of their composition of 1000 Genomes super populations: European (EUR), African (AFR), East Asian (EAS), South Asian (SAS) and American/Hispanic (AMR).

##### ADMIXTURE method

1. Use the set of 101,284 SNPs used by the UK Biobank analysis group for principal component analysis[55]: https://biobank.ctsu.ox.ac.uk/crystal/crystal/auxdata/snp_pca_map.txt
2. Download the 1000 Genomes Phase 3 VCF files: http://ftp.1000genomes.ebi.ac.uk/vol1/ftp/release/20130502/
3. Select the 99,085 SNPs present in both the 1000 Genomes samples and UK Biobank samples.
4. Run ADMIXTURE in supervised mode with the 1000 Genomes samples as the known ancestry, specifying that there are 5 founder populations.

It was not computationally feasible to run ADMIXTURE using all UK Biobank samples so we removed the 321,047 UK Biobank European samples used for discovery in our previous paper[52] (defined as European by k-means clustering) and ran it on the remaining 170,225 samples. We decided that the 321,047 samples removed from the ADMIXTURE run were unambiguously of European ancestry from plots of principal components.

##### ADMIXTURE results

ADMIXTURE returns the ancestry composition for each sample in terms of proportions of each of the 5 1000 Genomes founder populations e.g. 85.6% EUR, 0.01% AFR, 10.2% SAS, 3.6% EAS, 0.34% AMR. We compared the ancestry assignment using thresholds of 70%, 75%, 80% and 90% for the dominant super population in each sample's ancestry, with samples where no super population passed the threshold being set to missing. Using principal component plots we compared the distribution of 1000 Genomes samples of known ancestry with UK Biobank samples with ancestry defined by ADMIXTURE thresholds (Supplementary Figure 1). We decided a threshold of 75% gave a good balance between clearly demarking the appropriate ancestry groups and not excluding too many unassigned samples. The number of samples assigned to each ancestry group and finally included in the GWAS after quality control and matching to lung function data are shown in Supplementary Table 4.

#### Imputation

Before imputation sample and variant quality control filters were applied to the genotype data in each cohort: generally retaining individuals with a call rate of >95%, variants with a call rate of >95% and MAF >1% (the filters used in each cohort and number of SNPs included in the imputation are given in Supplementary Table 3; numbers of individuals that remained after filtering are given in Supplementary Table 2. Cohorts were imputed to either The Haplotype Reference Consortium (HRC) panel[56] or the 1000 genomes project panel[54] (Imputation software and reference panel used for each cohort in Supplementary Table 3). The imputation output was checked for consistency of imputation quality and allele frequencies across cohorts by comparing to the HRC or 1000 genomes allele frequencies.

#### Association testing in each cohort

For autosomal variants, all cohorts undertook the following analyses separately in ever smokers and never smokers: (i) restrict the dataset to never-smokers only with complete data on both $FEV_1$ and FVC; (ii) undertook linear regression of the trait on age, $age^2$, sex, and height; (iii) use the residuals as the phenotype for SNP association testing under an additive genetic model. Studies of unrelated individuals that did not account for population structure using a mixed model also included ancestry principal components as covariates in this model.

Studies with related individuals (i) additionally restricted the dataset to individuals with complete data on both $FEV_1$ and FVC; (ii) undertook linear regression of the trait on age, $age^2$, sex, height, and smoking status (never versus ever smoker); (iii) transformed the residuals to ranks and then to normally distributed z-scores; (iv) used the inverse-normal transformed residuals as the phenotype for SNP association testing under an additive genetic model.

For X chromosome variants we asked studies to undertake the analyses as for autosomal variants above, but separately in males and females. For males, SNP dosages were coded as 0 for 0 copies of the coded allele and 2 for 1 copy of the coded allele. Residuals calculated with males and females combined (as for the autosomal analysis) were then used for this analysis.

For studies with related individuals, as well as running the analysis separately in males and females, we additionally ran the analyses in males and females together, with sex as an additional covariate.

For 10 studies included in our previous lung function meta-analysis[52] updated imputation and association testing were not available hence the previous association results were used in this meta-analysis (indicated with an asterisk in Supplementary Table 3).

##### Untransformed association results

We had 2 data sources for untransformed results (i) studies that returned direct untransformed results; (ii) studies where we inferred approximate untransformed results by multiplying the transformed results by the standard deviation of the phenotype; when we compared this inference against actual untransformed results we found it to be a good approximation.

We requested cohorts to carry out the same association testing as in the discovery analysis but without rank inverse-normal transformation. In the case of cohorts EPIC-Norfolk, H2000, LBC1936, ORCADES and VIKING we previously received untransformed lung function GWAS results stratified across multiple age-bands, we therefore, used these results and meta-analysed across age-bands using Stouffer's method. For each lung function phenotype (FEV1, FVC, FEV1/FVC and PEF), we considered test units to be consistent across cohorts (e.g., L and L/s), and restricted variants with an imputation quality INFO >0.5 and a minor allele count >10.

#### Discovery meta-analysis

Firstly, we meta-analysed any within-cohort sets of results i.e. (i) smokers and non-smokers; (ii) males and females run separately for the X chromosome, such that we had 1 set of results contributing to the meta-analysis from each cohort. Before meta-analysis we removed variants with imputation INFO <0.3 or MAC <3. We also ran LD score regression[57] in each cohort (LDSC v1.0.1) with the appropriate ancestry specific reference LD scores from either the 1000 genomes project. Since transformed effect sizes are not on comparable scales we ran a P value meta-analysis (Stouffer's method) weighted by sample size as implemented in METAL[58] (release 2018-08-28 https://github.com/statgen/METAL) with the "SCHEME SAMPLESIZE" option. The LD score regression intercepts were specified as the genomic control factor for METAL to use for each cohort.

After meta-analysis, there were genomic inflation factors λ of 1.025, 1.022, 0.984 and 0.996 for $FEV_1$, FVC, $FEV_1$/FVC and PEF respectively, which were acceptable and hence no further genomic control was applied.

#### Signal selection

Variants with minor allele count (MAC) <20 across the meta-analysis were removed, leaving 66.8M variants for signal selection. As previously[52] a P value threshold of $P < 5 \times 10^{-9}$ was used to define significant SNPs. Region-based sentinel SNPs were defined from the trans-ethnic meta-analysis by the following method:

1. Select the most highly associated SNP as the sentinel for the region.
2. Remove all SNPs within 1 Mb of the sentinel SNP.
3. Repeat 1 & 2 until no SNPs $P < 5 \times 10^{-9}$ are left and we have a list of sentinel SNPs for each 2Mb "locus".
4. Where the extremes of adjacent loci are within 500 kb, merge the loci and pick the top SNP as the sentinel across this new combined locus.

##### Conditional analysis

Conditional analyses were performed on each study separately, for each locus (2Mb for most loci, unless regions are bigger due to overlapping loci). We used GCTA 1.93.2beta[59] (--cojo-cond method) which conditions on a pre-defined set of SNPs. Genome-wide unconditional results were used as input, with genotypes from the 2Mb (or bigger) locus from the appropriate LD reference panel. For European studies we used a random sample of 10,000 individuals from UK Biobank as the LD reference for all other ancestries we used all samples available in 1000 genomes phase 3 from the appropriate super-population (AFR, SAS, EAS or AMR).

Only 49 of the 50 studies in the initial discovery meta-analysis were included in conditional analyses as we were unable to determine an appropriate LD reference panel for QBB which displayed a broad mixture of ancestries within a single cohort.

##### Selecting conditionally independent signals

Our set of conditionally independent signals for each trait were selected by an iterative forward selection process, followed by backward elimination.

In forward selection: (i) In each study condition out the effect of the lead signal; (ii) meta-analyse across all cohorts using Stouffer's method with the same genomic control adjustment as in the unconditional meta-analysis; if any lead SNP is not present in a study, that study is dropped from the meta-analysis; (iii) if there are any lead SNPs with $P < 5x10^{-9}$ in the meta-analysis of conditional results, add that SNP to the list of lead signals for the locus and go back to step i, now conditioning on the extra lead signal as well; (vi) iterate over the steps above until no SNPs meet the $P < 5x10^{-9}$ threshold in the conditional meta-analysis.

Following forward selection we do a backward elimination process: (i) obtain joint P values for all SNPs in the conditional set for each locus; (ii) meta-analyse results of the joint association tests across studies, again using Stouffer's method and the same genomic control adjustment for each study as in the unconditional analysis; (iii) if any SNP in the conditional set does *not* attain genome-wide significant evidence of association, remove the SNP with the largest association P-value from the conditional set (backward elimination).

We then iterate between forward selection and backward elimination steps until: (i) no SNPs outside the conditional set attained genome-wide significant evidence of residual association in the meta-analysis (check after forward selection); (ii) all SNPs in the conditional set attained genome-wide significant evidence of association in the joint model after meta-analysis (check after backward elimination).

For some cohorts we only have a Z score in the association results because they were first meta-analysed across within-cohort strata such as smoking group or sex using Stouffer's method. GCTA requires an effect size (beta) and standard error (SE), so for these studies we calculate beta and SE from the Z score and effect allele frequency *p* using the following equations[60]:

$$SE = 1/\sqrt{2p(1-p)(n+Z^2)}$$
$$beta = zSE$$

##### Identifying distinct signals across traits

Colocalisation analyses were conducted for any signals (sentinel ± 500kb) overlapping across at least two traits and in the genomic region merged across the traits. Within the genomic region for colocalisation, the signals of any trait had to overlap each other. We found 1510 such genomic regions, among which 272 were overlapped by two traits, 405 were overlapped by three traits and 833 were overlapped by four traits. We used the moloc[61] R package (moloc_test function) to calculate the posterior probabilities (PPA) of there being a shared causal variant across traits or distinct causal variants underlying the association signals. Conditional summary association statistics (i.e. conditioning out the effect of other signals within the locus and meta-analysing across all cohorts) were used as input if multiple signals were detected within the locus. The criteria used to determine the colocalised signals were: (i) the model with highest PPA was selected as the most likely model for each colocalisation analysis; (ii) if the selected model was not the one with all the signals across traits being colocalised, PPA was recalculated for the subset of the colocalised signals; (iii) if a signal of one trait was colocalised with multiple signals of another trait, we chose the colocalisation result with highest PPA; (iv) if signals of different traits colocalize within pairs but are not colocalised over all traits, we chose the pair of signals colocalised with highest PPA; (v) if the highest PPA was <0.5 then we consulted other evidence at the locus (e.g. look at the LD between the signals and do colocalisation analysis with a subset of the signals). For the signals being colocalised, we reported the sentinel with lowest P value as the lead sentinel and the corresponding trait as the lead trait.

#### Novelty of signals with respect to previously reported curated lung function signals

We searched preprint servers Pubmed and GWAS Catalog to identify applied studies focused on curated lung function or COPD, identifying from those studies lung function associations reaching $P<5x10^{-9}$. These included 375 signals from the following sources: China Kadoorie Biobank[62]: 7; Global Biobank Meta-analysis Initiative[63]: 17; UK Biobank and the International COPD Genetics Consortium[64]: 19; UK Biobank and International Lung Cancer Consortium[65]: 53; UK Biobank and SpiroMeta consortium[52]: 279, giving a total of 375 signals. 101 previously reported sentinels are identical to one of our 1020 sentinels; 261 of these previously reported sentinels are within ±1 MB of our 1020 lung function signals: 68 previous sentinels are within ±1 MB of 1 of a single signal, 193 are near multiple signals.

To determine whether the above reported sentinels correspond to the same nearby signal from our results, we conditioned our lung function signals on the previously reported nearby sentinels. As our signals are conditionally independent from each other but the reported signals in literature may not be, we mapped each reported nearby sentinel to only one of our signals for checking independence i.e. the signal with largest change of P value (largest $log_{10}P/log_{10}P_{conditional}$) after conditional analysis.

For the 261 (68 + 193) pairs of our lung function signals and previously reported sentinels, we compare the original P value and the conditional P value of our signal to see whether the reported signal largely explains our lung function signal i.e. there is large attenuation of our lung function signal upon conditioning on the previously reported sentinel: we used $log_{10}P/log_{10}P_{conditional}$ >1.5 as the threshold for attenuation to define our signal as being explained by a previously reported signal.

For signals that were attenuated to this extent there were cases where the conditional P value was still significant. This may happen in two circumstances: i) the P value of the signal is affected by other signals in LD so the conditional P value would not be completely attenuated; ii) the current signal and the reported signal are independent despite conditional attenuation; to account for this case we did not define any signals as the same if the sentinels have $r^2$ <0.2.

After applying the above procedures, of our 1020 signals for lung function meeting a genome-wide significance threshold of $P < 5\times10^{-9}$, 713 meet similarly stringent criteria for the first time.

#### Prioritising putative causal genes by variant-to-gene mapping

##### eQTL/pQTL colocalisation

To identify putative causal genes, we colocalised 1020 lung function signals with expression quantitative trait loci (eQTLs) or protein quantitative trait loci (pQTLs) using coloc_susie method[66], which relaxes the single causal variant assumption and increases the power and accuracy of eQTL/pQTL colocalisation. We used three eQTL resources to identify associations with gene expression in different tissues: (i) GTEx V8 (downloaded from https://www.gtexportal.org/ July 2020; tissues: *Stomach, Small Intestine Terminal Ileum, Lung, Esophagus Muscularis, Esophagus Gastroesophageal Junction, Colon Transverse, Colon Sigmoid, Artery Tibial, Artery Coronary, Artery Aorta);* (ii) eQTLgen[67] blood eQTLs; 2 of 37 cohorts are non-European (~1.1% non-European samples); (iii) UBC lung eQTL. Two blood pQTL resources were used to identify associations with protein levels: (i) INTERVAL pQTL[68] and (ii) SCALLOP pQTL (http://scallop-consortium.com/).

Only European cohorts (17 cohorts for PEF and 39 cohorts for FVC, $FEV_1$ and $FEV_1$/FVC) were included in lung function meta-analysis results, making the lung function signals comparable with signals in eQTL/pQTL resources. For regions with multiple signals, we conditioned each signal on all other signals in the region (±1 MB) before colocalisation. If the eQTL/pQTL resources were built on GRCh38/hg38, the coordinates of the lung function results were lifted over to GRCh38/hg38 coordinates before colocalisation. For each signal, the gene-tissue pairs were selected if there were significant eQTLs/pQTLs (i.e. For UBC lung eQTL, eigenMT with a LD panel was used to calculate adjusted P value and Benjamini-Hochberg procedure was applied to control false discovery rate (FDR) at 5%. For other eQTL databases, significant eQTLs were defined and provided by the data source) within the region of lung function sentinel (±1 MB) in the eQTL/pQTL resources.

To make the analysis more computationally feasible, the signal-gene-tissue pairs were tested for colocalisation only if the common variants between lung function signal and eQTL/pQTL resources within the region of the lung function sentinel (±1 MB) covered the position of lung function sentinel. A random selection of 10,000 European ancestry individuals from UK Biobank were used as the LD reference for both lung function meta-analysis and the eQTL/pQTL resource. To ensure the LD matrix calculated from the reference panel was positive definite, variants with missing genotypes >0.5% in the reference panel were removed and then individuals with missing genotypes were removed, which resulted in a sufficient sample size of reference population and number of variants for colocalisation analysis. The LD matrix was calculated by plink v1.9 ("--recode A" option; by default the minor allele was counted) and R. For both lung function meta-analysis and eQTL/pQTL resource, the effect direction of the variant was flipped if the effect was not the effect of the minor allele, thereby making it consistent with the LD matrix. To avoid SuSiE multicollinearity problems within regions with many variants in high LD, "$r^2$.prune" parameter was set to 0.2. Since the lung function signals were conditioned on all the other signals, we set trim=1.0 for lung function statistics which was unlikely to lose anything interesting while speeding up the computation.

#### eQTL results

There were 2056 signal-gene pairs being colocalised (posterior probability of colocalisation >=0.9) in different tissues by coloc_susie (HLA, X chromosome and smoking signals excluded). We identified 831 candidate genes, implicated by 408 of the 1020 signals (Supplementary Table 28). These genes were identified in at least one tissue and up to 12 tissues. The genes that only came up in one tissue weremainly from tissues with larger sample sizes (e.g. blood in eQTLgen with 31,864 samples, lung in UBC lung eQTL with 1111 samples, and artery tibial/lung/esophagus muscularis in GTEx V8 with 584, 515 and 497 samples).

#### pQTL results

We identified 75 candidate genes, consisting of 14 cis genes and 62 trans genes (Supplementary Table 28). The 14 cis genes were implicated by 14 signals and the 62 trans genes were implicated by 15 signals.

#### Rare variant association

We checked for rare (MAF <1%) exonic associations near (±500 kb) our lung function sentinels using both single-variant and gene-based collapsing tests.

For single variant association, we looked up rare exonic associations ($P < 5\times10^{-6}$) near our lung function sentinels in the results provided at the AstraZeneca PheWAS Portal[69] (https://azphewas.com/), which contains single variant associations in 281,104 UK Biobank exomes for $FEV_1$ and FVC (UK Biobank field IDs 20150 & 20151). We use the results from Backman et al.[70] that tested the associations of loss-of-function and missense variants for $FEV_1$, FVC, $FEV_1$/FVC and PEF in 454,787 UK Biobank participants. The gene for any exonic association was added to our list of prioritised genes. 34 genes were implicated by a single variant association.

We used 3 resources for gene-based collapsing tests (i)The AstraZeneca PheWAS Portal (described above); (ii) Barton et al[71] designed their own imputation panel and performed burden tests for $FEV_1$/FVC and FVC in over ~500K UK Biobank participants (iii) Backman et al 2021 (described above) performed burden tests. 30 genes were implicated by a gene-based association.

There was a union of 54 genes implicated by single variant and/or gene-based test (Supplementary Table 31).

#### Polygenic priority score (PoPS)

The steps in the method were:

1. Apply MAGMA[72] to compute gene-level association statistics and gene-gene correlation using summary statistics from European specific meta-analysis and European LD reference panel.
2. Use PoPS to perform marginal gene feature selection at a nominal significance threshold (P <0.05).
3. Use PoPS to compute a joint enrichment of all selected gene features and calculate the polygenic priority scores for the genes in a leave one chromosome out framework (i.e. if the gene is on chromosome 1 then leave chromosome 1 out);
4. Prioritize gene with top polygenic priority score for each lung function signal within the 500kb/1Mb window.

Within 500kb windows, PoPS prioritized 724 genes (out of 18,383 protein coding genes) for 935 lung function signals. For the lung function signals without prioritized genes within 500kb windows by PoPS, we considered 1Mb windows for prioritised genes and prioritized an additional 31 genes for lung function across 49 signals (Supplementary Table 29 and Supplementary Table 30).

#### Annotation-informed credible sets

We used the ancestry-adjusted meta-regression effect estimates from MR-MEGA to calculate Bayesian posterior probabilities of a variant being causal within each of our 1020 independent signals for lung function (1028 in total minus 8 smoking signals). To calculate annotation-informed prior probabilities of variants being causal we first flagged each variant in our lung function associated loci for overlap with the following annotations:

- Genic location – 10 annotations: *exonic, intronic, 3' UTR, 5' UTR, splicing, upstream, downstream (variant overlaps 1 kb region upstream/downstream of transcription start site respectively), non-coding RNA (ncRNA) exonic, ncRNA intronic, ncRNA splicing*; annotated by ANNOVAR[73].

- Chromatin accessibility in respiratory relevant cells – 24 annotations:
    - Single-cell genome ATAC-seq data[74] from 19 cell types: *Myofibroblast, Pericyte, Ciliated, T_cell, Club, Capillary_endothelial_1/2, Basal, Matrix_fibroblast_1/2, Arterial_endothelial, Pulmonary_neuroendocrine, Natural_killer_cell, Macrophage, B_cell, Erythrocyte, Lymphatic_endothelial, Alveolar Type 1/2* (downloaded from https://www.lungepigenome.org/).
    - ATAC-seq data for 5 human primary lung-cell types implicated in COPD pathobiology[75]; cell types: *large and small airway epithelial cells, alveolar type II, pneumocytes and lung fibroblasts* (downloaded from http://www.copdconsortium.org/).
- Tissue-specific transcription factor binding sites – 1178 annotations: DNase-seq footprinting of 589 human transcription factors in lung and bronchus[76] (we selected footprints from the HINT method with a 20bp seed length and footprint score >200).

We thus had a flag for 1212 annotations in total at each variant in our associated loci. The annotations at each variant were combined with the Bayes factor for association from the MR-MEGA ancestry-adjusted meta-analysis. For 60 of our 1020 signals results were not available from >6 cohorts and hence a standard fixed-effects meta-analysis was used with no adjustment for axes of ancestry. For 3 HLA signals conditional results using GCTA were not available due to excessive collinearity in regions of high LD, hence we have a credible set calculation for 1017 signals in total.

fGWAS[77] was used to test for enrichment of annotations within 1017 lung function signals. We first considered each annotation separately and selected a set of nominally significant annotations for each phenotype (P <0.05 for enrichment) and then iteratively added annotations to a joint model for each phenotype until adding an annotation did not significantly improve the model (P <0.05). We next selected a penalty that maximised the cross-validation penalised likelihood. Then using the optimum penalty iteratively dropped annotations that resulted in a decrease in the cross-validation likelihood. The annotations included in the final joint model for each phenotype used for calculating SNP prior probabilities of causality are shown in Supplementary Table 13.

At each lung function signal we used the annotation-informed posterior probabilities from fGWAS to calculate a 95% credible set. Compared to a fixed-effect inverse-variance weighted meta-analysis, incorporating axes of genetic ancestry reduced the median size of 95% credible sets from 11 to 10 variants, increased the median posterior probability of the most likely causal variant from 0.396 to 0.453 and increased the number of signals with a variant accounting for >50% of the posterior probability from 325 to 367; additionally incorporating functional annotation to weight prior causal probabilities reduced the median credible set size to 9 variants, increased the median posterior probability of the most likely causal variant to 0.491 (Supplementary Figure 7) and increased the number of signals with a variant with >50% posterior probability to 438.

We annotated variants in the 95% credible sets using VEP. At each signal we selected a gene as putatively causal if there was a variant with >50% posterior probability in the credible set that was also either a missense variant, annotated as “deleterious” by SIFT, annotated as “damaging” by PolyPhen or had a CADD PHRED score >=20. Using these criteria, we implicated 33 genes (Supplementary Table 15)

### Supplementary Figures

**Supplementary Figure 1: UK Biobank ancestry clustering with ADMIXTURE**

Principal component plots for 1000 Genomes samples (left panels) and UK Biobank samples (right panels) where UK Biobank ancestry has been assigned to 1000 Genomes super population (EUR, AFR, EAS, SAS or AMR) that exceeds 70%, 75%, 80% or 90% of that sample's ancestry (top to bottom) as determined by ADMIXTURE. UK Biobank ancestry set to missing when no super population exceeds the threshold.

a) **PC2 vs. PC1**

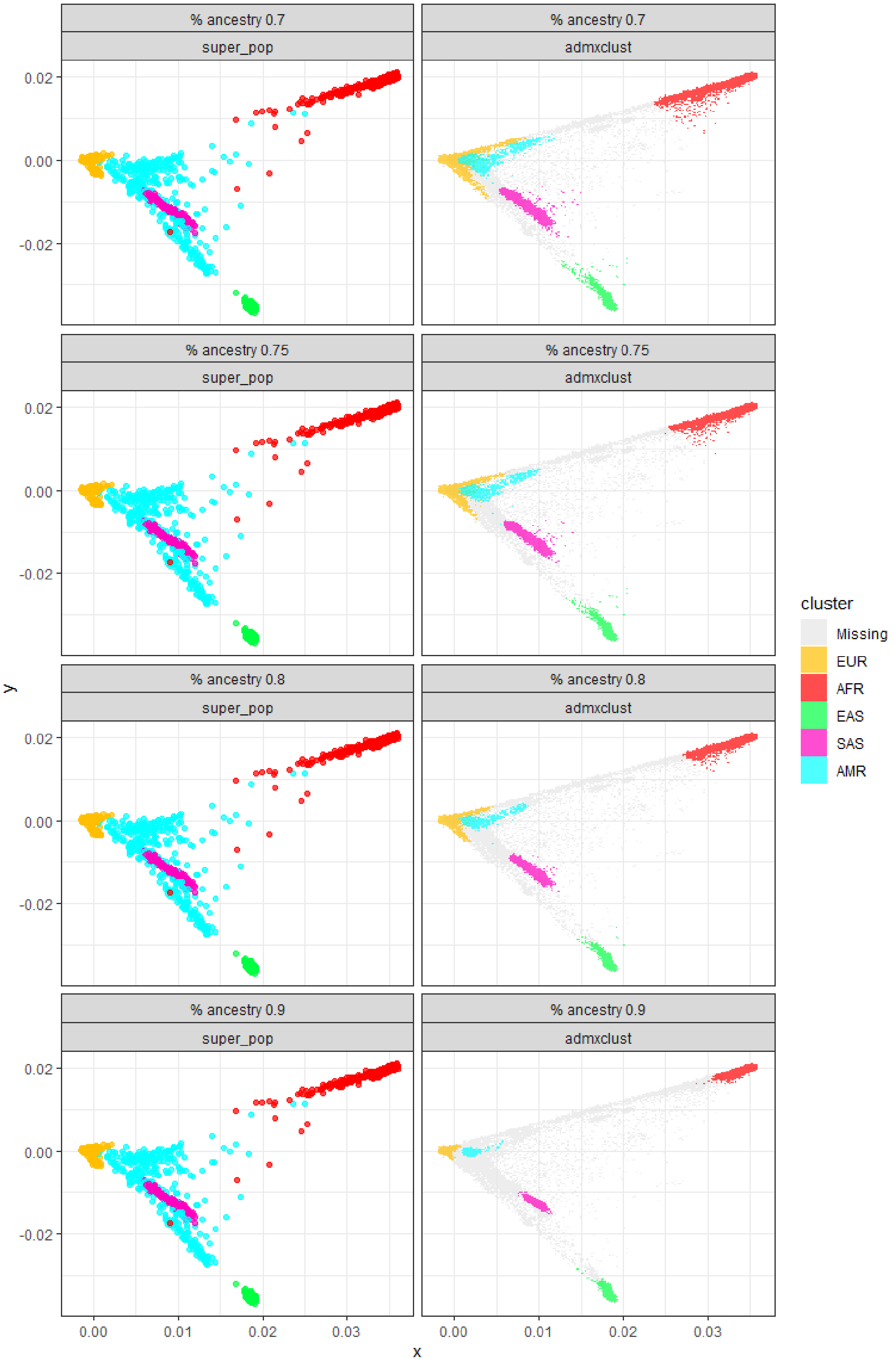

**b) PC4 vs. PC3**

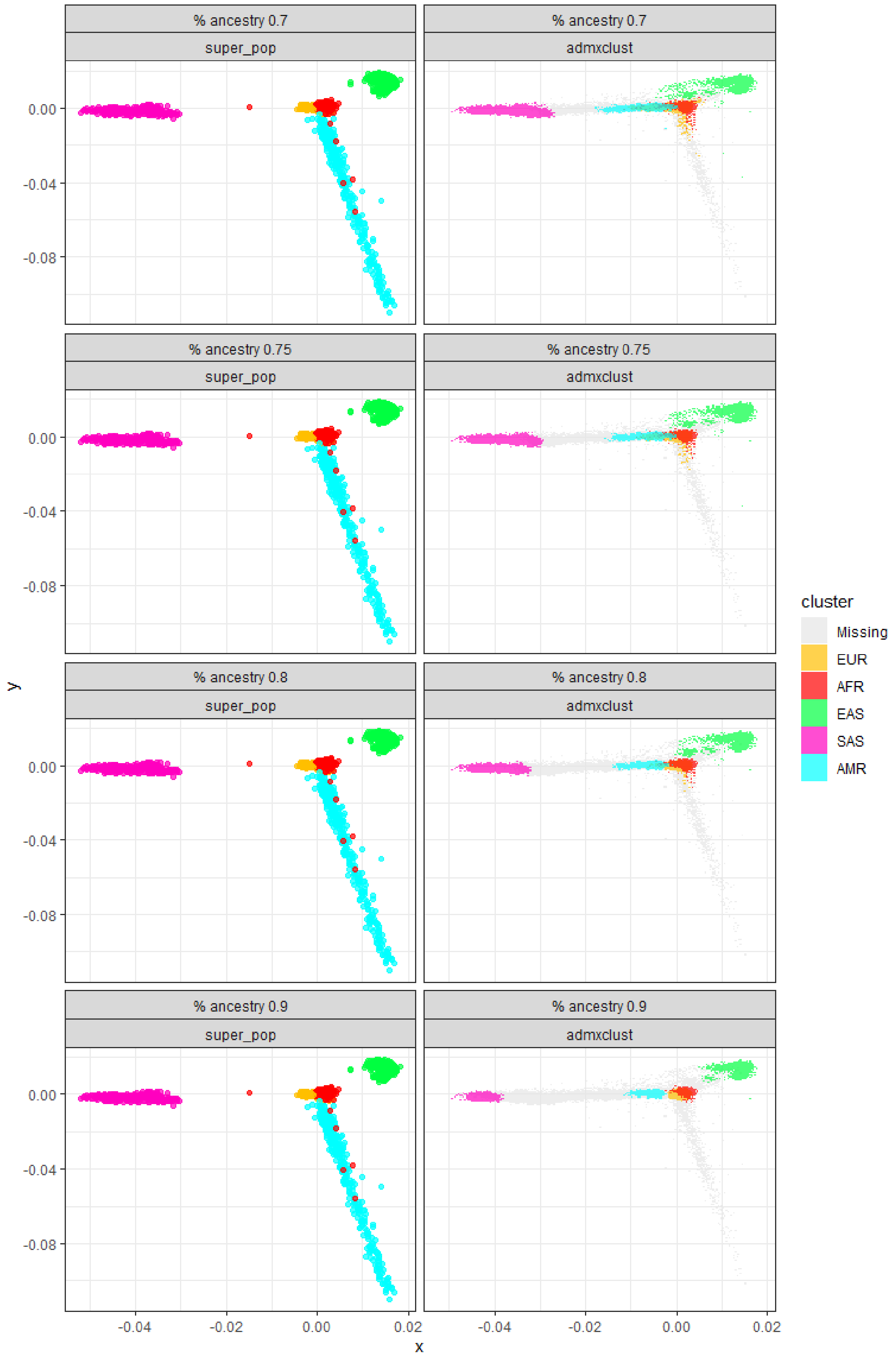
% ancestry 0.7
super_pop
% ancestry 0.7
admxclust
% ancestry 0.75
super_pop
% ancestry 0.75
admxclust
% ancestry 0.8
super_pop
% ancestry 0.8
admxclust
% ancestry 0.9
super_pop
% ancestry 0.9
admxclust
0.00
-0.04
-0.08
y
-0.04
-0.02
0.00
0.02
x
cluster
Missing
EUR
AFR
EAS
SAS
AMR

**Supplementary Figure 2: QQ plots for unconditional multi-ancestry meta-analysis across 49 cohorts**
66.8M variants with minor allele count >= 20.

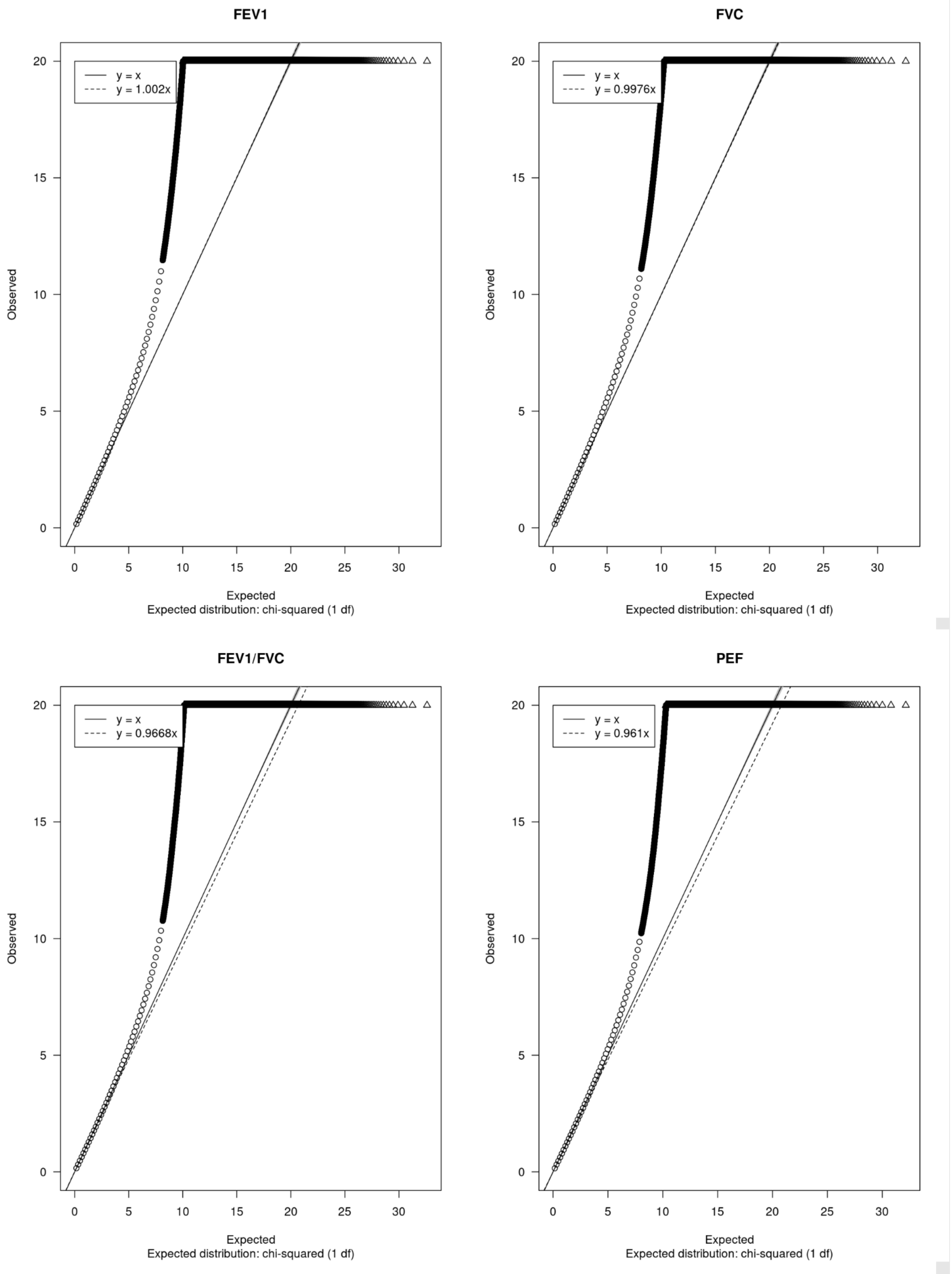

**Supplementary Figure 3: Manhattan plots for unconditional multi-ancestry meta-analysis across 49 cohorts**
Green dots indicate the reported sentinels.

**a)**

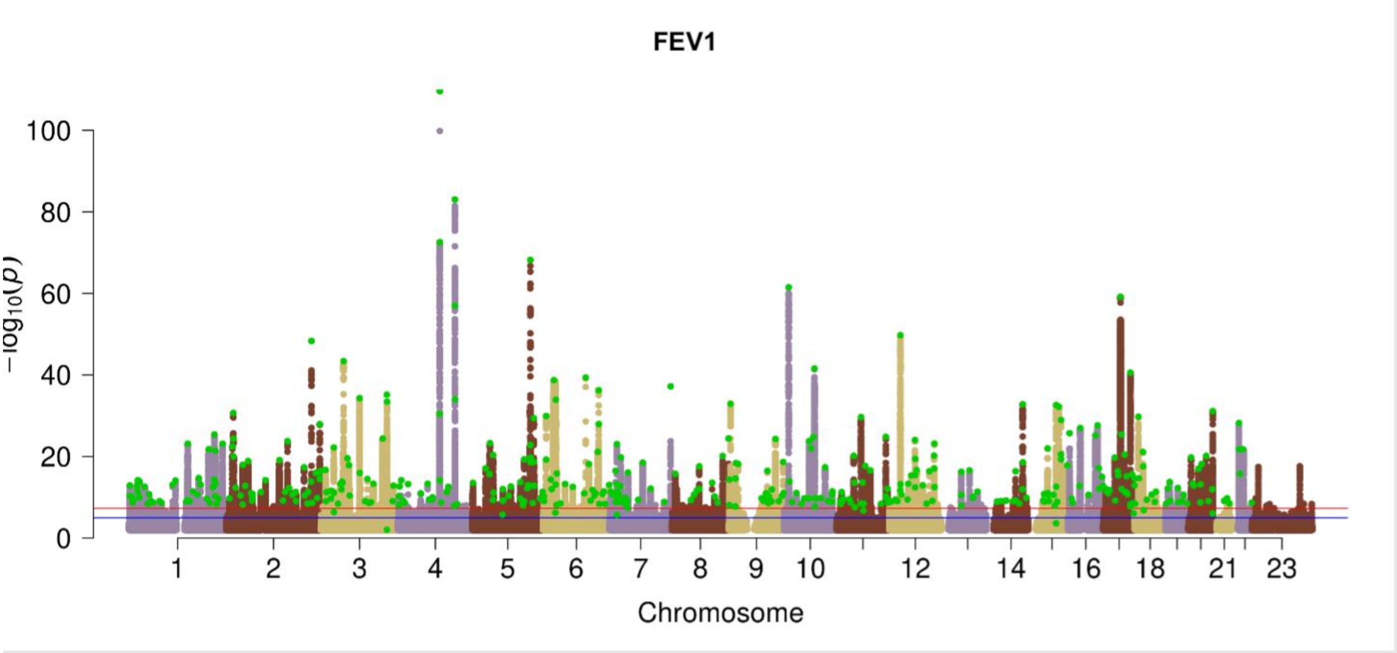

**b)**

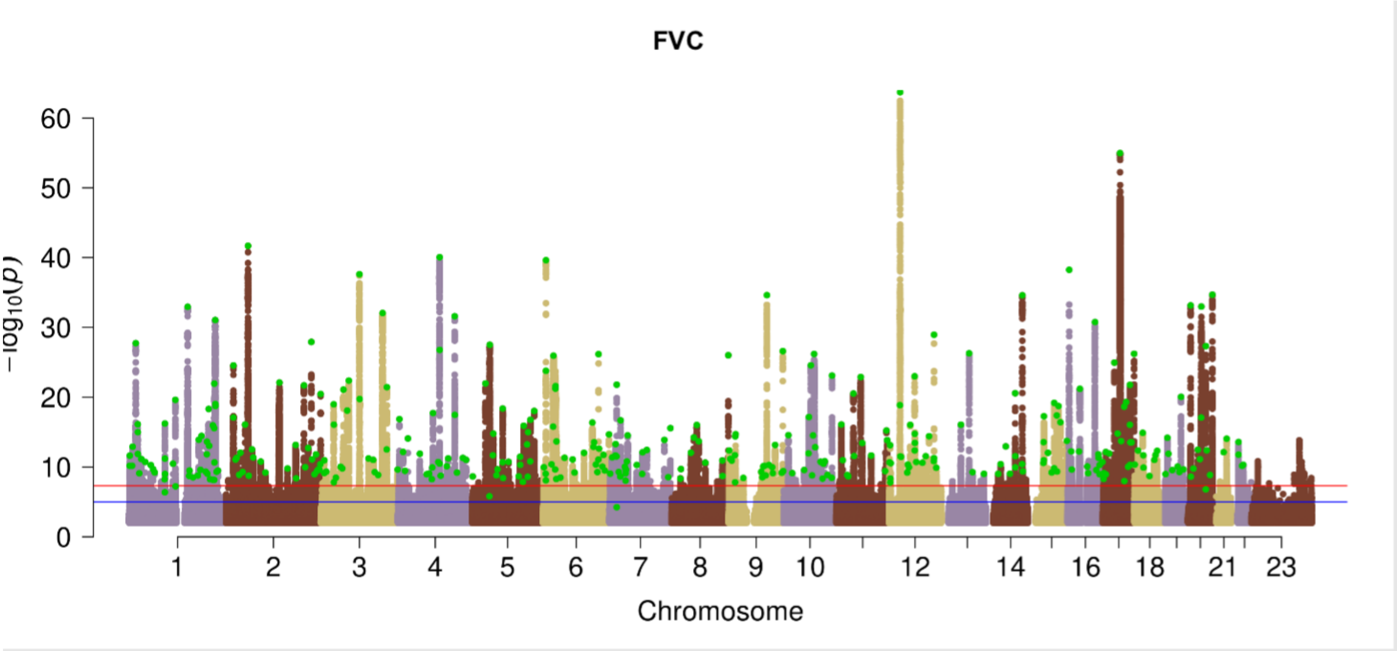

c)

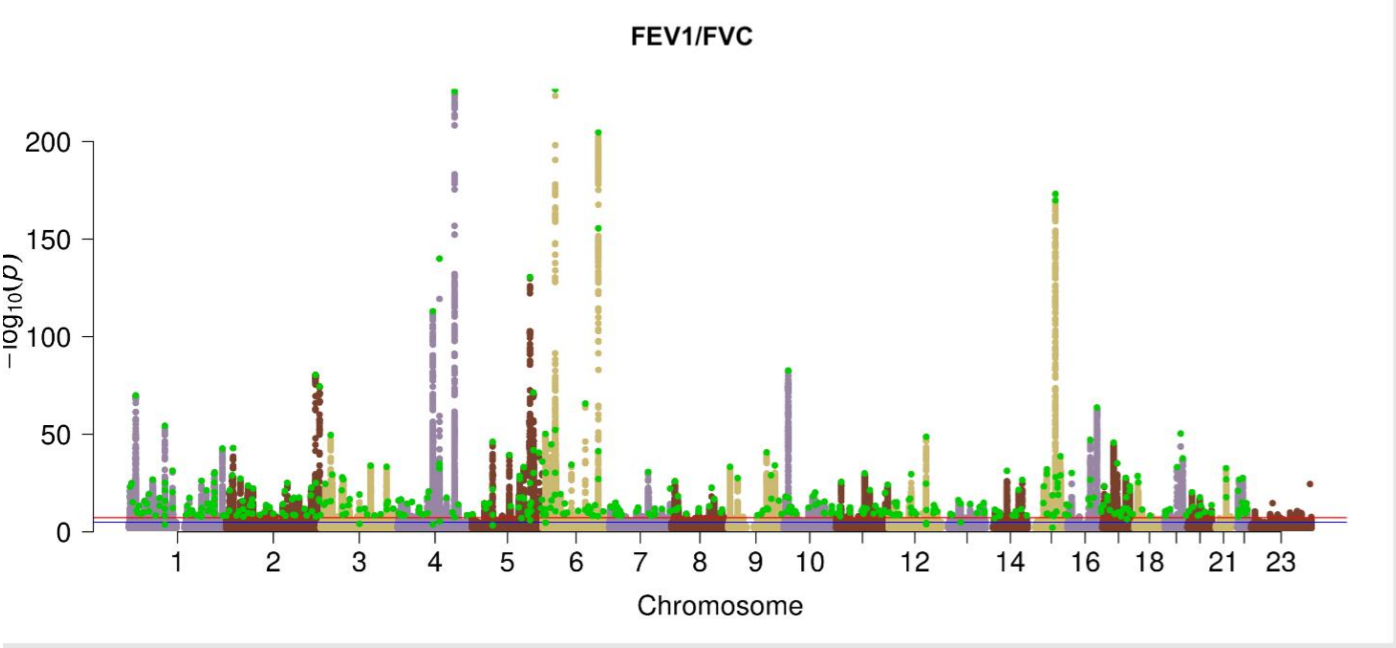
FEV1/FVC
−log10(p)
200
150
100
50
0
1
2
3
4
5
6
7
8
9
10
12
14
16
18
21
23
Chromosome

d)

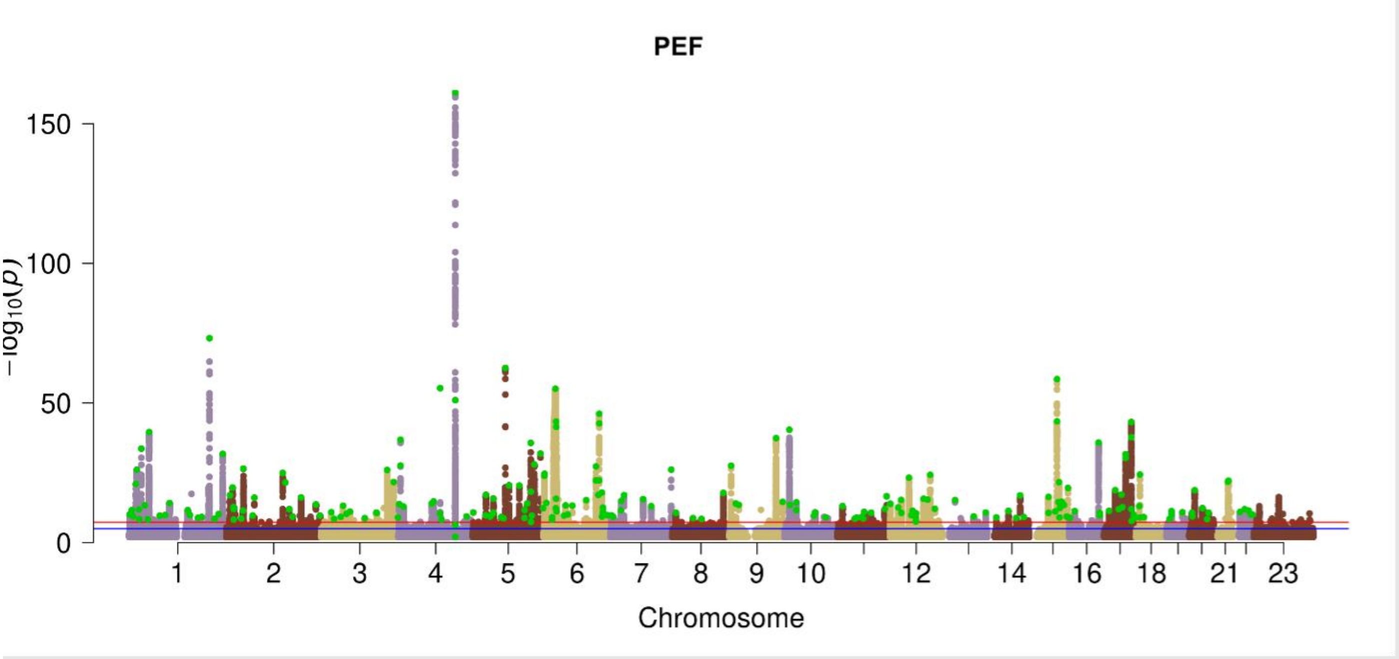
PEF
−log10(p)
150
100
50
0
1
2
3
4
5
6
7
8
9
10
12
14
16
18
21
23
Chromosome

**Supplementary Figure 4: Location of cohorts on first two principal components derived by MR-MEGA multidimensional scaling of allele frequencies.**

The colours show the 5 ancestry groups assigned to each cohort, either African (AFR), American/Hispanic (AMR), East Asian (EAS), European (EUR) or South Asian (SAS).

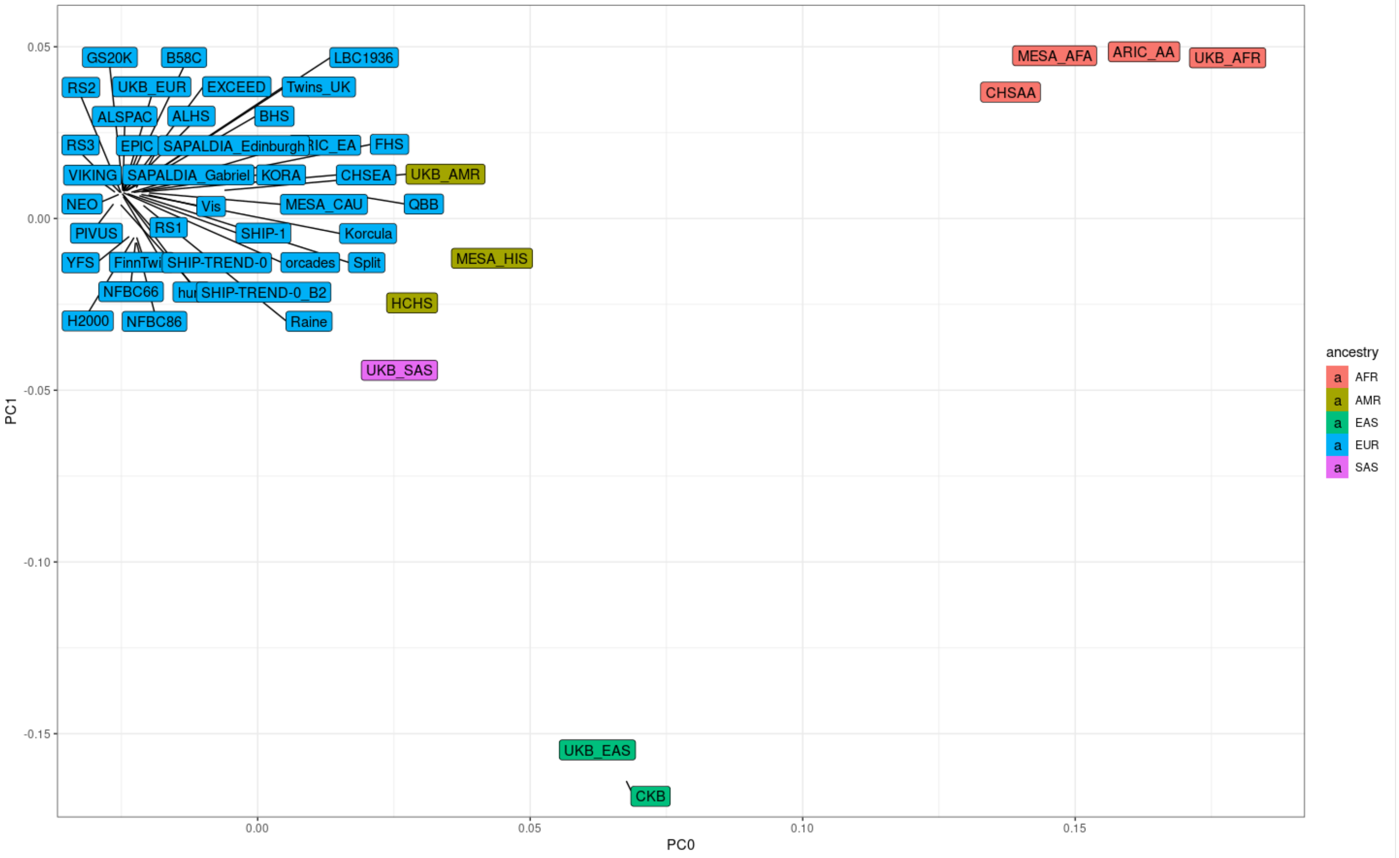

**Supplementary Figure 5. Forest plots for the effect sizes of the five signals showing significant ancestry-correlated heterogeneity at 5% Bonferroni corrected for 960 signals tested**

**a) rs9393688:** Allelic effects are aligned to allele A across GWAS with different ancestry origin (AFR=African, AMR=Admixture American, EAS=East Asian, EUR=European and SAS=South Asian), denoted by black points. The fixed-effect meta-analysed effect sizes within ancestry groups are denoted by blue diamonds. This signal attained nominal significance only in EUR ($P=6.88 \times 10^{-19}$, freq=73.48%) ancestry specific meta-analyses. The heterogeneity due to ancestry was significant at $P=9.78 \times 10^{-6}$.

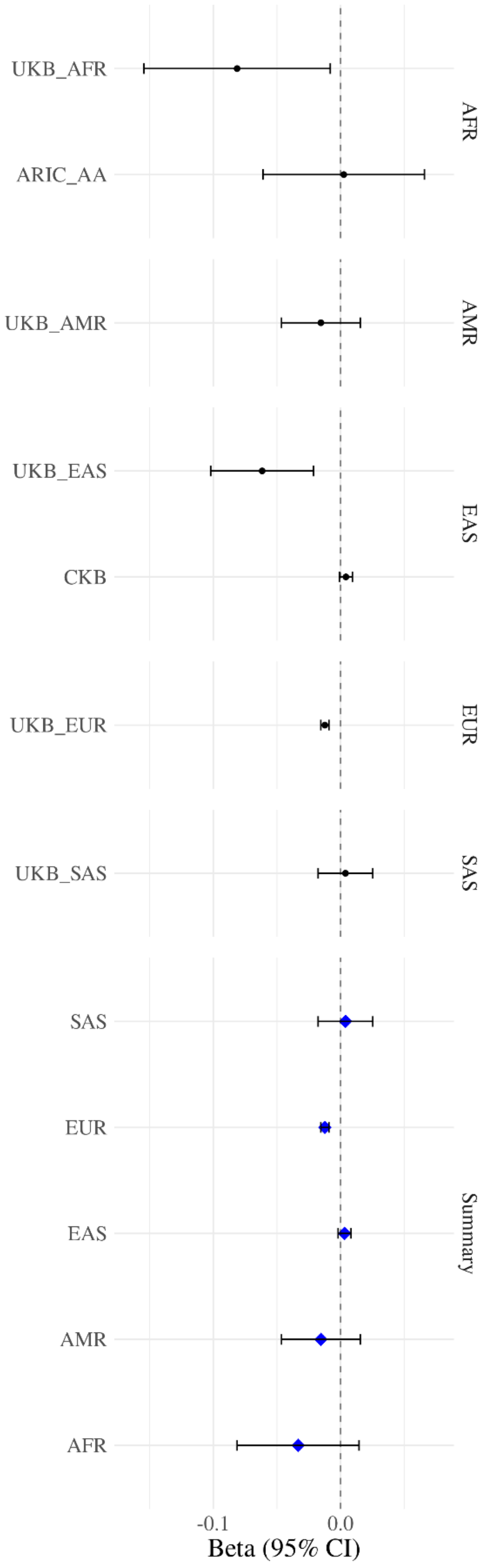

**b) rs28574670(*LTBP4*):** Allelic effects are aligned to allele A across GWAS with different ancestry origin (AFR=African, AMR=Admixture American, EAS=East Asian, EUR=European and SAS=South Asian), denoted by black points. The fixed-effect meta-analysed effect sizes within ancestry groups are denoted by blue diamonds. This signal attained nominal significance in EUR ($P=6.77\times10^{-23}$, freq.=9.6%) and SAS (P=0.0204, freq=46.09%) ancestry specific meta-analyses. The heterogeneity due to ancestry was significant at $P=1.71\times10^{-5}$.

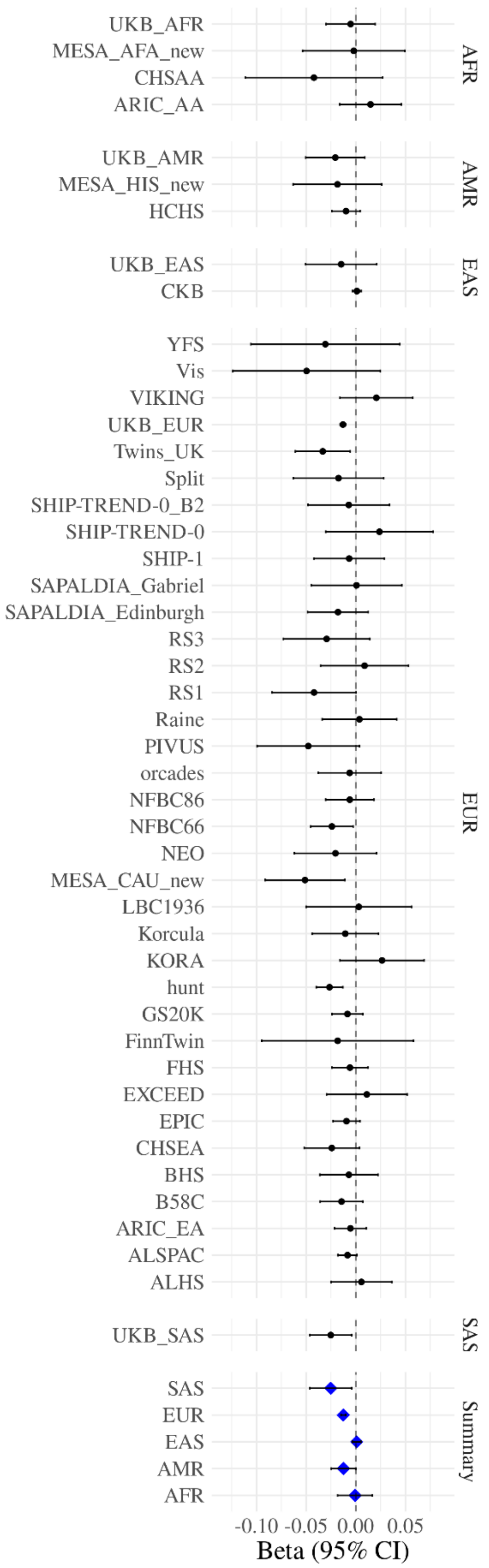

**c) rs7183859 (*THSD4*):** Allelic effects are aligned to allele T across GWAS with different ancestry origin (AFR=African, AMR=Admixture American, EAS=East Asian, EUR=European and SAS=South Asian), denoted by black points. The fixed-effect meta-analysed effect sizes within ancestry groups are denoted by blue diamonds. This signal attained nominal significance in AFR ($P=8.62 \times 10^{-4}$, freq.=9.6%), AMR ($P = 5.36 \times 10^{-4}$, freq.=16.0%) and EUR ($P=3.06 \times 10^{-17}$, freq=17.7%) ancestry specific meta-analyses. The heterogeneity due to ancestry was significant at $P=3.33 \times 10^{-5}$.

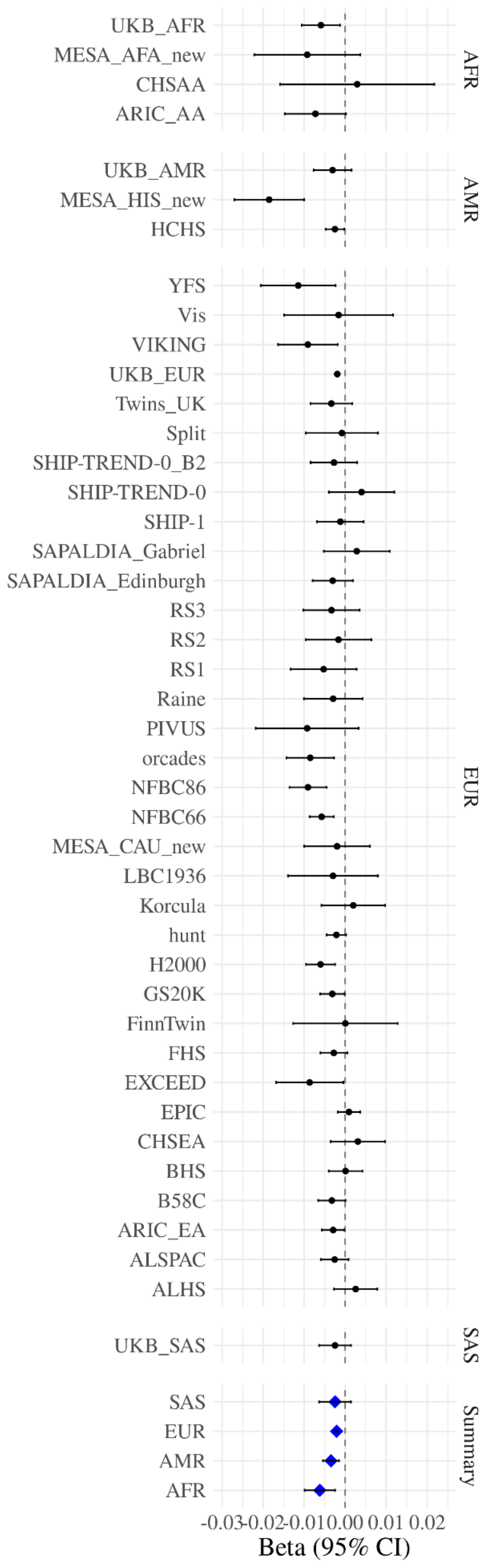

**d) rs59985551 (*EFEMP1*):** Allelic effects are aligned to allele T across GWAS with different ancestry origin (AFR=African, AMR=Admixture American, EAS=East Asian, EUR=European and SAS=South Asian), denoted by black points. The fixed-effect meta-analysed effect sizes within ancestry groups are denoted by blue diamonds. This signal attained nominal significance in AMR (P = 6.83 x10$^{-3}$, freq=21.97%) and EUR (P=5.60x10$^{-39}$, freq=23.0%) ancestry specific meta-analyses. The heterogeneity due to ancestry was significant at P=3.77x10$^{-5}$.

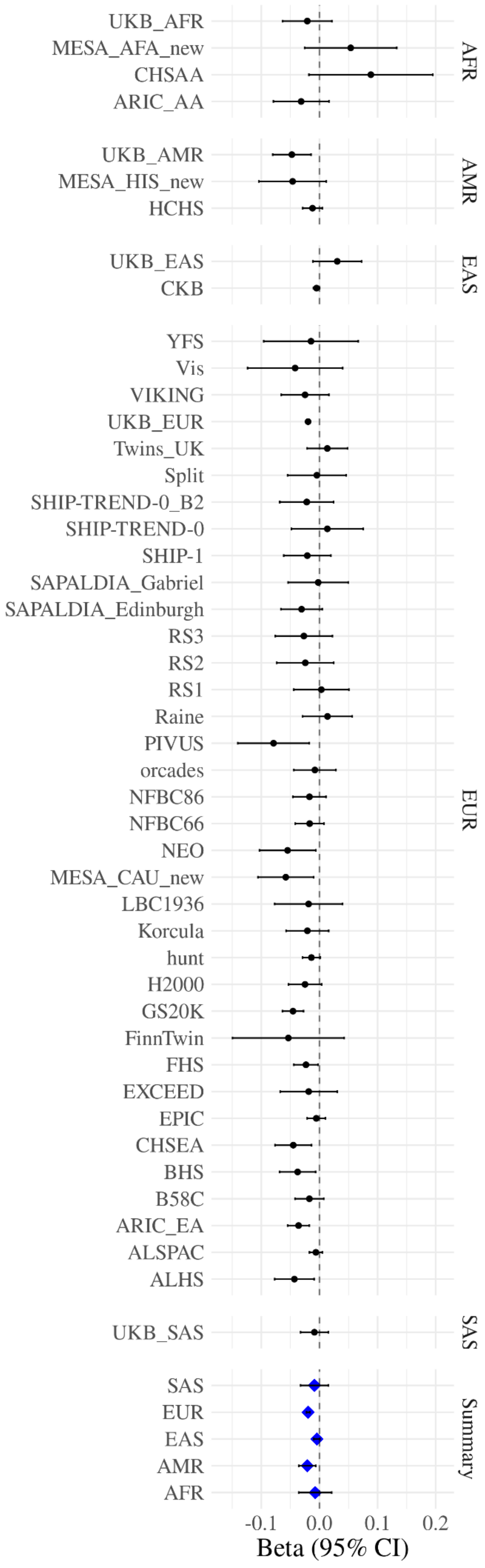

**e) rs78101726(*MECOM*):** Allelic effects are aligned to allele A across GWAS with different ancestry origin (AFR=African, AMR=Admixture American, EAS=East Asian, EUR=European and SAS=South Asian), denoted by black points. The fixed-effect meta-analysed effect sizes within ancestry groups are denoted by blue diamonds. This signal attained nominal significance in AMR ($P = 4.29 \times 10^{-4}$, freq=78.14%) and EUR ($P=2.13 \times 10^{-27}$, freq=17.7%) ancestry specific meta-analyses. The heterogeneity due to ancestry was significant at $P=3.86 \times 10^{-5}$.

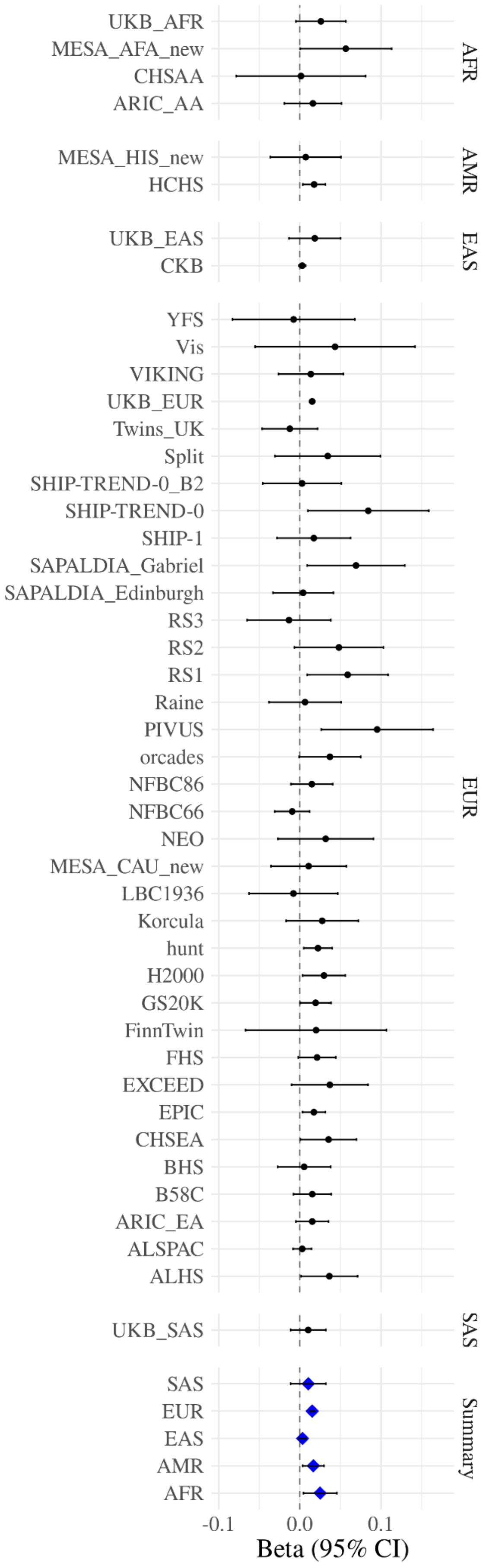

**Supplementary Figure 6: Distribution of effect sizes with allele frequency.**

The absolute effect size (beta) for $FEV_1$ (L), FVC(L), $FEV_1$/FVC and PEF(L/sec) is plotted against the minor allele frequency (MAF) for 1020 sentinel variants (26 of which are also the putative causal variant for the signal) and 7 putative causal variants that are not the sentinel for the signal. A putative causal variant accounts for >50% of the posterior probability of being causal in an annotation-informed credible set for the signal. A variant only appears in panels for traits other than the one for which it is a sentinel or putative causal variant if it also associated with that trait ($P < 5 \times 10^{-5}$). Variants with large effect sizes or with outlying effect sizes compared to other variants of comparable frequency are labelled with the putative causal gene implicated at that signal.

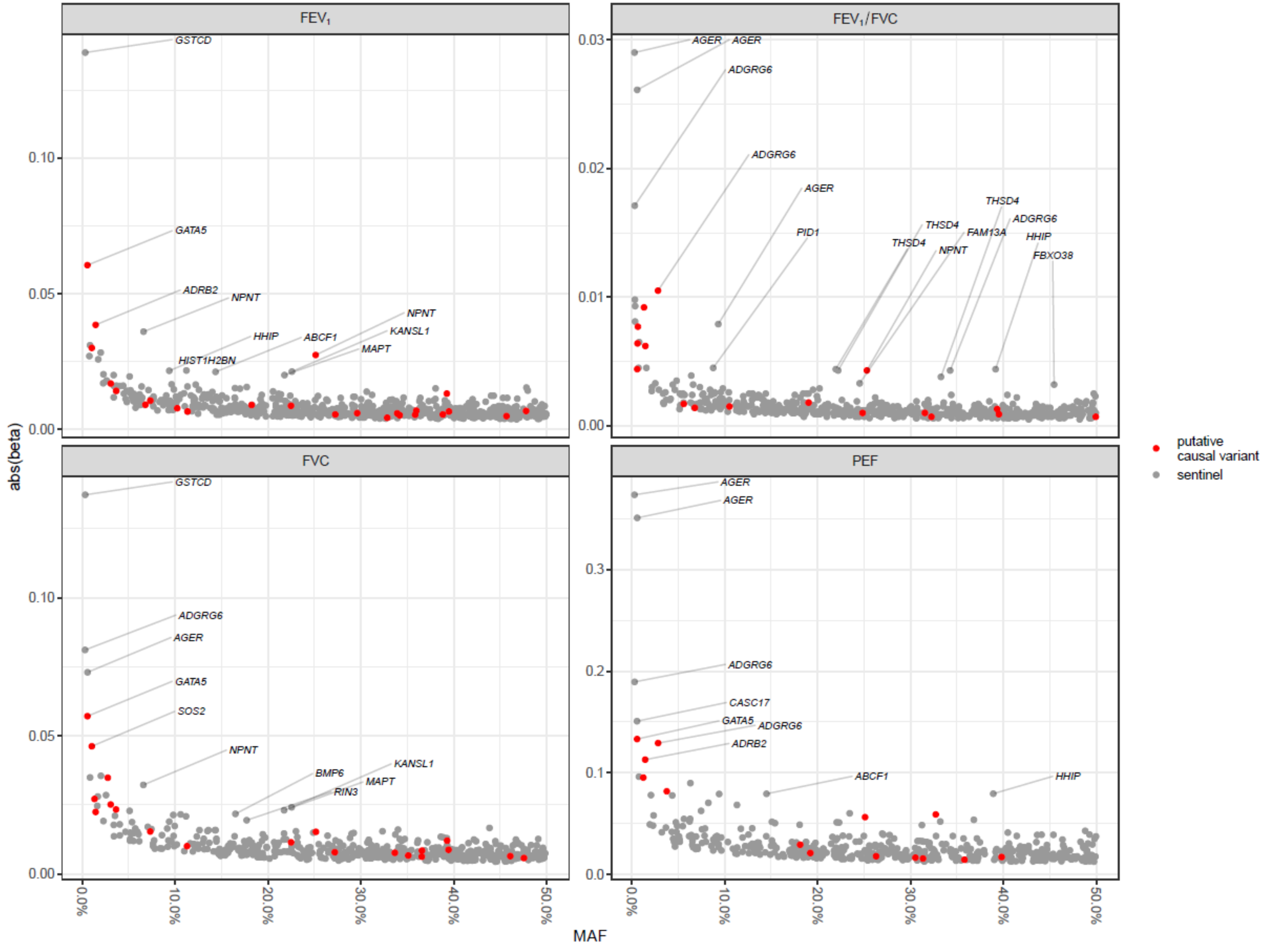

**Supplementary Figure 7: Distribution of number of variants and maximum posterior probability in 95% credible sets**

95% credible sets were constructed based on log Bayes factors from (i) a standard fixed effects meta-analysis ("METAL") across our 49 cohorts; (ii) a meta-regression including axes of genetic ancestry as covariates ("MRMEGA"); (iii) MRMEGA method but additionally with prior probabilities weighted by functional annotation ("Annotation"). The METAL analysis results in credible sets with the distributions shifted towards more variants (n) and lower maximum posterior probability (maxPPA); the Annotation analysis credible sets are shifted towards smaller credible sets with higher maximum posterior probability, with the MRMEGA credible sets in-between the two.

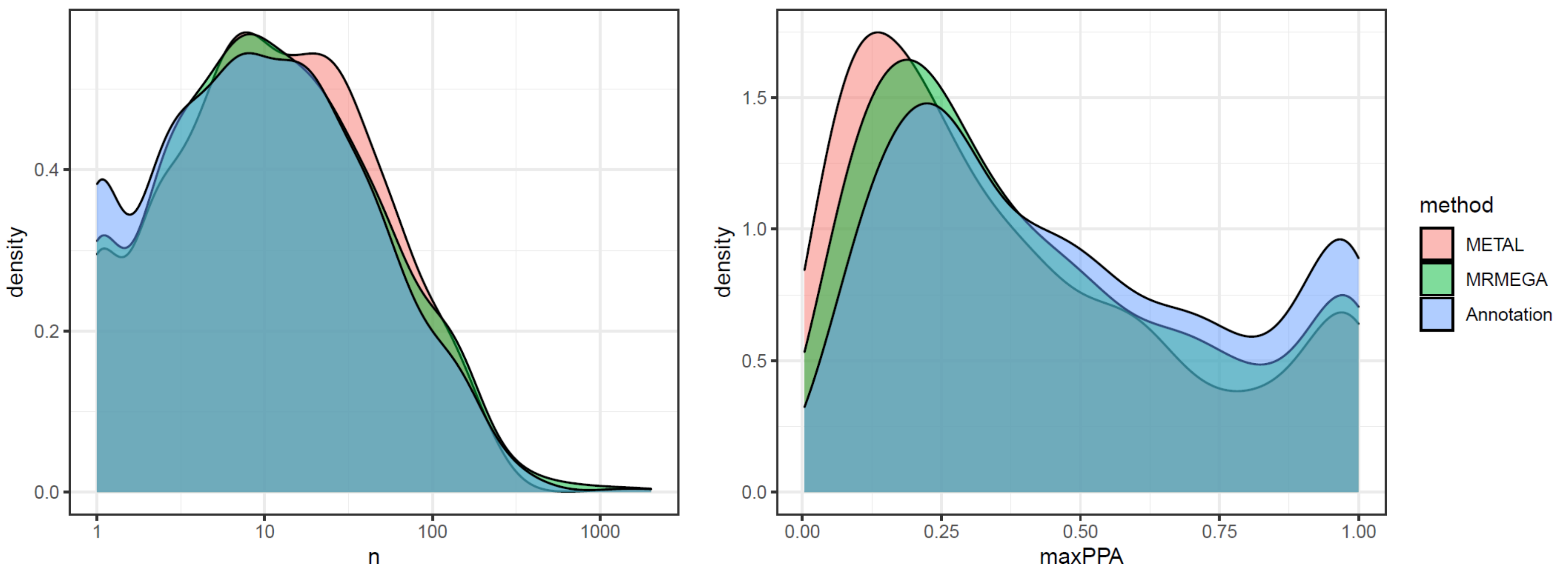

**Supplementary Figure 8: GARFIELD functional enrichment**

The wheel plots display functional enrichment for associations with $FEV_1$, FVC, $FEV_1$/FVC and PEF within different types of functional annotation regions in the ENCODE and Roadmap Epigenomics project: **(a)** DHS Hotspots, **(b)** transcription factor footprints, **(c)** genic annotations, **(d)** chromatin segmentation states, **(e)** transcription factor binding sites, **(f)** open chromatin peaks, **(g)** FAIRE and **(h)** histone modifications. The radial axis shows fold enrichment estimates at trait-genotype association P-value thresholds at $P < 5x10^{-5}$ and $P < 5x10^{-9}$ for each of the cell types tested. Cell types are sorted by tissue, represented along the outside edge of the plot with font size proportional to the number of cell lines from that tissue. Fold enrichment values at the different thresholds are plotted with different colours inside the plot (e.g. values at $P < 5\times10^{-9}$ are in black). If present, the dots along the inside edge of the plot denote significant enrichment for a given cell type at $P < 5\times10^{-5}$ (outermost dot) to $P < 5\times10^{-9}$ (innermost dot).

**a) DNaseI hypersensitive sites (DHS) hotspots**

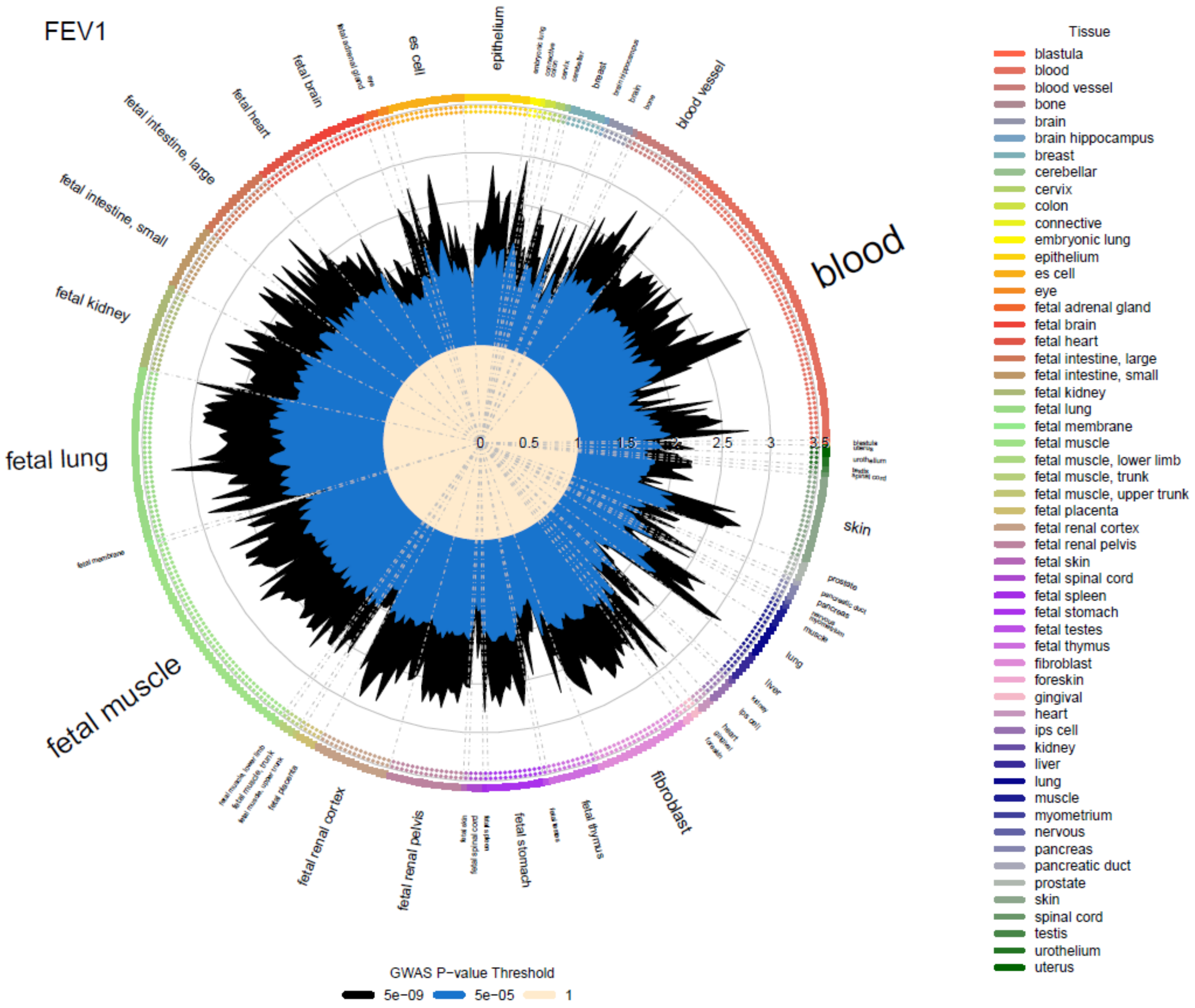

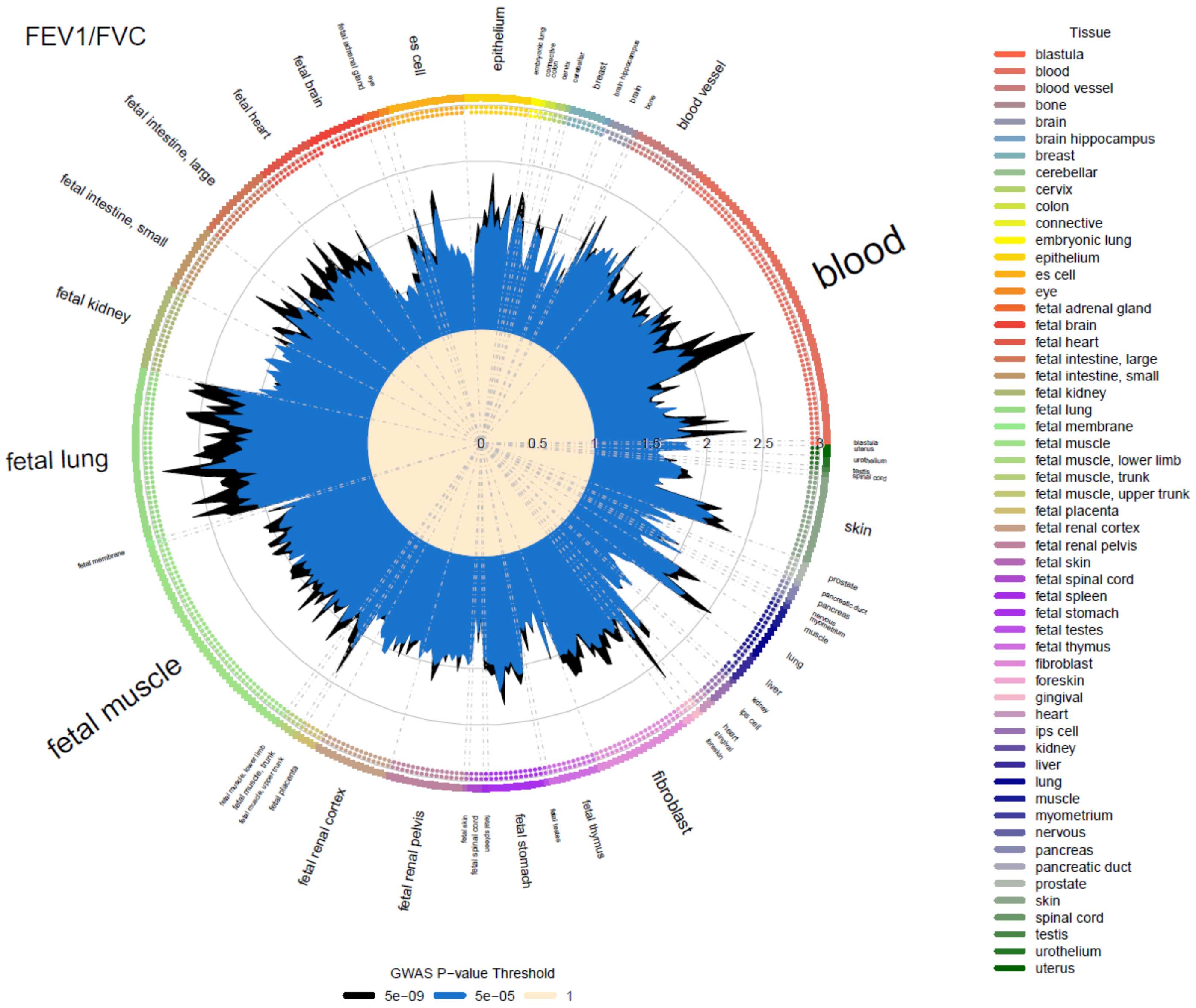
FEV1/FVC
blood
fetal lung
fetal muscle
skin
fibroblast
fetal renal pelvis
fetal renal cortex
fetal stomach
fetal thymus
fetal kidney
fetal intestine, small
fetal intestine, large
fetal heart
fetal brain
es cell
epithelium
blood vessel
fetal membrane
prostate
pancreatic duct
pancreas
lung
liver
kidney
ips cell
heart
gingival
foreskin
breast
brain hippocampus
brain
bone
cerebellar
cervix
colon
connective
embryonic lung
eye
fetal adrenal gland
fetal muscle, lower limb
fetal muscle, trunk
fetal muscle, upper trunk
fetal placenta
fetal skin
fetal spinal cord
fetal spleen
fetal testes
blastula
uterus
urothelium
testis
spinal cord
nervous
myometrium
muscle
0
0.5
1
1.5
2
2.5
3
Tissue
blastula
blood
blood vessel
bone
brain
brain hippocampus
breast
cerebellar
cervix
colon
connective
embryonic lung
epithelium
es cell
eye
fetal adrenal gland
fetal brain
fetal heart
fetal intestine, large
fetal intestine, small
fetal kidney
fetal lung
fetal membrane
fetal muscle
fetal muscle, lower limb
fetal muscle, trunk
fetal muscle, upper trunk
fetal placenta
fetal renal cortex
fetal renal pelvis
fetal skin
fetal spinal cord
fetal spleen
fetal stomach
fetal testes
fetal thymus
fibroblast
foreskin
gingival
heart
ips cell
kidney
liver
lung
muscle
myometrium
nervous
pancreas
pancreatic duct
prostate
skin
spinal cord
testis
urothelium
uterus
GWAS P−value Threshold
5e−09
5e−05
1

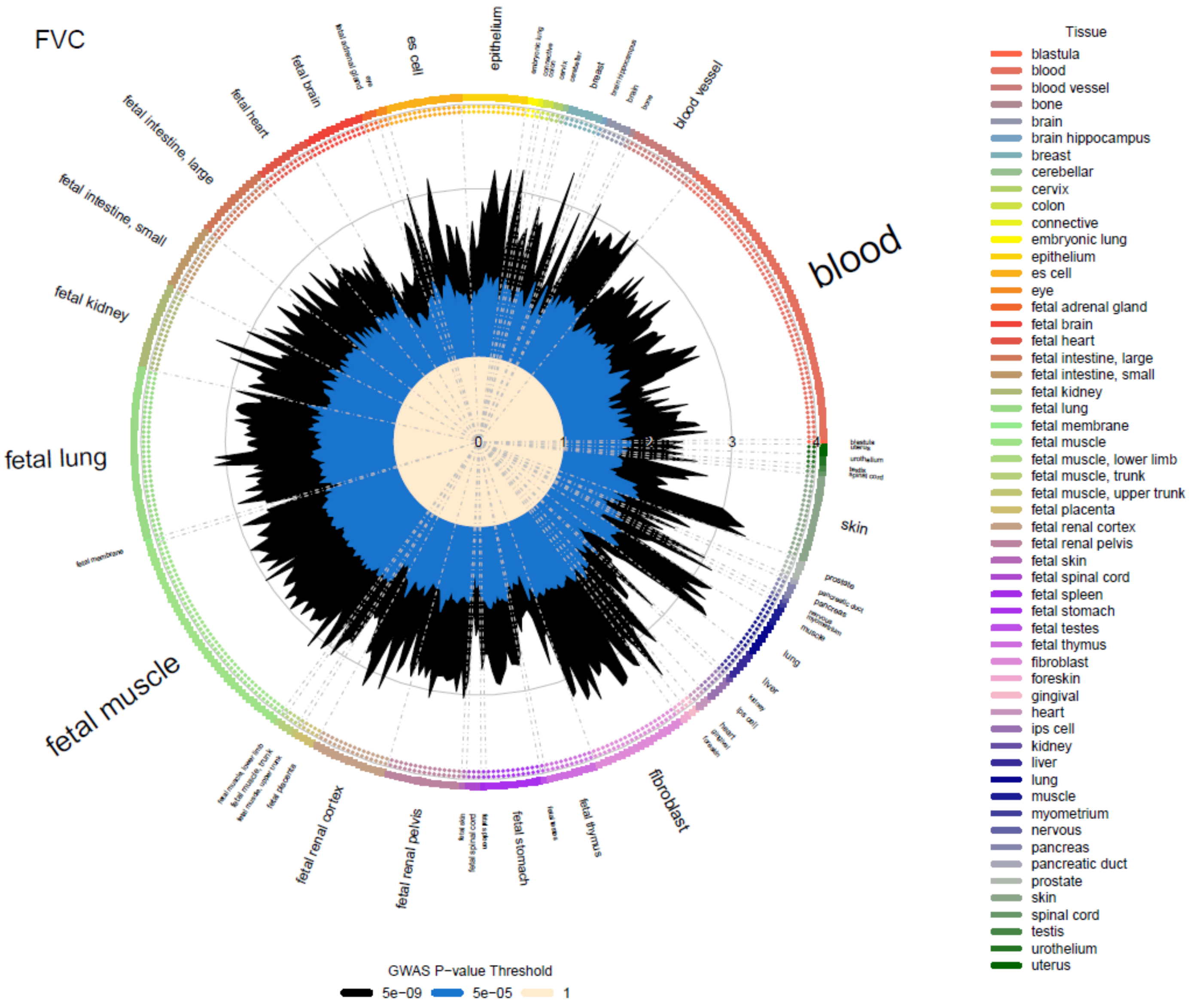
FVC
blood
fetal lung
fetal muscle
skin
fibroblast
epithelium
es cell
fetal brain
fetal heart
fetal intestine, large
fetal intestine, small
fetal kidney
blood vessel
fetal renal cortex
fetal renal pelvis
fetal stomach
fetal thymus
lung
liver
prostate
0
1
2
3
4
Tissue
blastula
blood
blood vessel
bone
brain
brain hippocampus
breast
cerebellar
cervix
colon
connective
embryonic lung
epithelium
es cell
eye
fetal adrenal gland
fetal brain
fetal heart
fetal intestine, large
fetal intestine, small
fetal kidney
fetal lung
fetal membrane
fetal muscle
fetal muscle, lower limb
fetal muscle, trunk
fetal muscle, upper trunk
fetal placenta
fetal renal cortex
fetal renal pelvis
fetal skin
fetal spinal cord
fetal spleen
fetal stomach
fetal testes
fetal thymus
fibroblast
foreskin
gingival
heart
ips cell
kidney
liver
lung
muscle
myometrium
nervous
pancreas
pancreatic duct
prostate
skin
spinal cord
testis
urothelium
uterus
GWAS P−value Threshold
5e−09
5e−05
1

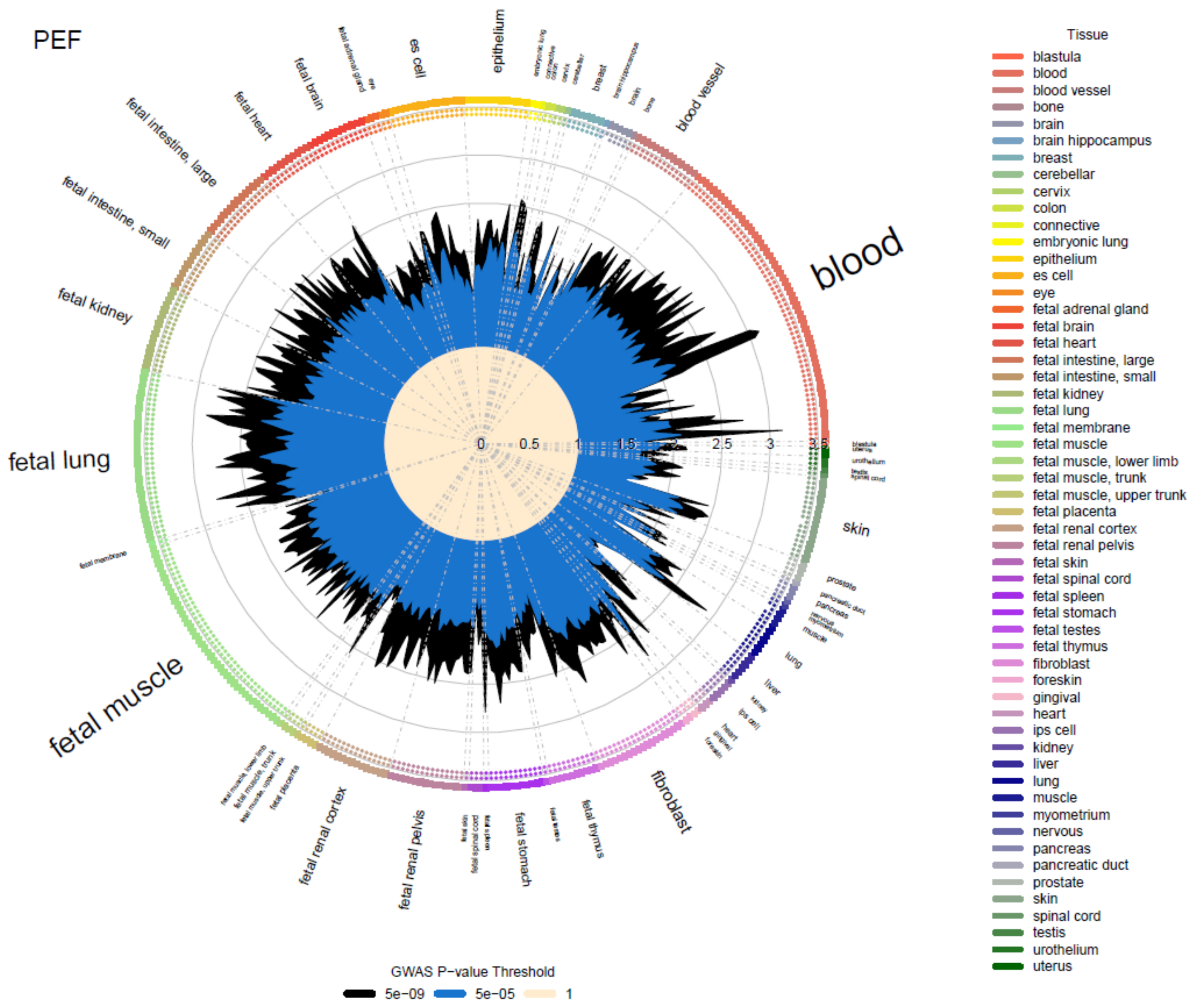
PEF
blood
fetal lung
fetal muscle
fetal kidney
fetal intestine, small
fetal intestine, large
fetal heart
fetal brain
es cell
epithelium
blood vessel
skin
fibroblast
fetal thymus
fetal stomach
fetal renal pelvis
fetal renal cortex
lung
liver
0
0.5
1
1.5
2
2.5
3
3.5
Tissue
blastula
blood
blood vessel
bone
brain
brain hippocampus
breast
cerebellar
cervix
colon
connective
embryonic lung
epithelium
es cell
eye
fetal adrenal gland
fetal brain
fetal heart
fetal intestine, large
fetal intestine, small
fetal kidney
fetal lung
fetal membrane
fetal muscle
fetal muscle, lower limb
fetal muscle, trunk
fetal muscle, upper trunk
fetal placenta
fetal renal cortex
fetal renal pelvis
fetal skin
fetal spinal cord
fetal spleen
fetal stomach
fetal testes
fetal thymus
fibroblast
foreskin
gingival
heart
ips cell
kidney
liver
lung
muscle
myometrium
nervous
pancreas
pancreatic duct
prostate
skin
spinal cord
testis
urothelium
uterus
GWAS P−value Threshold
5e−09
5e−05
1

**b) Transcription factor footprints**

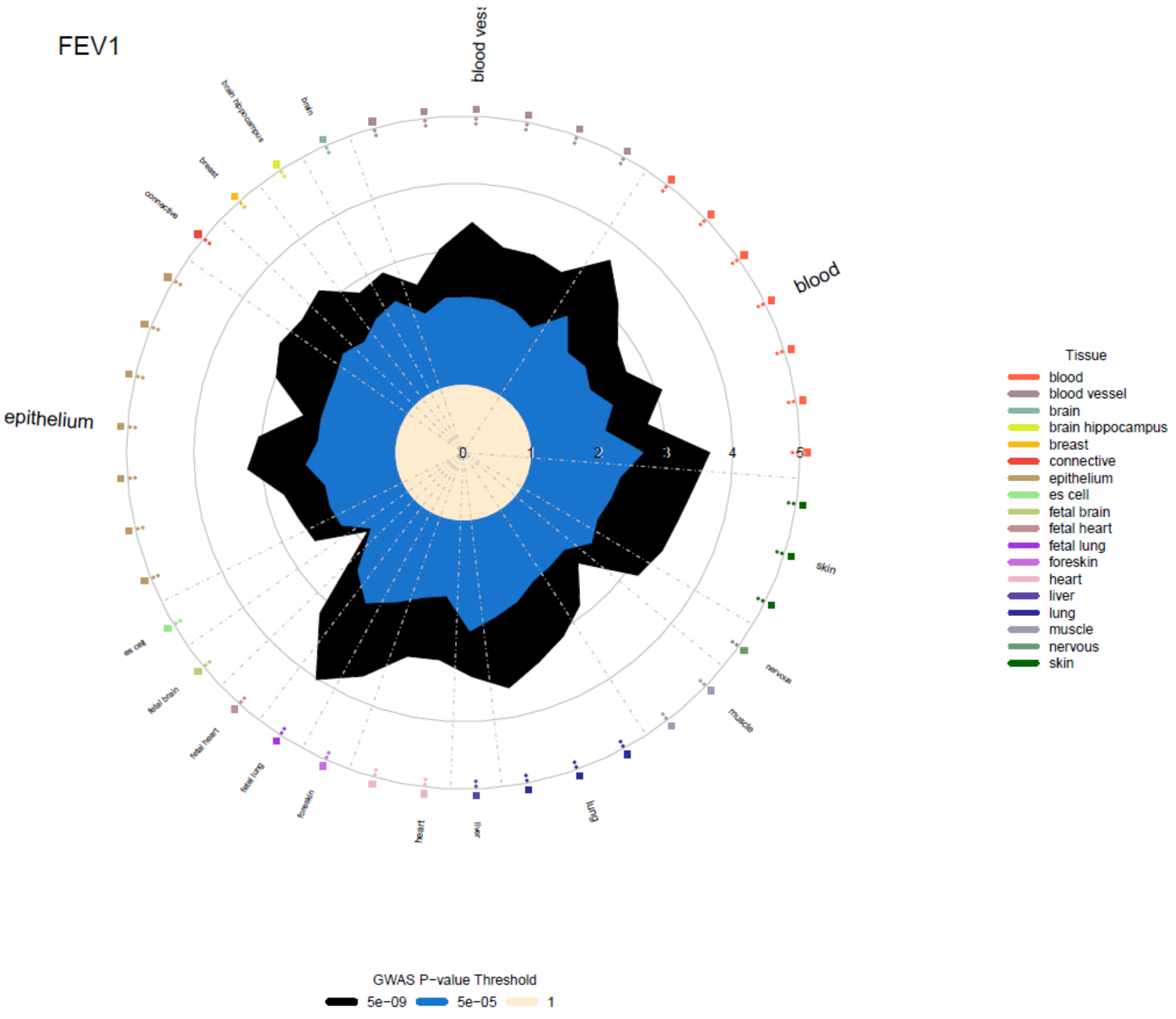

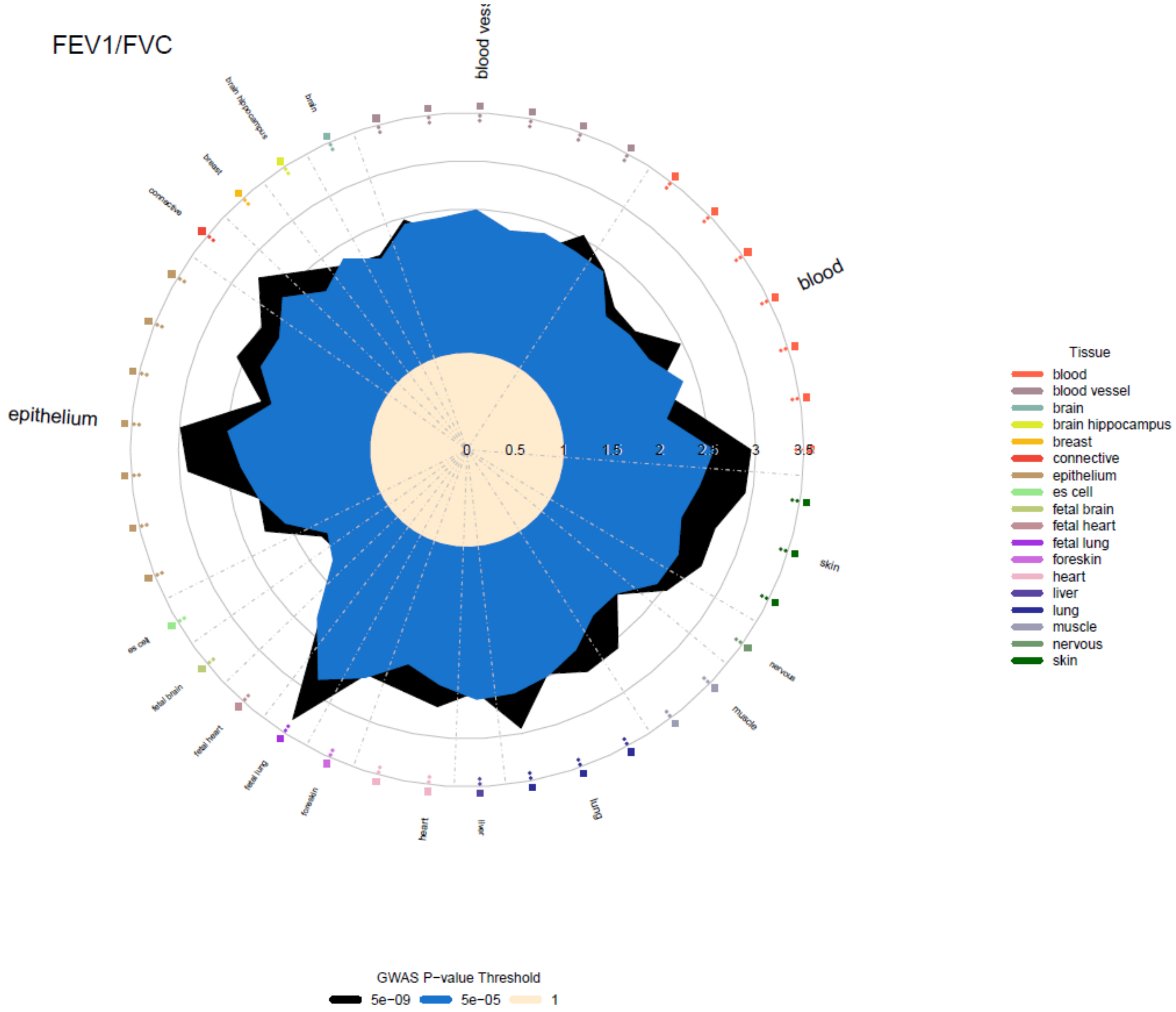
FEV1/FVC
blood vess
blood
epithelium
skin
nervous
muscle
lung
liver
heart
foreskin
fetal lung
fetal heart
fetal brain
es cell
connective
breast
brain hippocampus
brain
0
0.5
1
1.5
2
2.5
3
3.5
Tissue
blood
blood vessel
brain
brain hippocampus
breast
connective
epithelium
es cell
fetal brain
fetal heart
fetal lung
foreskin
heart
liver
lung
muscle
nervous
skin
GWAS P−value Threshold
5e−09
5e−05
1

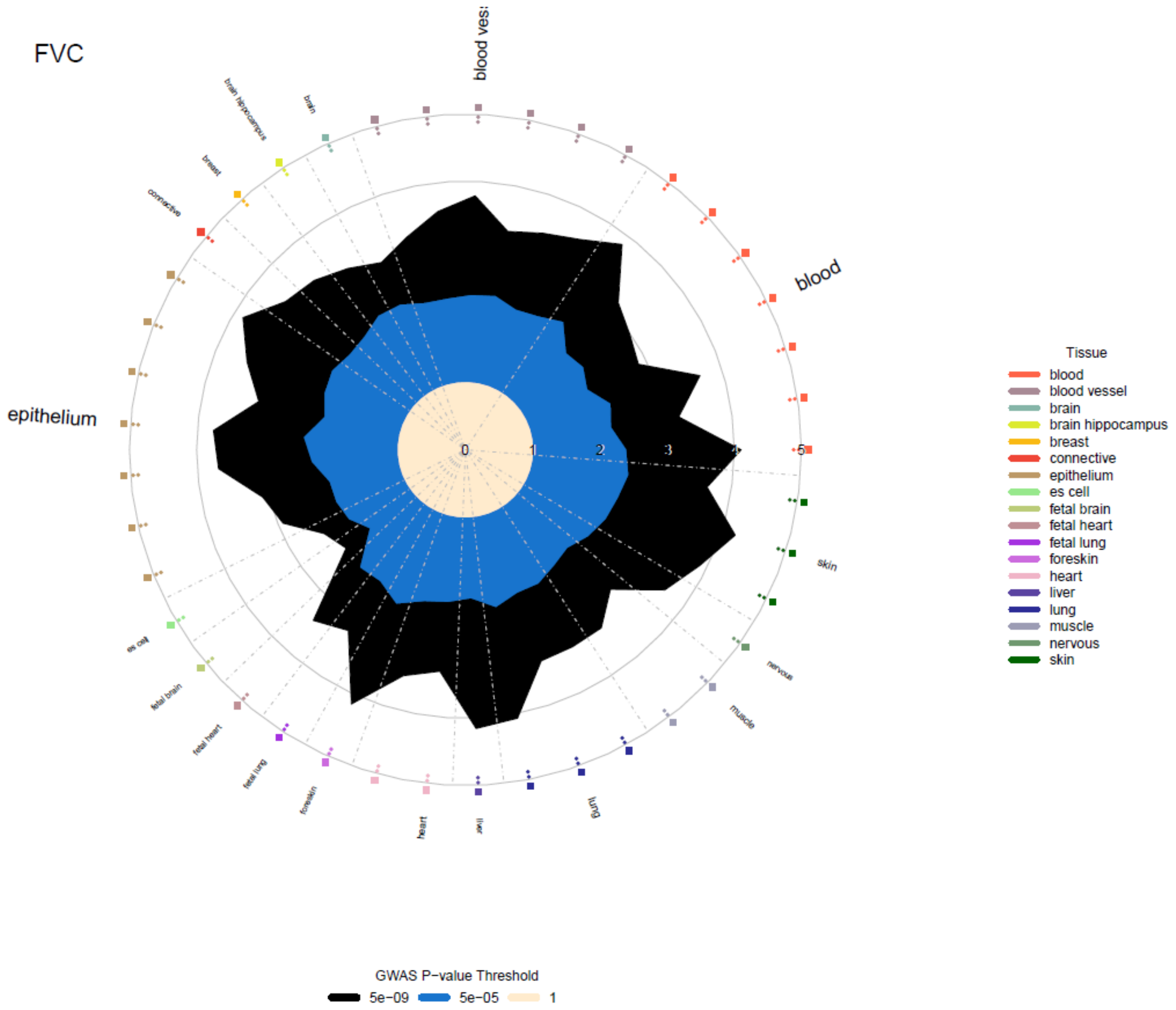
FVC
blood vessel
brain hippocampus
brain
breast
connective
epithelium
blood
skin
nervous
muscle
lung
liver
heart
foreskin
fetal lung
fetal heart
fetal brain
es cell
0
1
2
3
4
5
Tissue
blood
blood vessel
brain
brain hippocampus
breast
connective
epithelium
es cell
fetal brain
fetal heart
fetal lung
foreskin
heart
liver
lung
muscle
nervous
skin
GWAS P−value Threshold
5e−09
5e−05
1

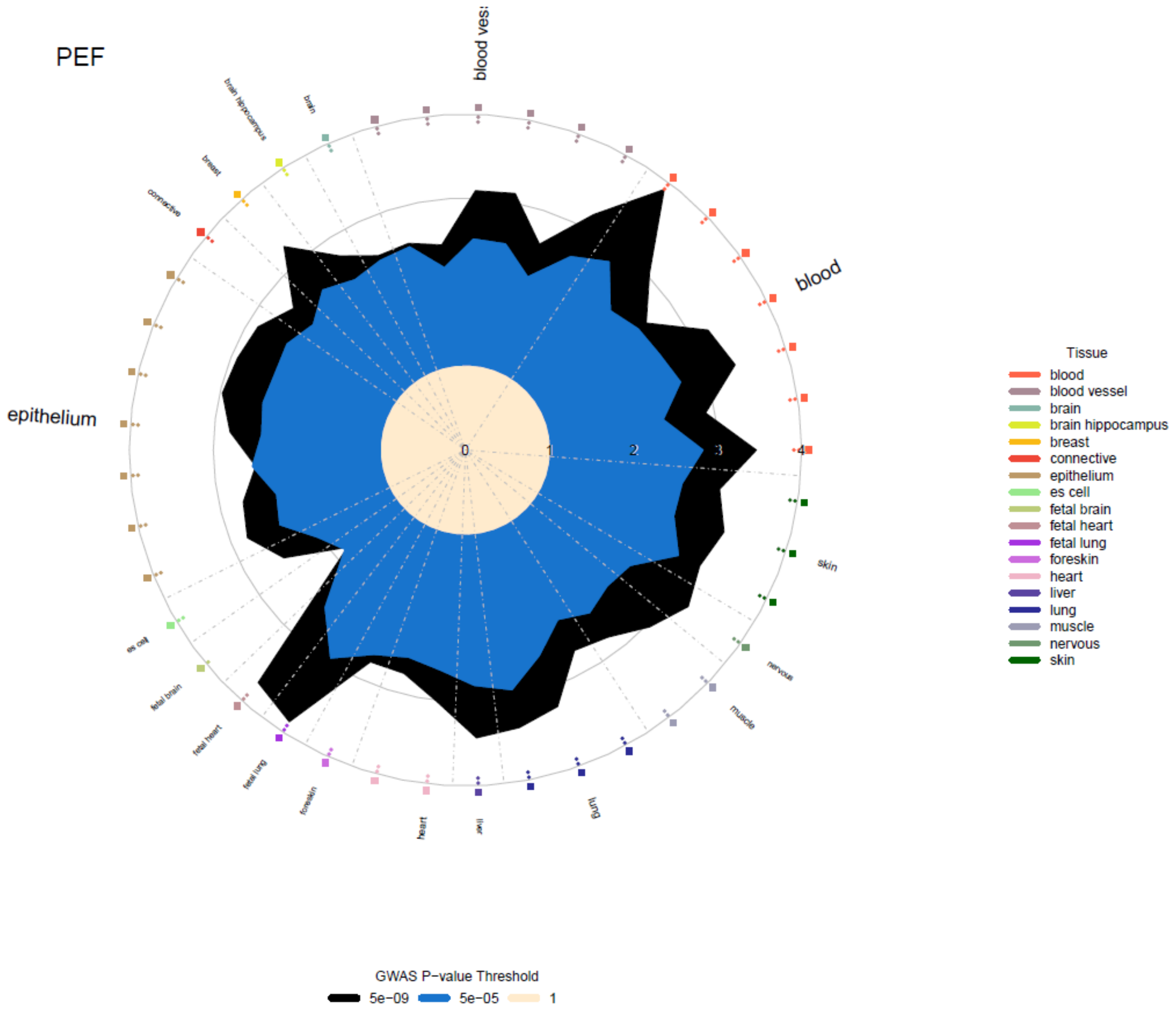

PEF
blood ves:
blood
epithelium
skin
nervous
muscle
lung
liver
heart
foreskin
fetal lung
fetal heart
fetal brain
es cell
connective
breast
brain hippocampus
brain
0
1
2
3
4
Tissue
blood
blood vessel
brain
brain hippocampus
breast
connective
epithelium
es cell
fetal brain
fetal heart
fetal lung
foreskin
heart
liver
lung
muscle
nervous
skin
GWAS P−value Threshold
5e−09
5e−05
1

c) Genic

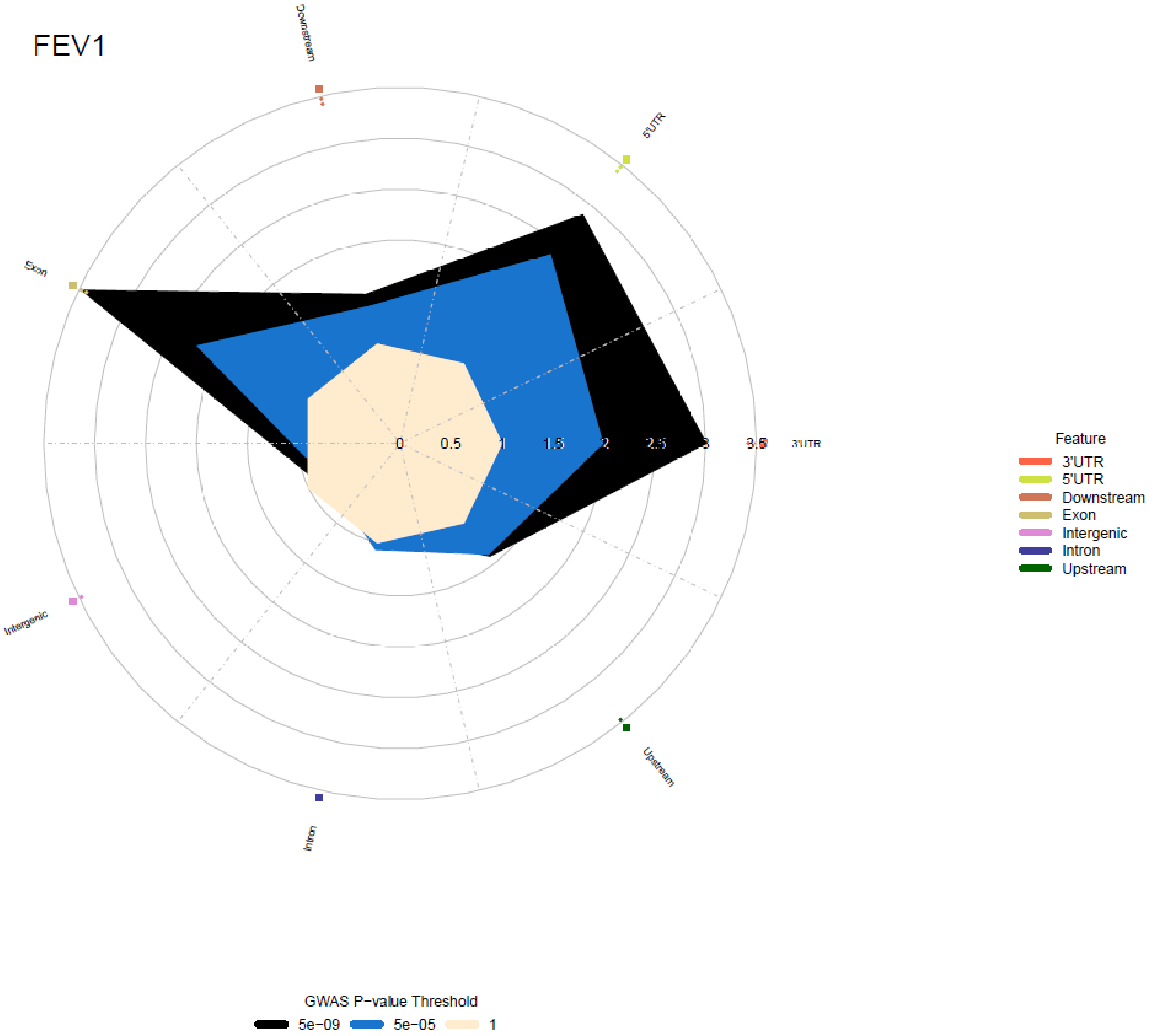

FEV1
Downstream
5'UTR
Exon
3'UTR
Intergenic
Intron
Upstream
0
0.5
1
1.5
2
2.5
3
3.5
Feature
3'UTR
5'UTR
Downstream
Exon
Intergenic
Intron
Upstream
GWAS P−value Threshold
5e−09
5e−05
1

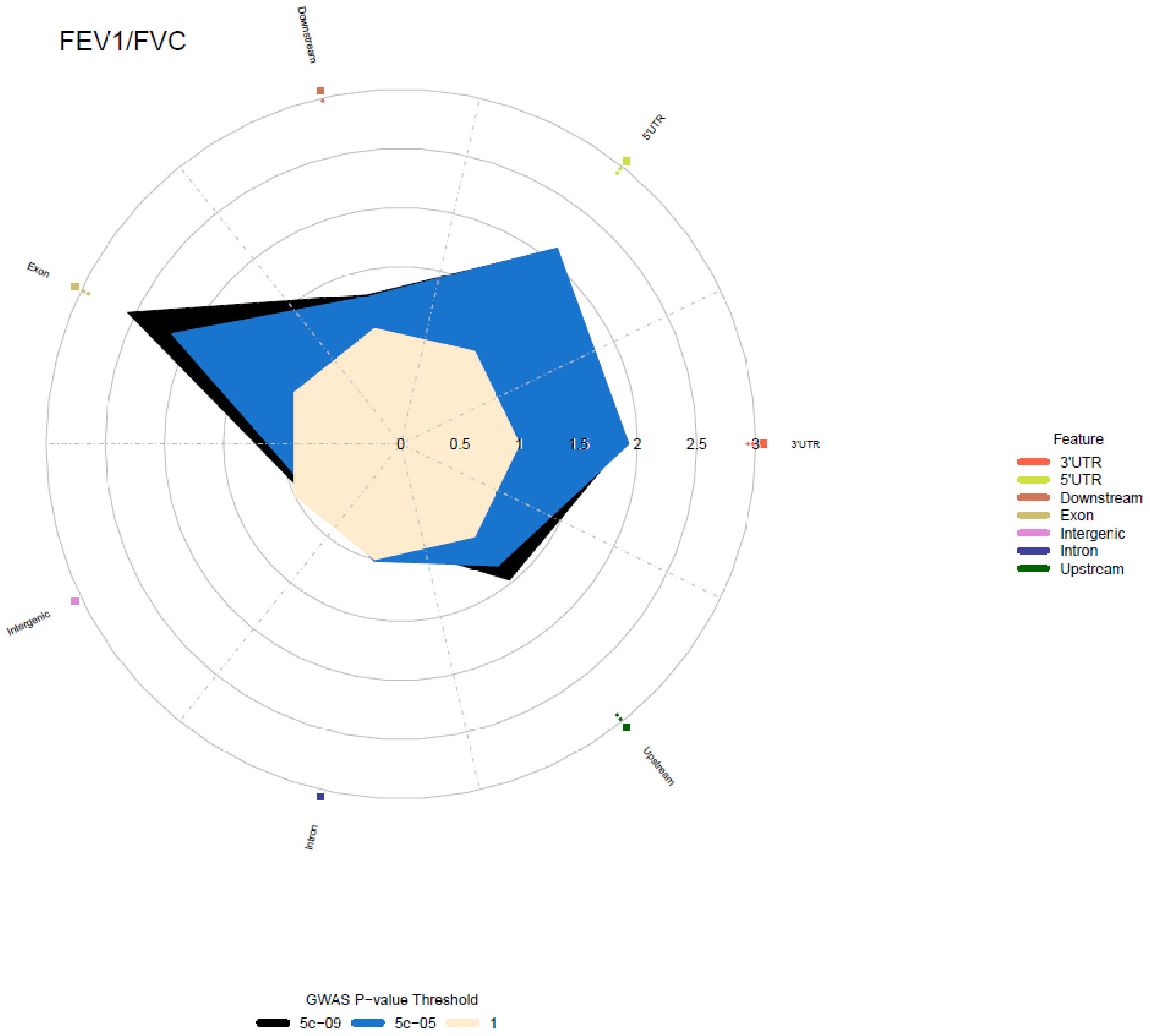
FEV1/FVC
Downstream
5'UTR
Exon
3'UTR
Intergenic
Upstream
Intron
0
0.5
1
1.5
2
2.5
3
Feature
3'UTR
5'UTR
Downstream
Exon
Intergenic
Intron
Upstream
GWAS P−value Threshold
5e−09
5e−05
1

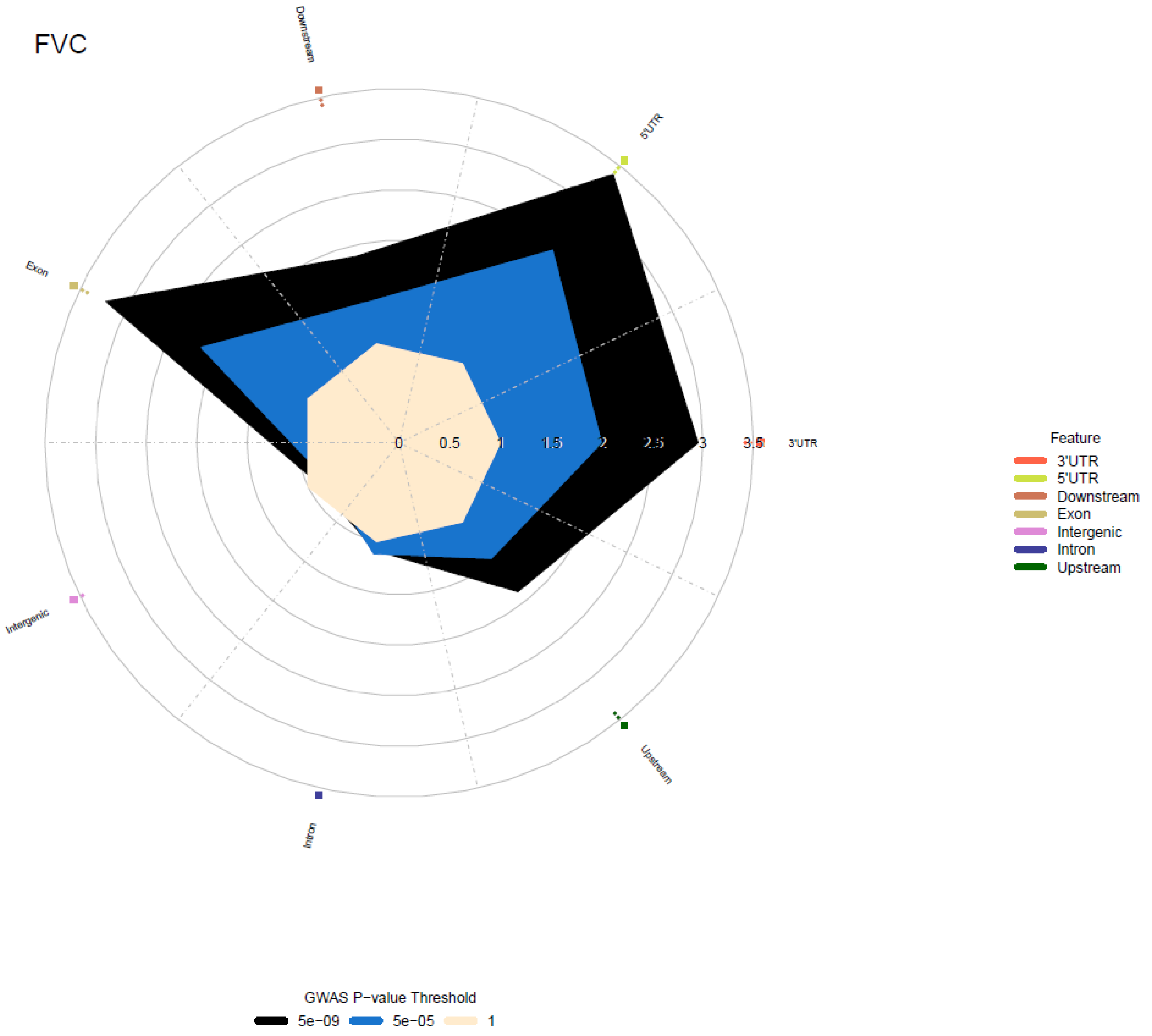
FVC
Downstream
5'UTR
Exon
3'UTR
Intergenic
Upstream
Intron
0
0.5
1
1.5
2
2.5
3
3.5
Feature
3'UTR
5'UTR
Downstream
Exon
Intergenic
Intron
Upstream
GWAS P-value Threshold
5e-09
5e-05
1

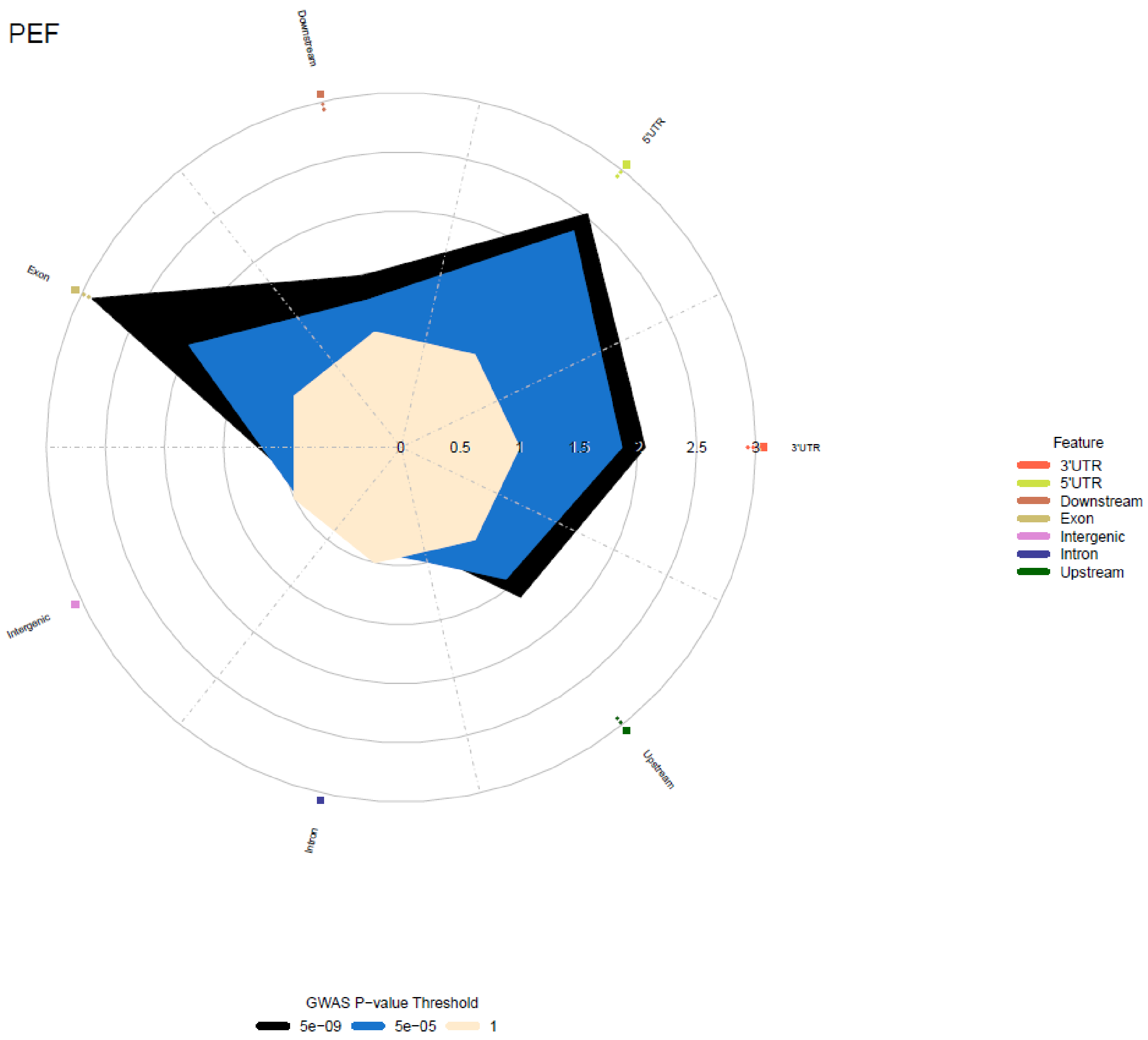

PEF
Downstream
5'UTR
Exon
3'UTR
Intergenic
Intron
Upstream
0
0.5
1
1.5
2
2.5
3
Feature
3'UTR
5'UTR
Downstream
Exon
Intergenic
Intron
Upstream
GWAS P−value Threshold
5e−09
5e−05
1

d) Chromatin states

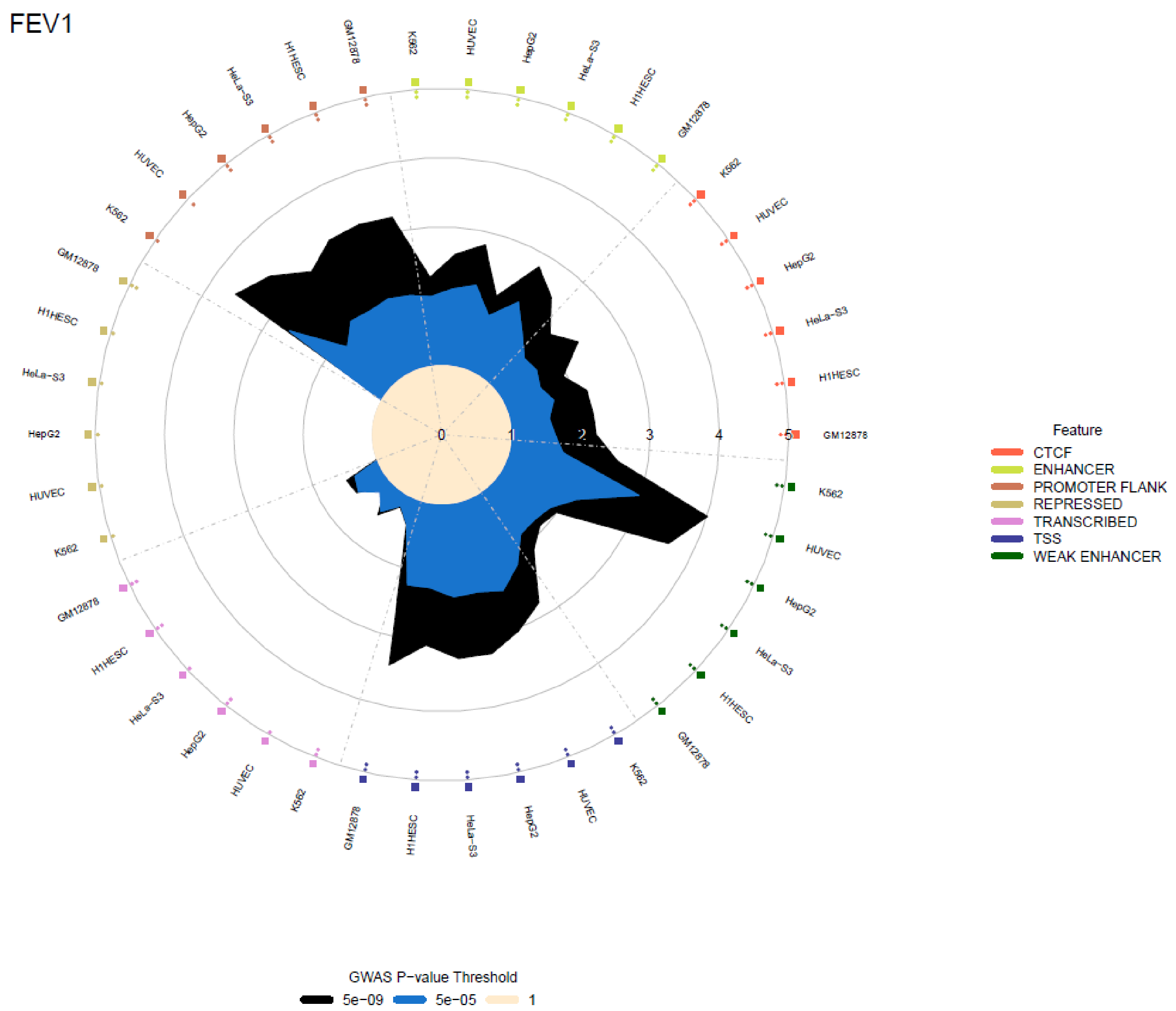

FEV1/FVC

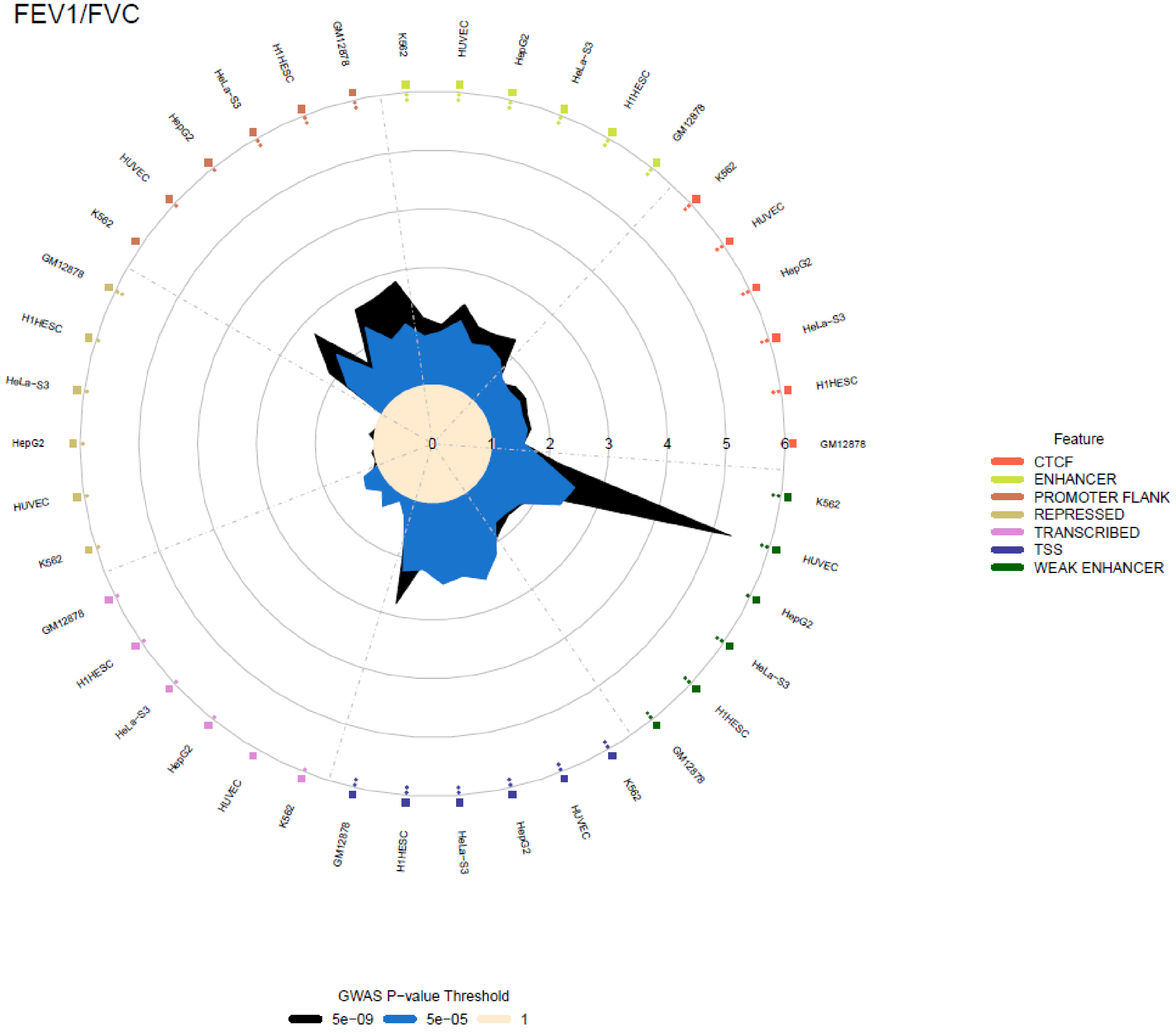
Feature
CTCF
ENHANCER
PROMOTER FLANK
REPRESSED
TRANSCRIBED
TSS
WEAK ENHANCER
GWAS P−value Threshold
5e−09
5e−05
1
GM12878
K562
HUVEC
HepG2
HeLa−S3
H1HESC
0
1
2
3
4
5
6

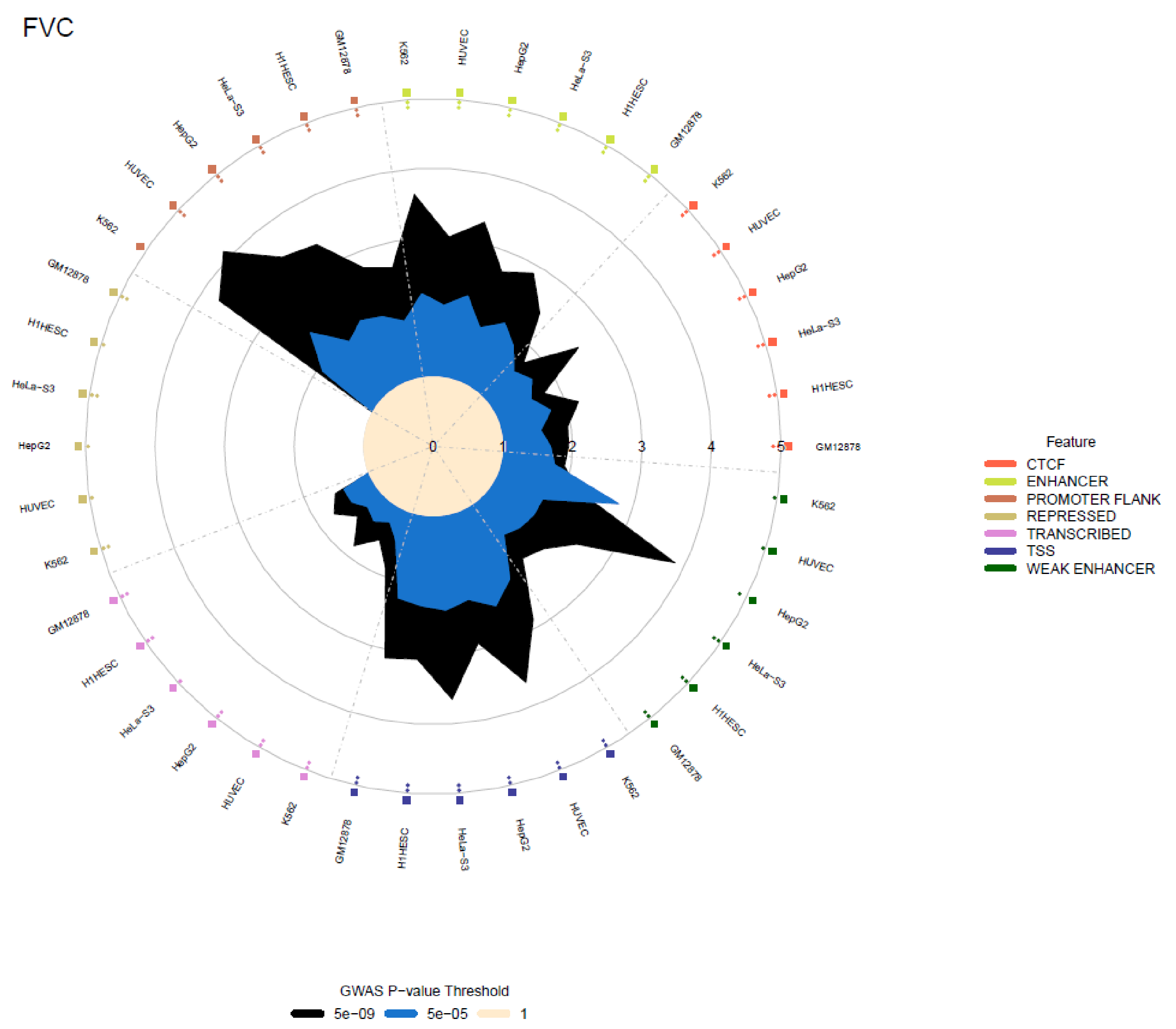
FVC
Feature
CTCF
ENHANCER
PROMOTER FLANK
REPRESSED
TRANSCRIBED
TSS
WEAK ENHANCER
GWAS P−value Threshold
5e−09
5e−05
1
0
1
2
3
4
5
GM12878
H1HESC
HeLa−S3
HepG2
HUVEC
K562

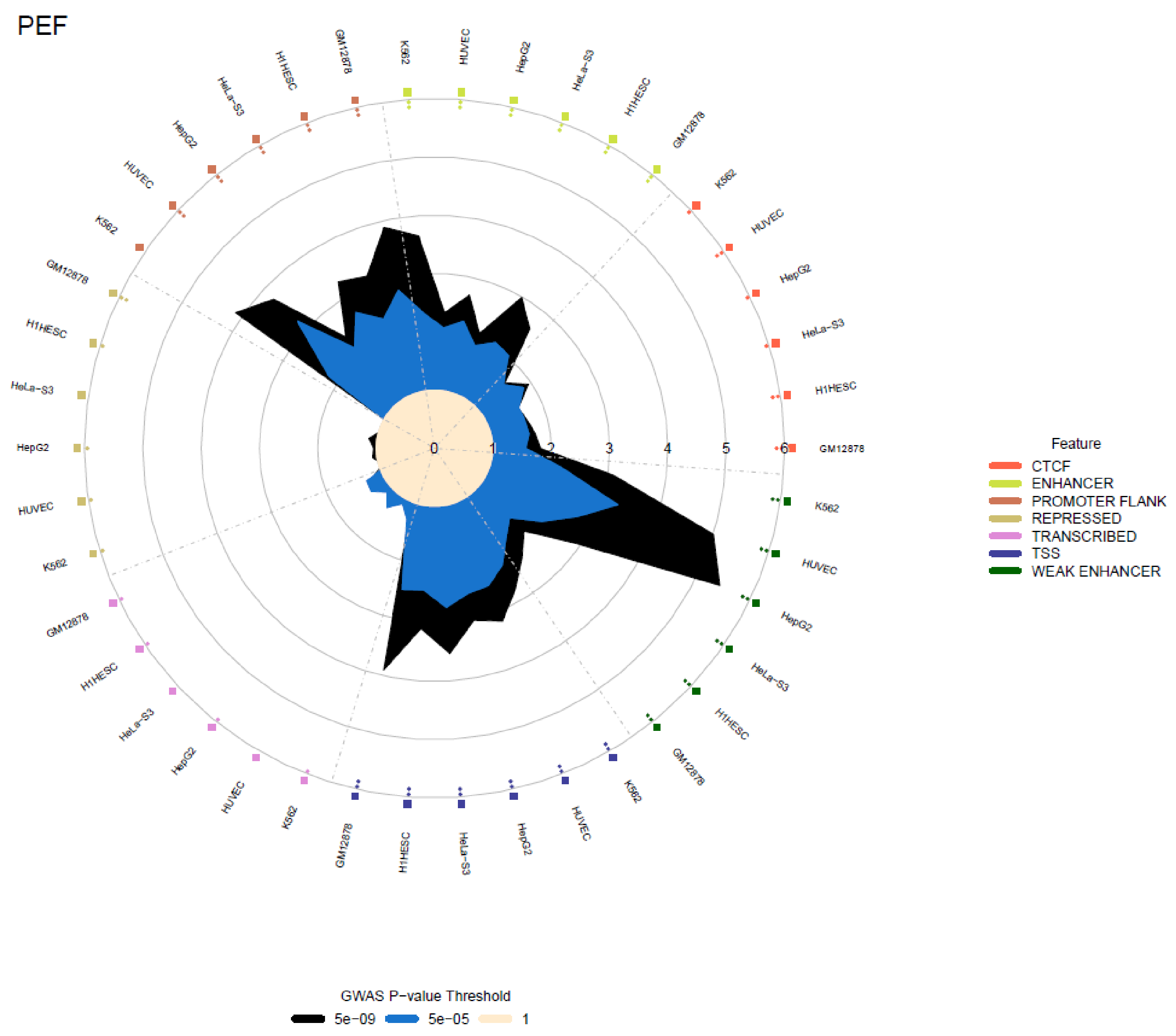
PEF
0
1
2
3
4
5
6
GM12878
K562
HUVEC
HepG2
HeLa-S3
H1HESC
Feature
CTCF
ENHANCER
PROMOTER FLANK
REPRESSED
TRANSCRIBED
TSS
WEAK ENHANCER
GWAS P-value Threshold
5e-09
5e-05
1

**e) Transcription factor binding sites**

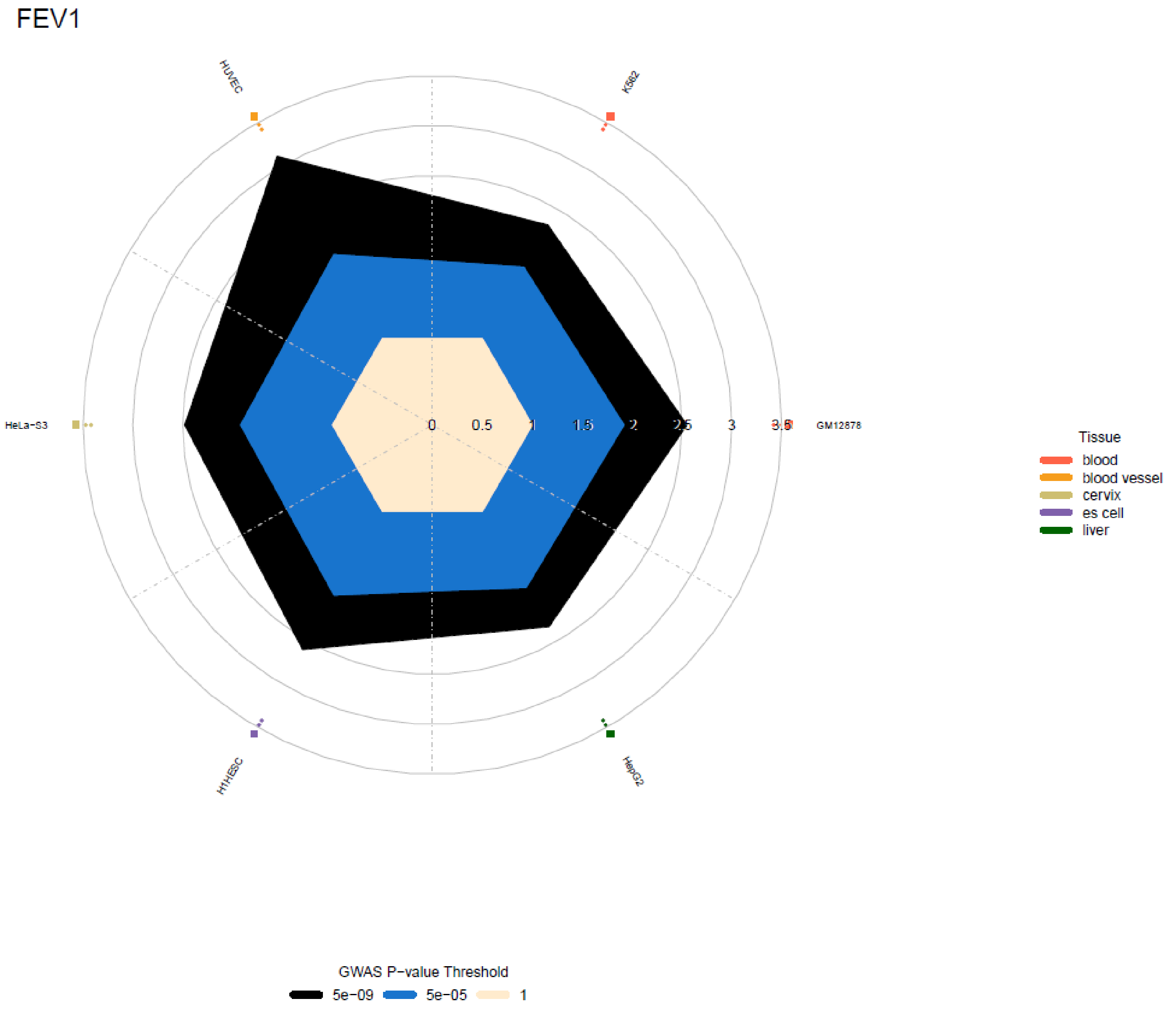

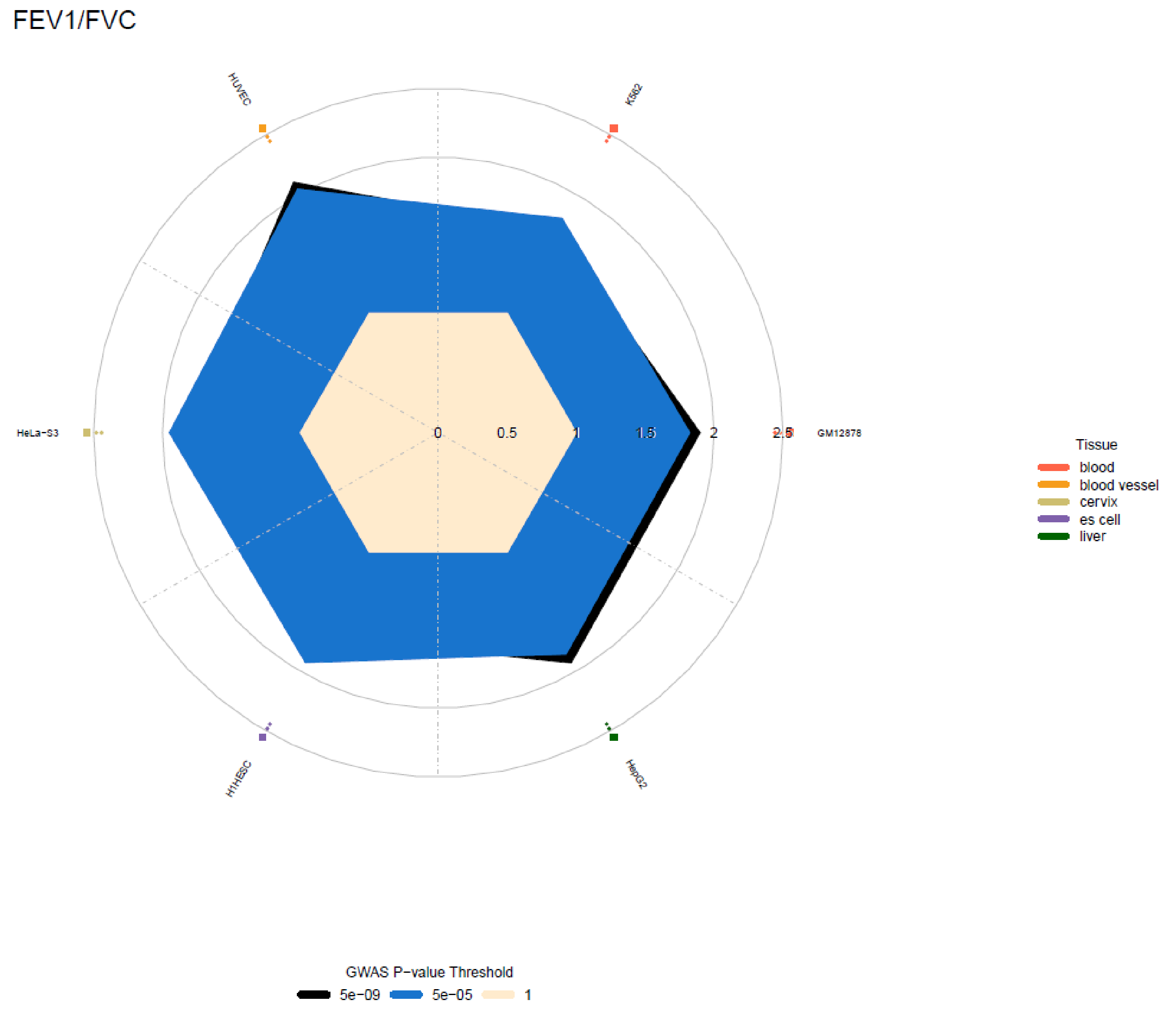
FEV1/FVC
HUVEC
K562
HeLa−S3
GM12878
H1HESC
HepG2
0
0.5
1
1.5
2
2.5
Tissue
blood
blood vessel
cervix
es cell
liver
GWAS P−value Threshold
5e−09
5e−05
1

### FVC

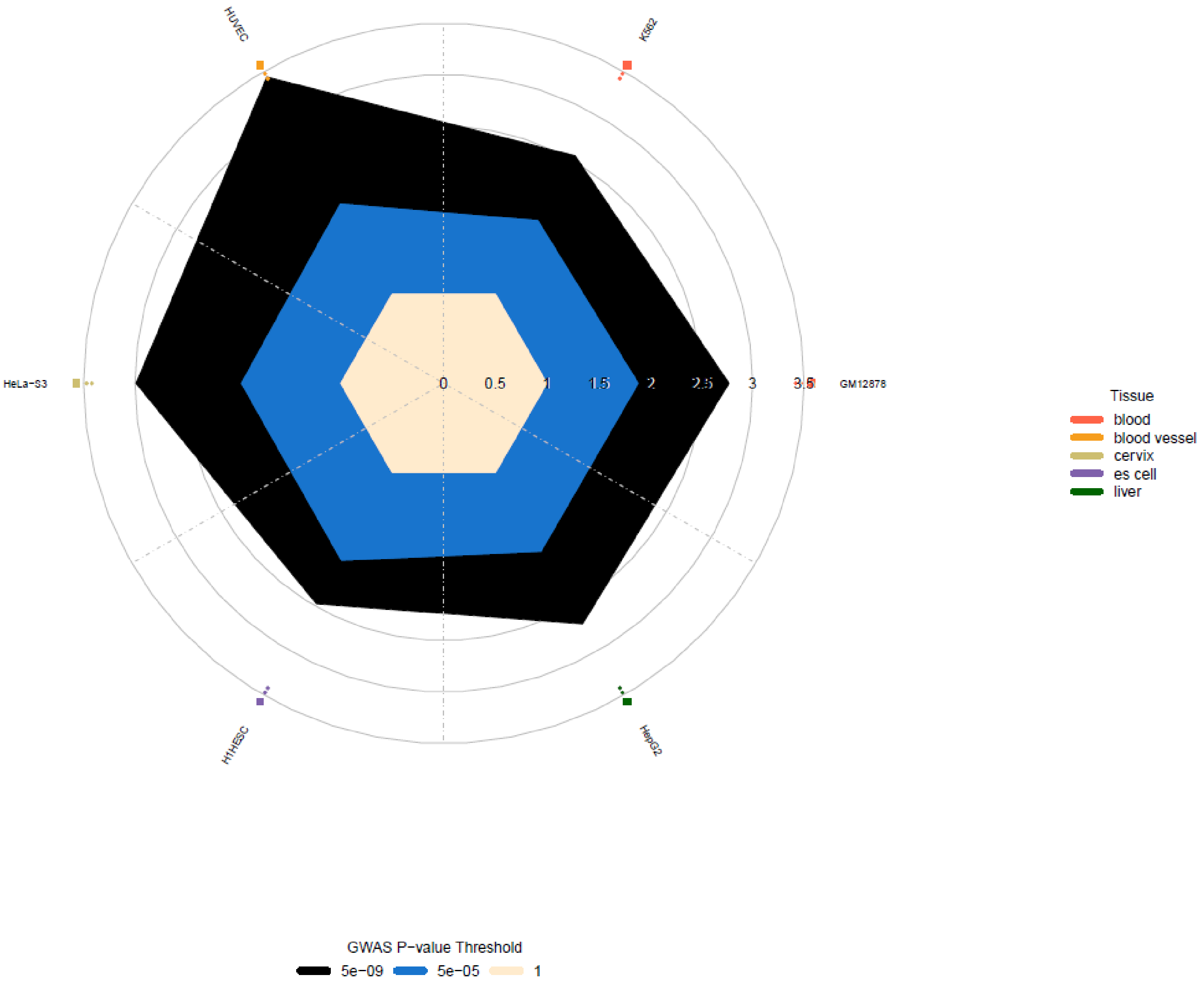

HUVEC
K562
HeLa−S3
GM12878
H1HESC
HepG2
0
0.5
1
1.5
2
2.5
3
3.5
Tissue
blood
blood vessel
cervix
es cell
liver
GWAS P−value Threshold
5e−09
5e−05
1

PEF

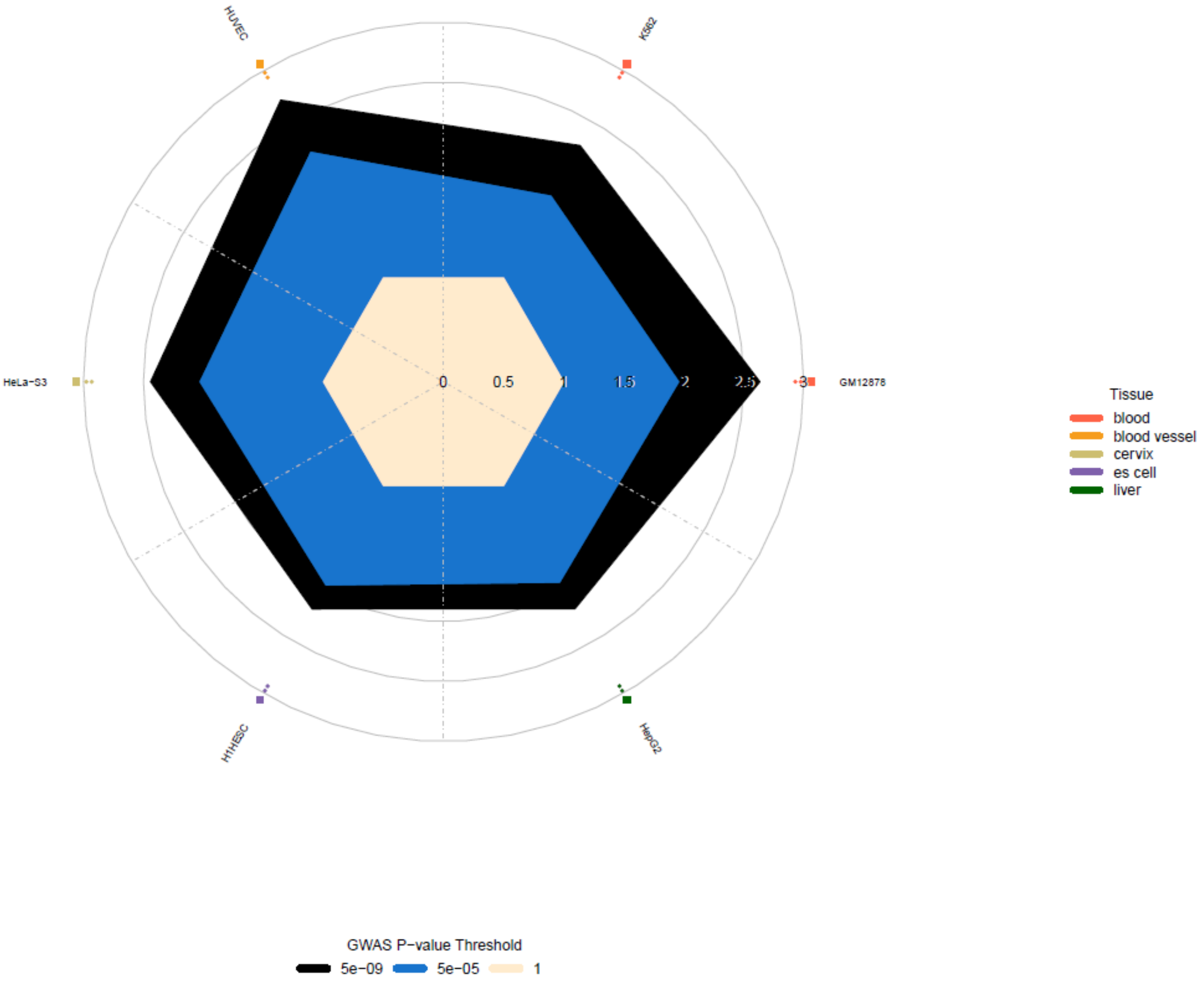

HUVEC
K562
HeLa−S3
GM12878
H1HESC
HepG2
0
0.5
1
1.5
2
2.5
3
Tissue
blood
blood vessel
cervix
es cell
liver
GWAS P−value Threshold
5e−09
5e−05
1

#### f) Open chromatin peaks

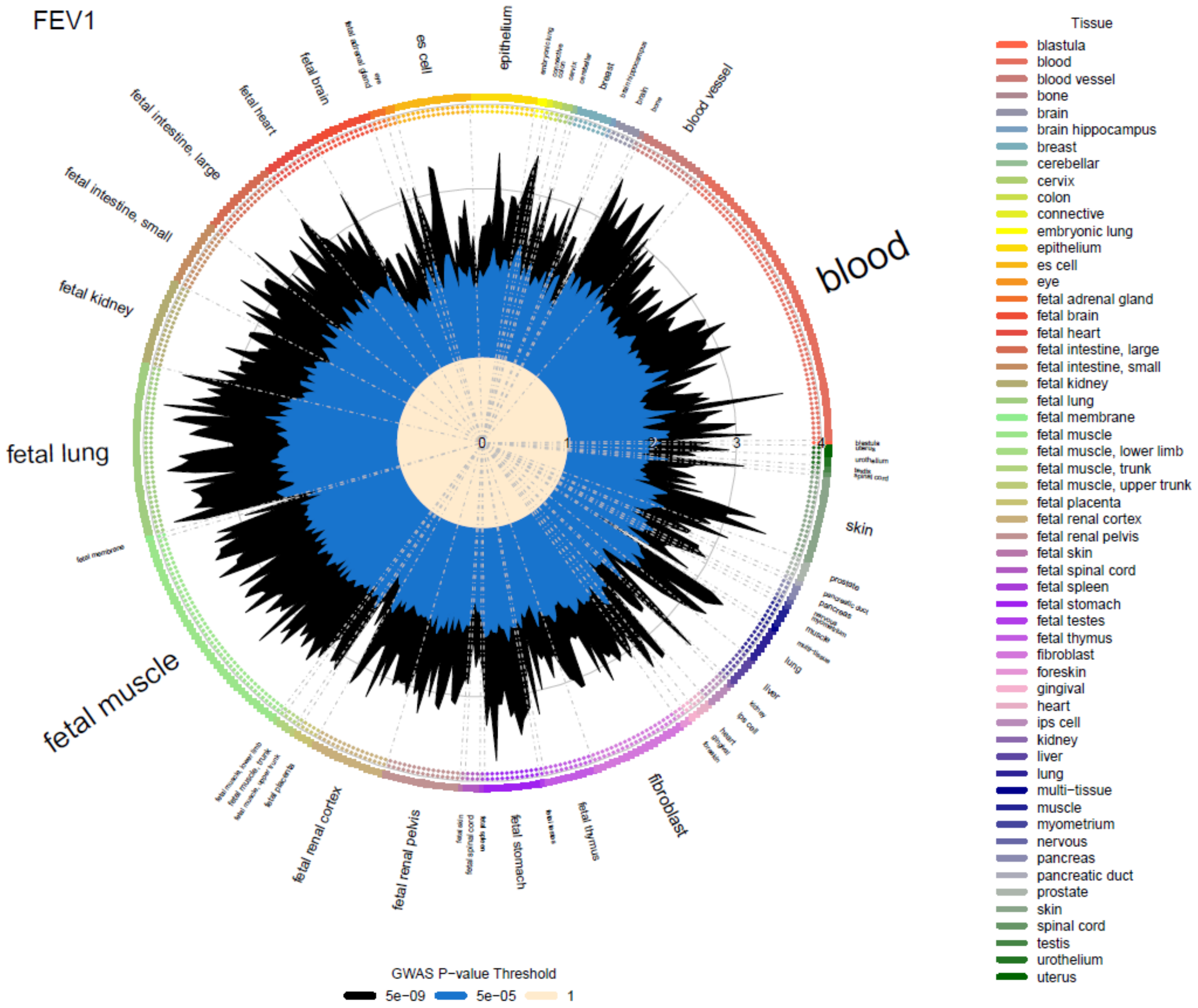

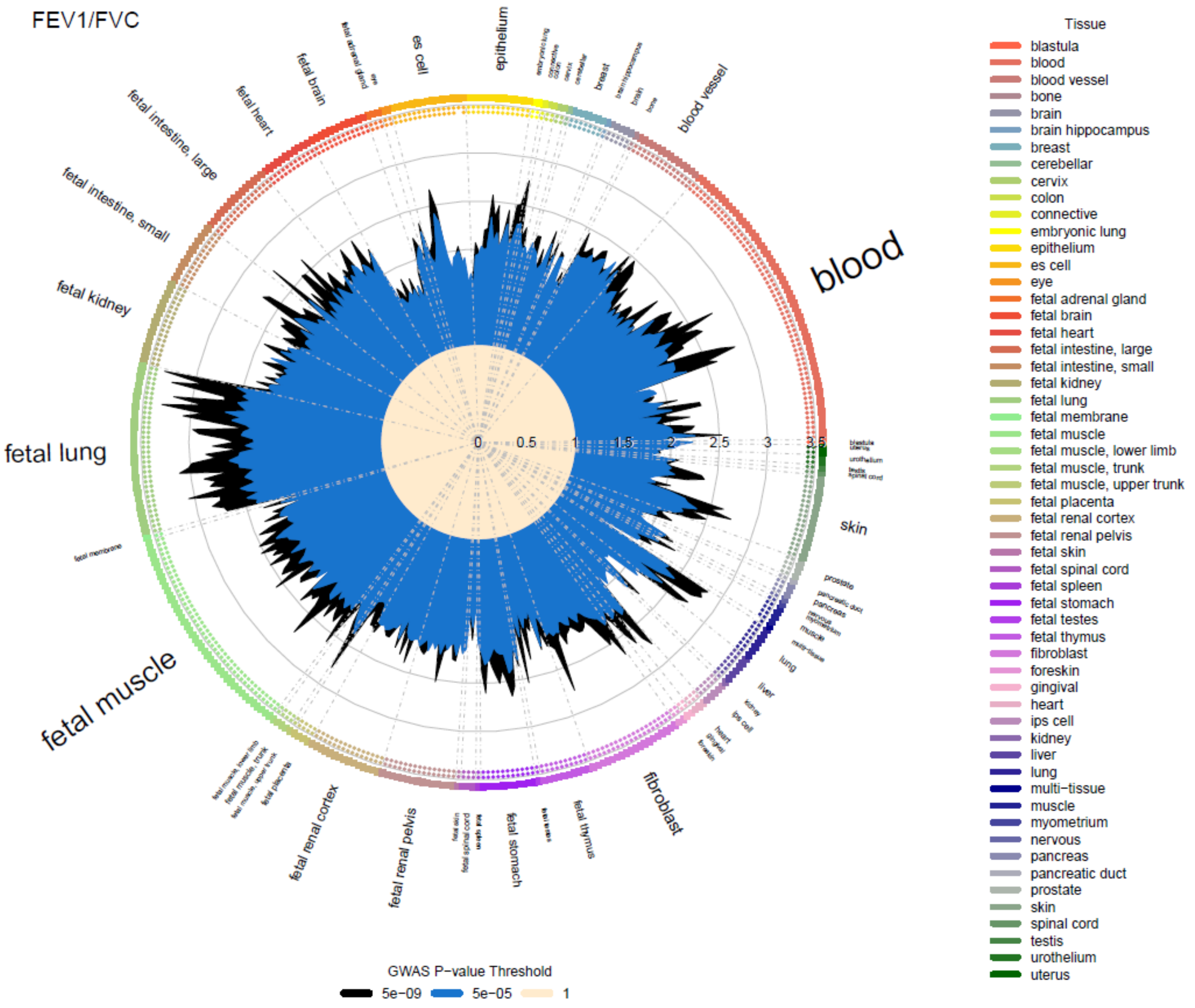
FEV1/FVC
blood
fetal lung
fetal muscle
epithelium
es cell
fetal brain
fetal heart
fetal intestine, large
fetal intestine, small
fetal kidney
blood vessel
breast
skin
fibroblast
fetal renal cortex
fetal renal pelvis
fetal stomach
fetal thymus
0
0.5
1
1.5
2
2.5
3
3.5
GWAS P−value Threshold
5e−09
5e−05
1
Tissue
blastula
blood
blood vessel
bone
brain
brain hippocampus
breast
cerebellar
cervix
colon
connective
embryonic lung
epithelium
es cell
eye
fetal adrenal gland
fetal brain
fetal heart
fetal intestine, large
fetal intestine, small
fetal kidney
fetal lung
fetal membrane
fetal muscle
fetal muscle, lower limb
fetal muscle, trunk
fetal muscle, upper trunk
fetal placenta
fetal renal cortex
fetal renal pelvis
fetal skin
fetal spinal cord
fetal spleen
fetal stomach
fetal testes
fetal thymus
fibroblast
foreskin
gingival
heart
ips cell
kidney
liver
lung
multi−tissue
muscle
myometrium
nervous
pancreas
pancreatic duct
prostate
skin
spinal cord
testis
urothelium
uterus

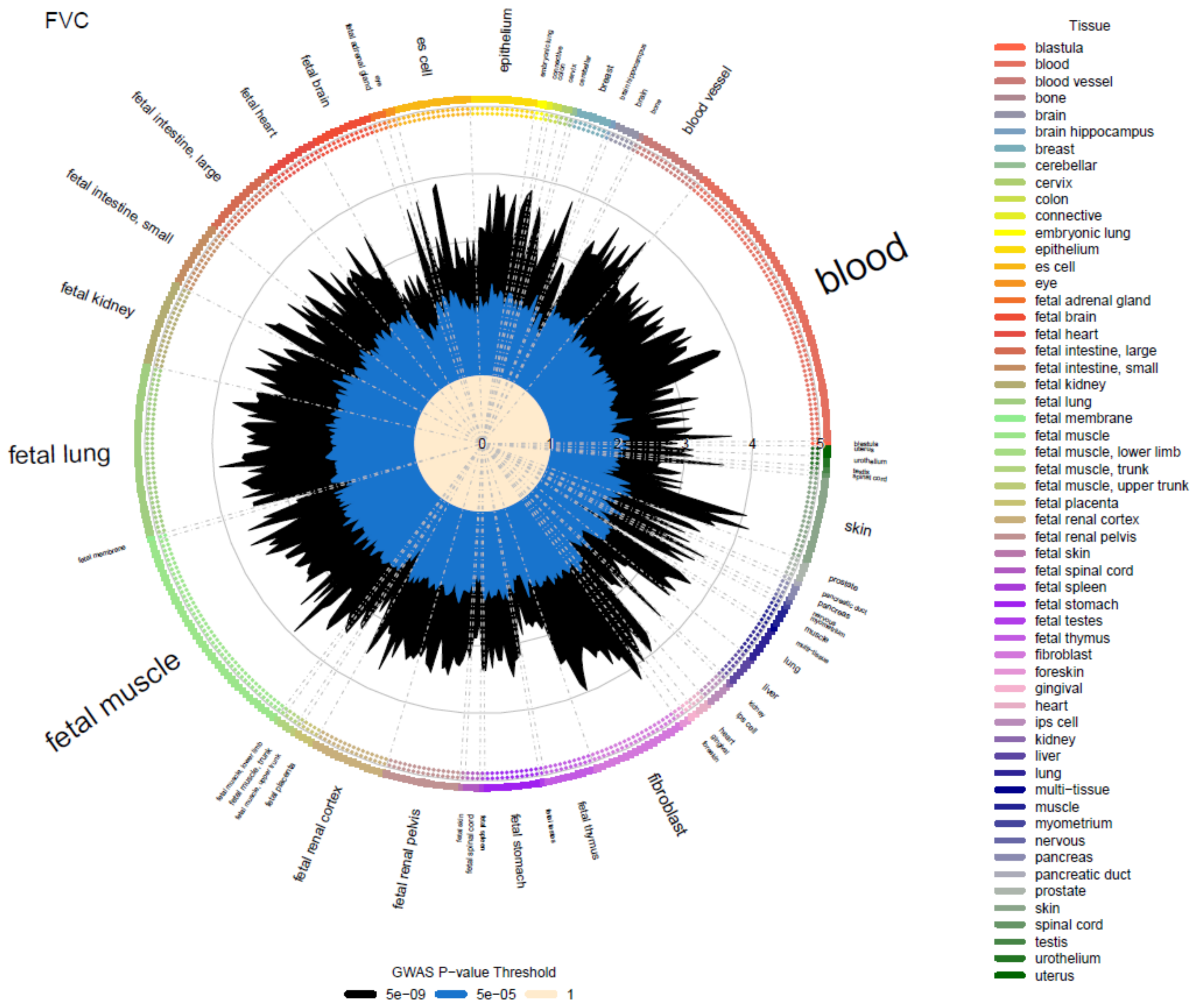
FVC
blood
fetal lung
fetal muscle
fibroblast
skin
epithelium
es cell
fetal brain
fetal heart
fetal intestine, large
fetal intestine, small
fetal kidney
blood vessel
fetal renal cortex
fetal renal pelvis
fetal stomach
fetal thymus
Tissue
blastula
blood
blood vessel
bone
brain
brain hippocampus
breast
cerebellar
cervix
colon
connective
embryonic lung
epithelium
es cell
eye
fetal adrenal gland
fetal brain
fetal heart
fetal intestine, large
fetal intestine, small
fetal kidney
fetal lung
fetal membrane
fetal muscle
fetal muscle, lower limb
fetal muscle, trunk
fetal muscle, upper trunk
fetal placenta
fetal renal cortex
fetal renal pelvis
fetal skin
fetal spinal cord
fetal spleen
fetal stomach
fetal testes
fetal thymus
fibroblast
foreskin
gingival
heart
ips cell
kidney
liver
lung
multi−tissue
muscle
myometrium
nervous
pancreas
pancreatic duct
prostate
skin
spinal cord
testis
urothelium
uterus
GWAS P-value Threshold
5e−09
5e−05
1

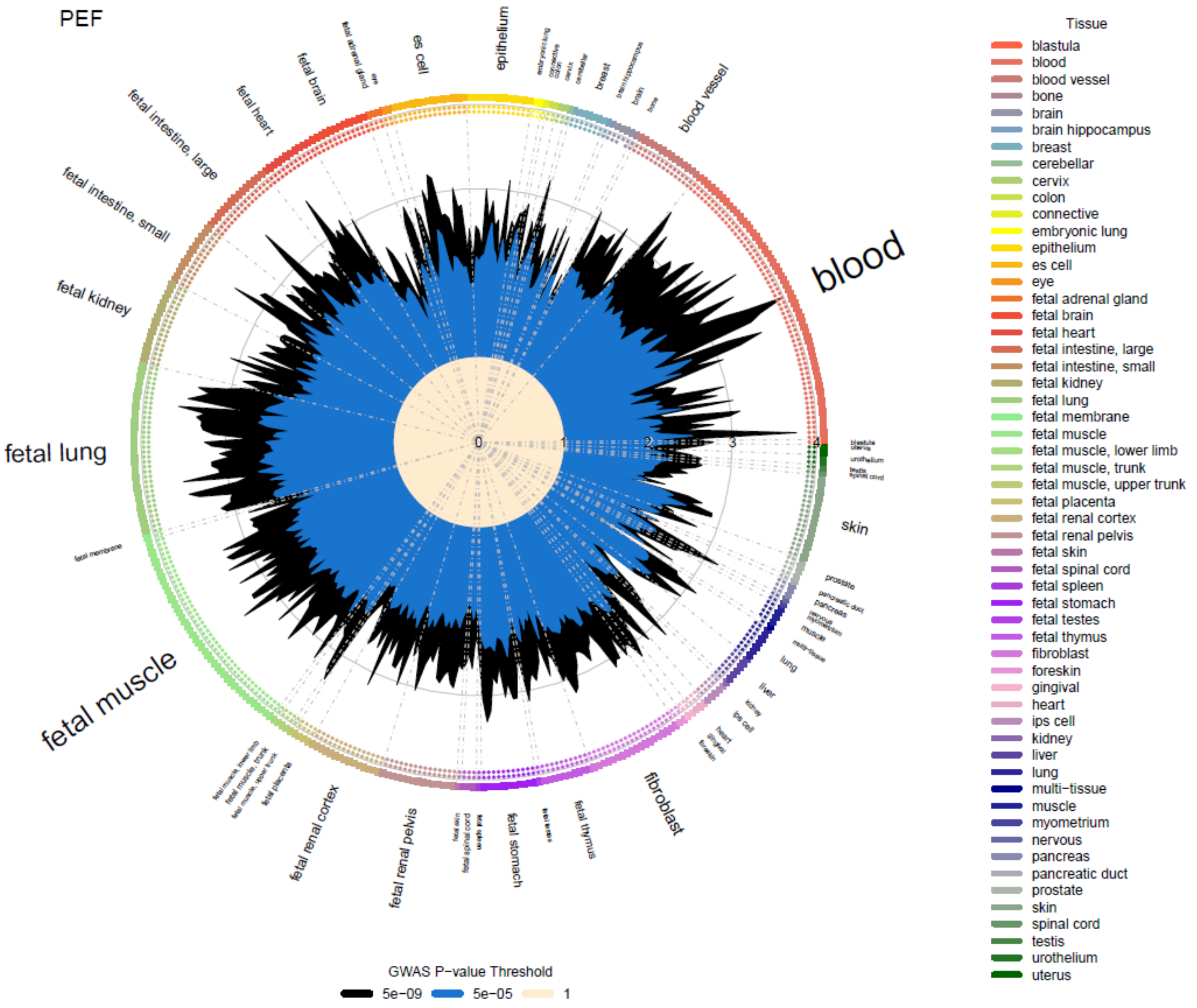
PEF
blood
fetal lung
fetal muscle
skin
fibroblast
epithelium
es cell
fetal brain
fetal heart
fetal intestine, large
fetal intestine, small
fetal kidney
blood vessel
breast
fetal renal cortex
fetal renal pelvis
fetal stomach
fetal thymus
prostate
lung
liver
fetal membrane
0
1
2
3
4
GWAS P−value Threshold
5e−09
5e−05
1
Tissue
blastula
blood
blood vessel
bone
brain
brain hippocampus
breast
cerebellar
cervix
colon
connective
embryonic lung
epithelium
es cell
eye
fetal adrenal gland
fetal brain
fetal heart
fetal intestine, large
fetal intestine, small
fetal kidney
fetal lung
fetal membrane
fetal muscle
fetal muscle, lower limb
fetal muscle, trunk
fetal muscle, upper trunk
fetal placenta
fetal renal cortex
fetal renal pelvis
fetal skin
fetal spinal cord
fetal spleen
fetal stomach
fetal testes
fetal thymus
fibroblast
foreskin
gingival
heart
ips cell
kidney
liver
lung
multi−tissue
muscle
myometrium
nervous
pancreas
pancreatic duct
prostate
skin
spinal cord
testis
urothelium
uterus

**g) FAIRE**

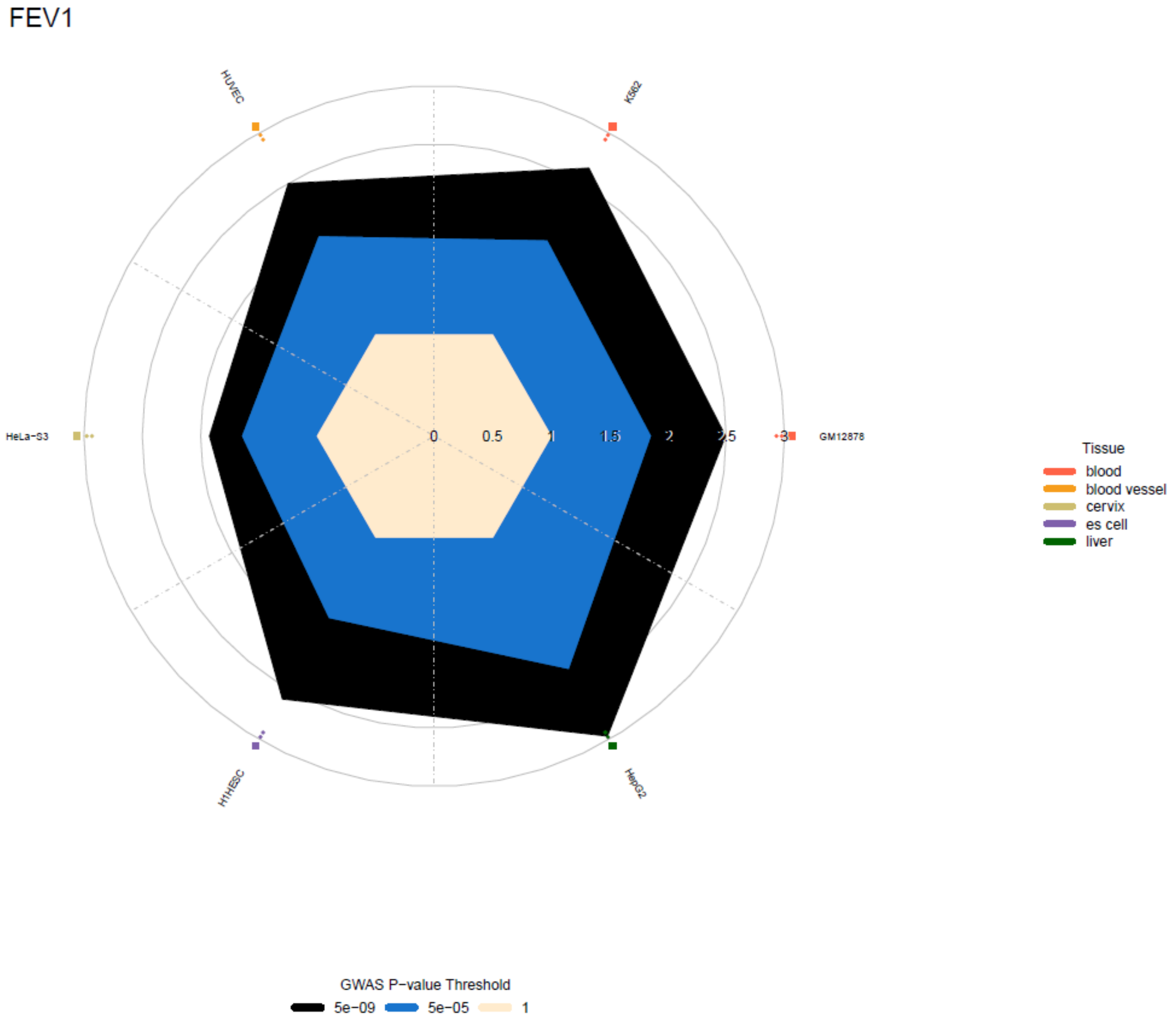
FEV1
HUVEC
K562
HeLa−S3
GM12878
H1HESC
HepG2
0
0.5
1
1.5
2
2.5
3
Tissue
blood
blood vessel
cervix
es cell
liver
GWAS P−value Threshold
5e−09
5e−05
1

### FEV1/FVC

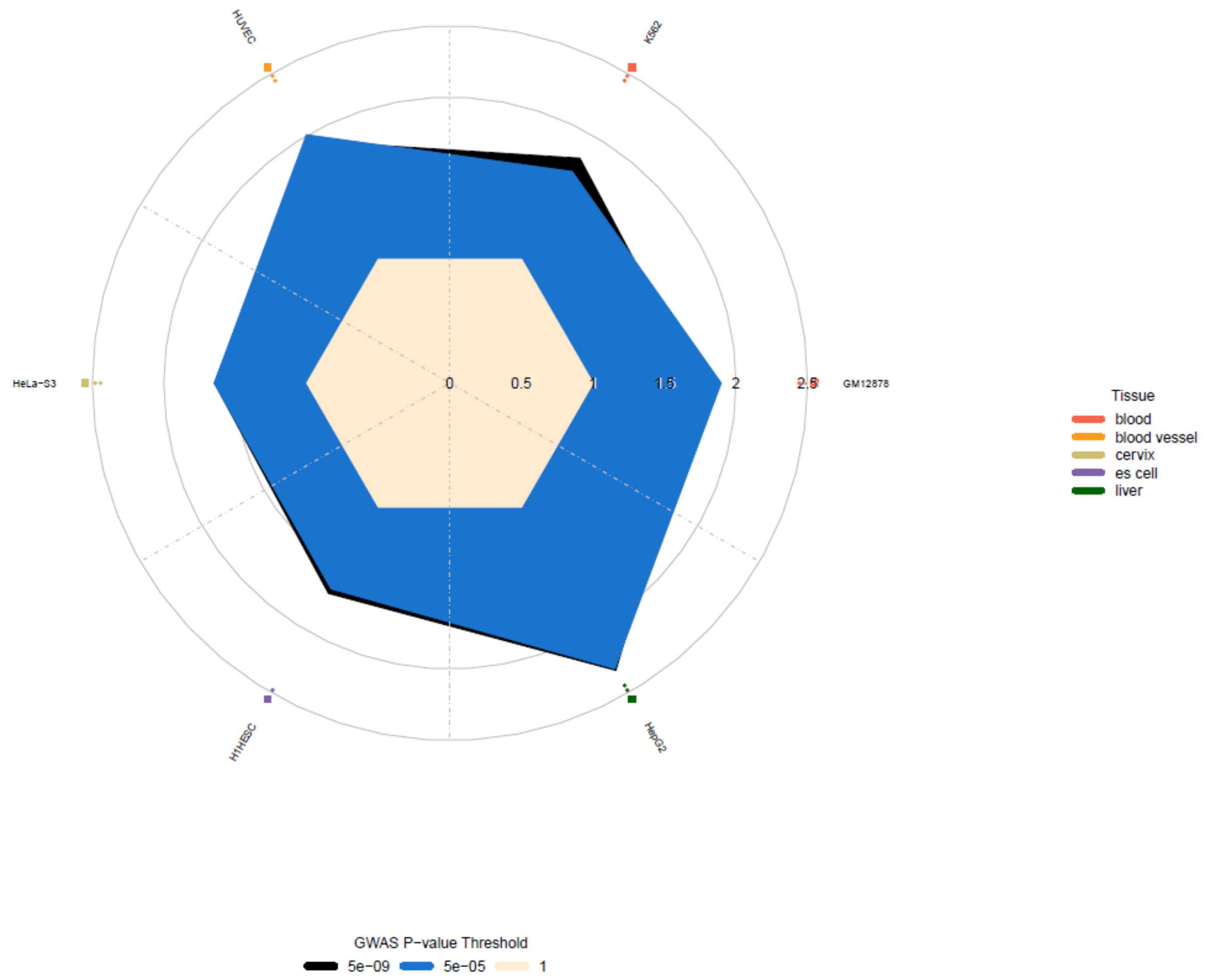

HUVEC
K562
HeLa-S3
GM12878
H1HESC
HepG2
0
0.5
1
1.5
2
2.5
Tissue
blood
blood vessel
cervix
es cell
liver
GWAS P-value Threshold
5e-09
5e-05
1

FVC

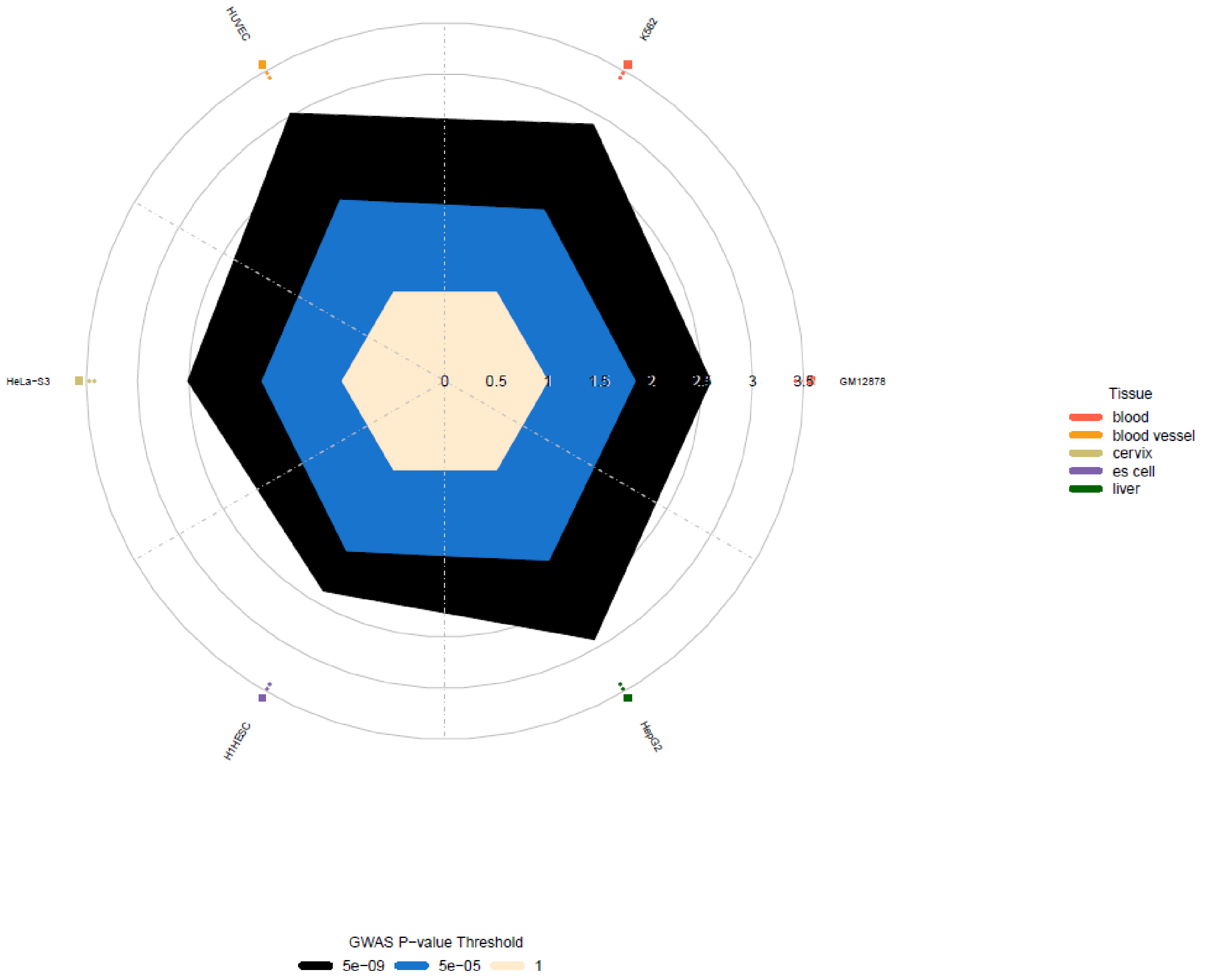
HUVEC
K562
HeLa-S3
GM12878
H1HESC
HepG2
0
0.5
1
1.5
2
2.5
3
3.5
Tissue
blood
blood vessel
cervix
es cell
liver
GWAS P−value Threshold
5e−09
5e−05
1

PEF

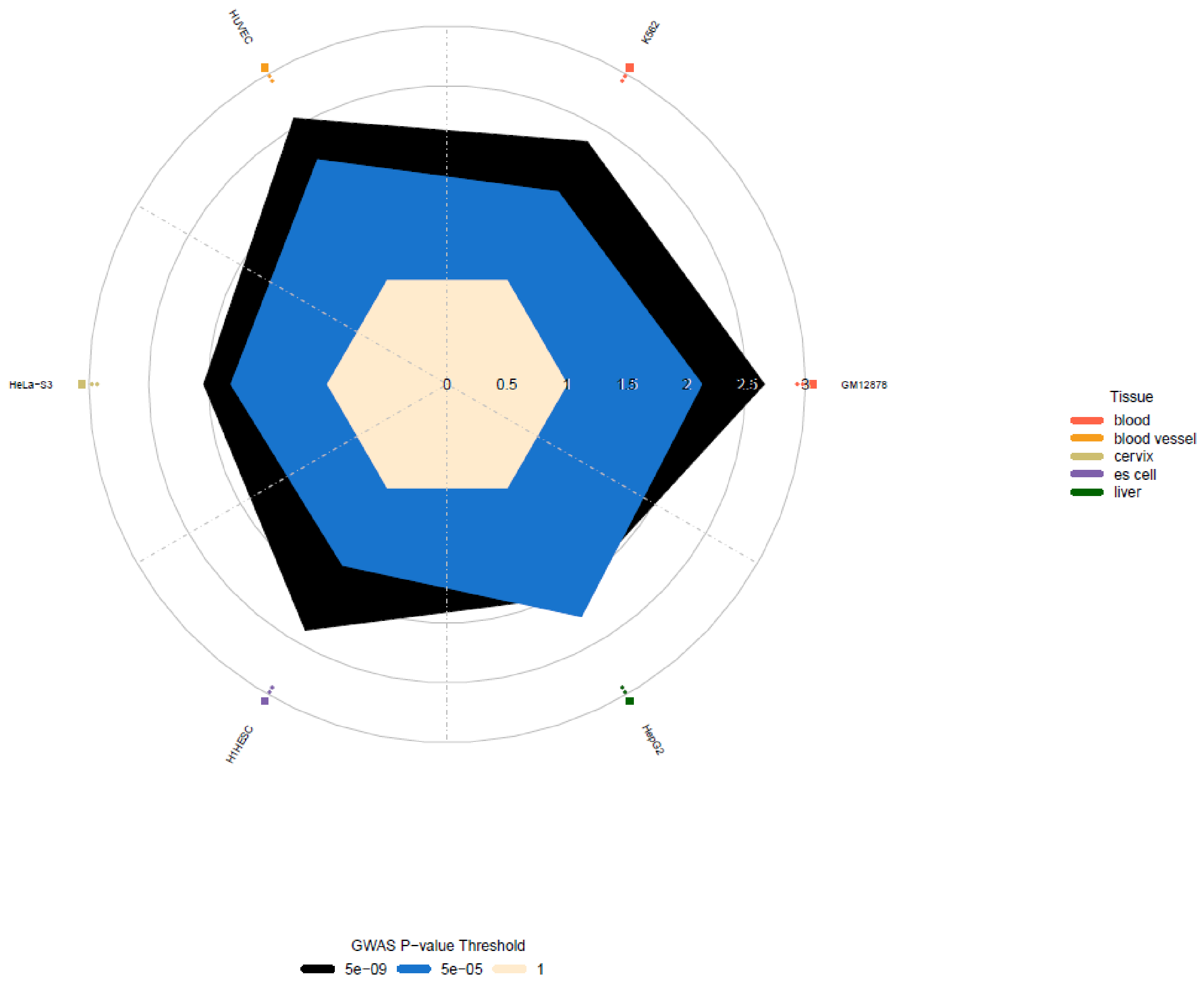

HUVEC
K562
HeLa−S3
GM12878
H1HESC
HepG2
0
0.5
1
1.5
2
2.5
3
Tissue
blood
blood vessel
cervix
es cell
liver
GWAS P−value Threshold
5e−09
5e−05
1

**h) Histone modifications**

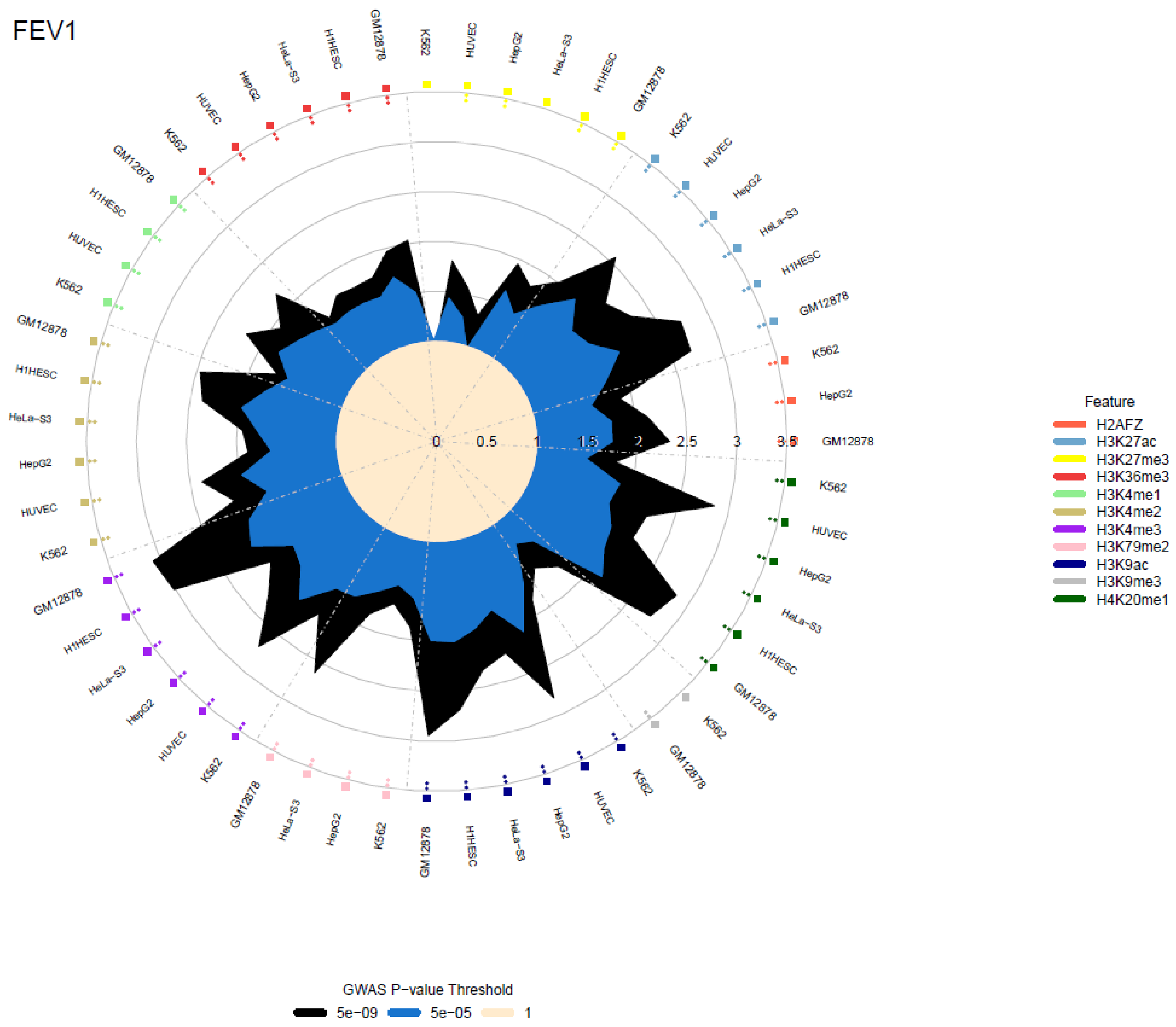

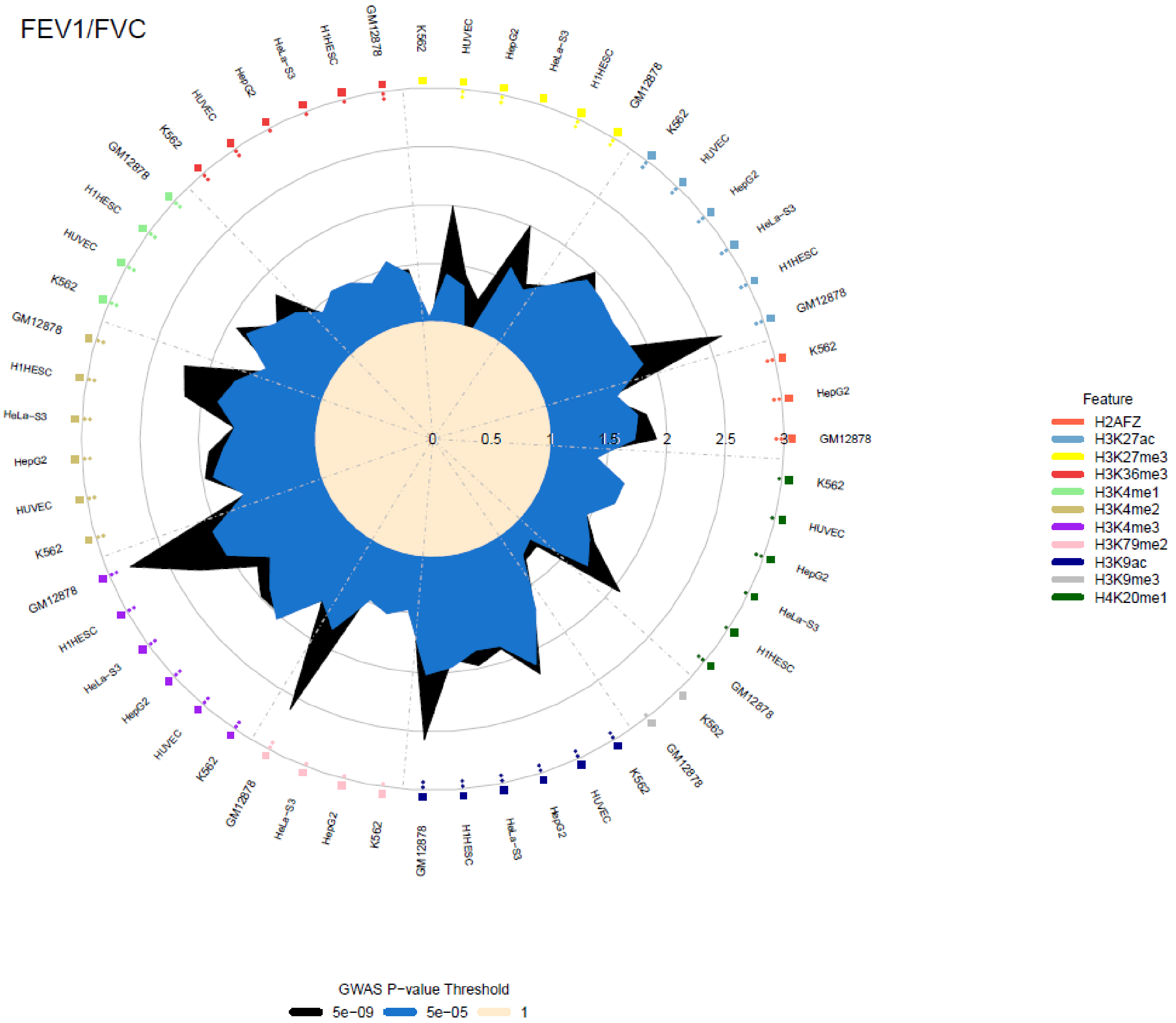
FEV1/FVC
0
0.5
1
1.5
2
2.5
3
Feature
H2AFZ
H3K27ac
H3K27me3
H3K36me3
H3K4me1
H3K4me2
H3K4me3
H3K79me2
H3K9ac
H3K9me3
H4K20me1
GWAS P−value Threshold
5e−09
5e−05
1

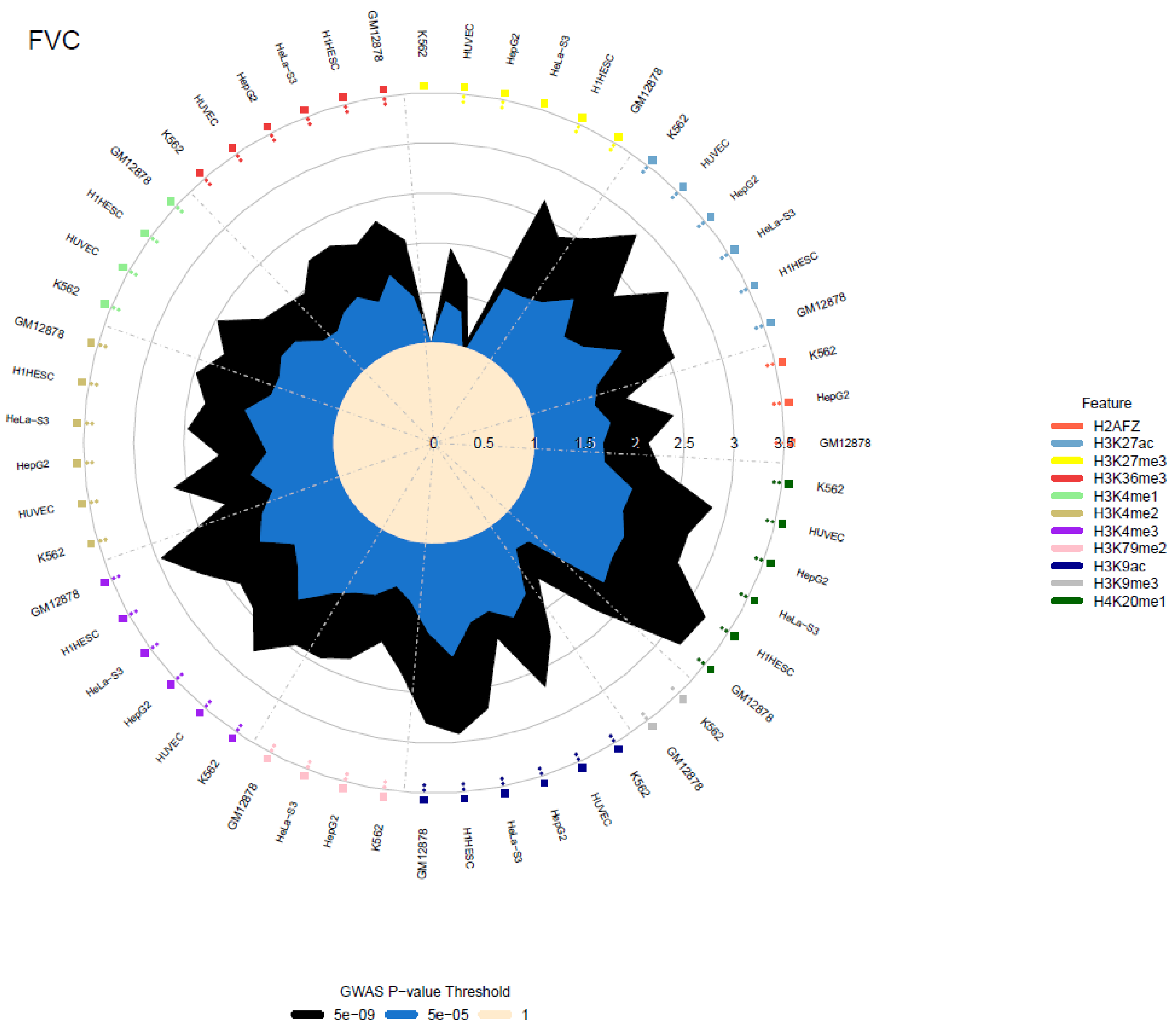
FVC
Feature
H2AFZ
H3K27ac
H3K27me3
H3K36me3
H3K4me1
H3K4me2
H3K4me3
H3K79me2
H3K9ac
H3K9me3
H4K20me1
GM12878
K562
HUVEC
HepG2
HeLa-S3
H1HESC
0
0.5
1
1.5
2
2.5
3
3.5
GWAS P-value Threshold
5e-09
5e-05
1

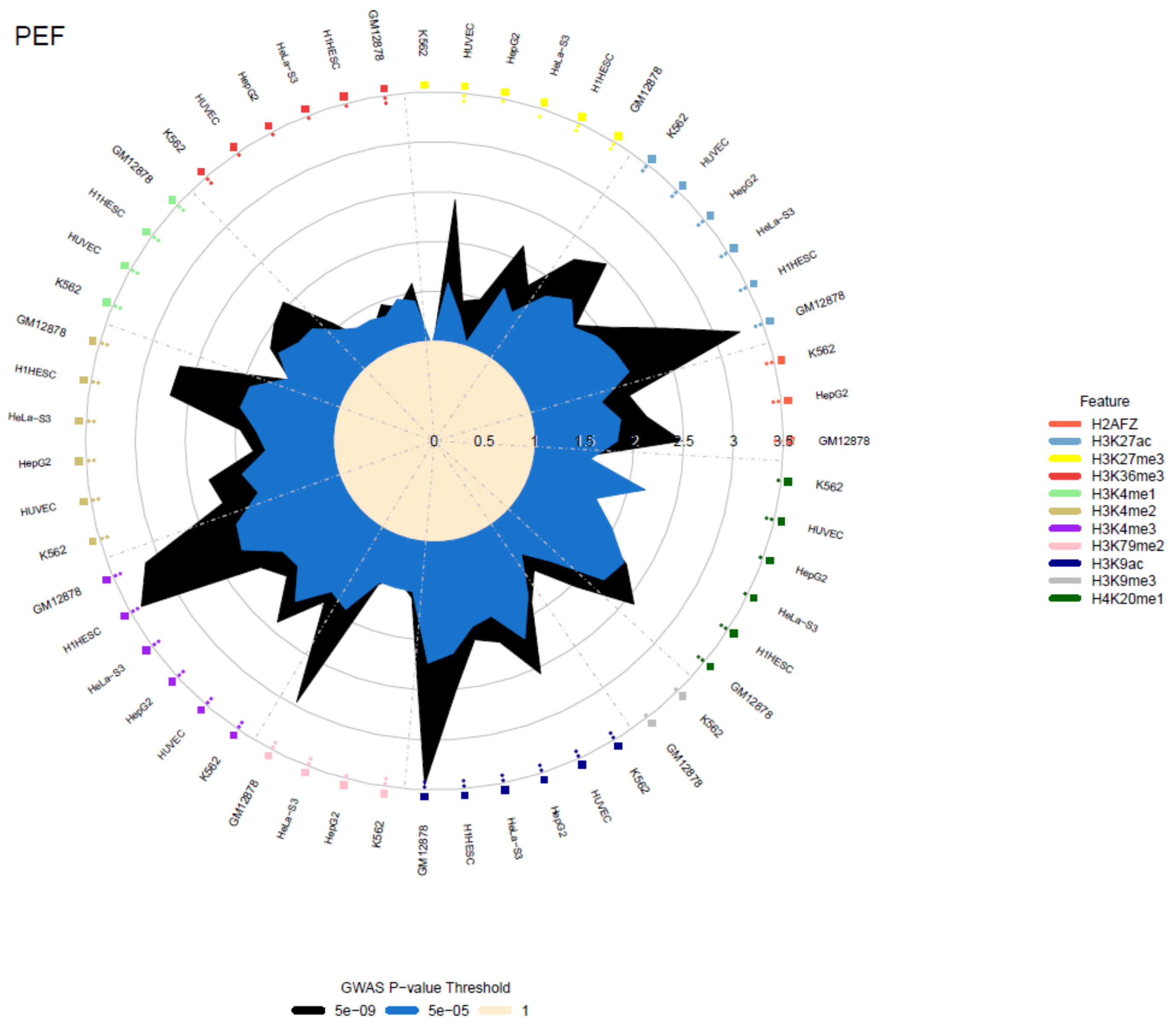
PEF
Feature
H2AFZ
H3K27ac
H3K27me3
H3K36me3
H3K4me1
H3K4me2
H3K4me3
H3K79me2
H3K9ac
H3K9me3
H4K20me1
GWAS P−value Threshold
5e−09
5e−05
1
0
0.5
1
1.5
2
2.5
3
3.5
GM12878
K562
HUVEC
HepG2
HeLa−S3
H1HESC

**Supplementary Figure 9: Comparing effect sizes in children of the top lung function signals with adults in the meta-analysis.**

The effect size estimated from European (EUR) children meta-analysis (ALSPAC, Raine Study and USC children's study) is plotted against that estimated from EUR adults meta-analysis (N = 35 cohorts for $FEV_1$ and FVC; 34 for $FEV_1$/FVC; 16 for PEF) for the 972 genetic signals. The four signals showed significant age-dependent effects (Welch's t test, $P < 5.14 \times 10^{-5}$) were highlighted in red: $FEV_1$ – *MAPT* rs11079718, FVC – *CCDC91* rs7977418 (top), *MECOM* rs6806825 (bottom), PEF – *CYTL1* rs11722554 (Supplementary Table 9).

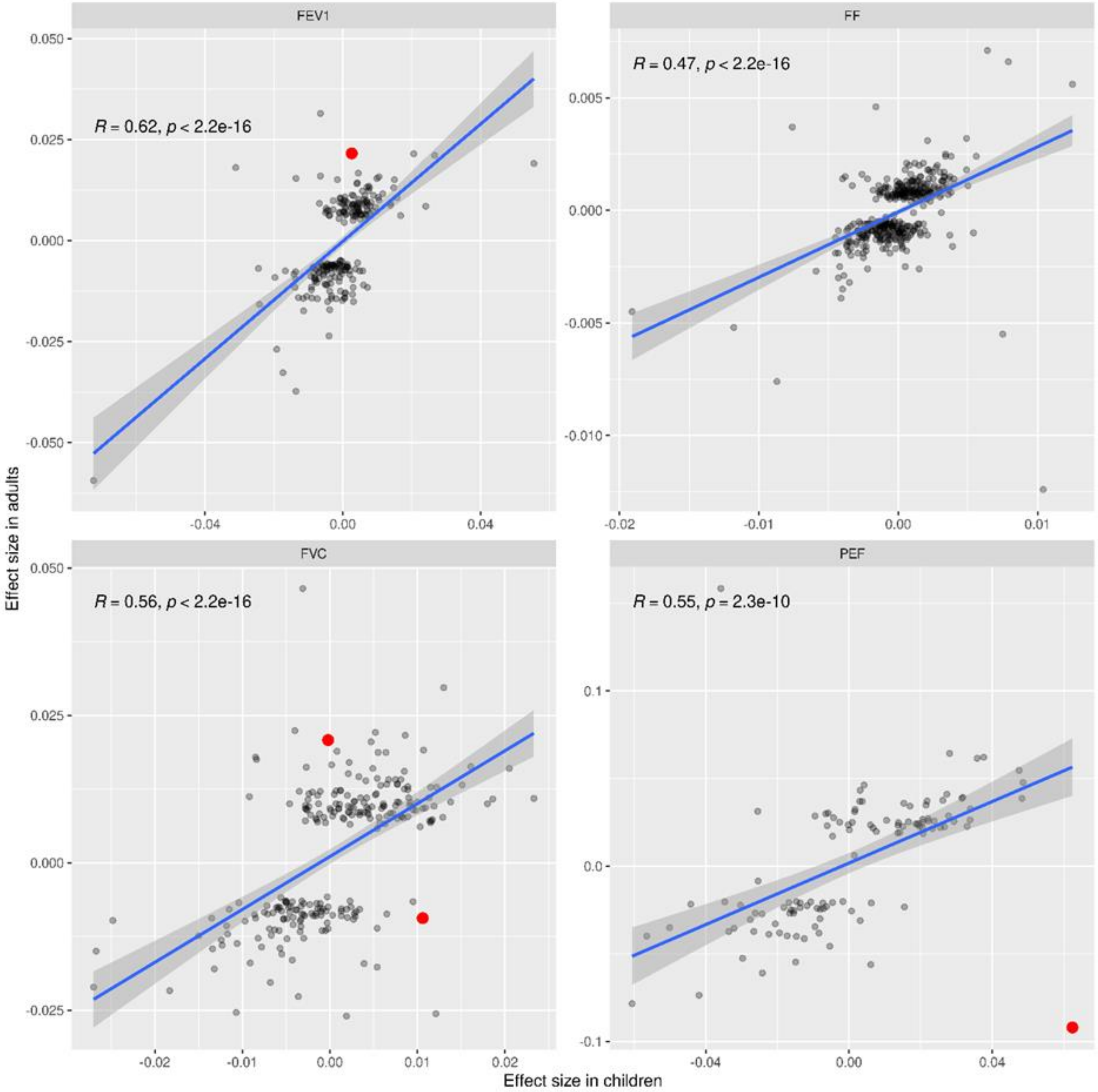

**Supplementary Figure 10: Correlation of height effects vs lung function effects**

Height effects from the GIANT consortium meta-analysis[78] are plotted against lung function effects from a fixed-effects meta-analysis of untransformed traits in this study. We included 899 of lung function 1,020 signals, 403 were exactly the same sentinel in the GIANT height look up and 496 are proxies with an $r^2$ >0.4; 318, 269, 229 and 202 SNPs for $FEV_1$, FVC, $FEV_1$/FVC and PEF, respectively are plotted that have both P <$10^5$ for lung function and P < 0.01 for height. No significant correlation of height effect with lung function effect is observed.

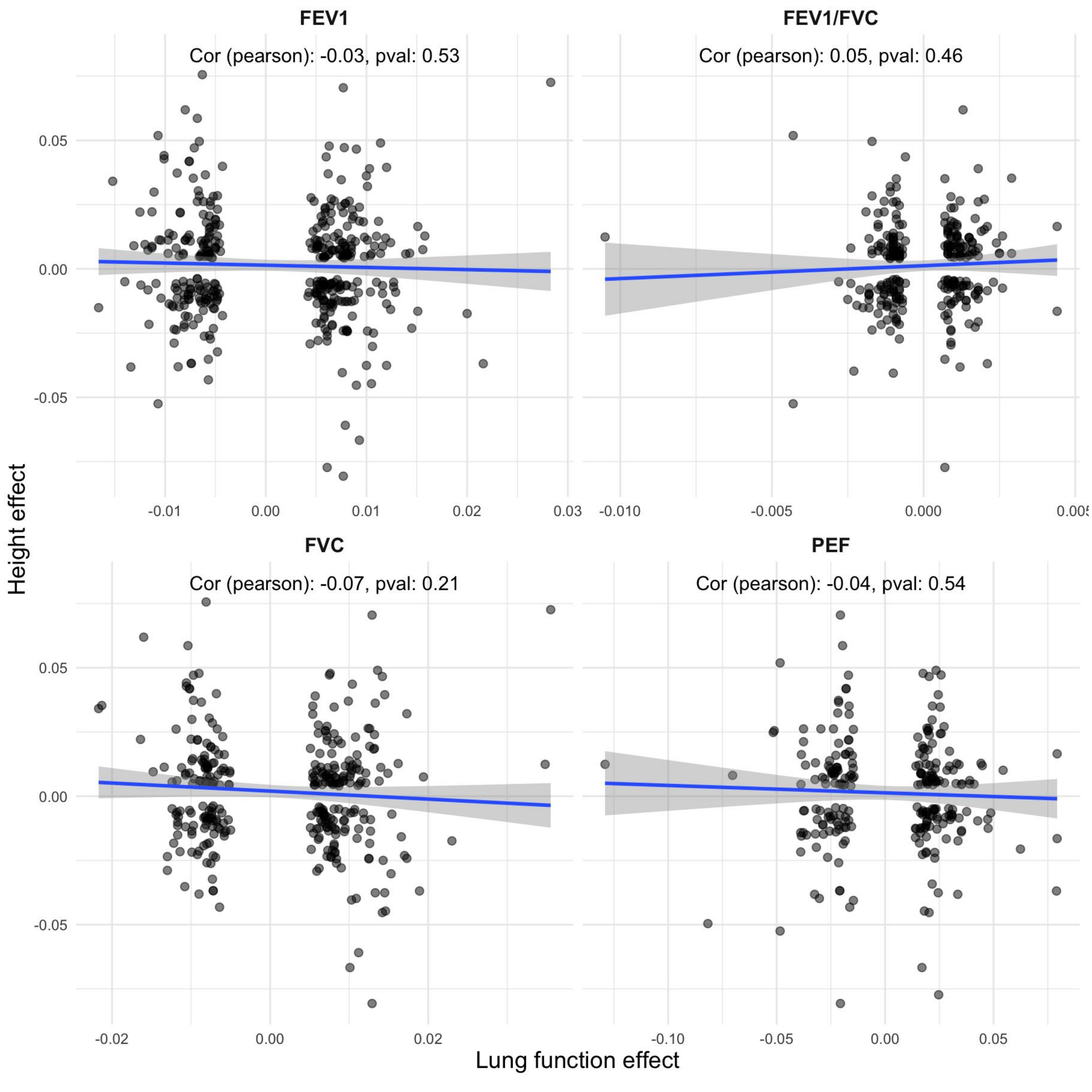

**Supplementary Figure 11: Sensitivity analysis: adjustment for height$^2$ and height$^3$.**
Comparison of effect sizes in UK Biobank European ancestry (N=320,656) when adding **a)** height$^2$ as an additional covariate (age, age$^2$, sex, height, smoking status already included as covariates) and **b)** then additionally height$^3$

**a)**

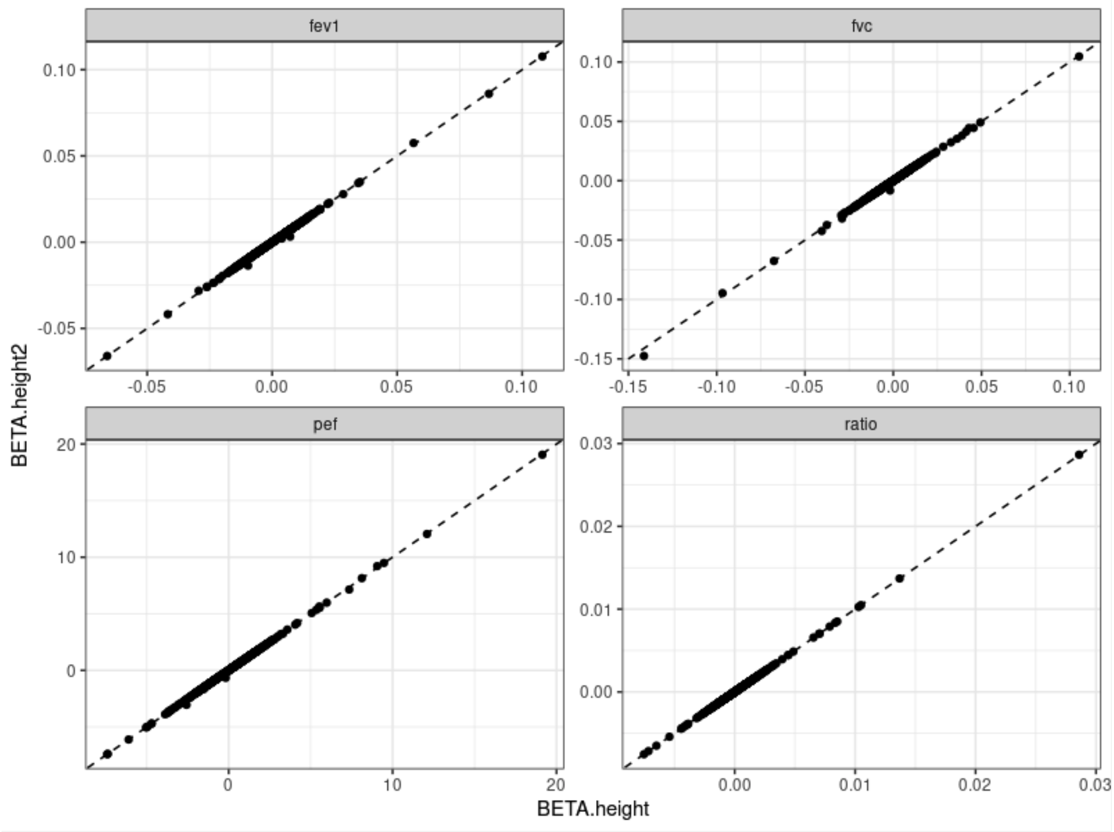

**b)**

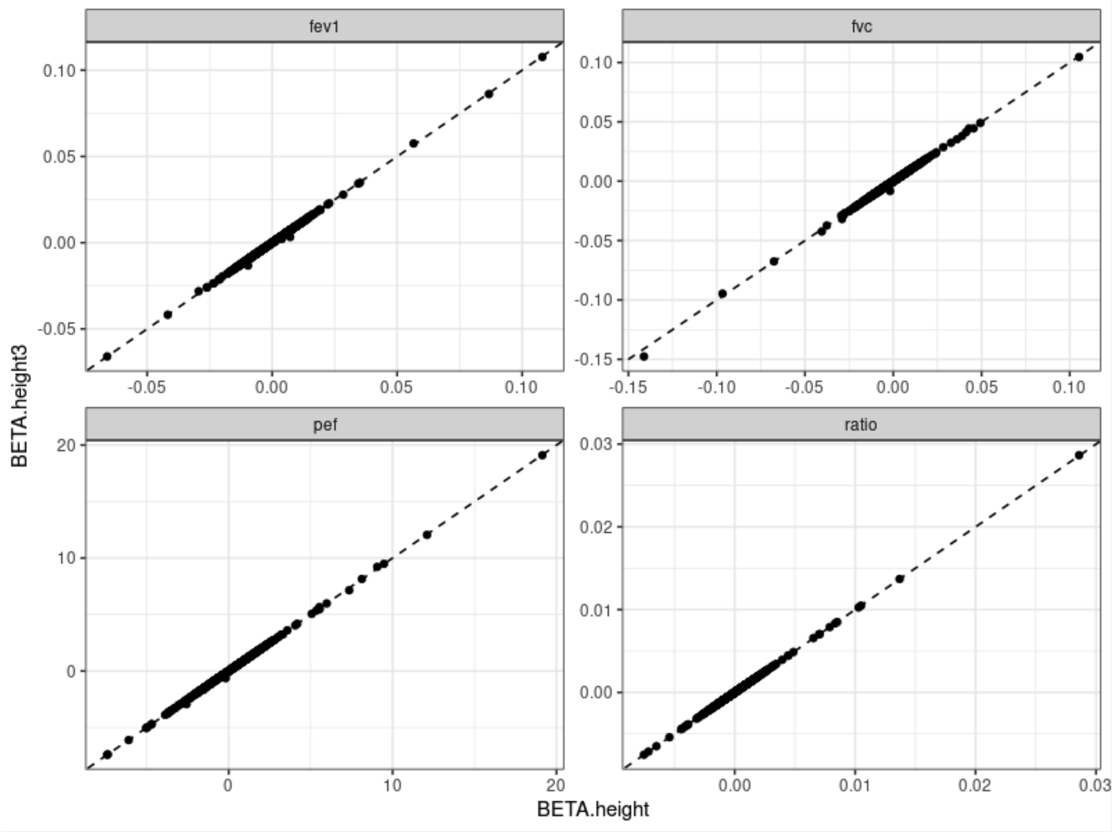

**Supplementary Figure 12: SNP by smoking interaction in 1020 signals.**

Effects of EUR ever smokers meta-analysis against EUR never smokers meta-analysis for lung function traits $FEV_1$, FVC, $FEV_1$/FVC and PEF (N=206,162 ever smokers and 229,046 never smokers). Signals in red showed significantly smoking interaction effects (z test, $P < 4.0x10^{-5}$).

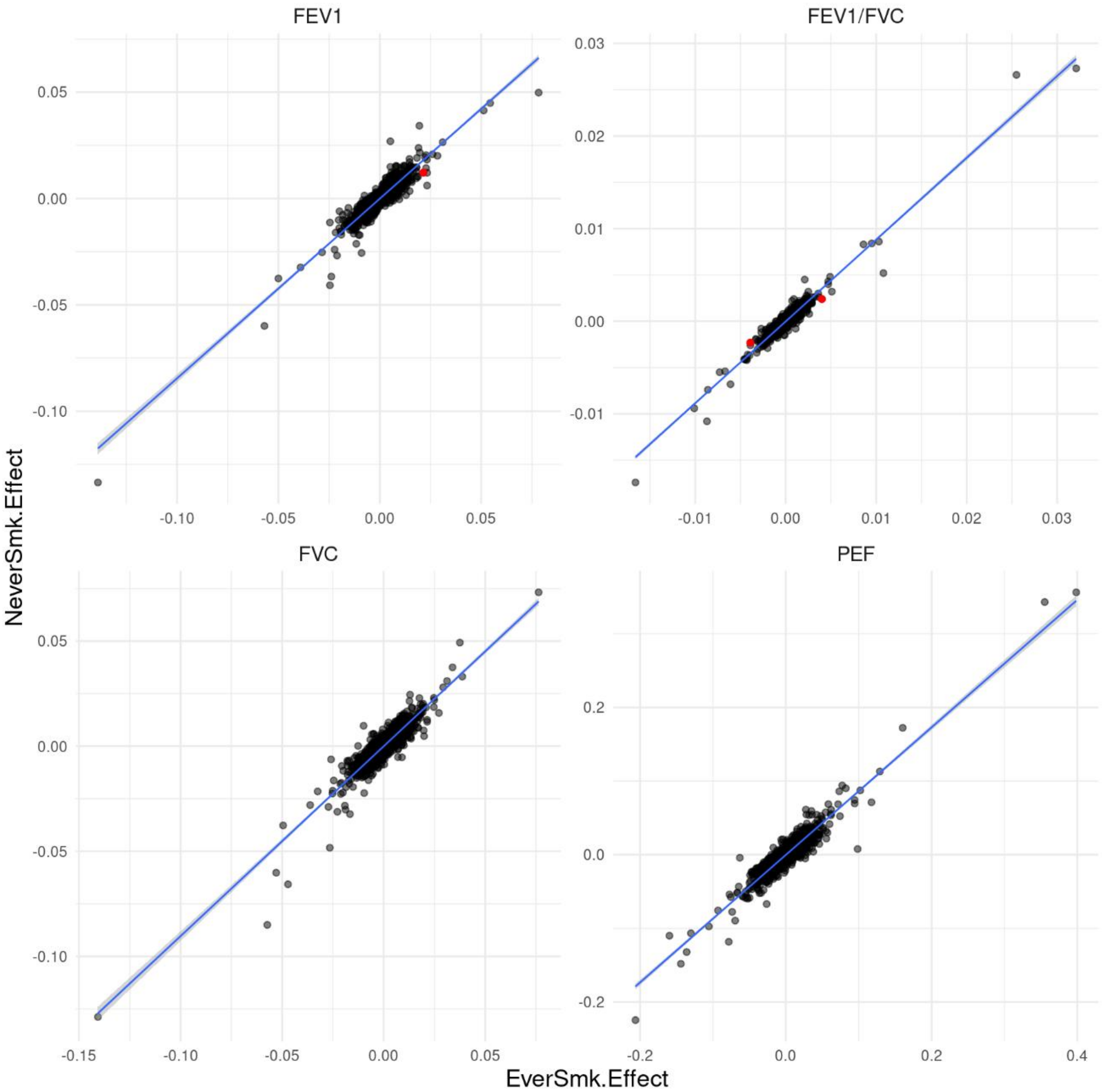

**Supplementary Figure 13: Pathway correlation.**

**a)** Heatmap displaying the proportion of the union of genes in each pair of pathways for the 29 pathways enriched at an FDR $<10^{-5}$ for 559 implicated genes by 2 or more lines of evidence.

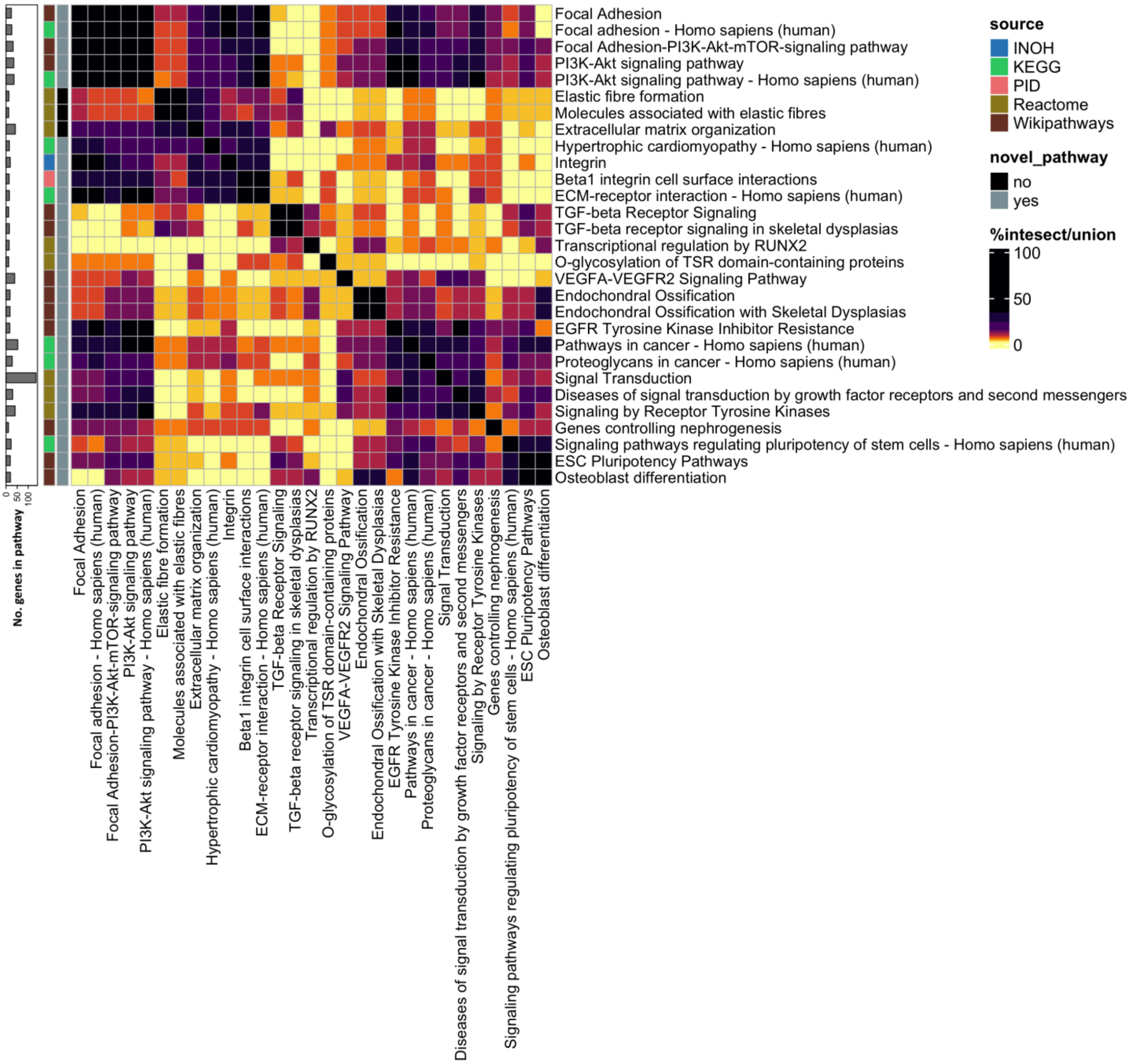

**b)** Heatmap displaying the Pearson r correlation between individuals' genetic risk scores in each pair of pathways in the 29 pathways enriched at an FDR <$10^{-5}$ for 559 implicated genes by 2 or more lines of evidence.

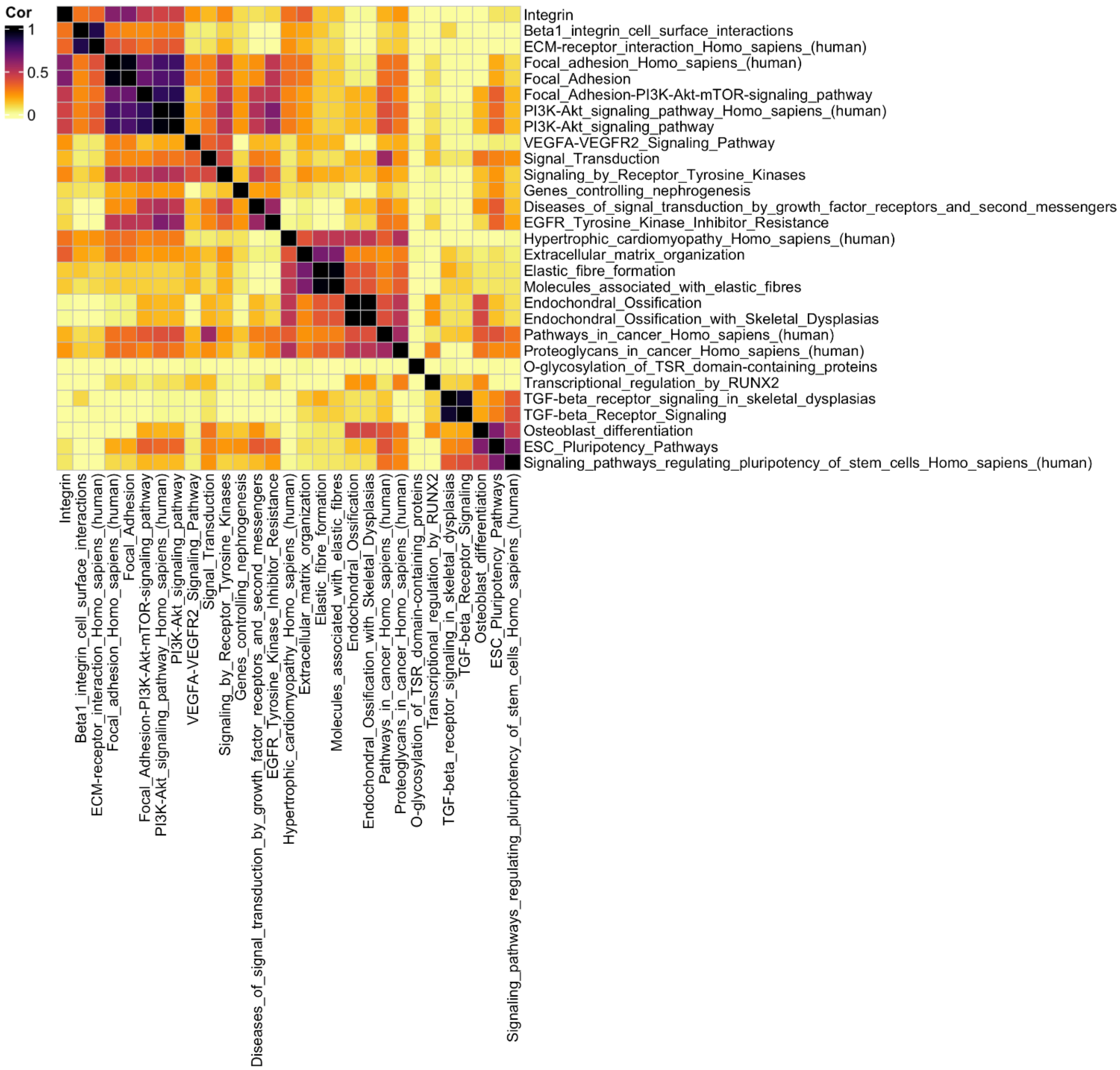

**c)** Pairwise scatter plots of individual pathway genetic risk scores for pathways Elastic fibre formation (Reactome), PI3K-Akt signalling pathway Homo sapiens (human) (KEGG), Hypertrophic cardiomyopathy Homo sapiens (human) (KEGG) and Signal Transduction (Reactome).
The x-axis corresponds to the scores for the pathways with the horizontal labels and vice-versa.

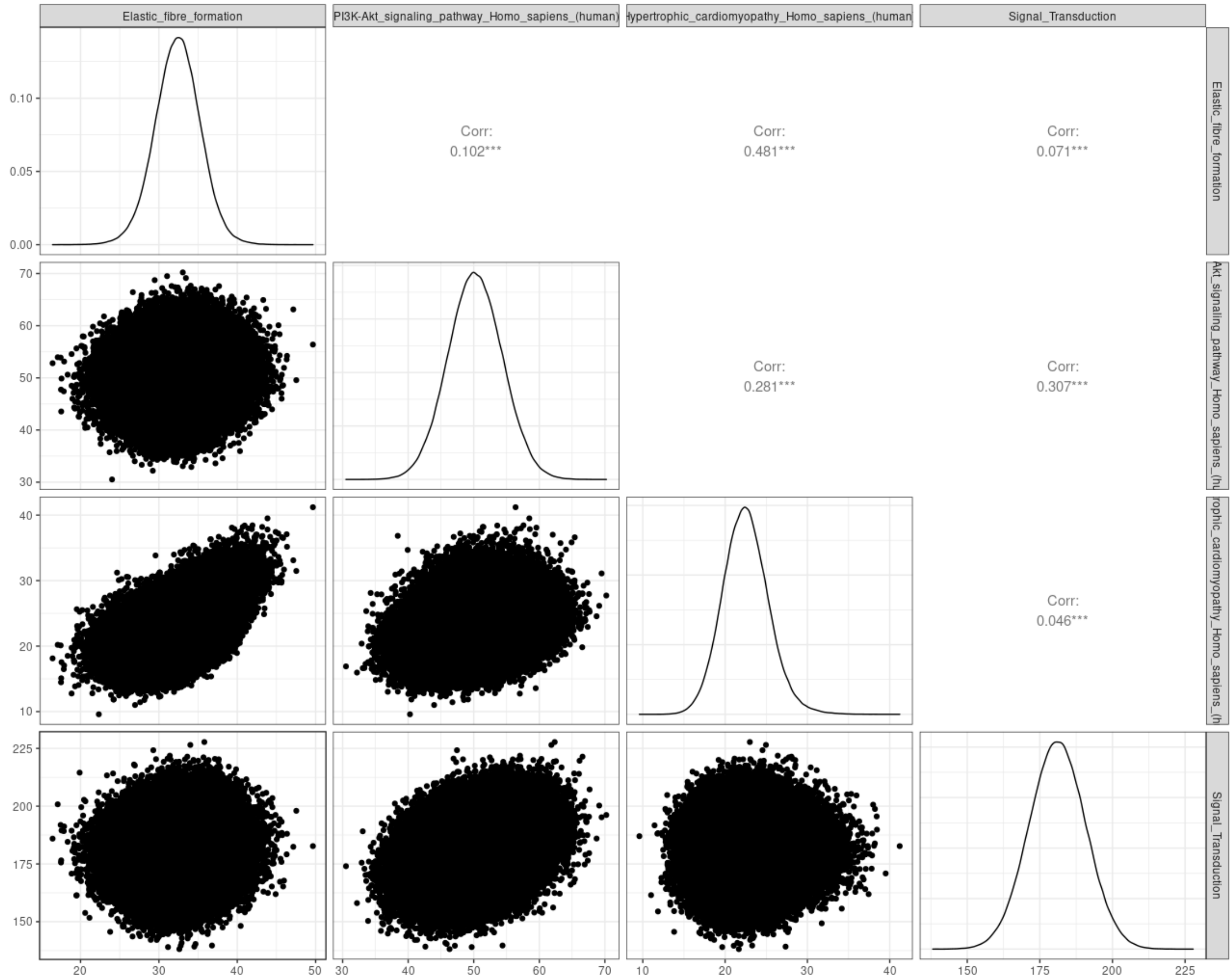

**Supplementary Figure 14: Single variant PheWAS results**

27 single variant PheWAS associations in up to 430,402 unrelated European samples from UK Biobank. PheWAS were aligned to the lung function risk allele i.e. the trait decreasing allele for the lung function trait for which the variant is reported here.

a) ***ACAN*** **rs34949187 – aligned to $FEV_1$/FVC decreasing G allele**

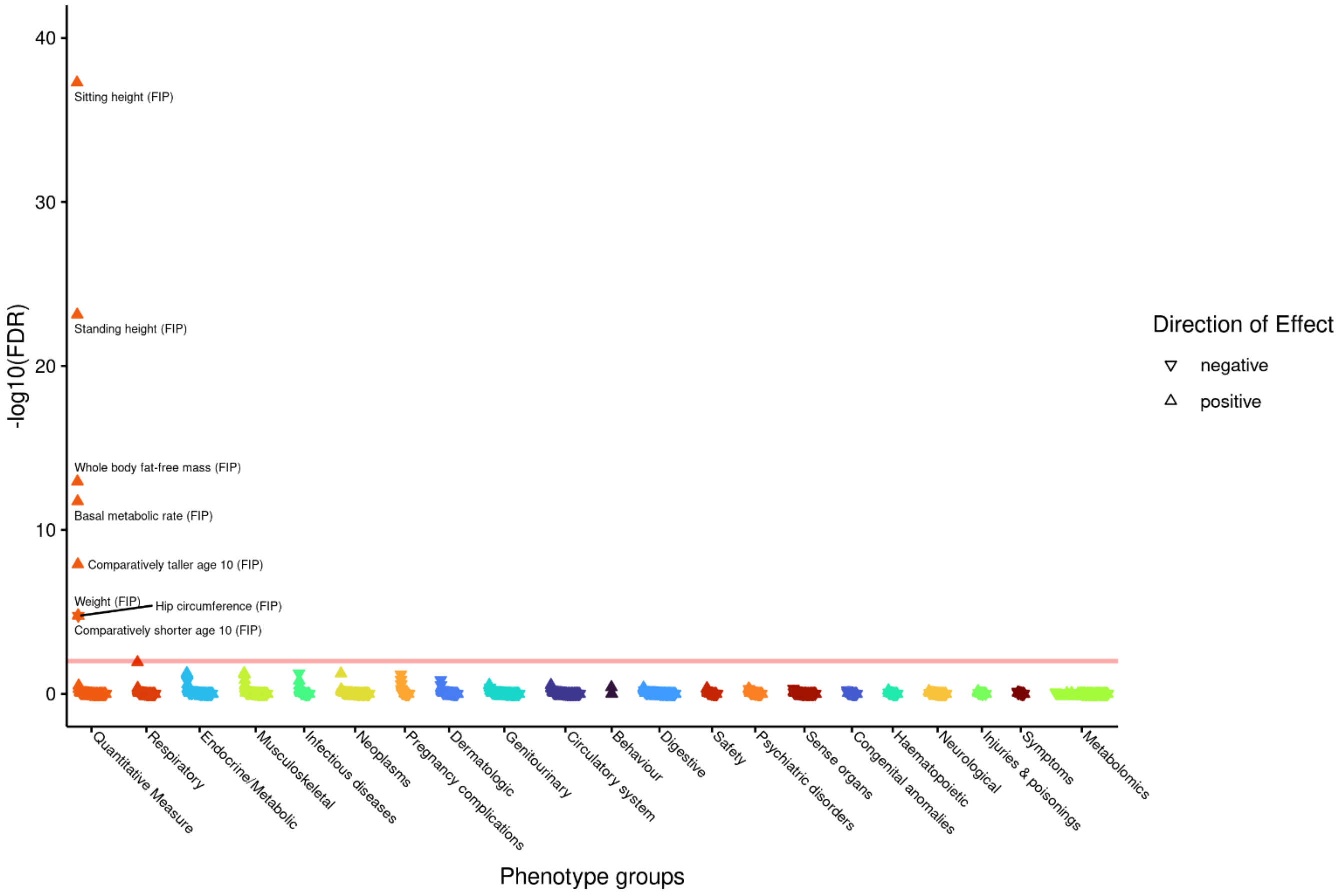

b) *ADAMTS10* rs62621197 – aligned to FVC decreasing T allele

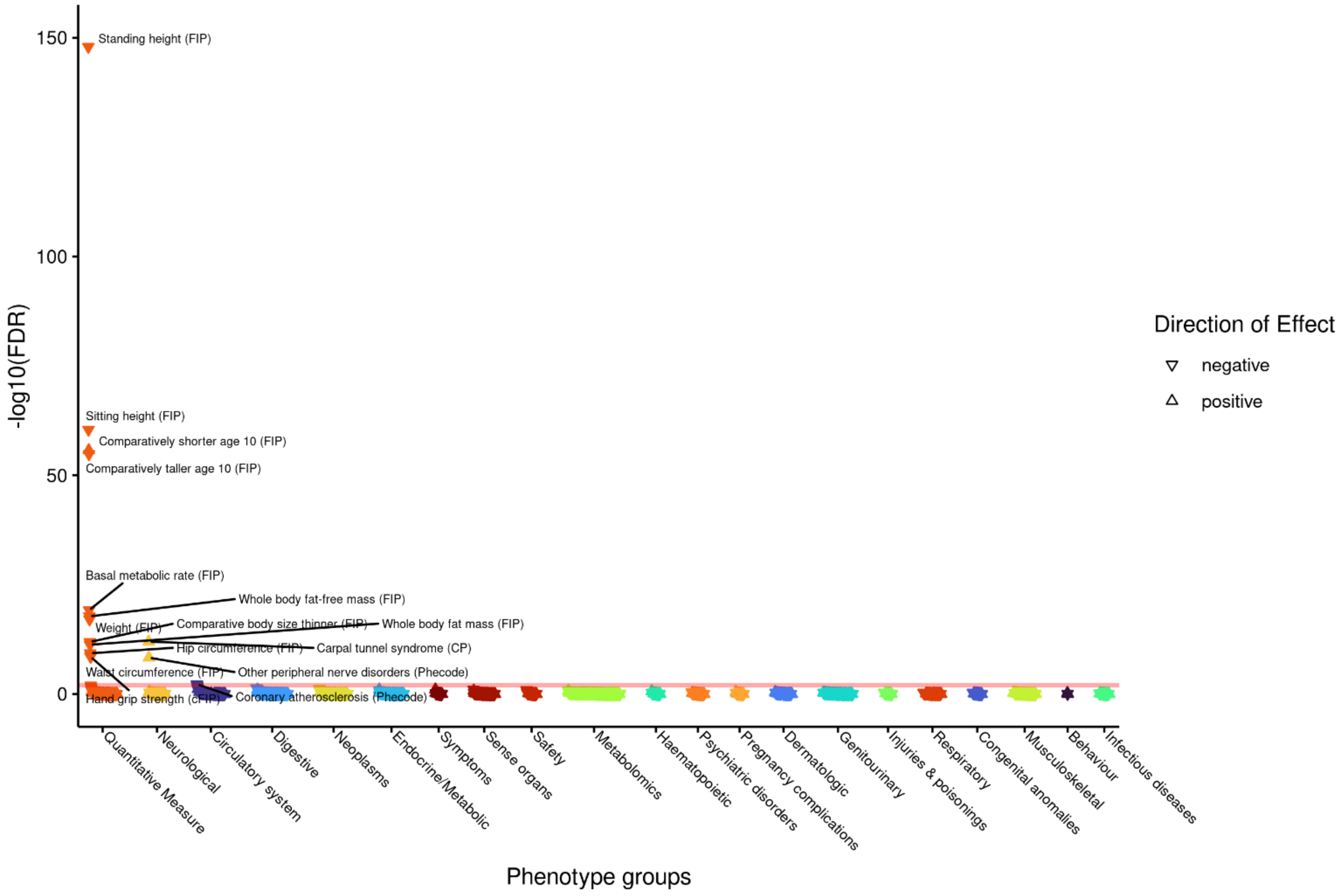

c) *ADGRG6* rs17280293 – aligned to $FEV_1$/FVC decreasing A allele

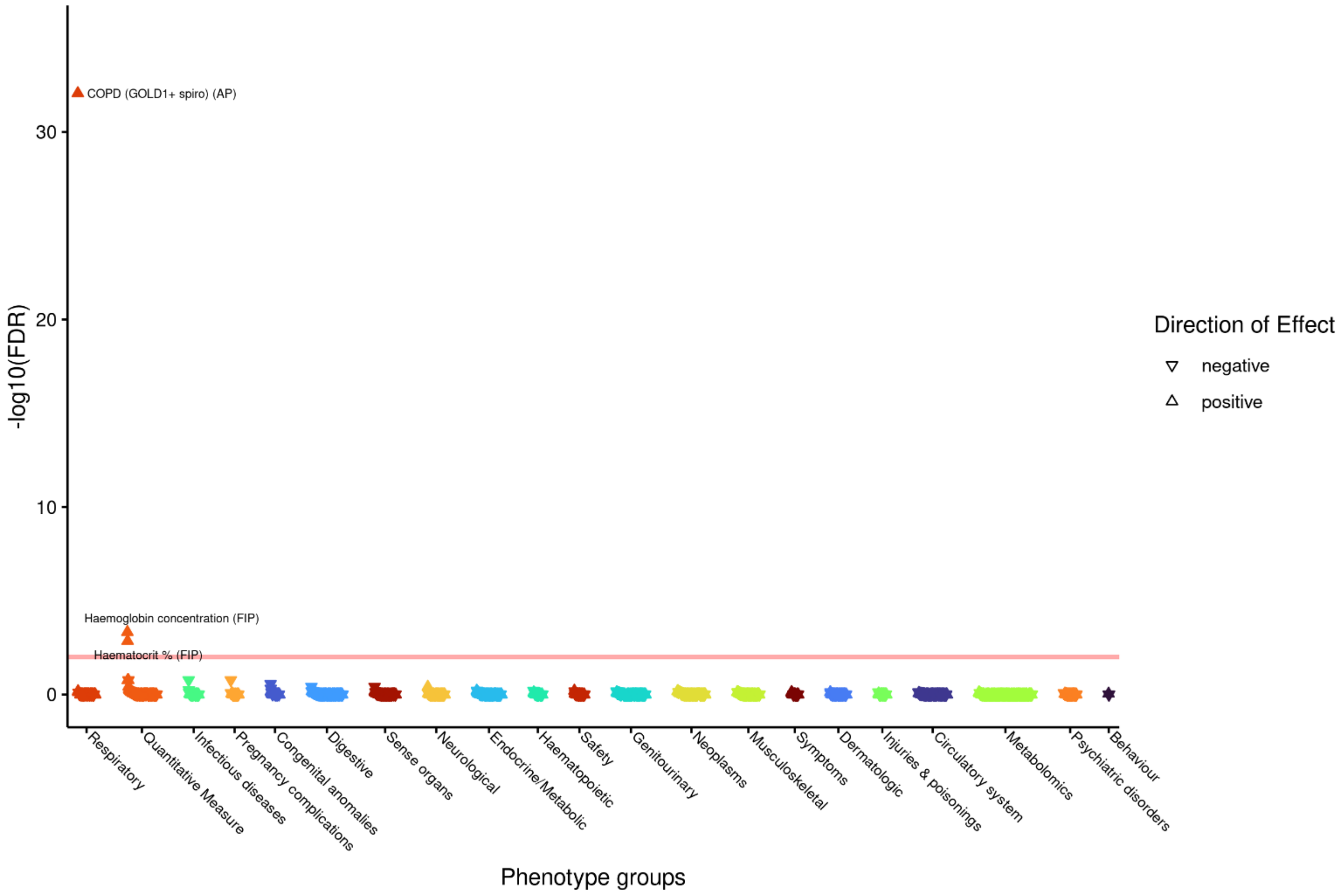

**d)** ***ADRB2*** **rs1800888 – aligned to $FEV_1$/FVC decreasing T allele**

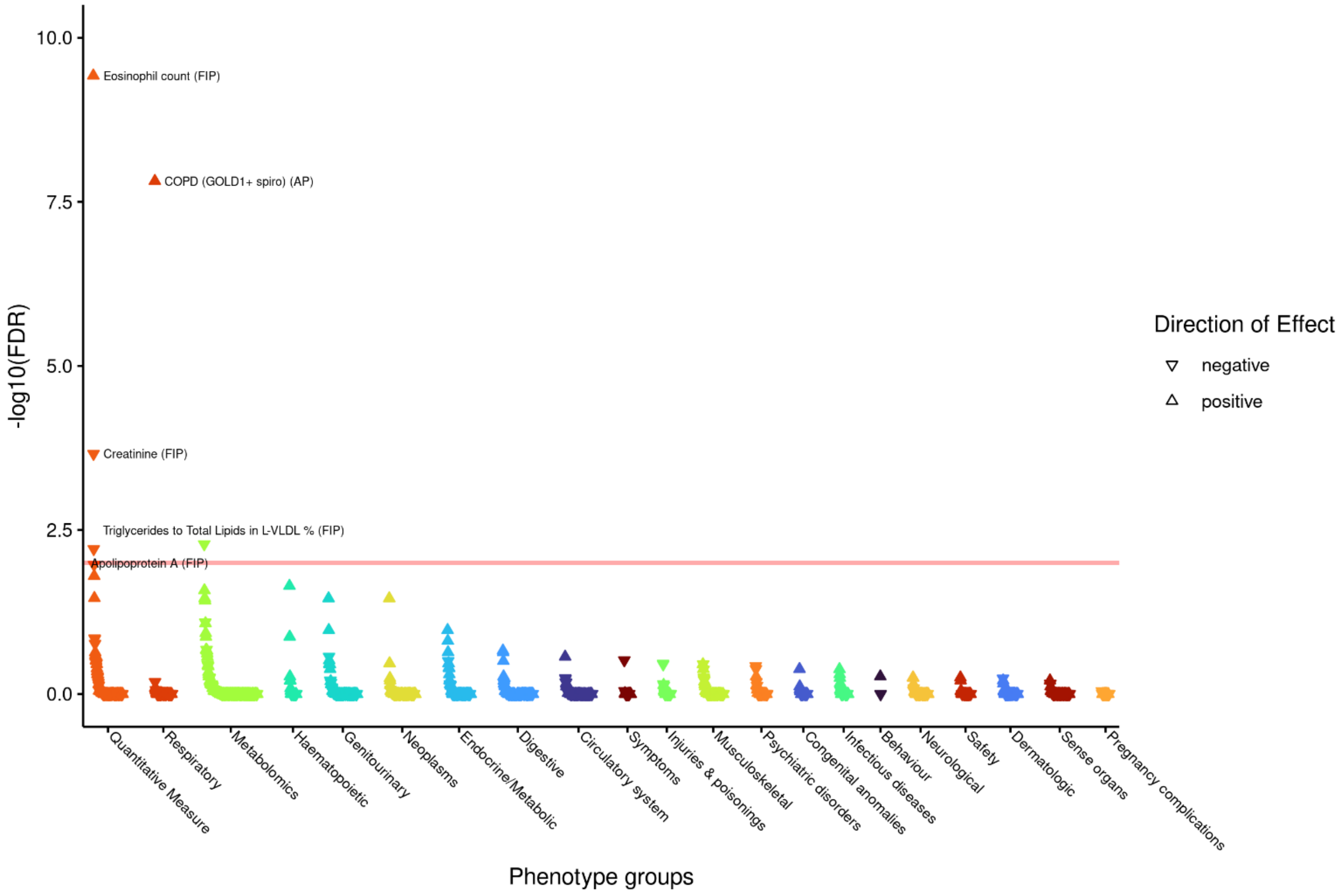

**e)** ***AP3B1*** **rs55769512 – aligned to FVC decreasing C allele**

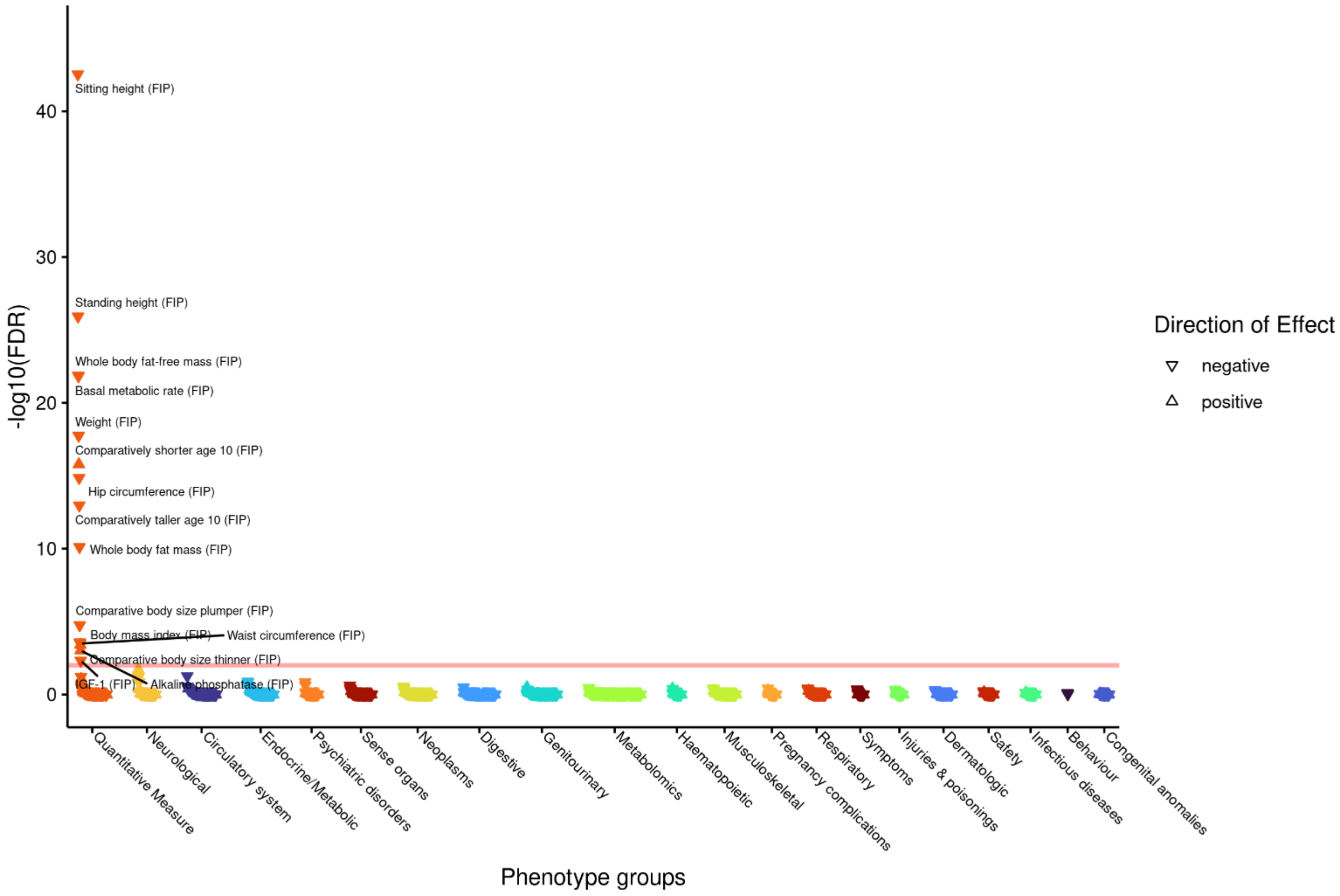

**f)** ***CACNA1S*** **rs3850625 – aligned to FVC decreasing A allele**

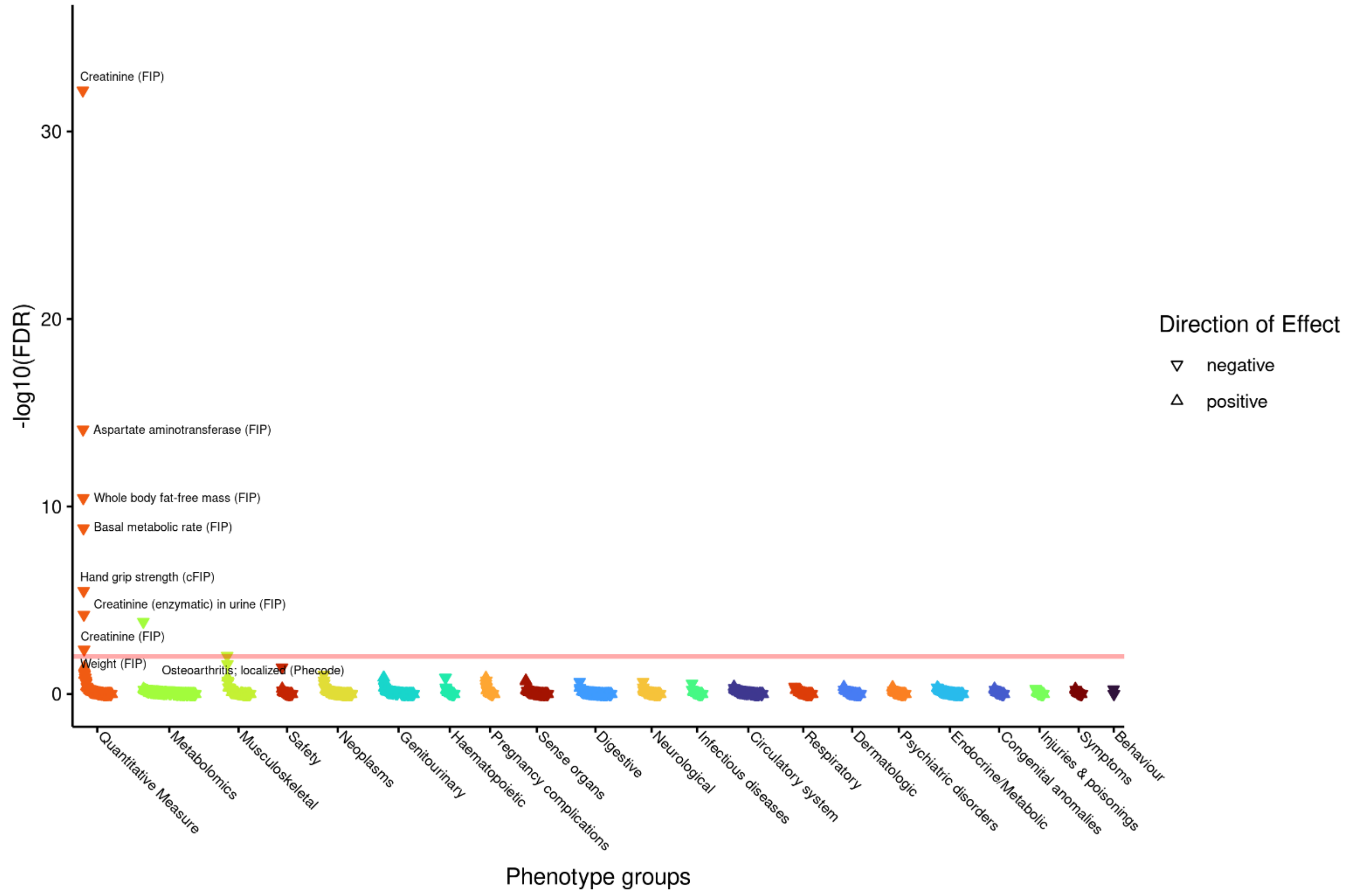

**g)** ***CCDC91*** **rs7977418 – aligned to FVC decreasing C allele**

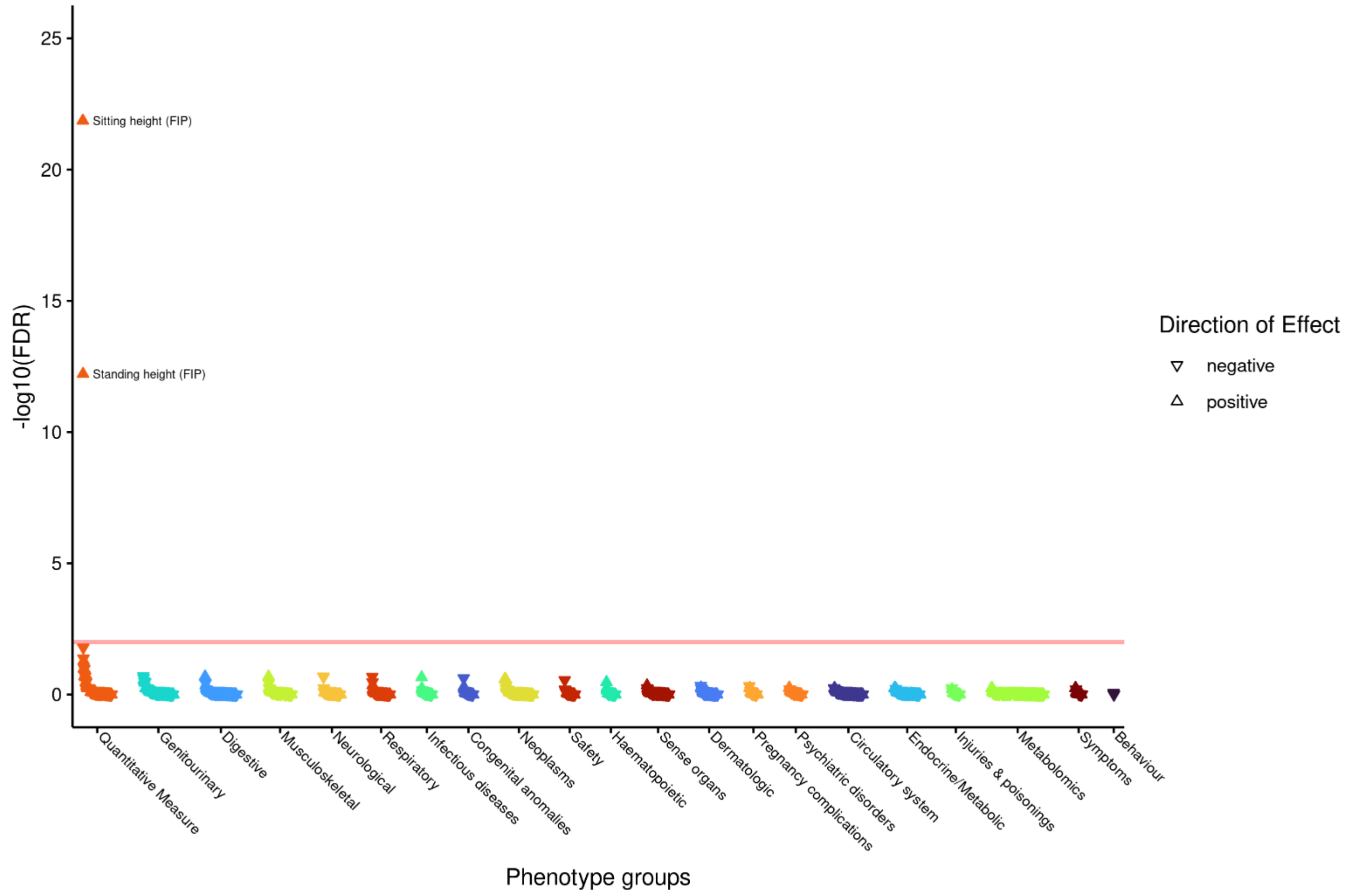

h) *CFH* rs7514261 – aligned to FVC decreasing A allele

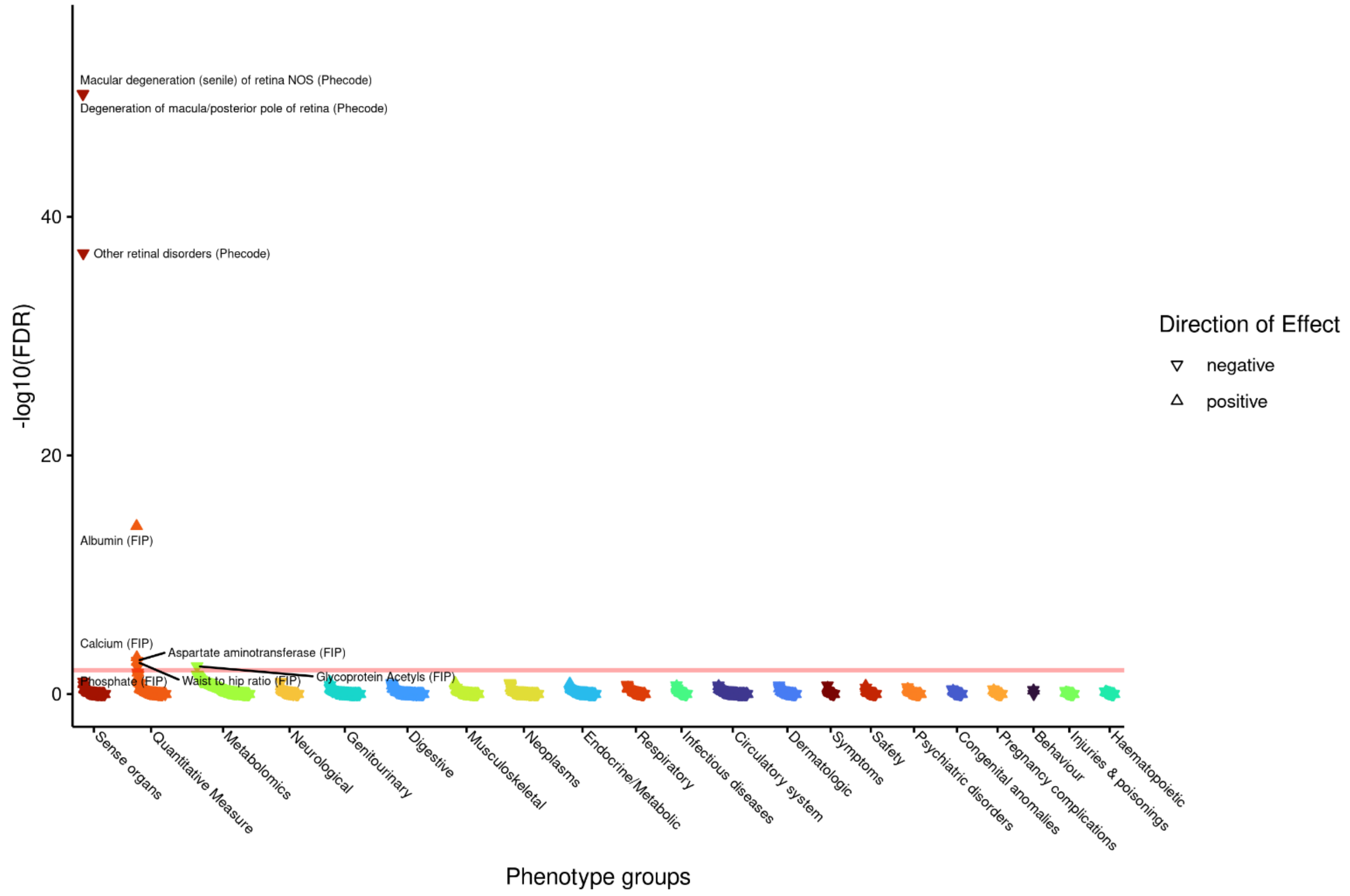

i) *CYTL1* rs11722554 – aligned to PEF decreasing A allele

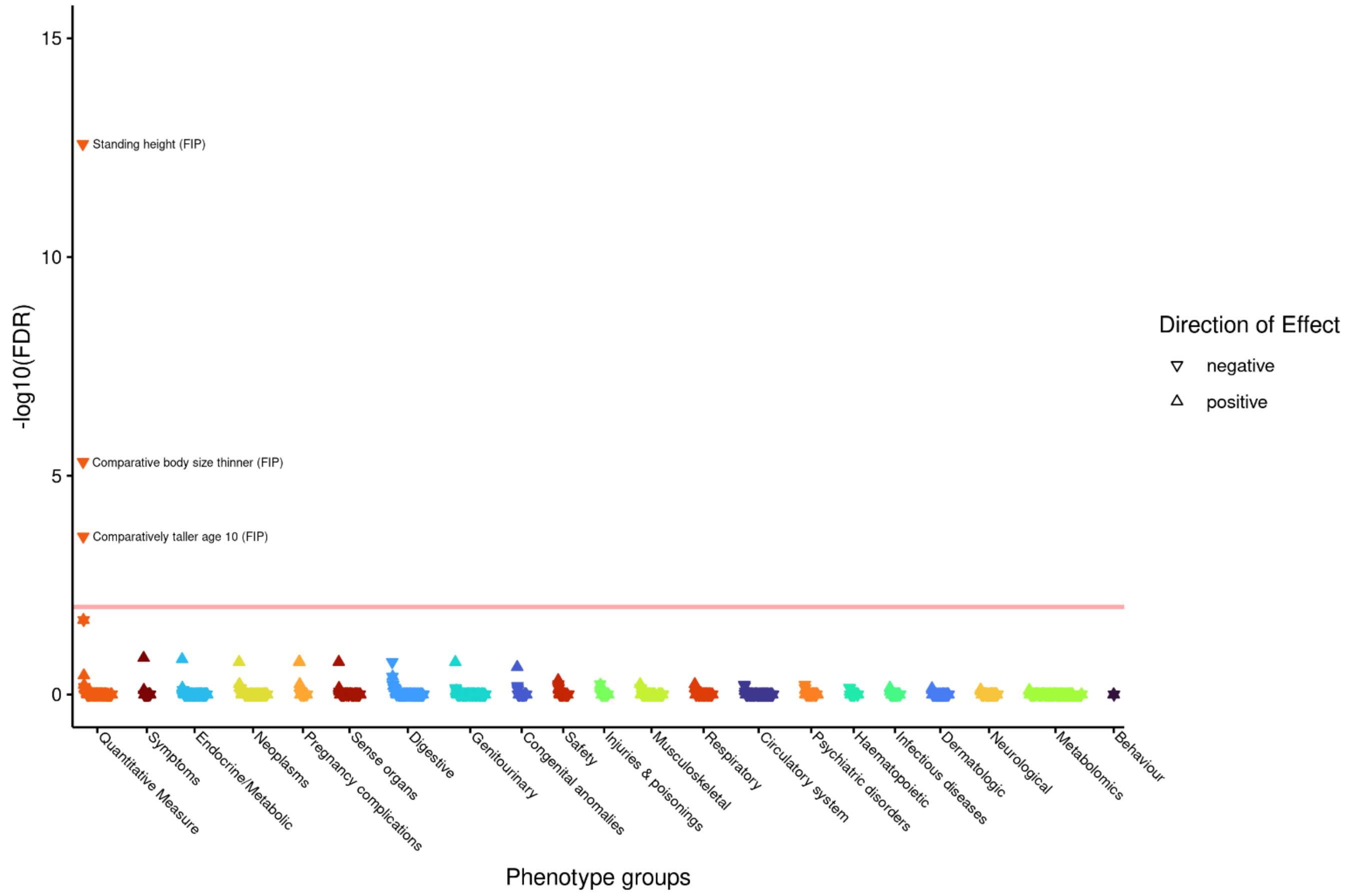

j) *GATA5* rs200383755 – aligned to $FEV_1$ decreasing C allele

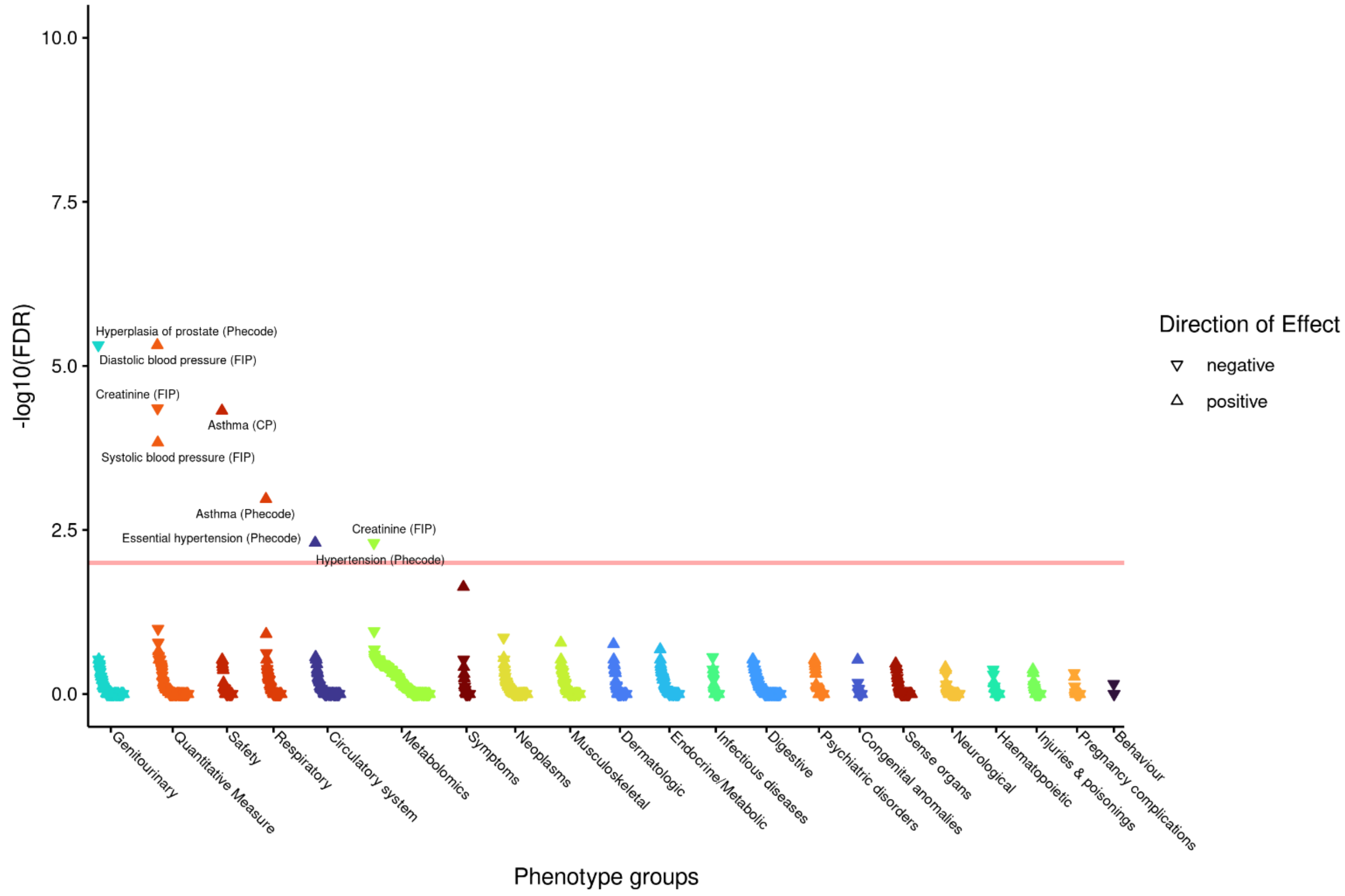

k) *HIST1H2BE* rs9393688 – aligned to FVC decreasing A allele

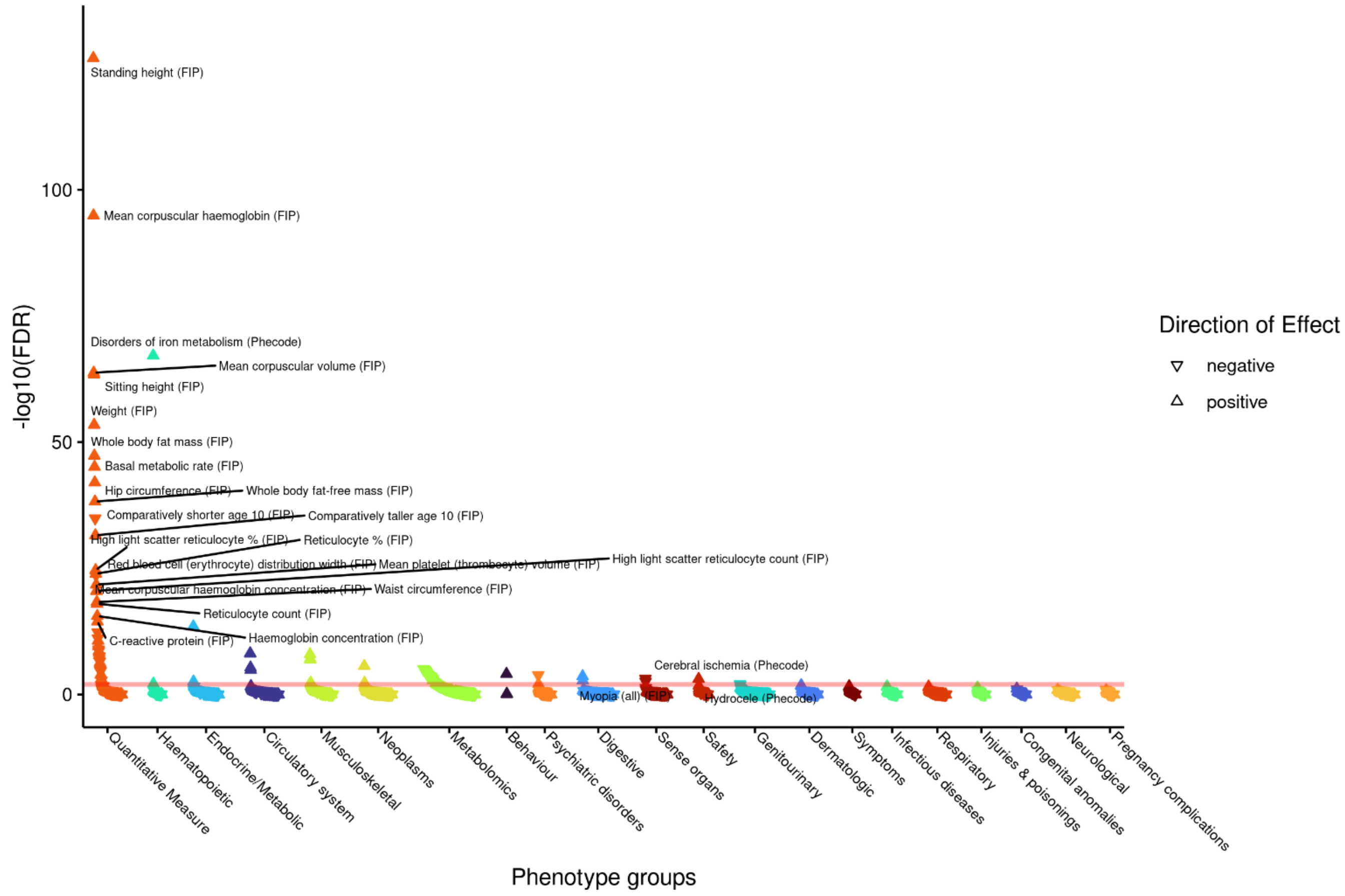

l) *HMCN1* rs17531405 – aligned to $FEV_1$/FVC decreasing G allele

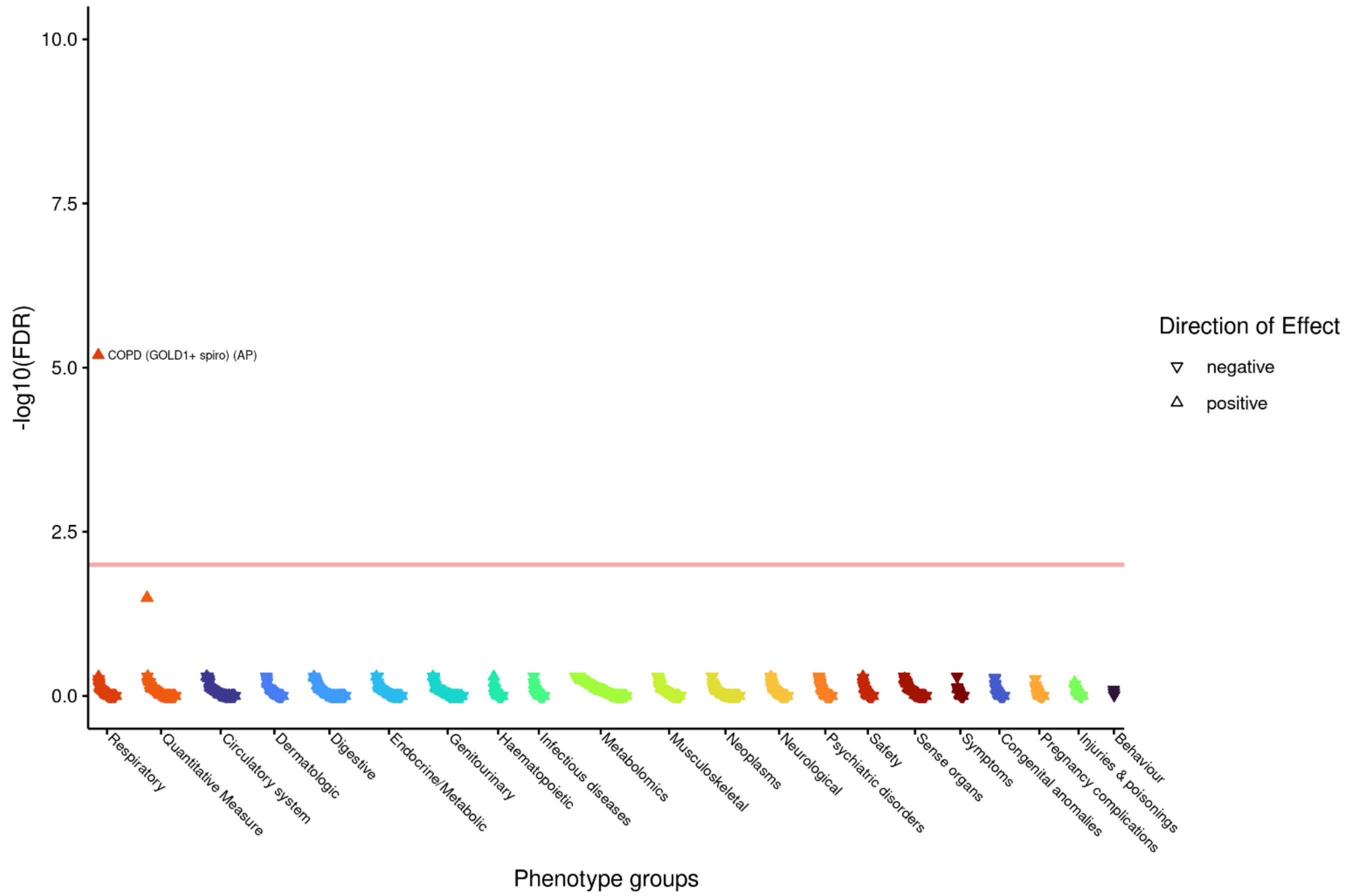

m) *IGF2BP2* rs7615045 – aligned to PEF decreasing G allele

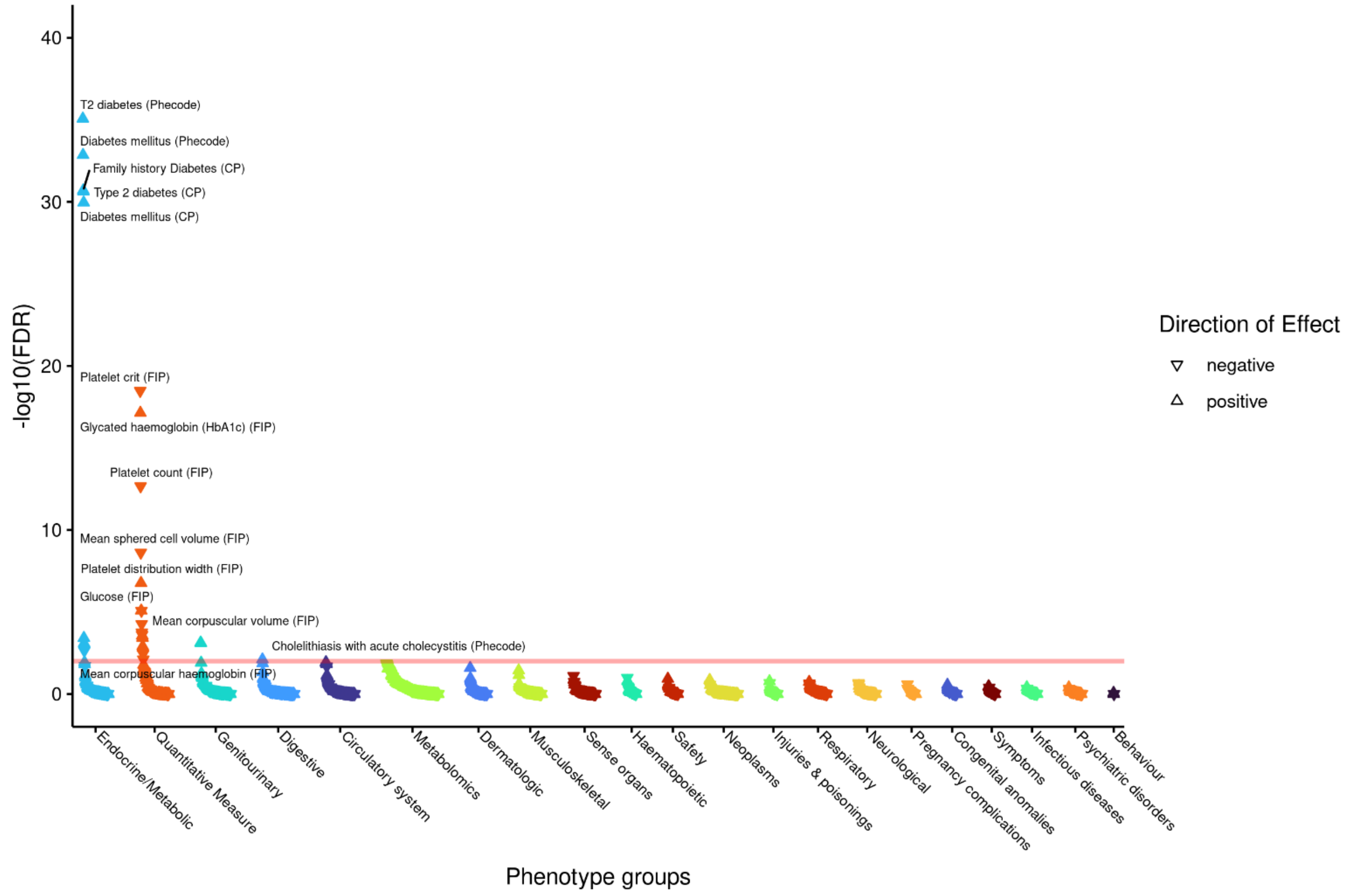

**n)** ***IGHMBP2*** **rs901823 – aligned to PEF decreasing C allele**

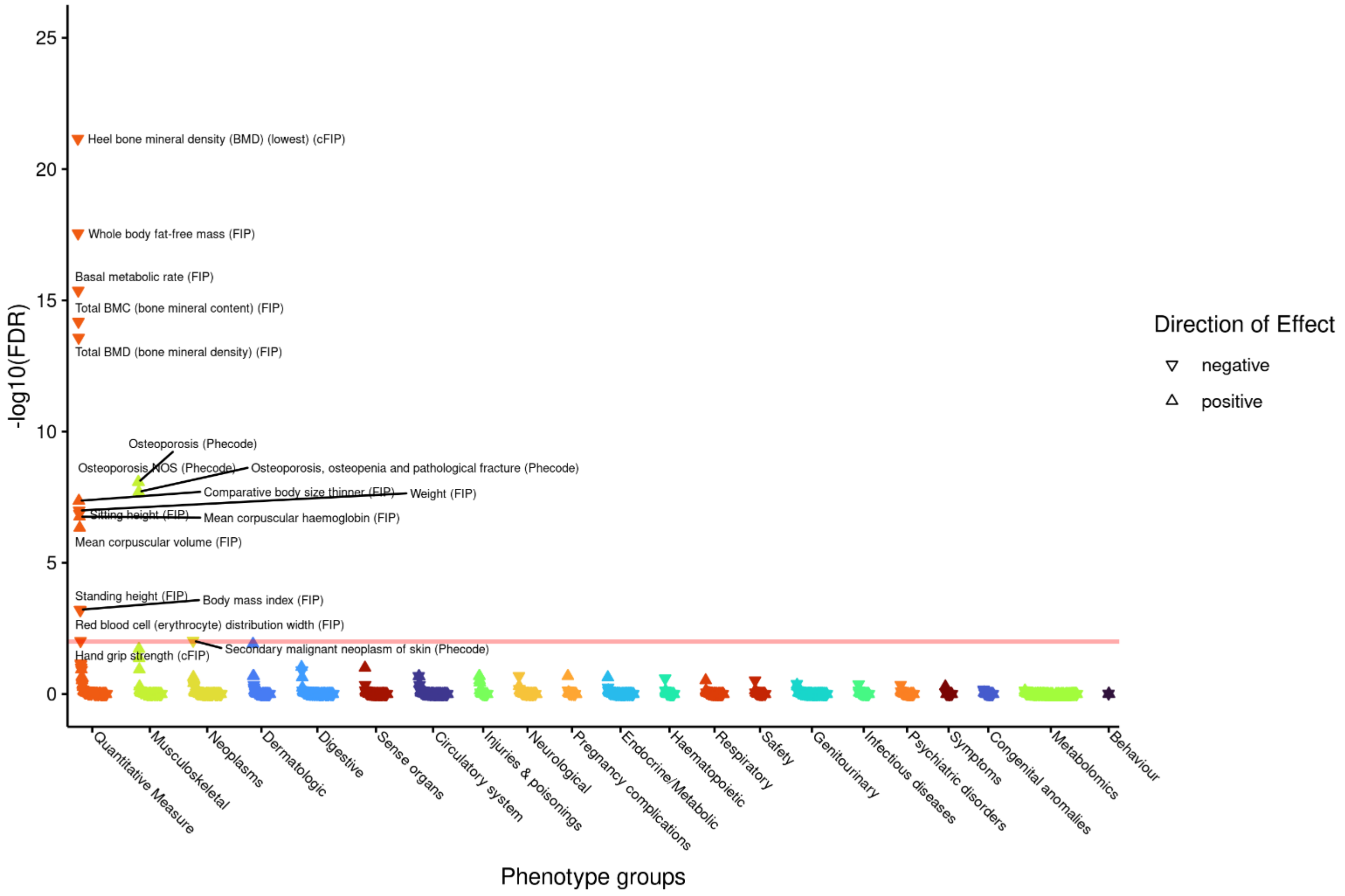

**o)** ***LRBA*** **rs2290846 – aligned to FVC decreasing G allele**

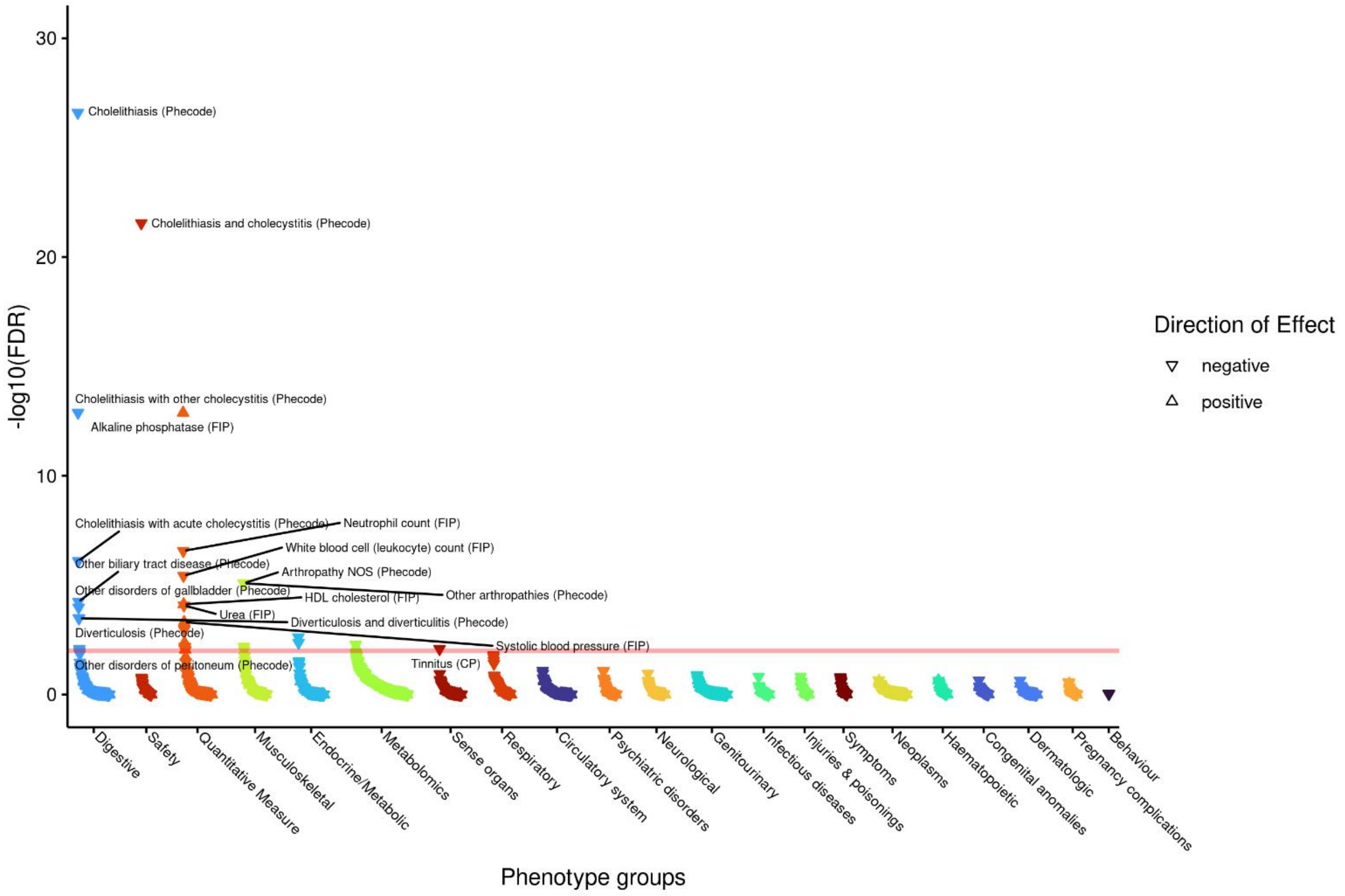

**p)** ***LTBP4*** **rs34093919 – aligned to $FEV_1$/FVC decreasing G allele**

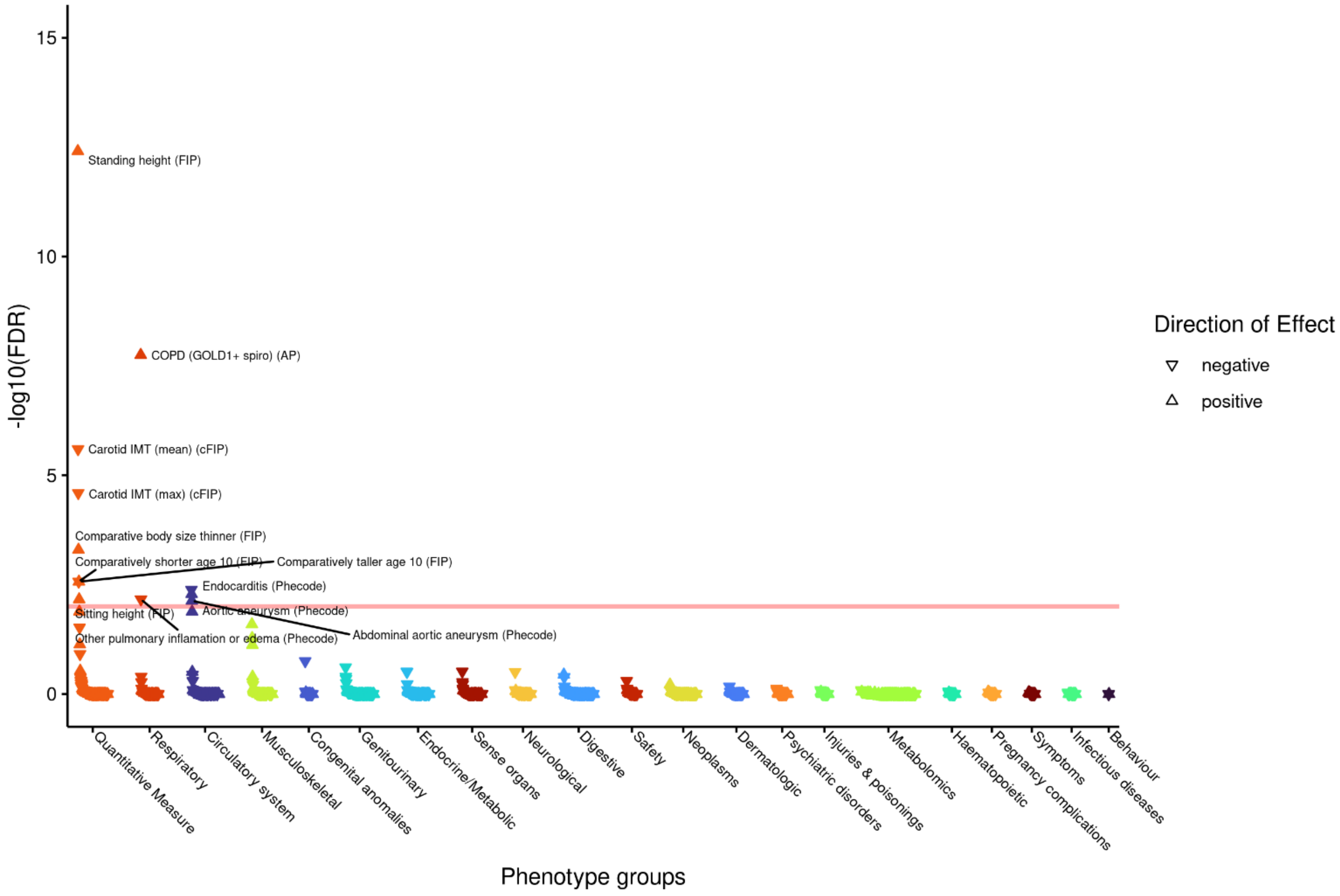

**q)** ***MAPT*** **rs11079718 – aligned to $FEV_1$ decreasing T allele**

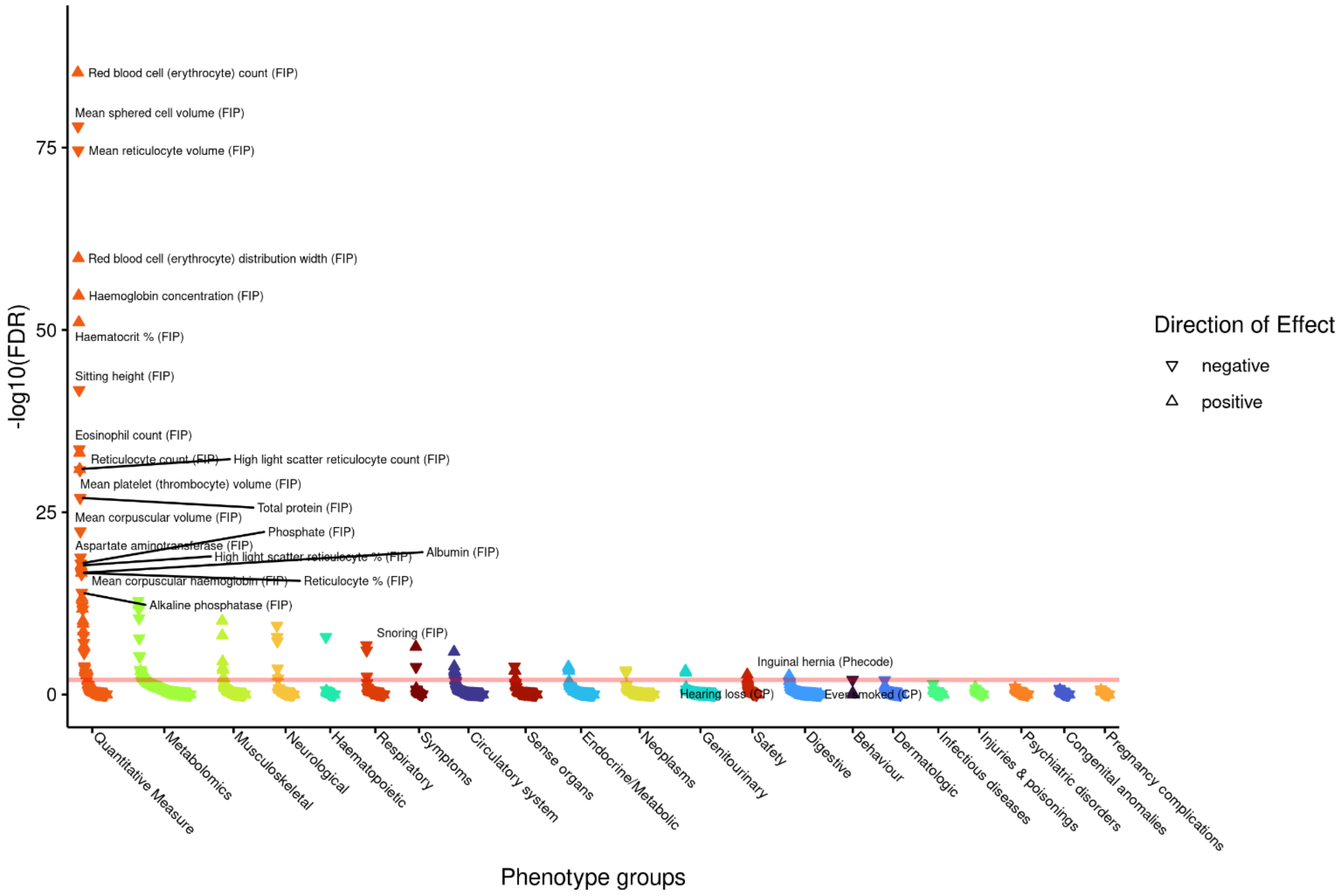

r) *MECOM* rs879394 – aligned to $FEV_1$ decreasing T allele

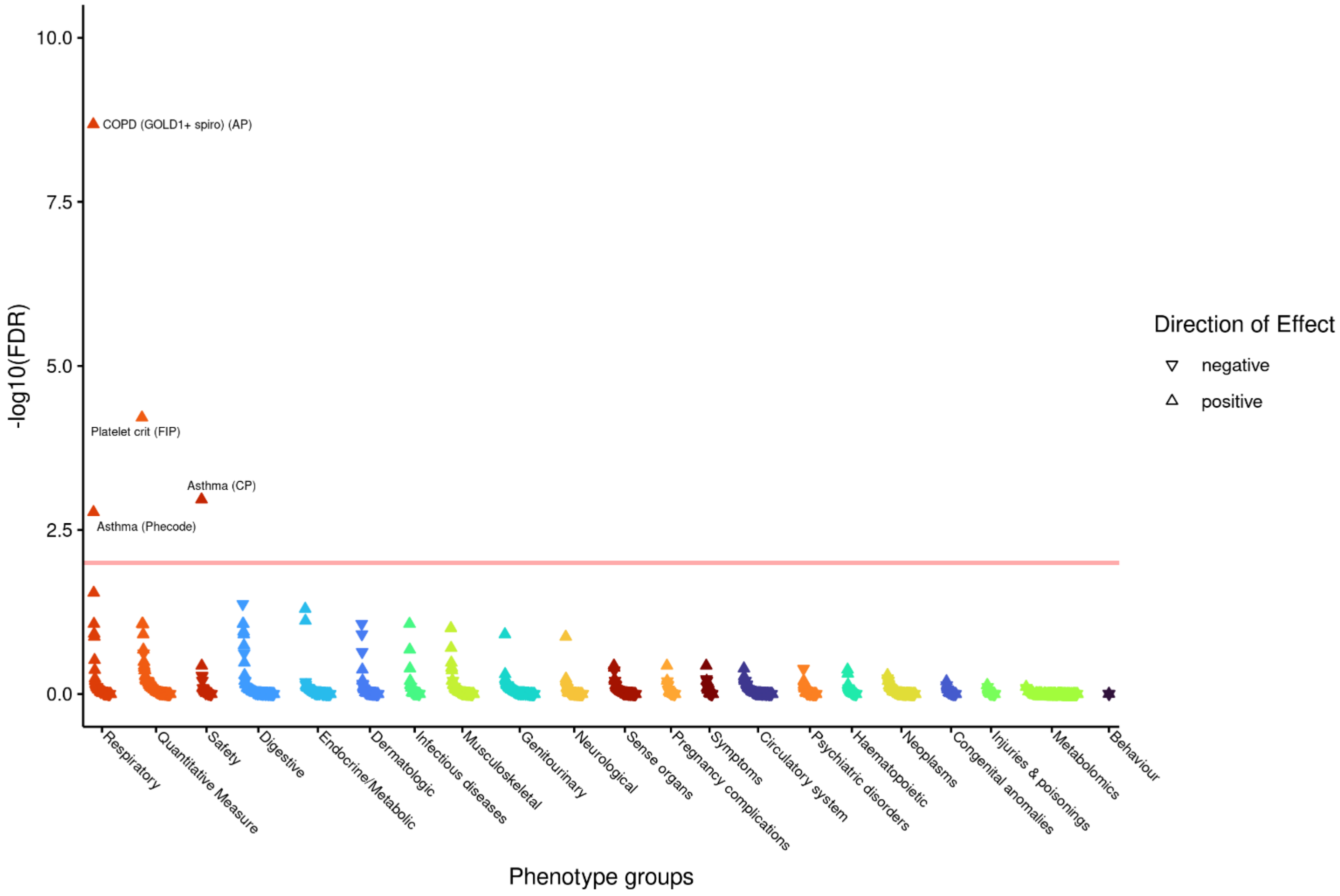

s) *NPNT* rs34712979 – aligned to $FEV_1$/FVC decreasing A allele

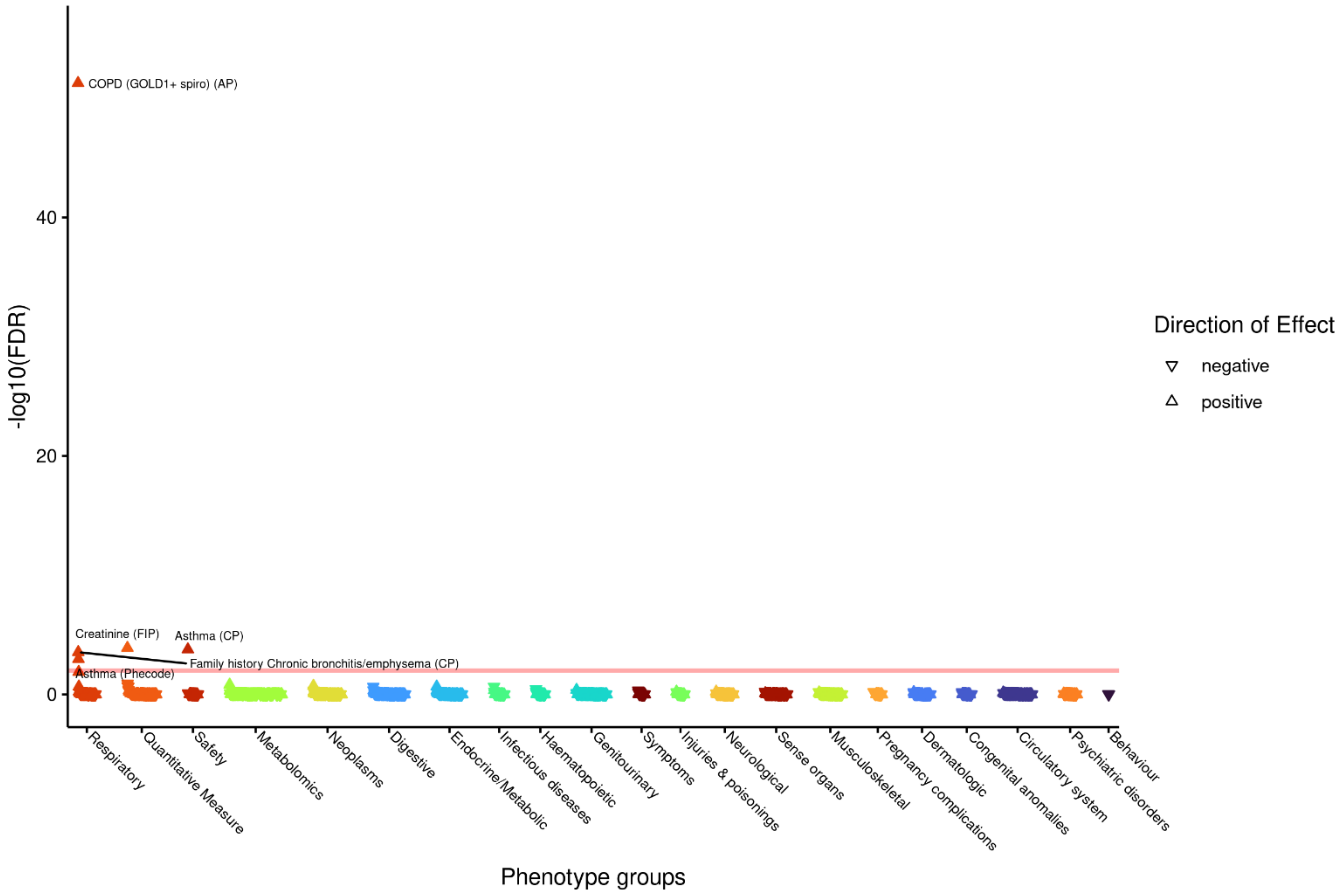

t) *SCARF2* rs5763025 - aligned to $FEV_1$/FVC decreasing A allele

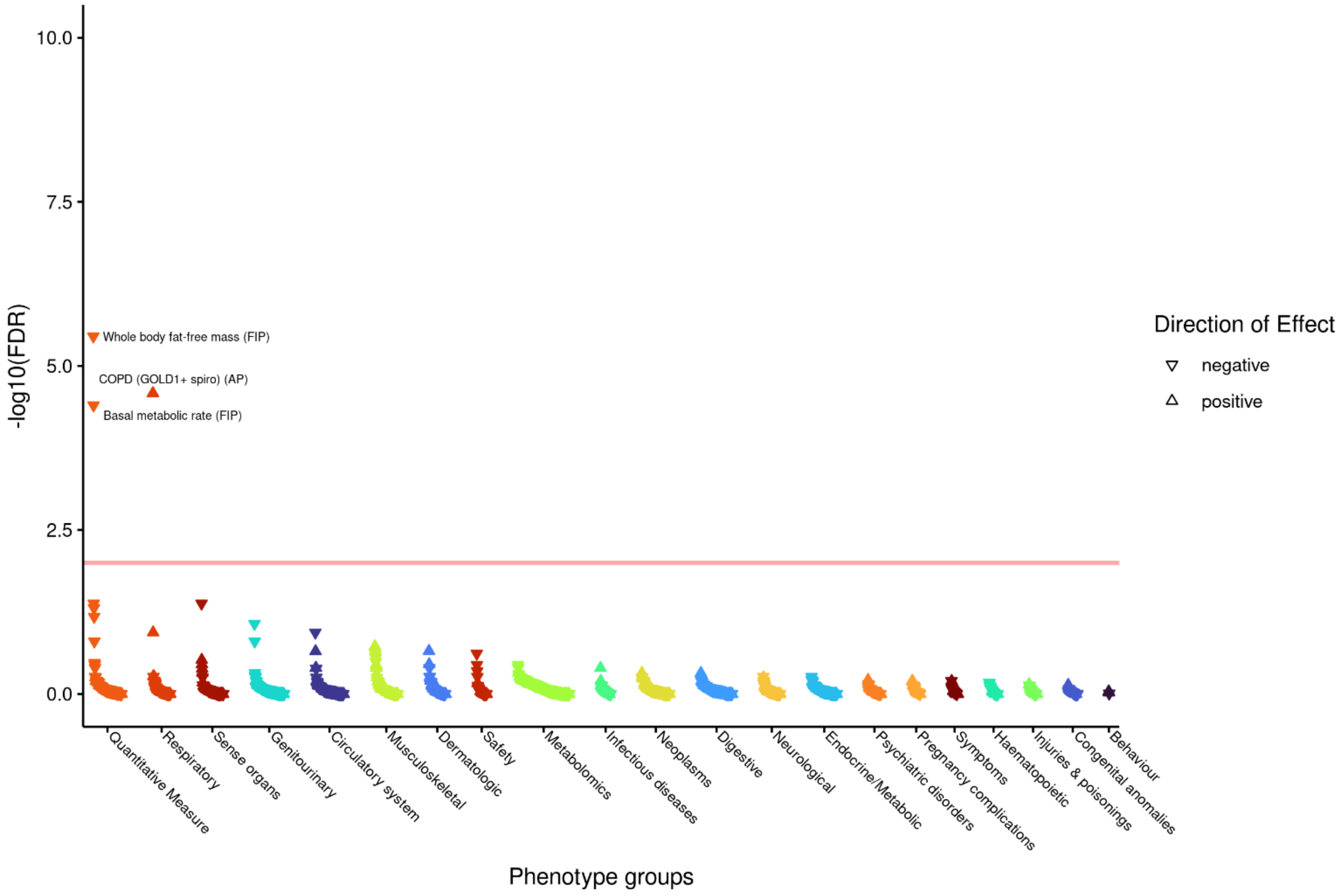

u) *SCMH1* rs2885697 – aligned to FVC decreasing T allele

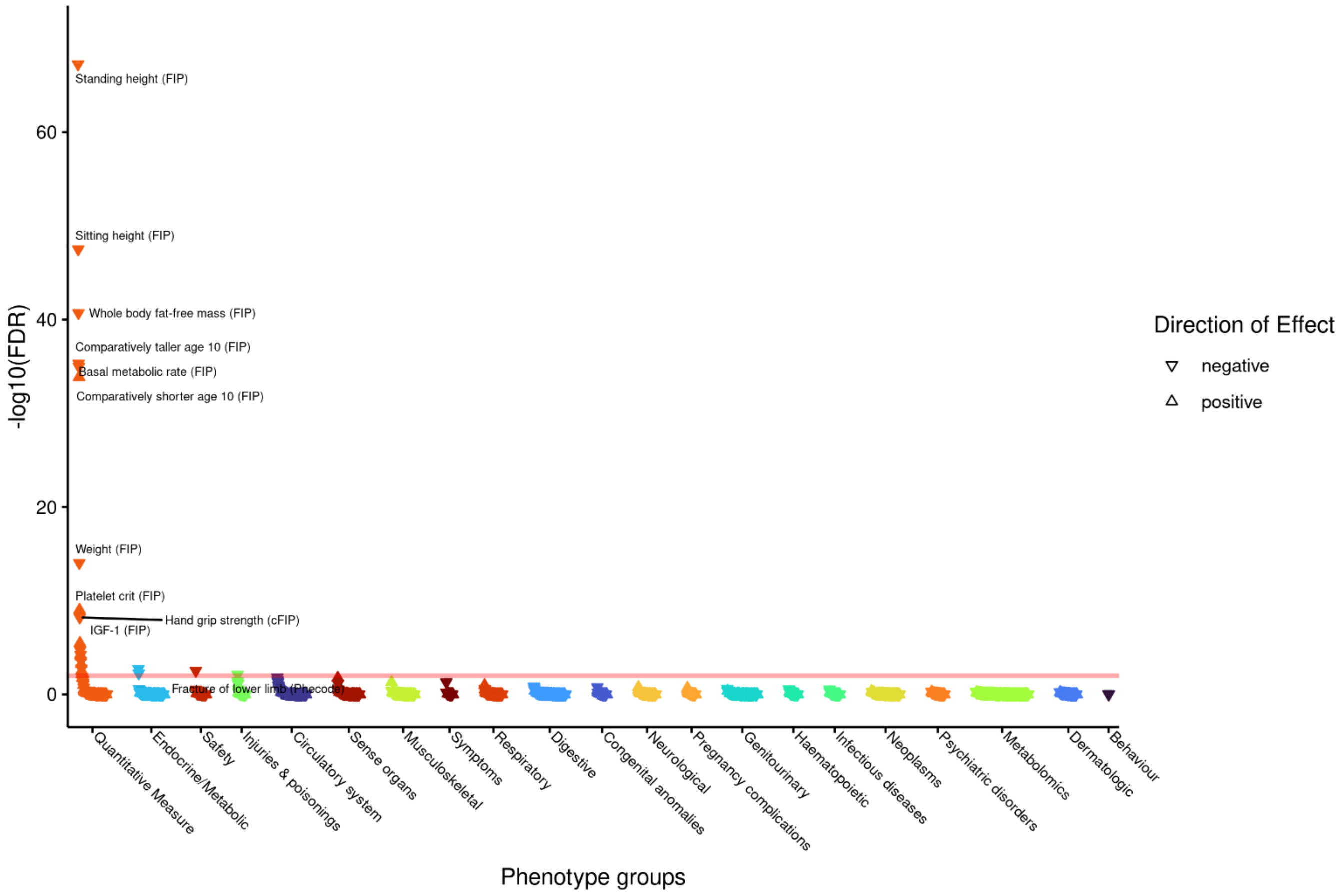

v) *SMAD3* rs35688953 – aligned to FVC decreasing C allele

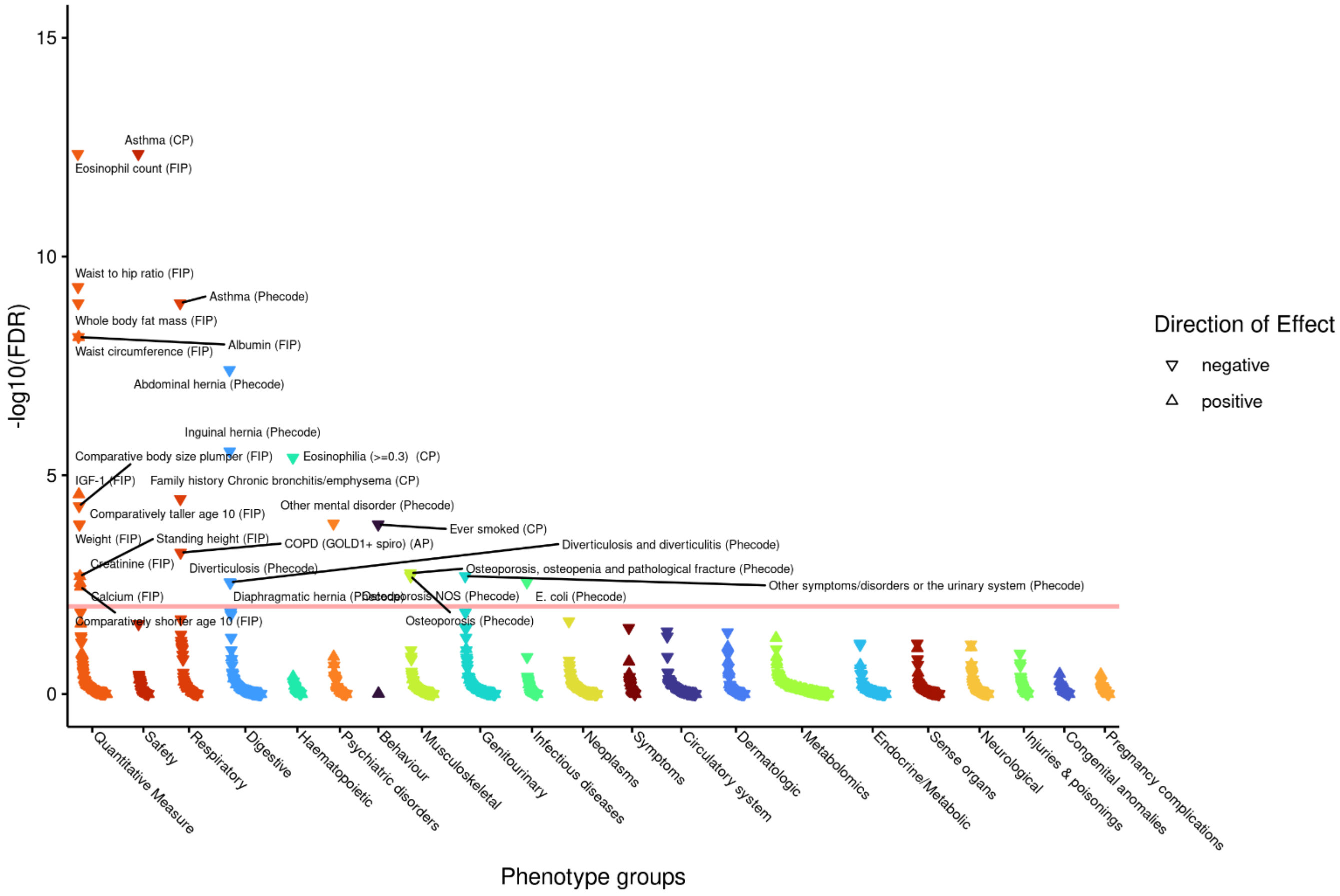

w) *STIM1* 11:4014295_GA_G – aligned to $FEV_1$/FVC decreasing A insertion

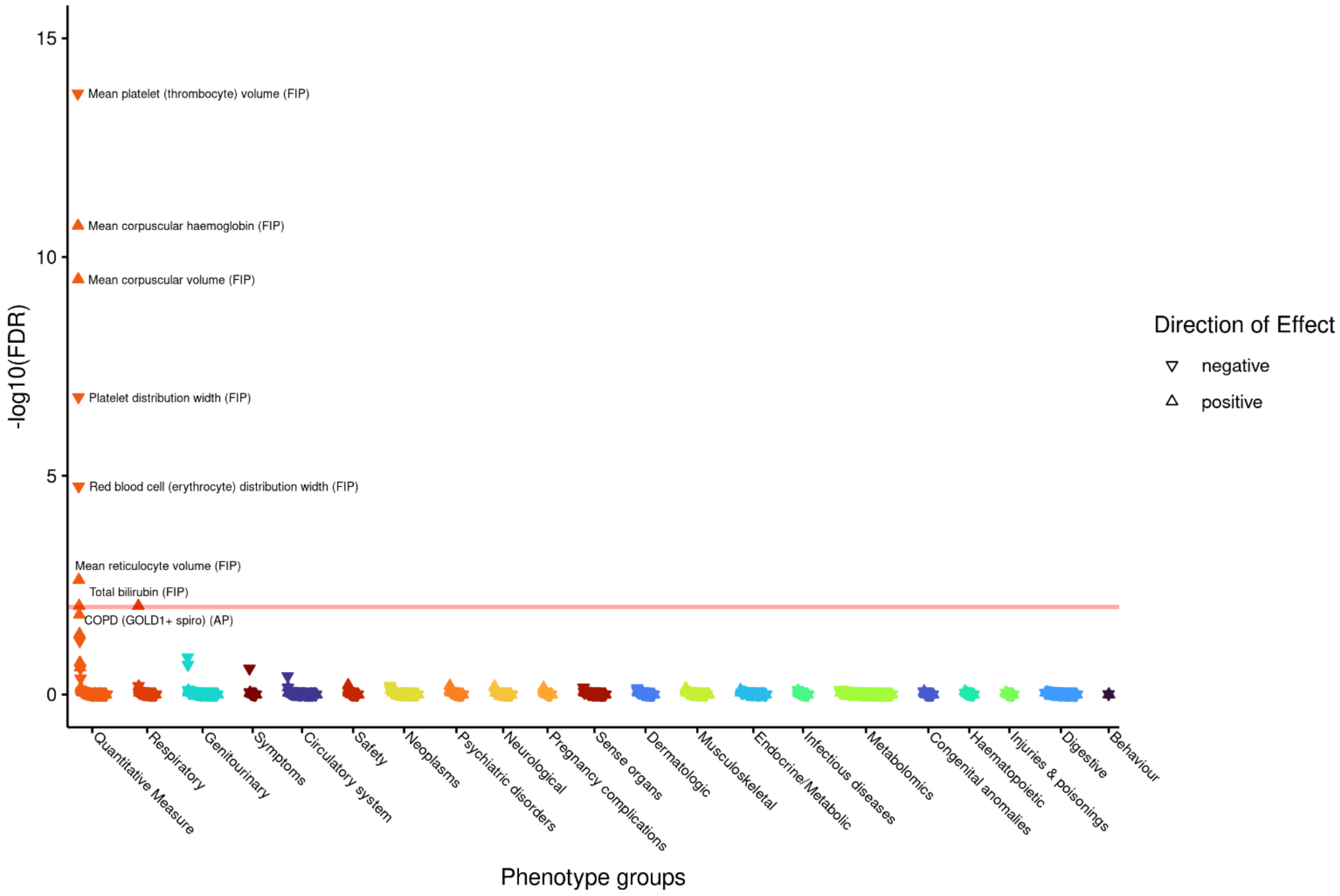

**x)** ***TGFB2* rs10482810 – aligned to $FEV_1$/FVC decreasing C allele**

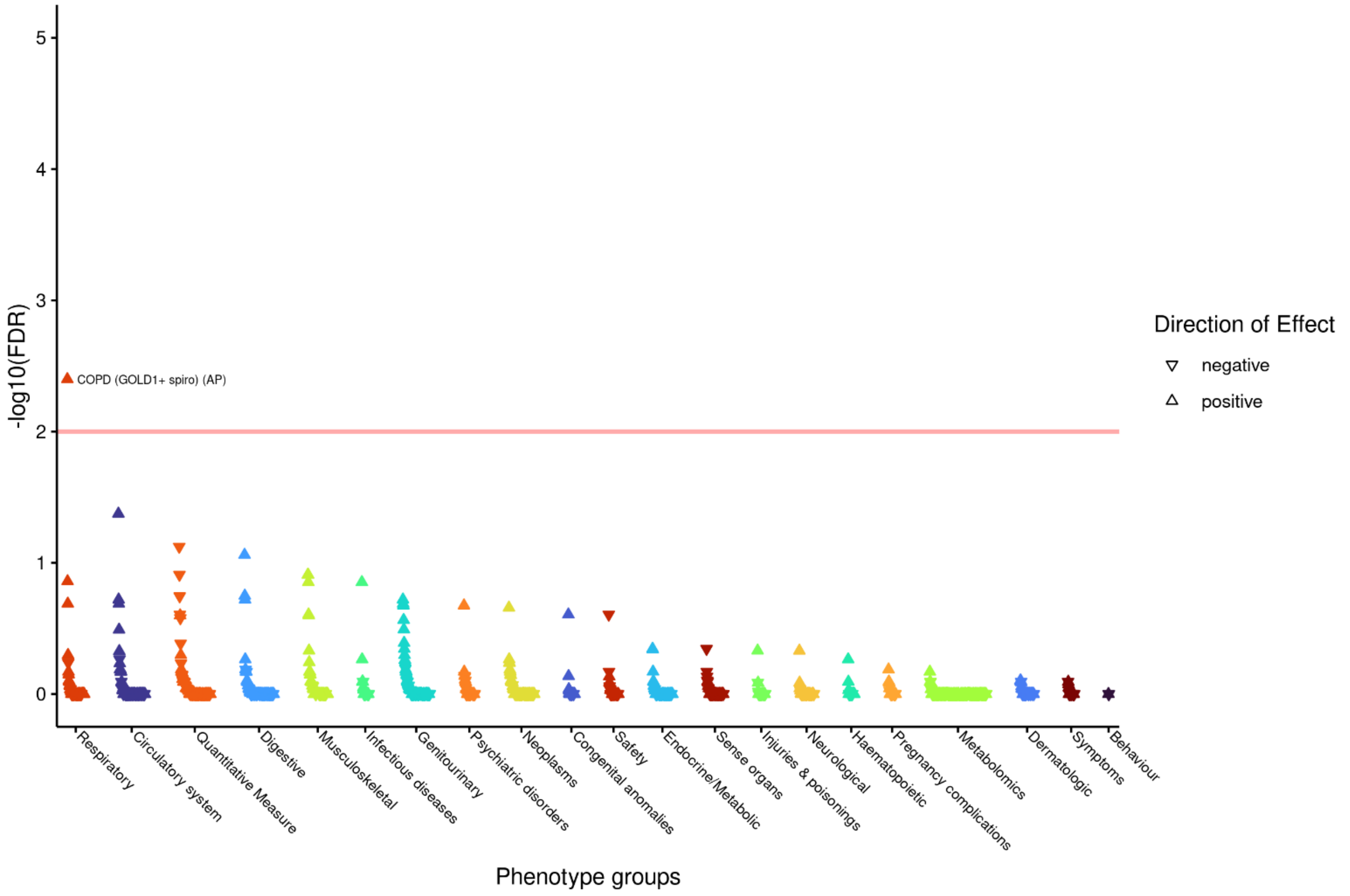

**y)** ***TNS1* rs2571445 – aligned to $FEV_1$ decreasing A allele**

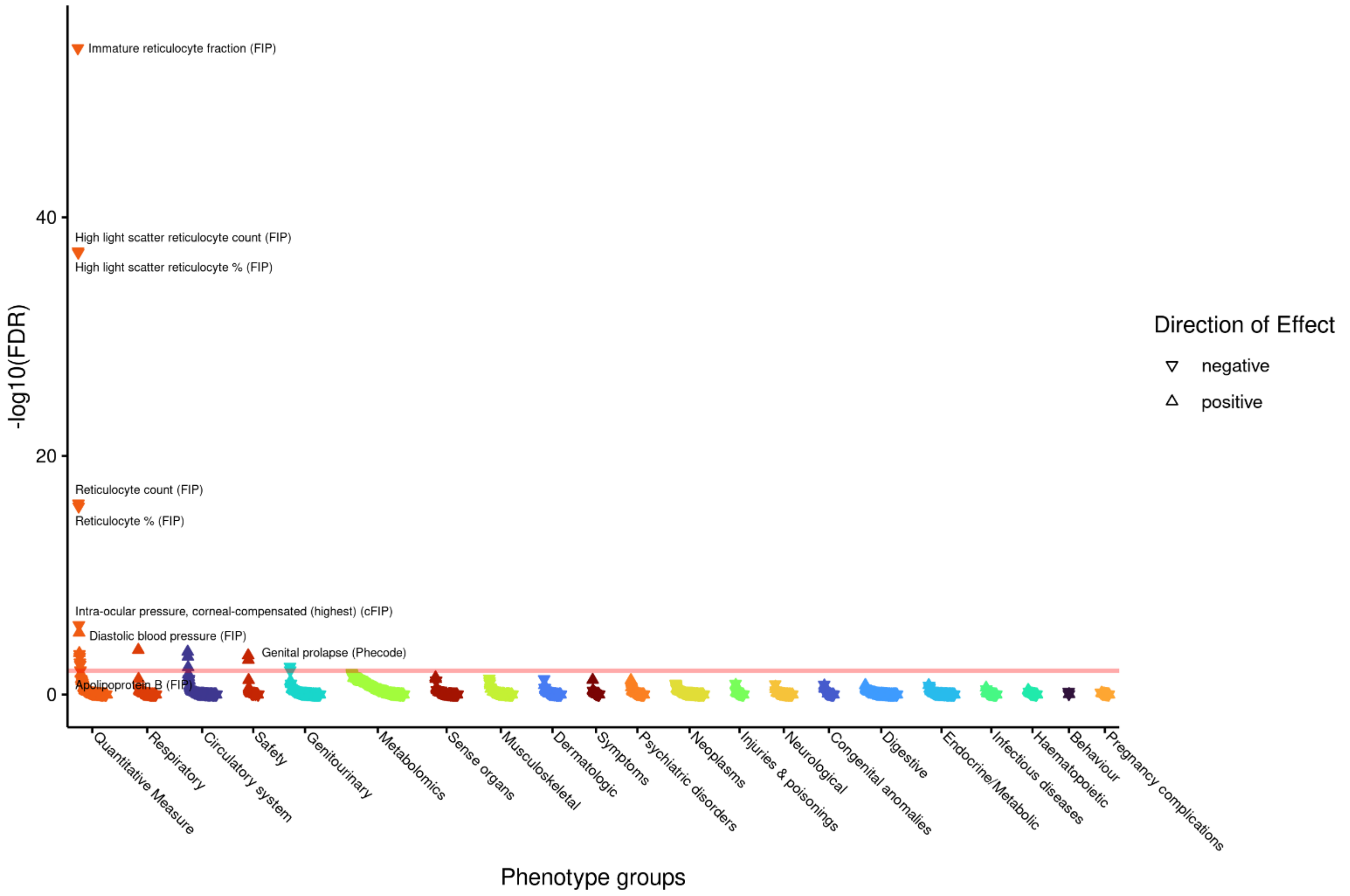

#### z) FGFR1 rs881299 – aligned to $FEV_1$/FVC decreasing T allele

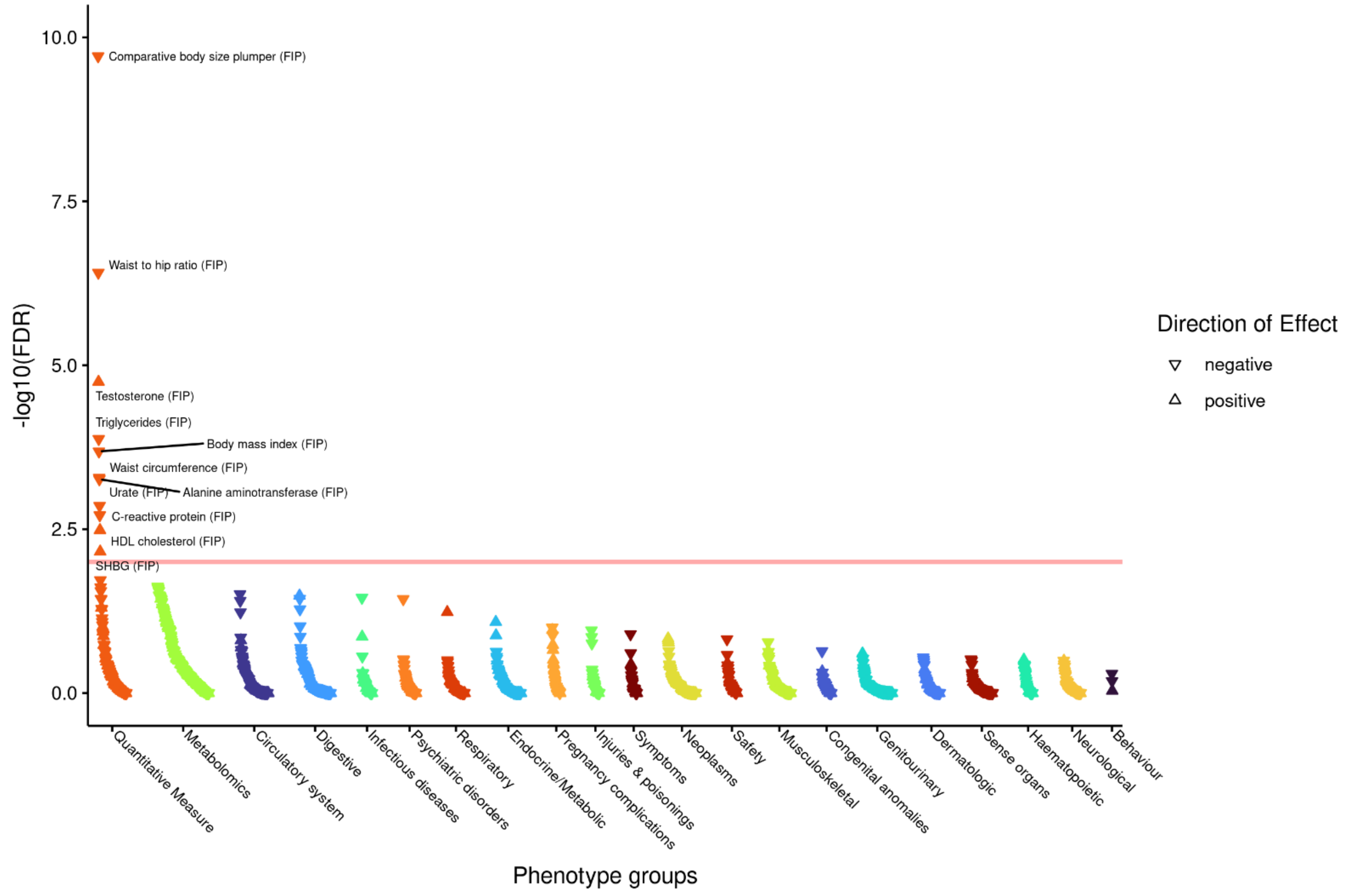

#### aa) FGFR1 rs881299 – aligned to $FEV_1$/FVC decreasing T allele (male only)

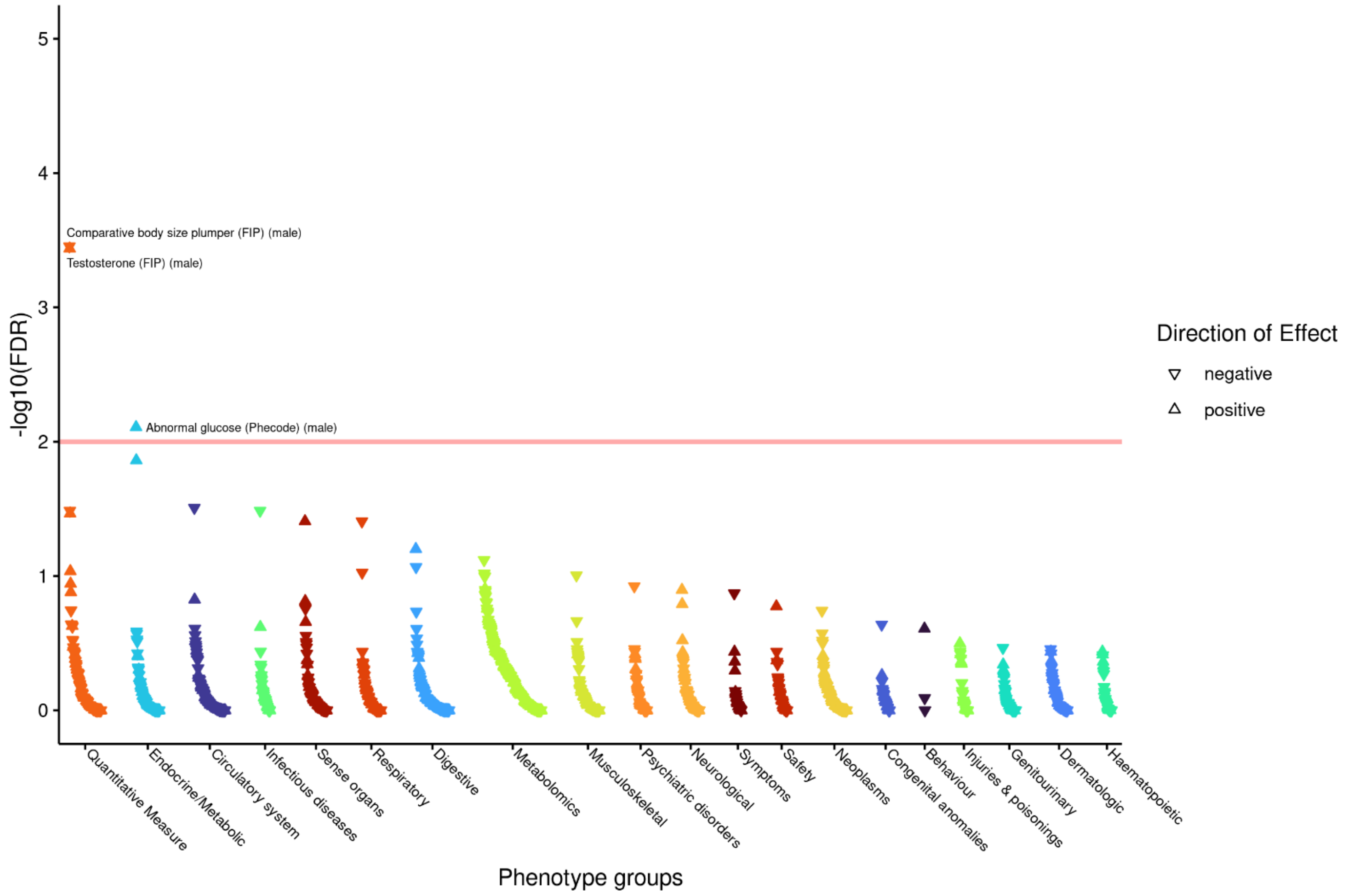

**ab) FGFR1 rs881299 – aligned to FEV$_1$/FVC decreasing T allele (female only)**

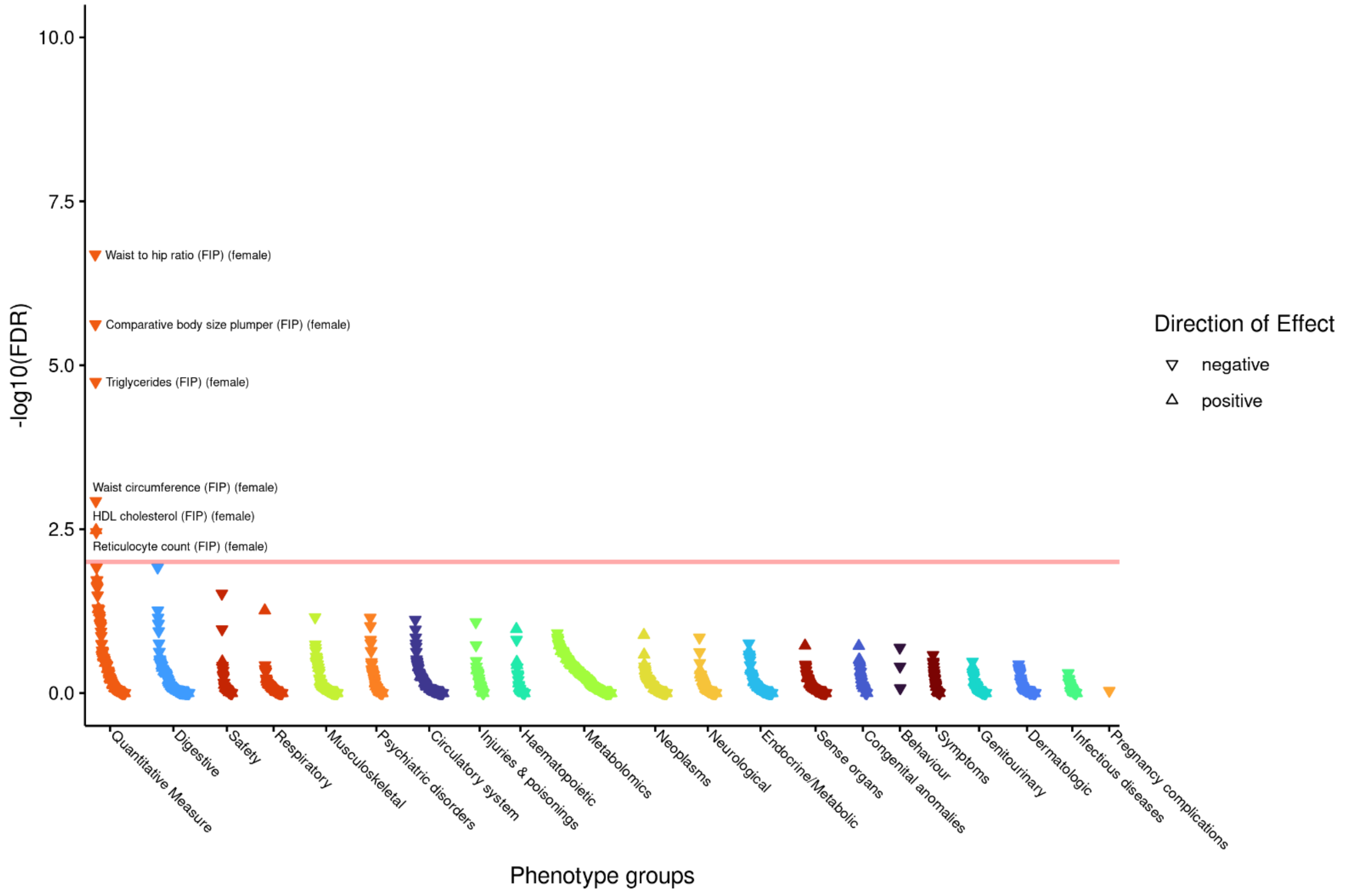

**ac) SOS2 rs72681869 – aligned to FVC decreasing G allele**

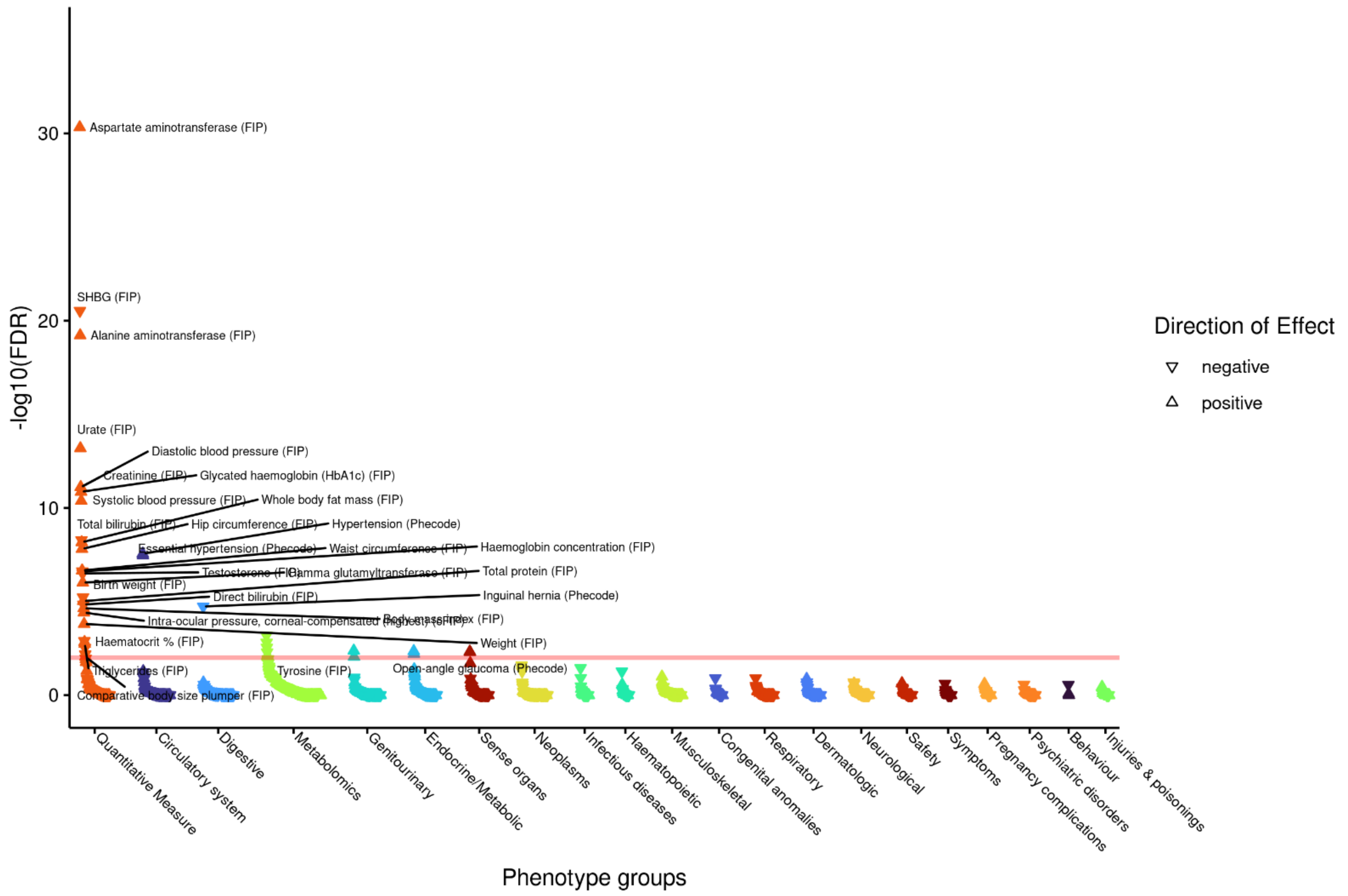

**ad) SOS2 rs72681869 – aligned to FVC decreasing G allele (male only)**

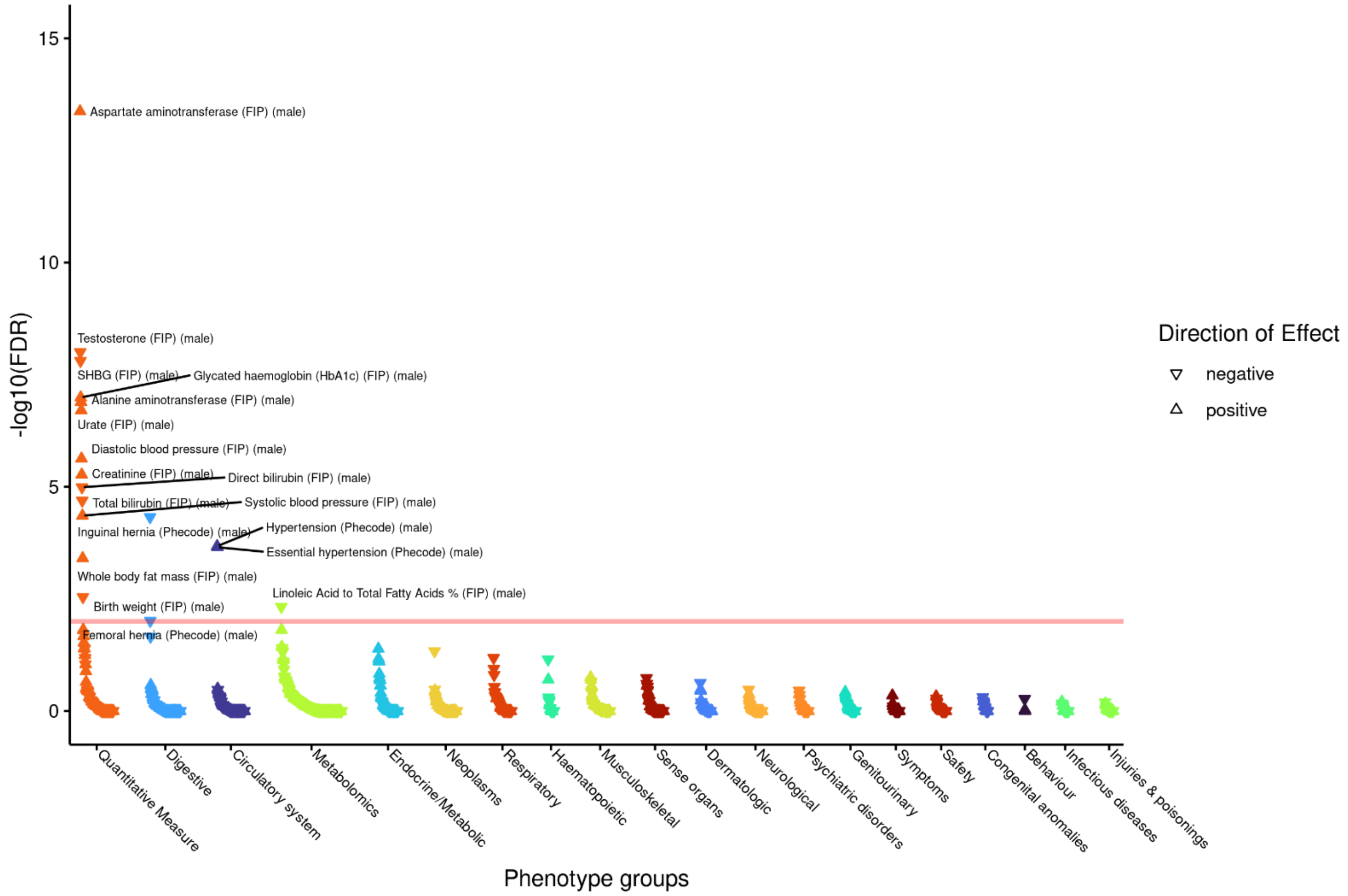

**ae) SOS2 rs72681869 – aligned to FVC decreasing G allele (female only)**

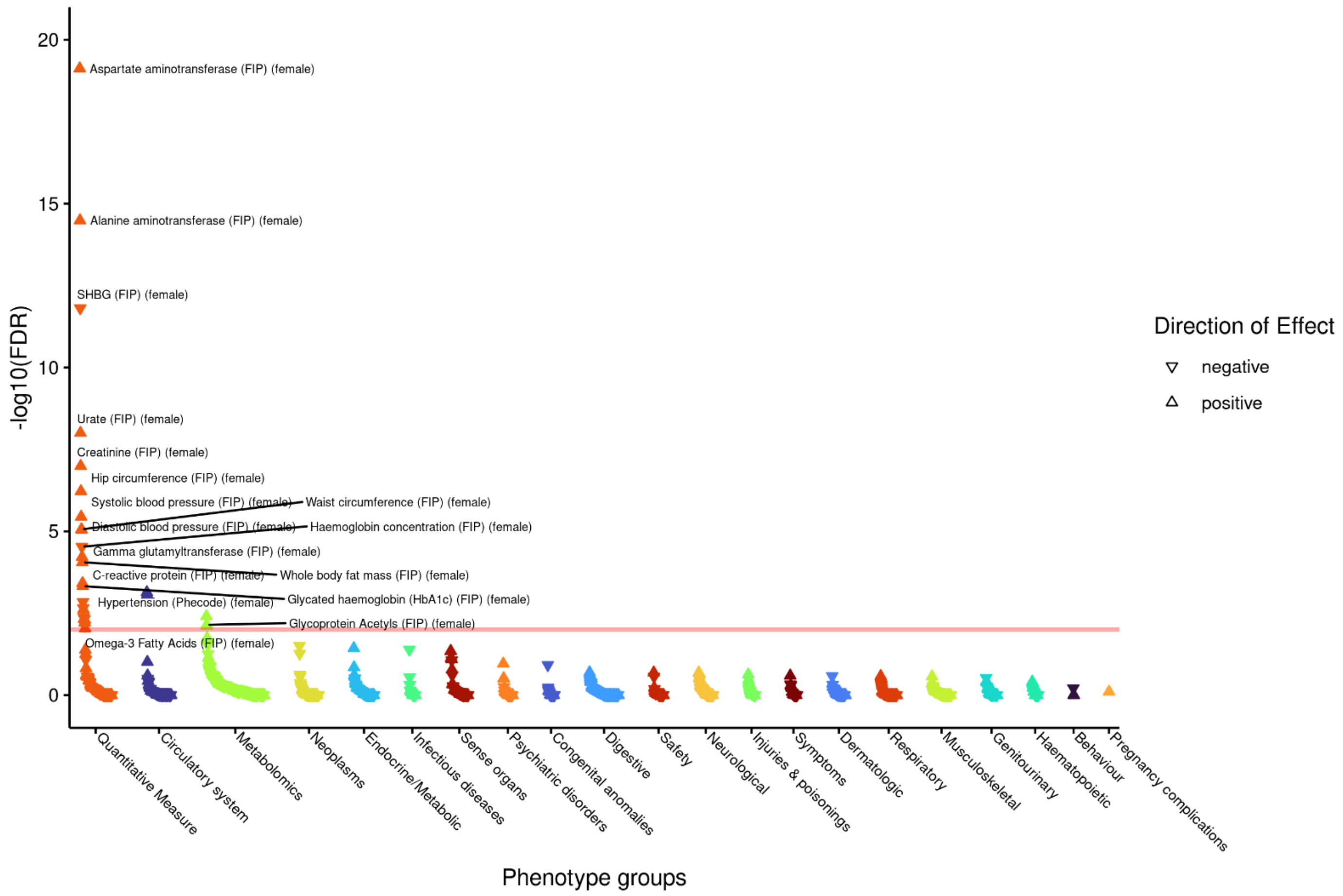

**Supplementary Figure 15: Pathway partitioned PheWAS results**

A genetic risk score was created for each of the 29 pathways by including for each gene in the pathway, the variant with the most significant P value for the trait that implicates the gene. Weights are the multi-ancestry meta-regression effect sizes for $FEV_1$/FVC. The data source ConsensuspathDB used for the pathway definition is in brackets. Only 21 of 29 pathway GRS tested are shown that have at least 1 result with FDR <1% (Supplementary Table 27).

a) **Beta1 integrin cell surface interactions (PID)**

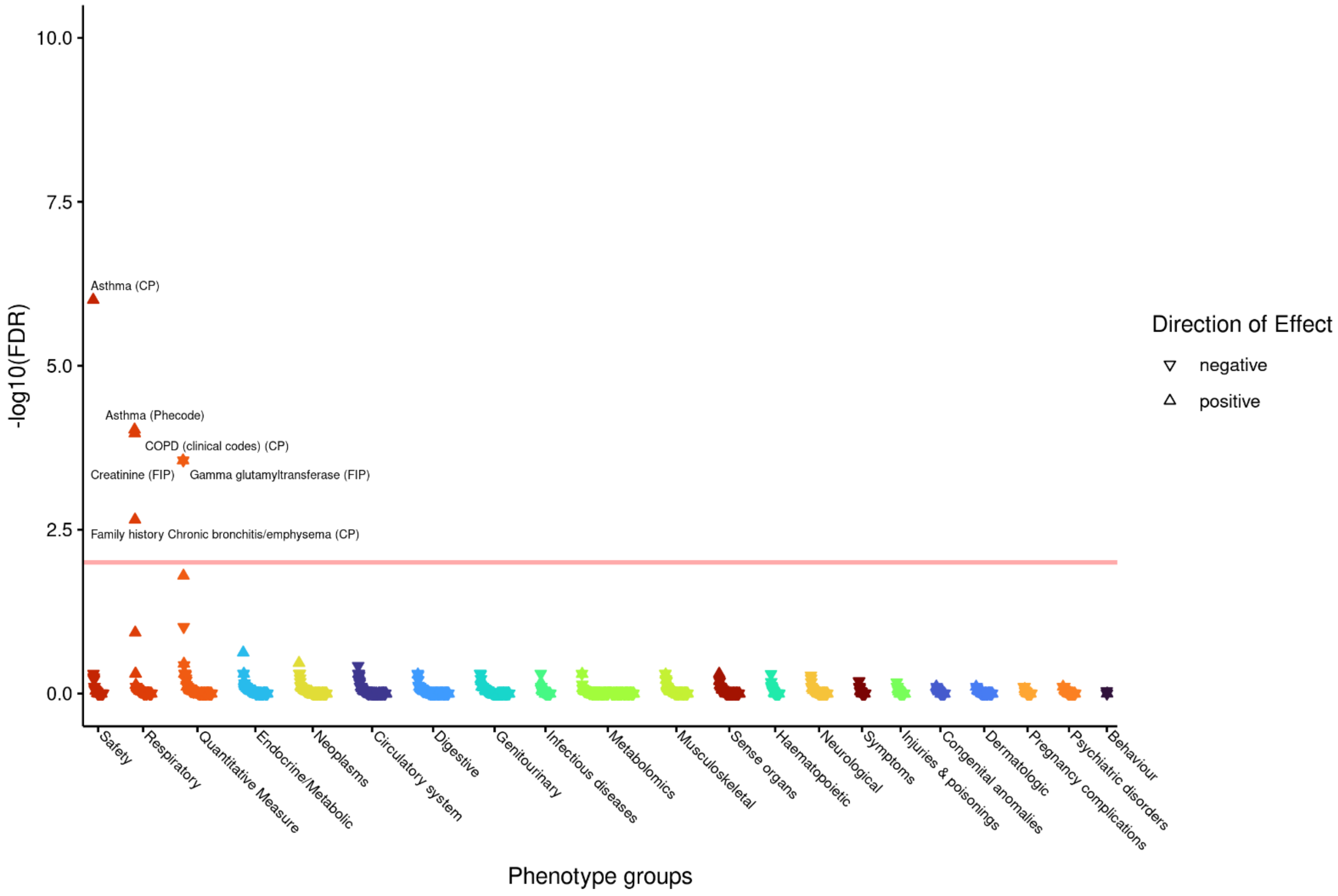

**b) Diseases of signal transduction by growth factor receptors and second messengers (Reactome)**

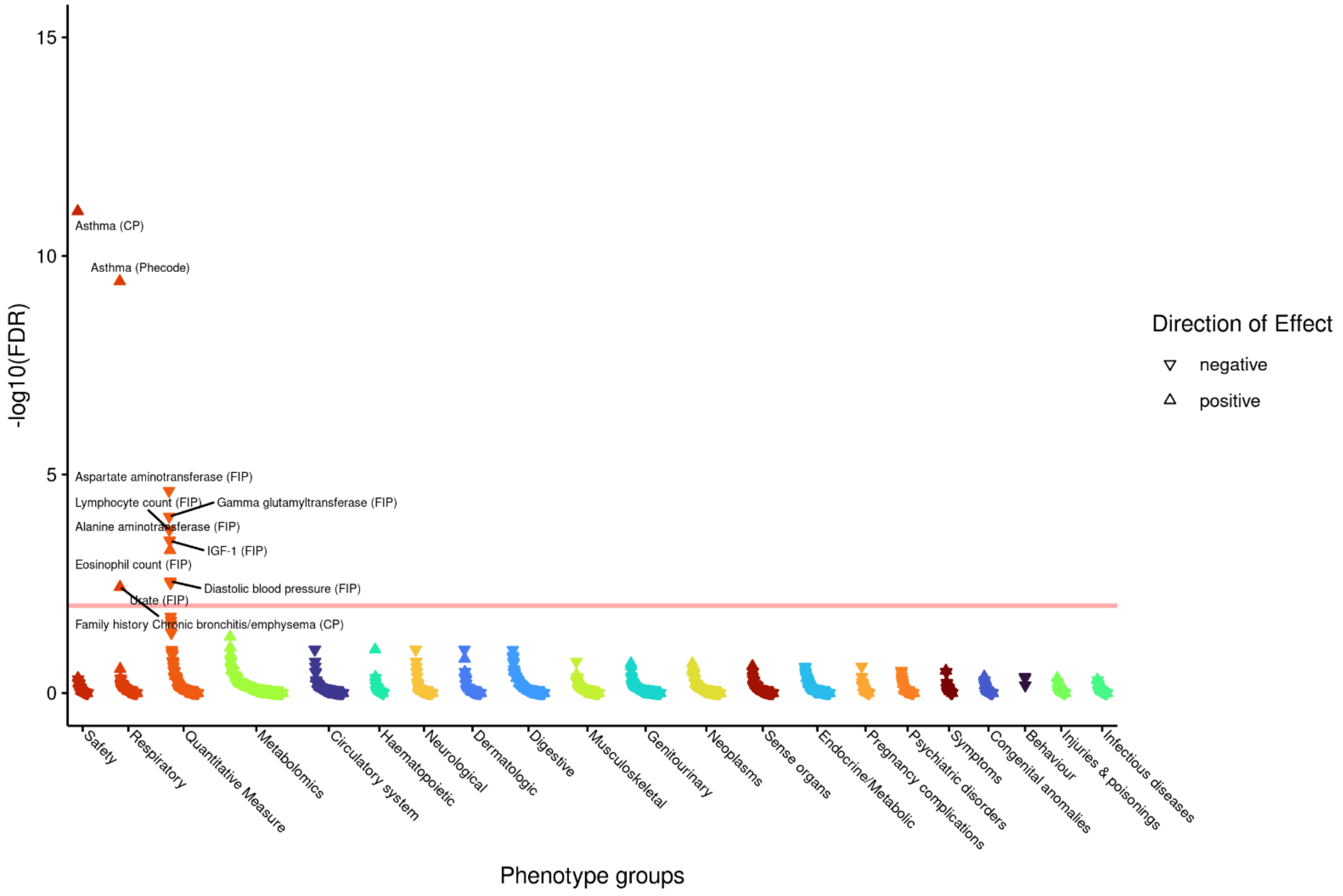

**c) ECM-receptor interaction Homo sapiens (human) (KEGG)**

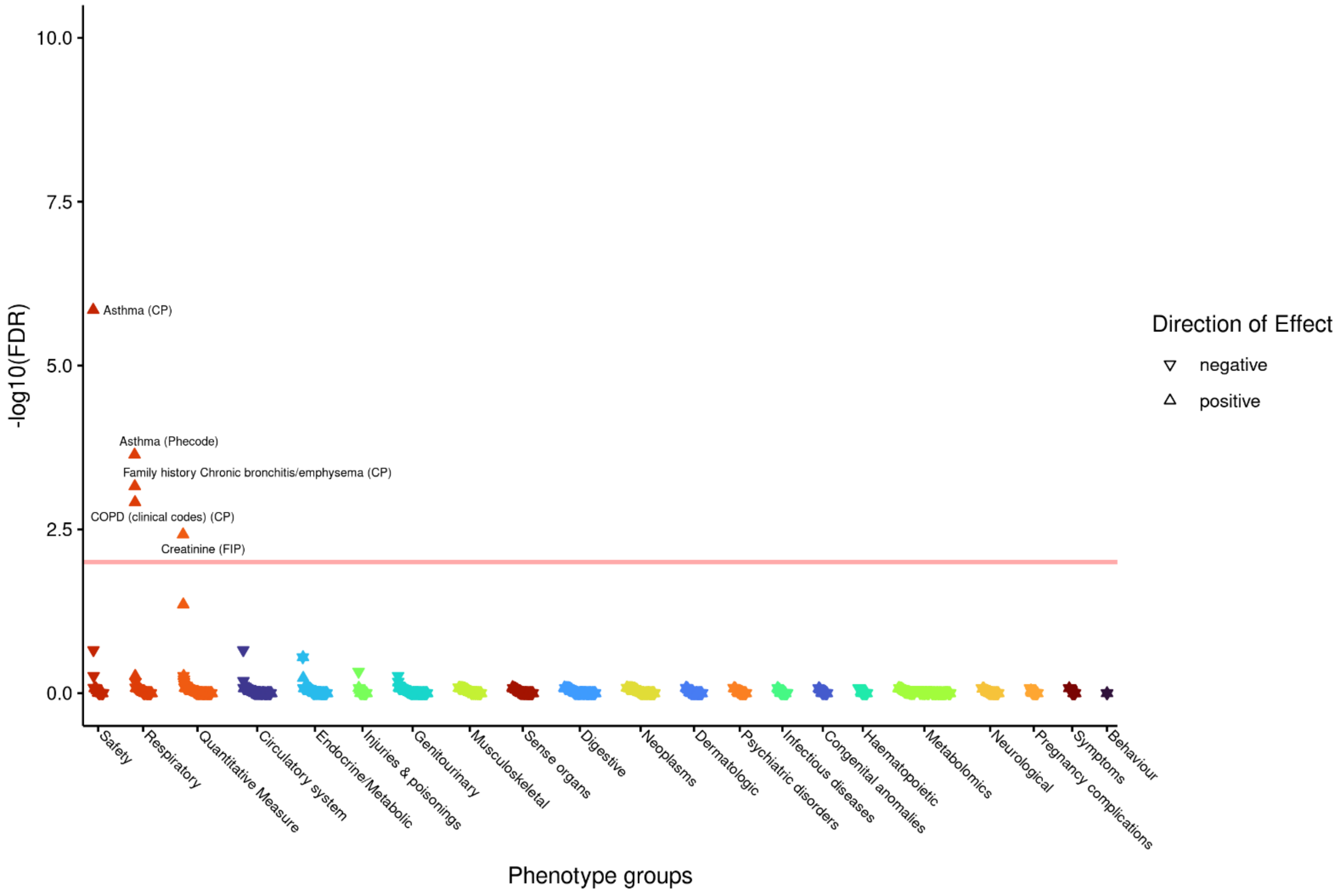

**d) EGFR Tyrosine Kinase Inhibitor Resistance (Wikipathways)**

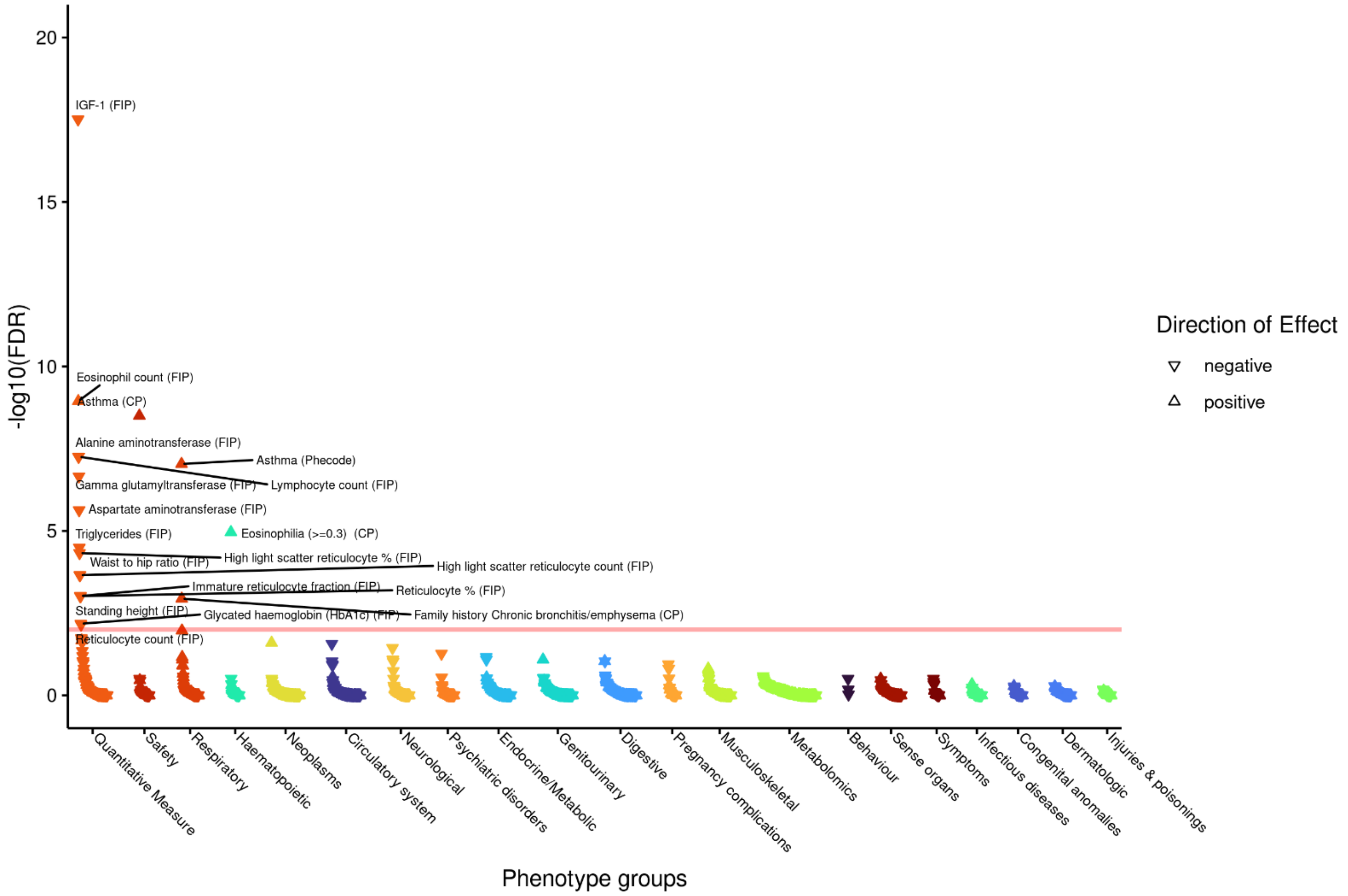

**e) Endochondral Ossification (Wikipathways)**

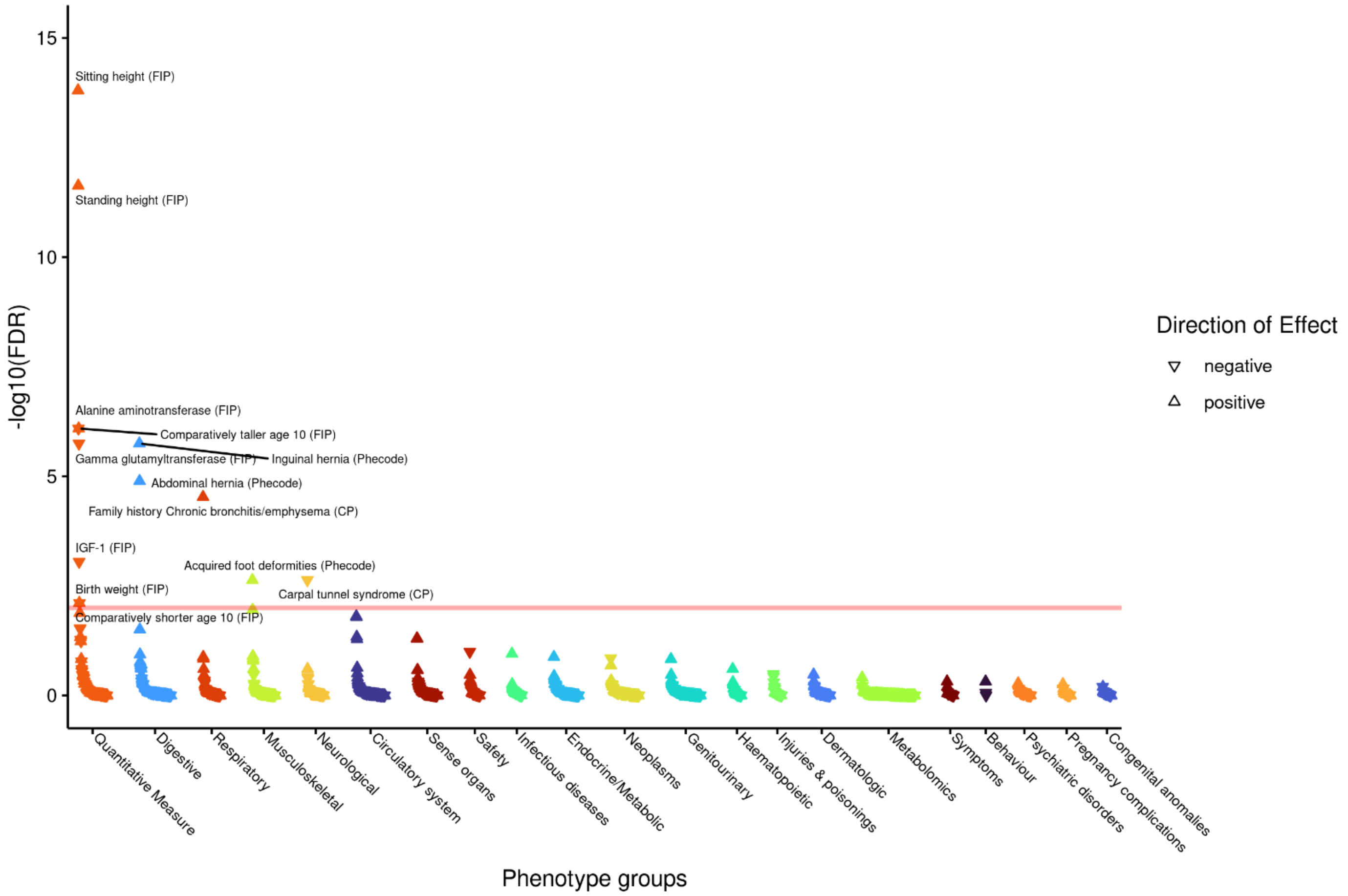

**f) Endochondral Ossification with Skeletal Dysplasias (Wikipathways)**

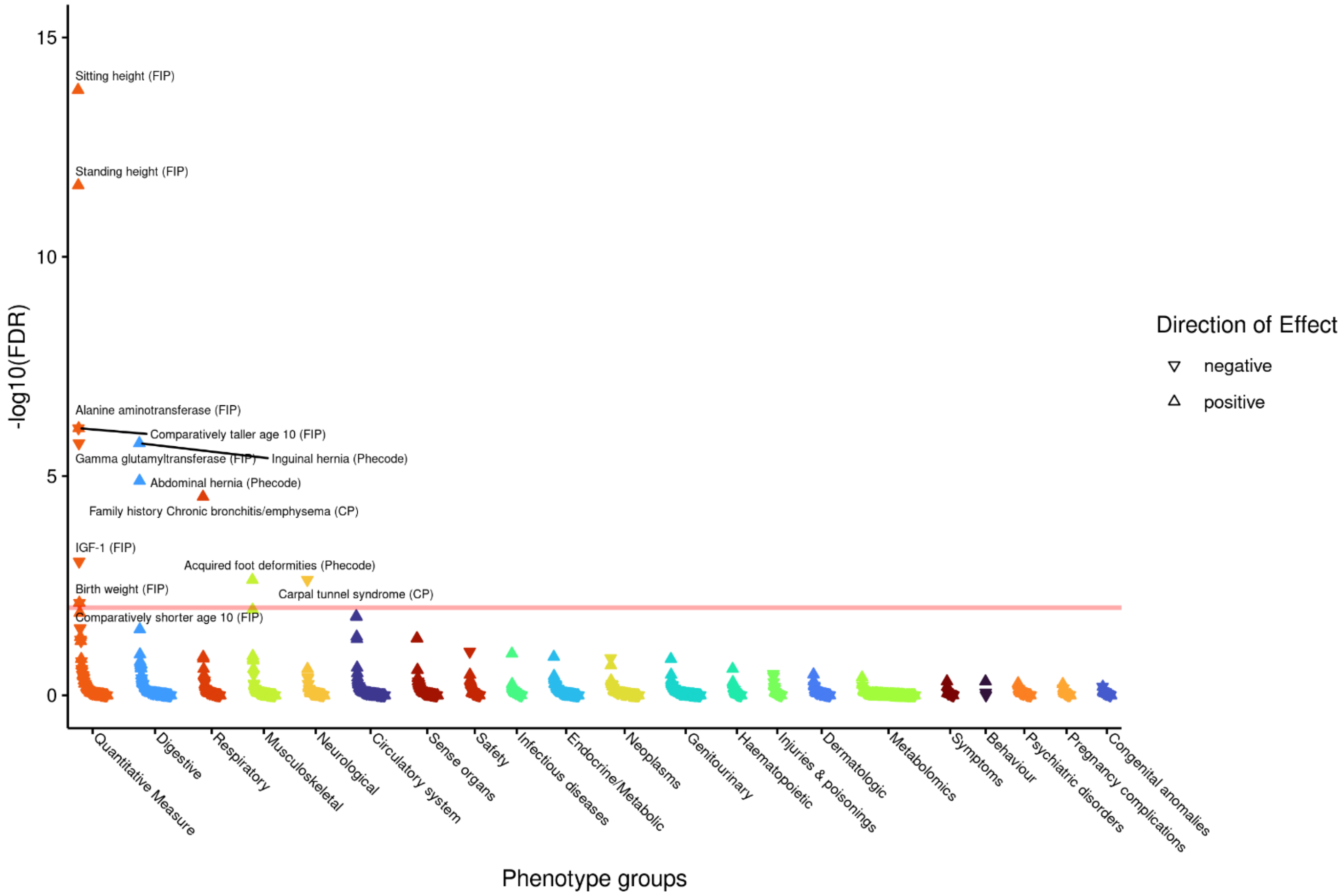

**g) ESC Pluripotency Pathways (Wikipathways)**

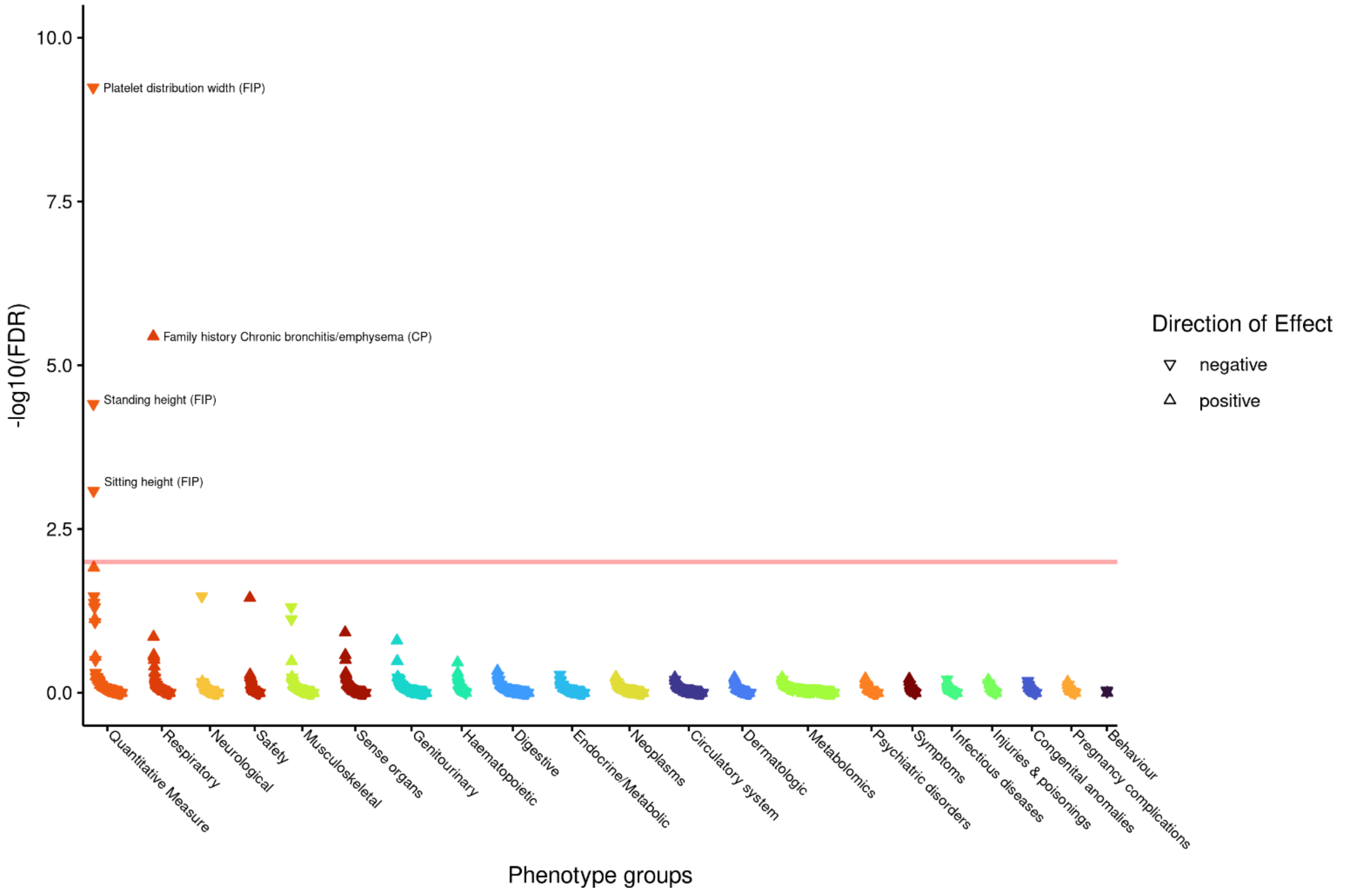

**h) Extracellular matrix organization (Reactome)**

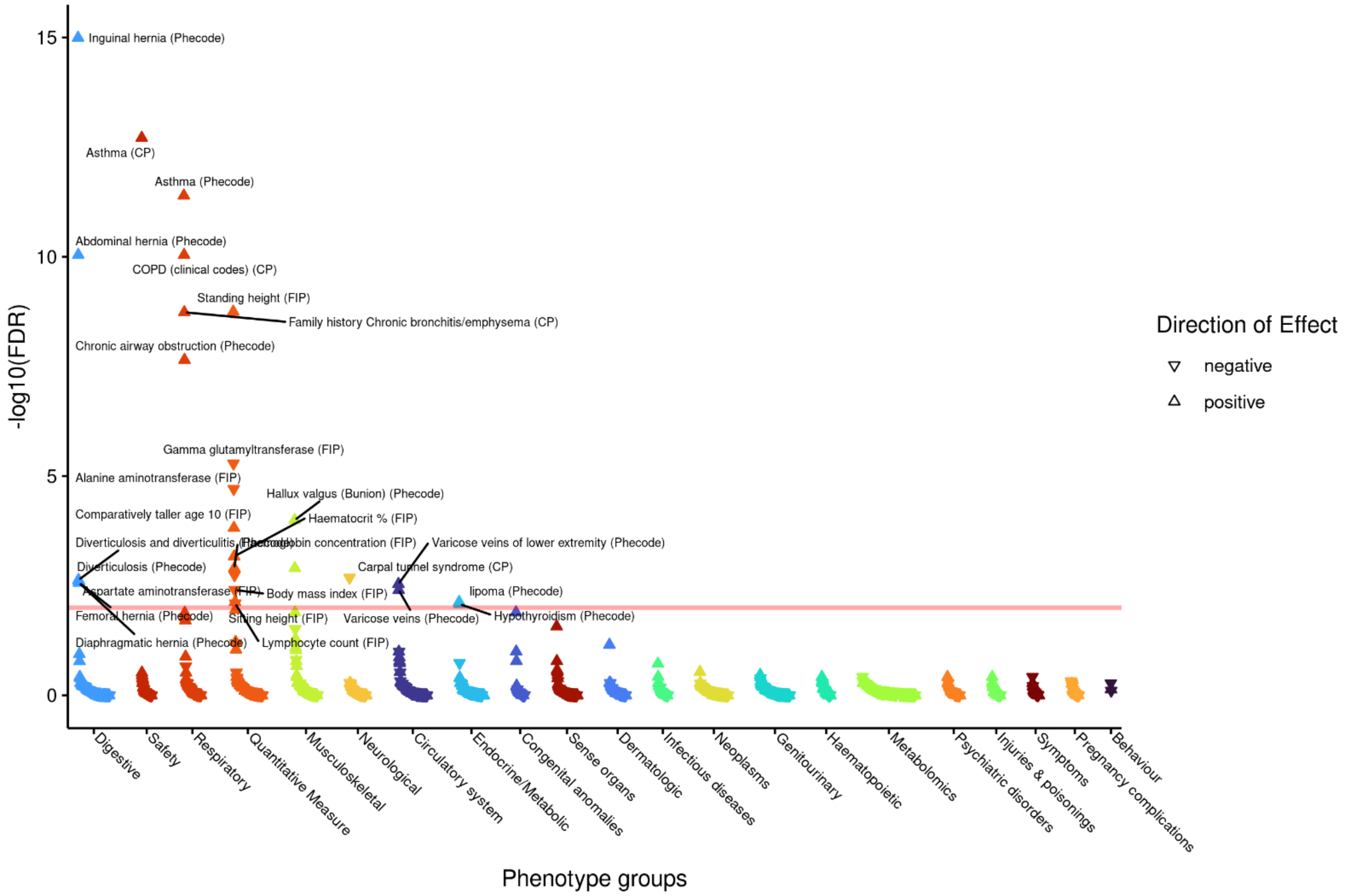

**i) Focal adhesion Homo sapiens (human) (KEGG)**

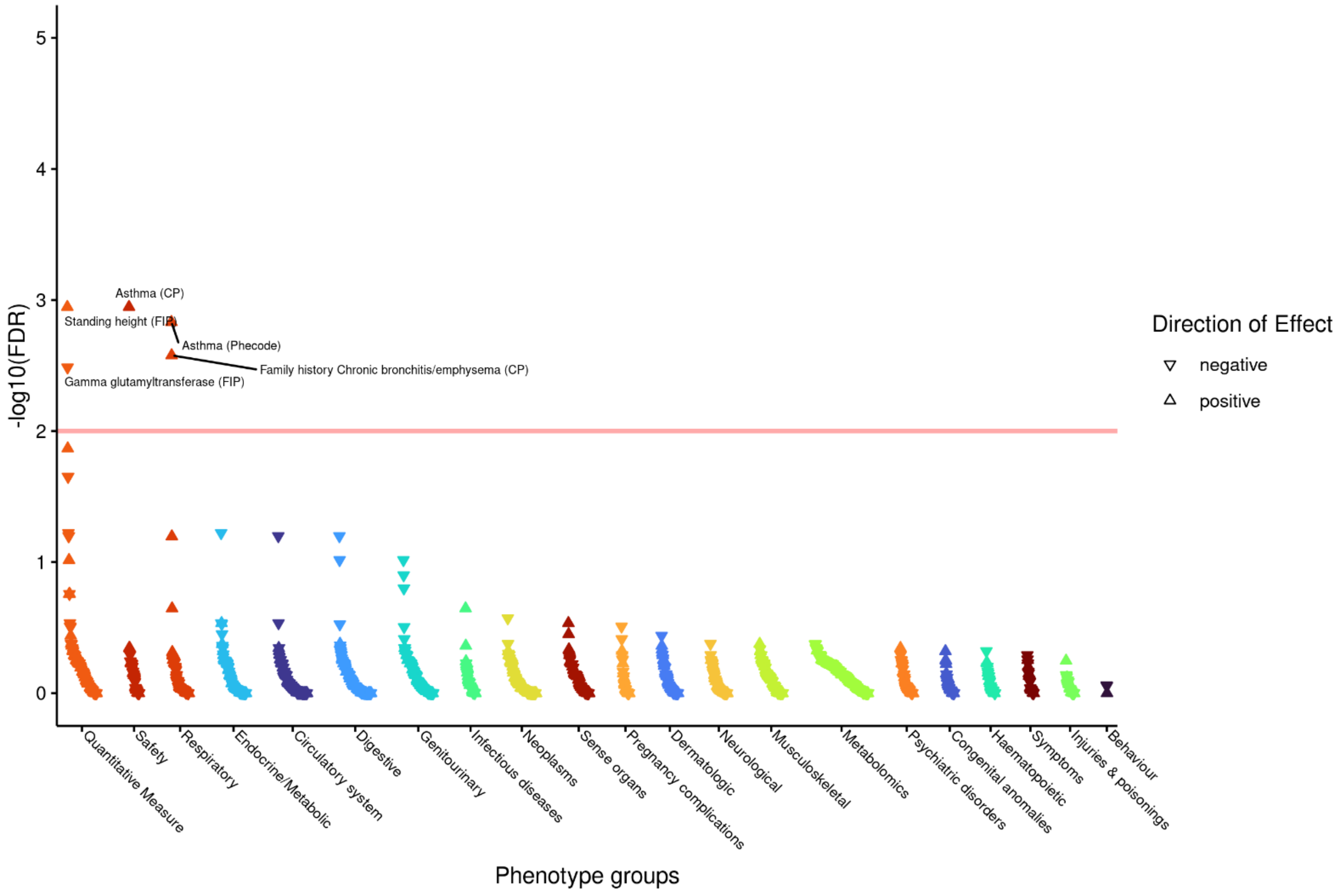

**j) Focal Adhesion (Wikipathways)**

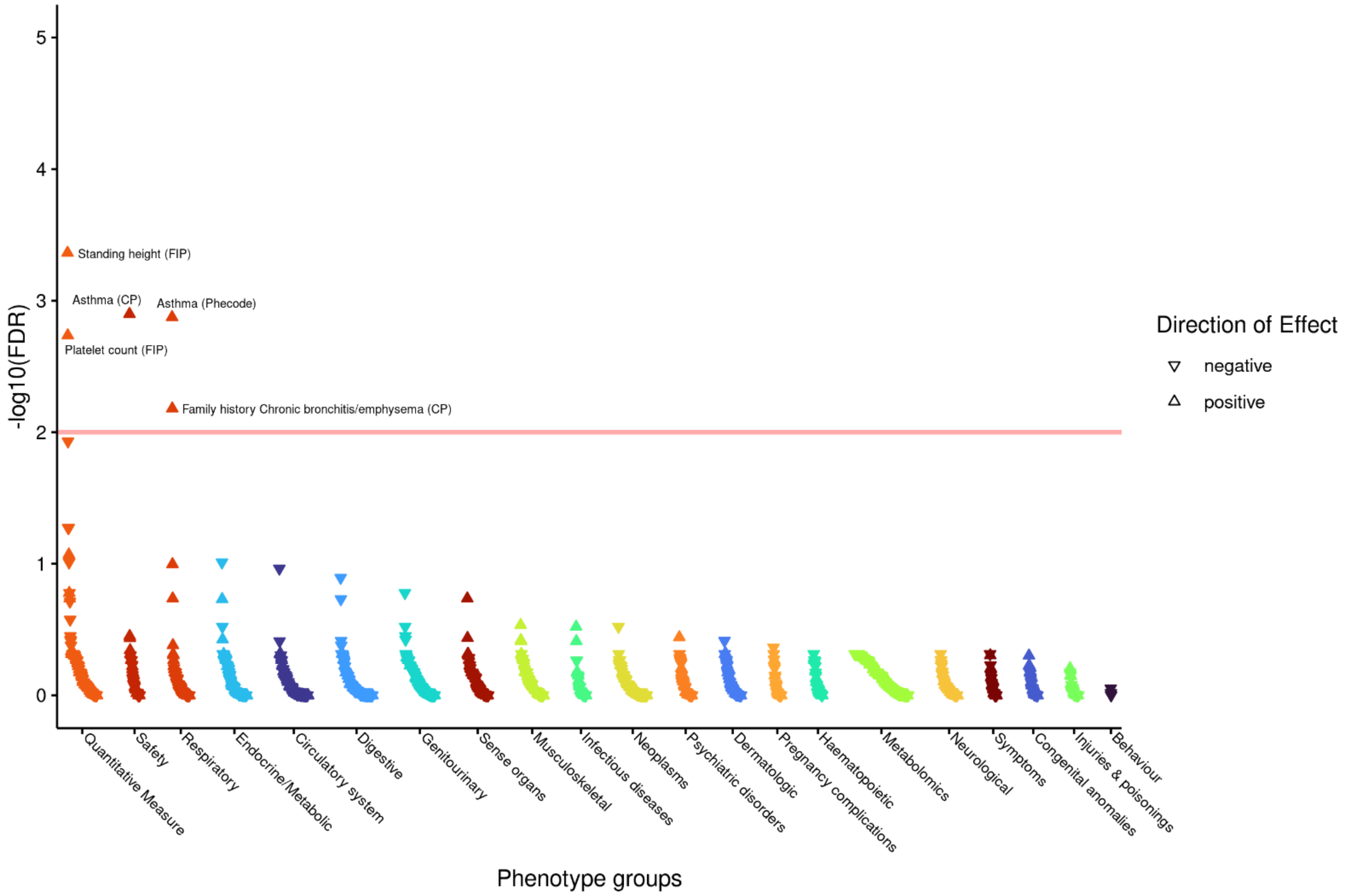

**k) Focal Adhesion-PI3K-Akt-mTOR-signaling pathway (Wikipathways)**

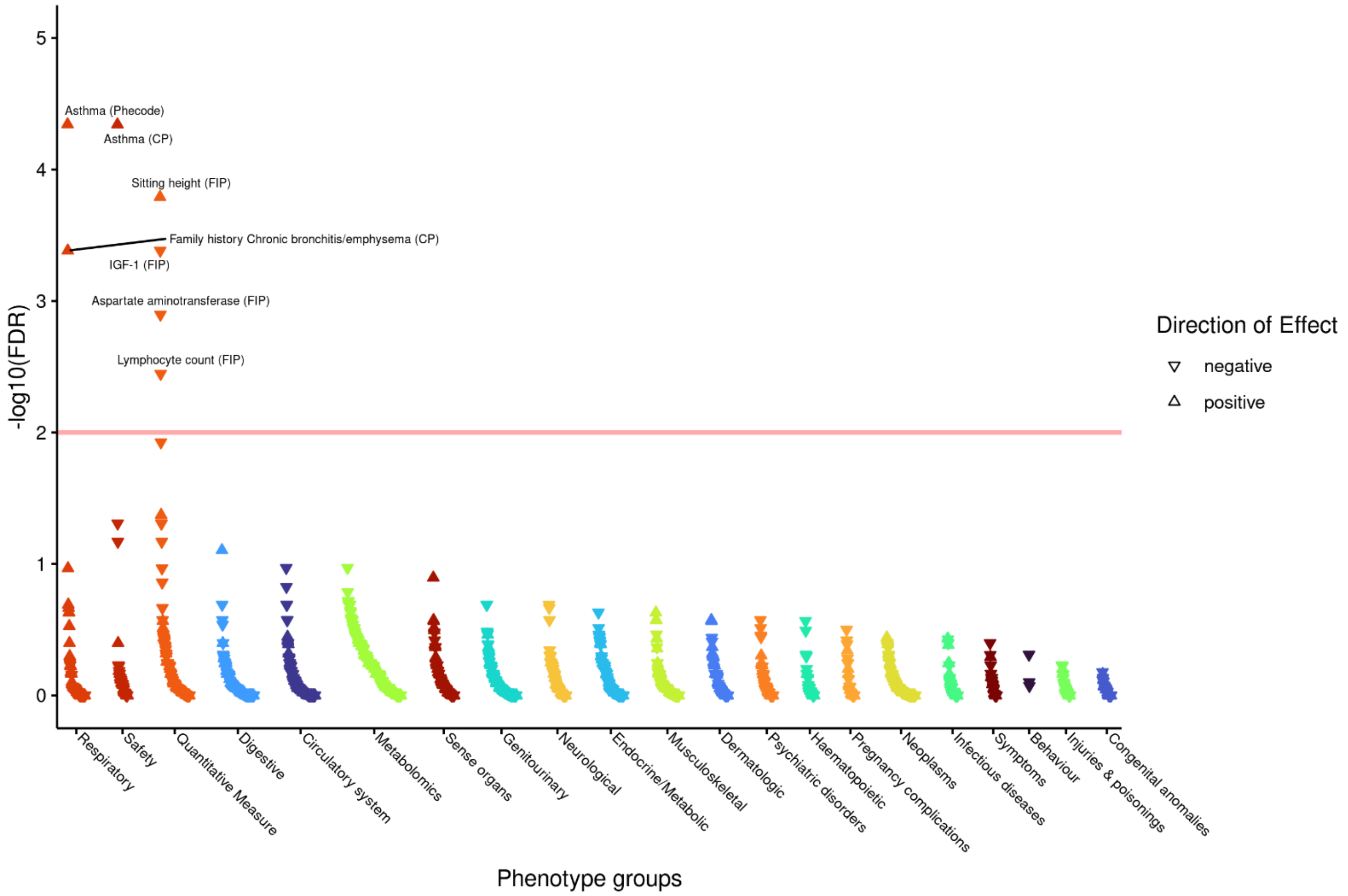

l) **Genes controlling nephrogenesis (Wikipathways)**

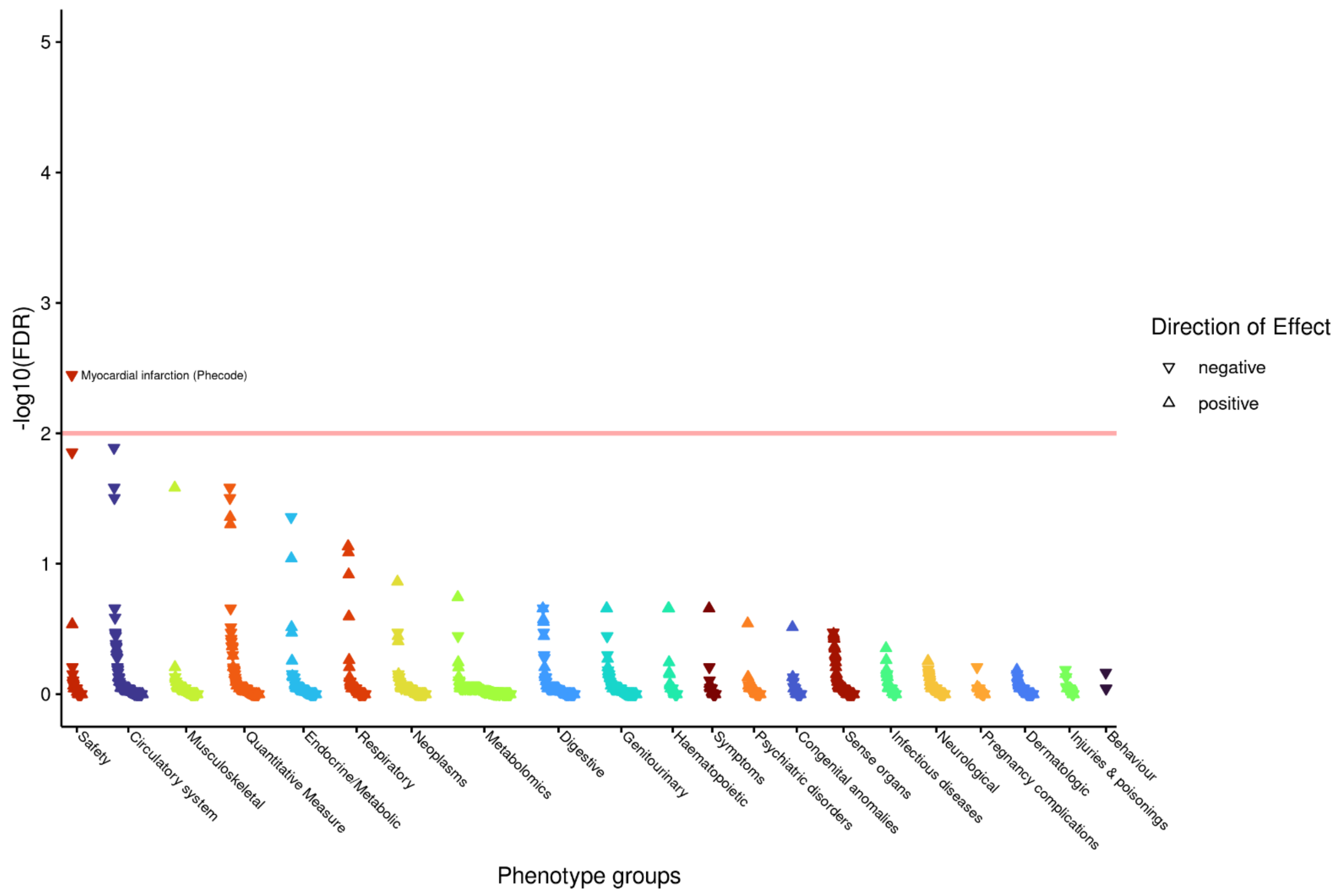

m) **Integrin (INOH)**

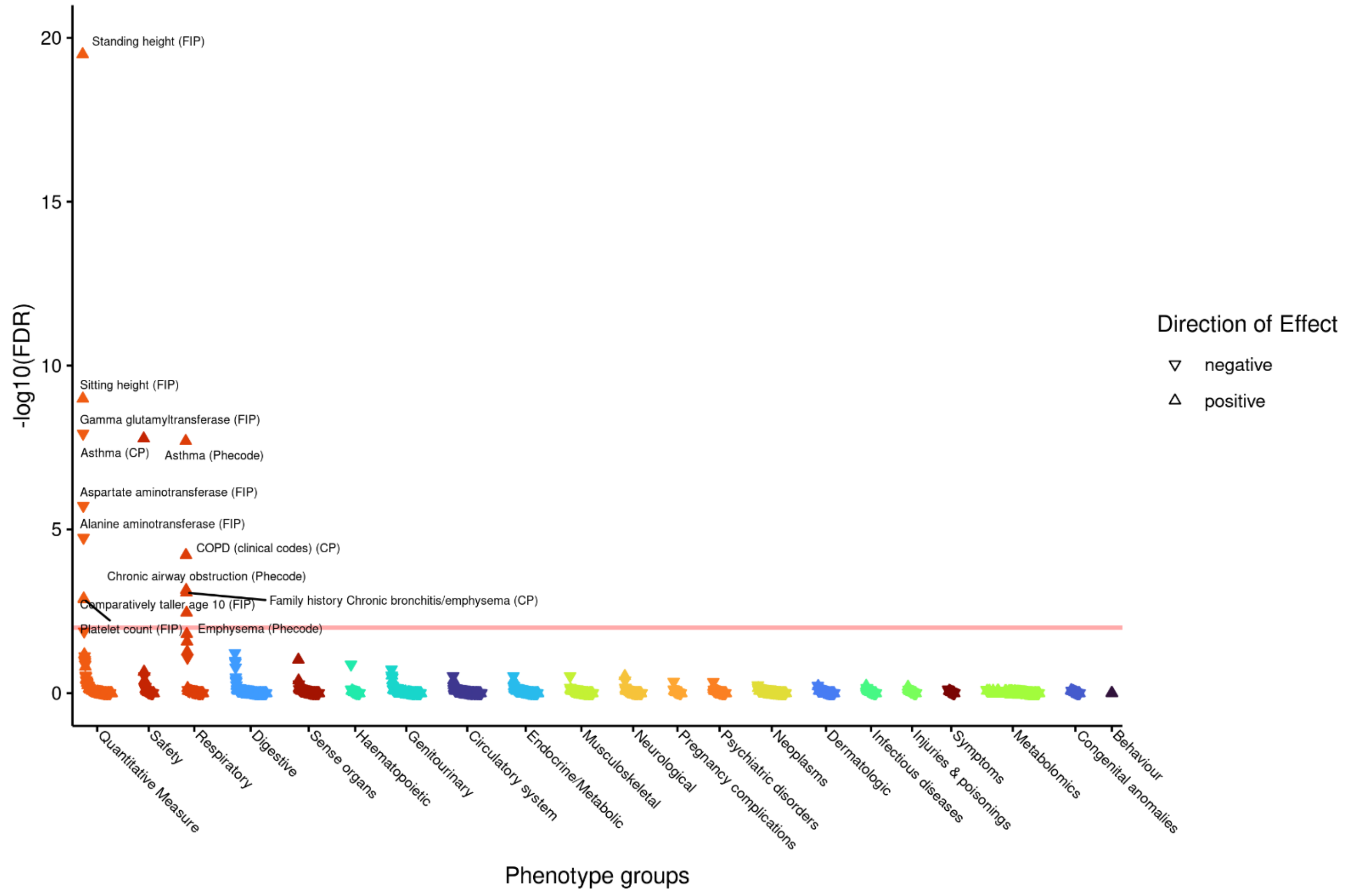

#### n) Molecules associated with elastic fibres (Reactome)

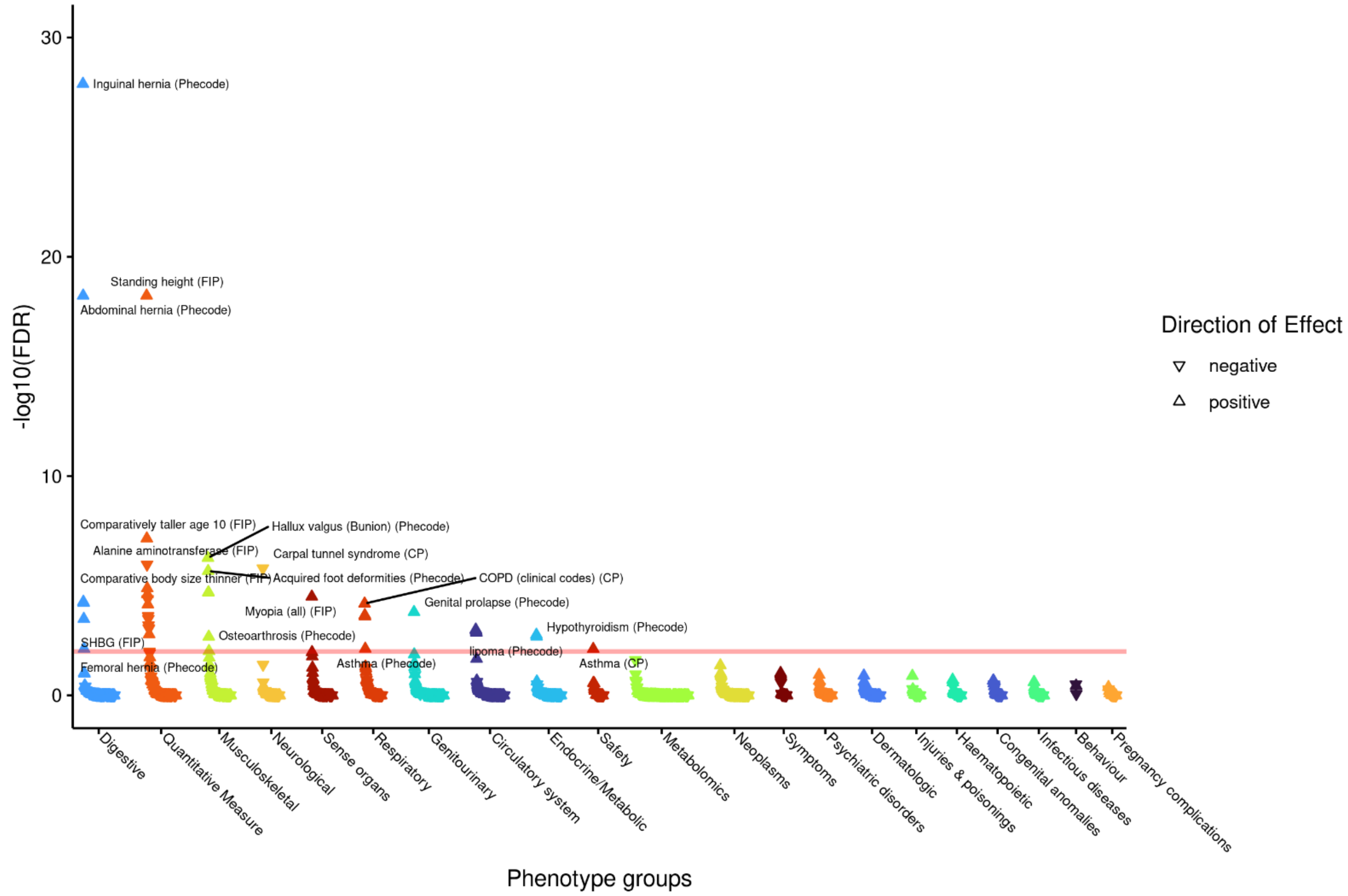

#### o) Osteoblast differentiation (Wikipathways)

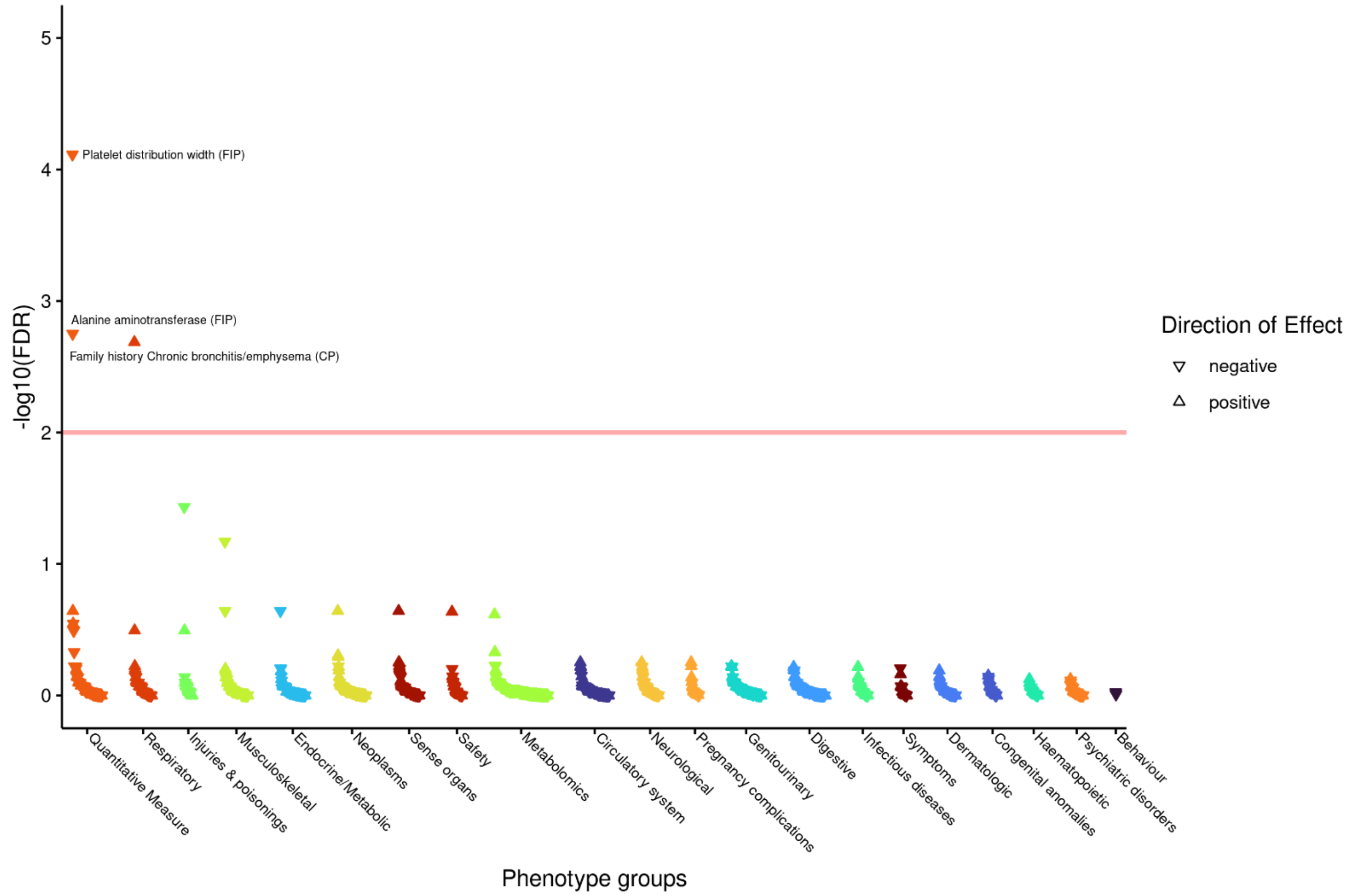

p) **Pathways in cancer Homo sapiens (human) (KEGG)**

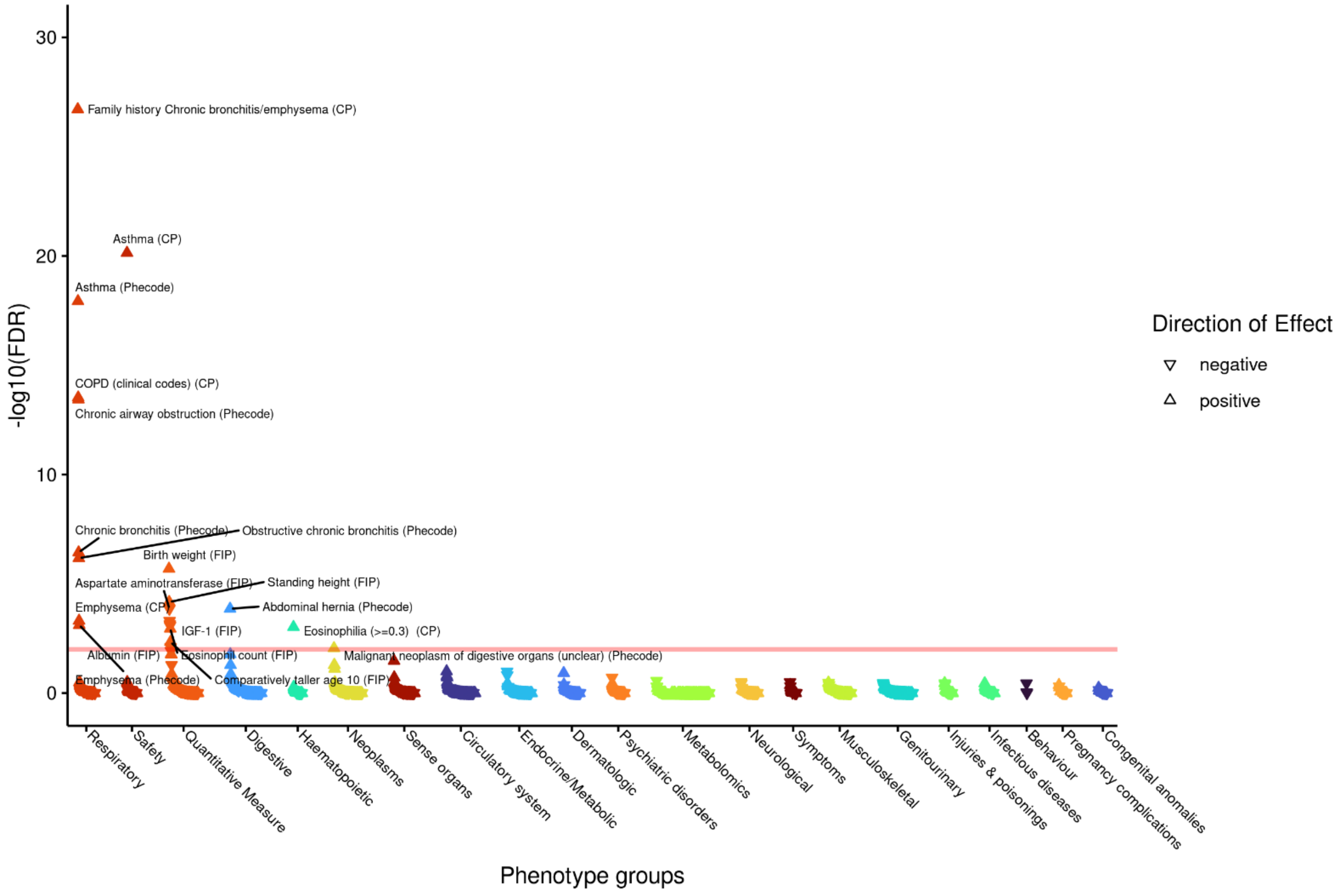

q) **PI3K-Akt signaling pathway (Wikipathways)**

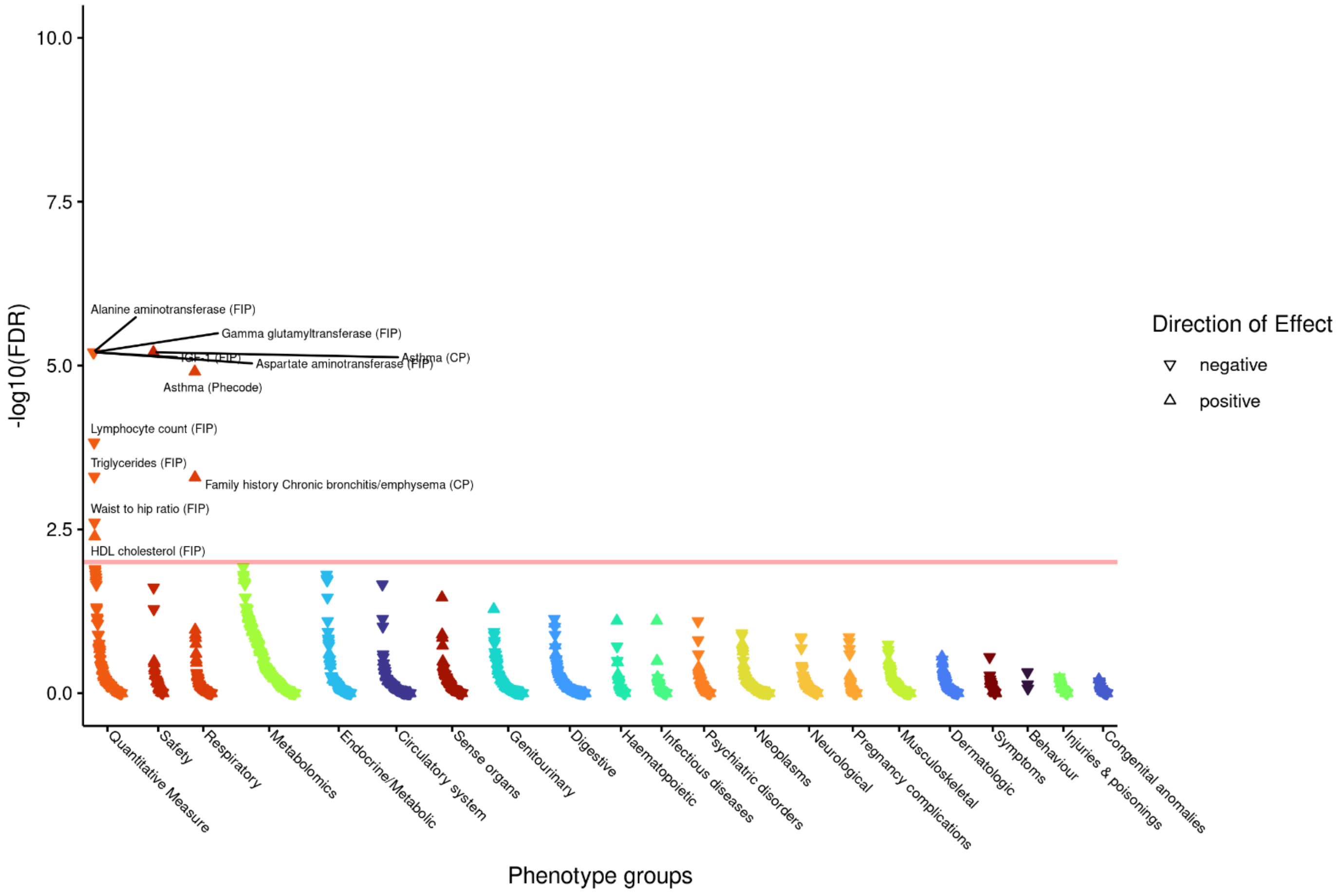

**r) Proteoglycans in cancer Homo sapiens (human) (KEGG)**

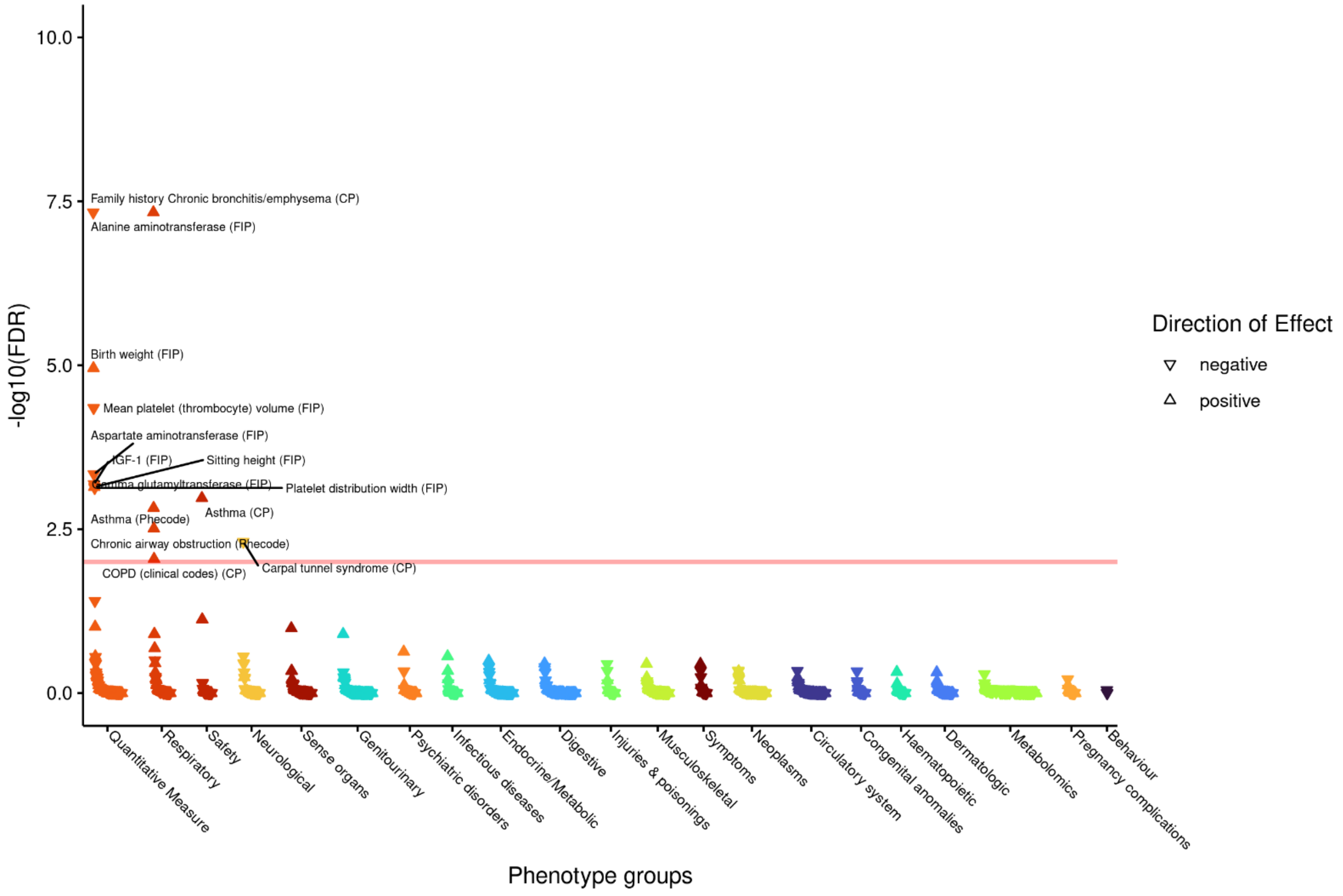

**s) Signaling by Receptor Tyrosine Kinases (Reactome)**

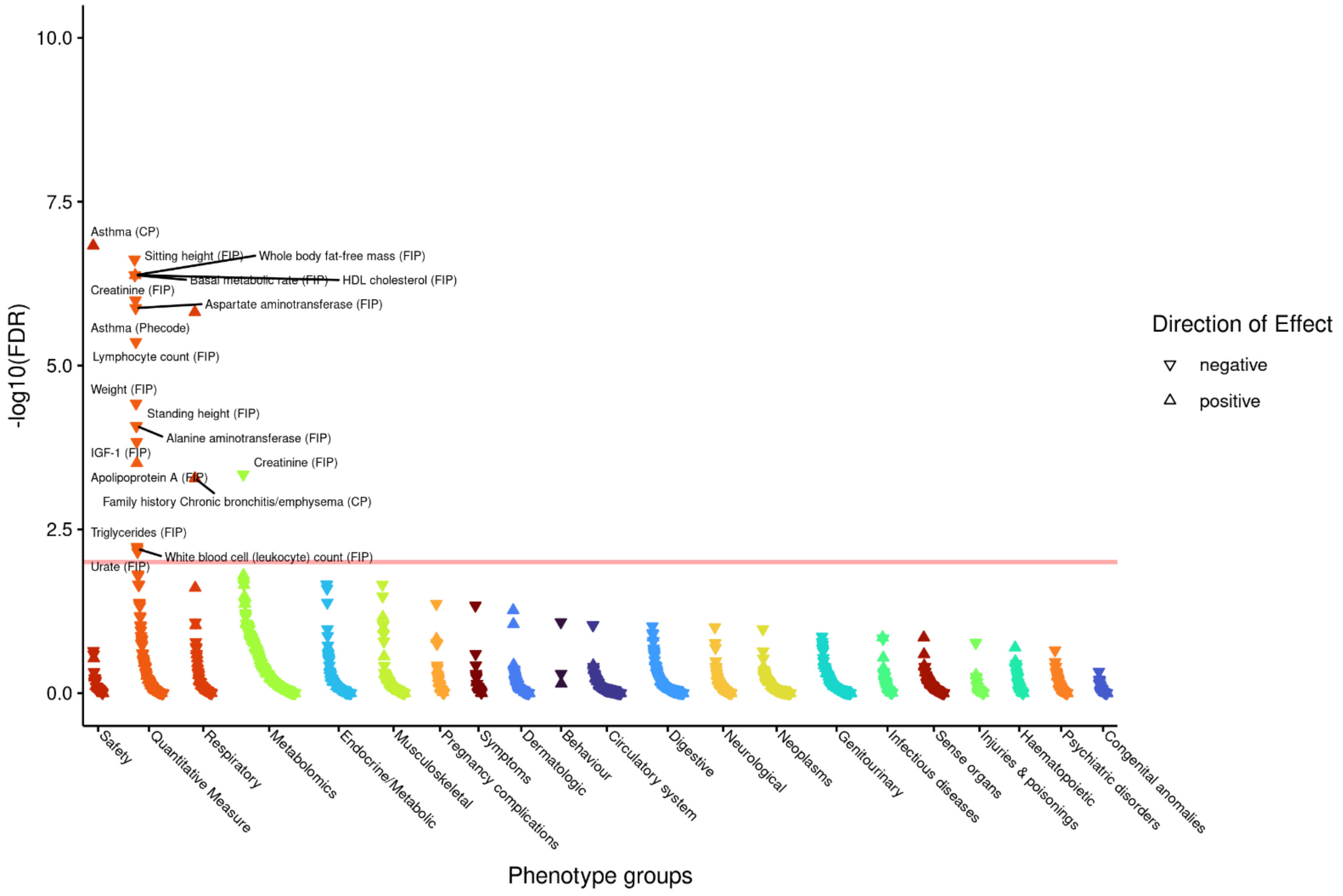

**t) Signalling pathways regulating pluripotency of stem cells Homo sapiens (human) (KEGG)**

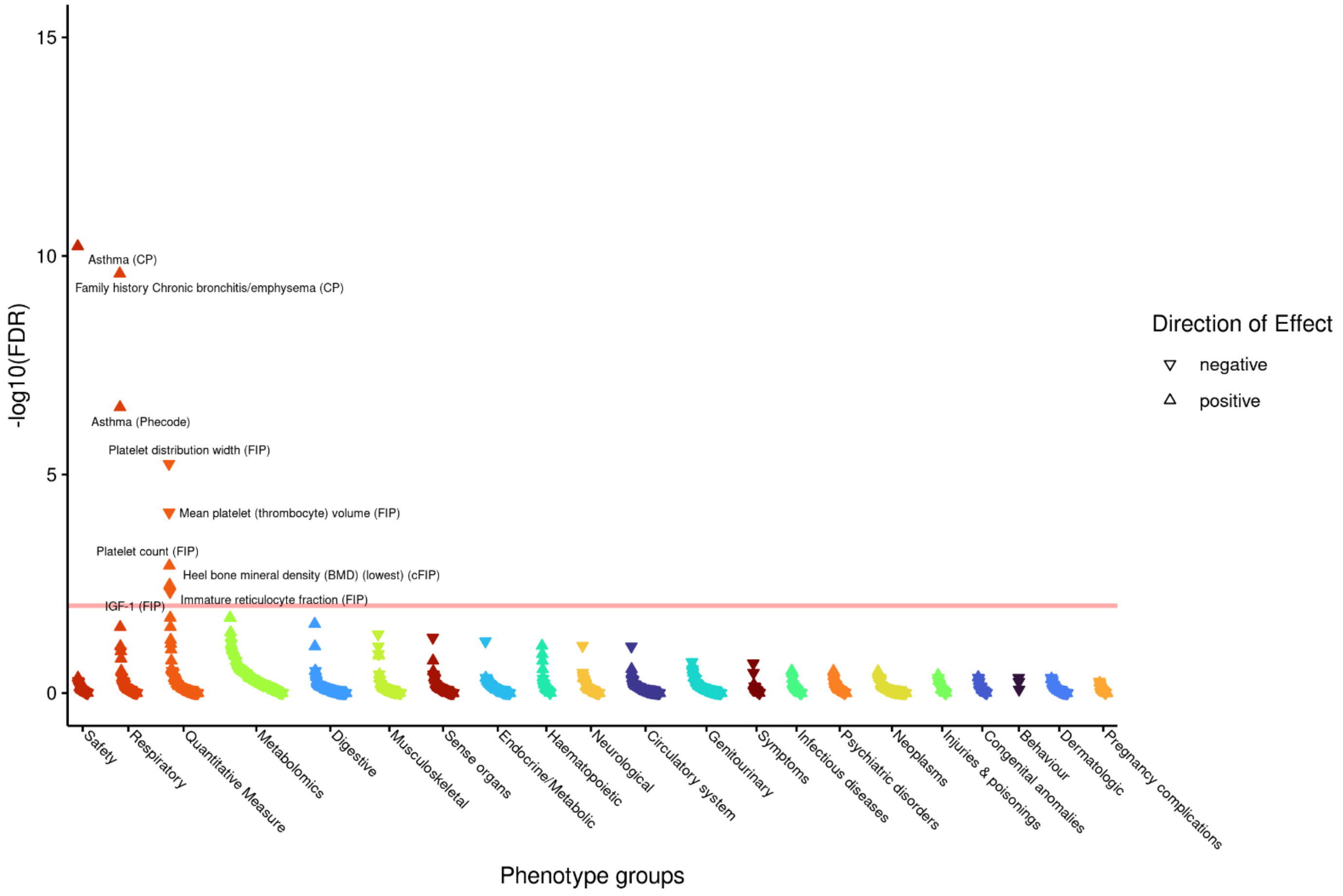

**u) TGF-beta receptor signalling in skeletal dysplasias (Wikipathways)**

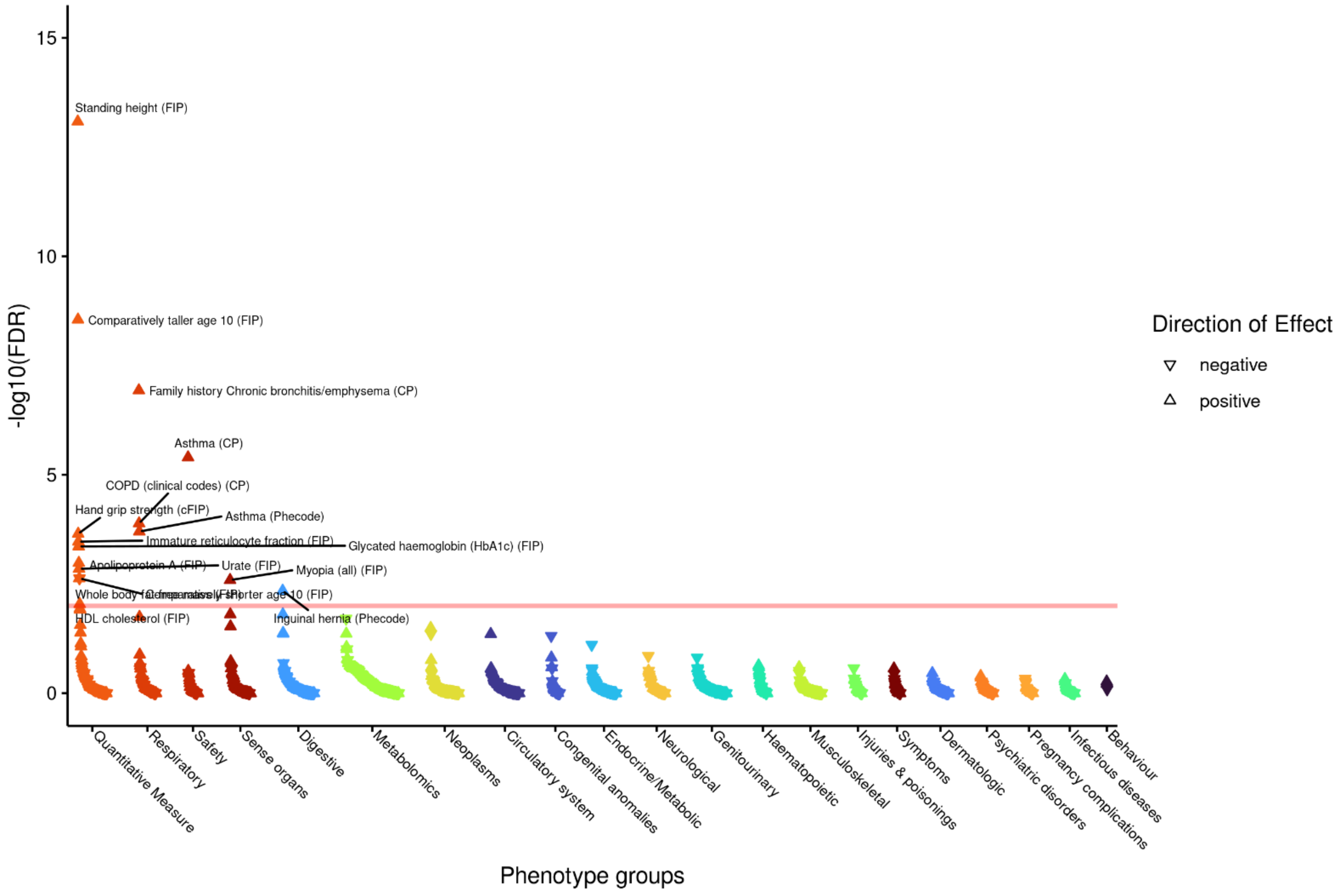

#### v) TGF-beta receptor signalling (Wikipathways)

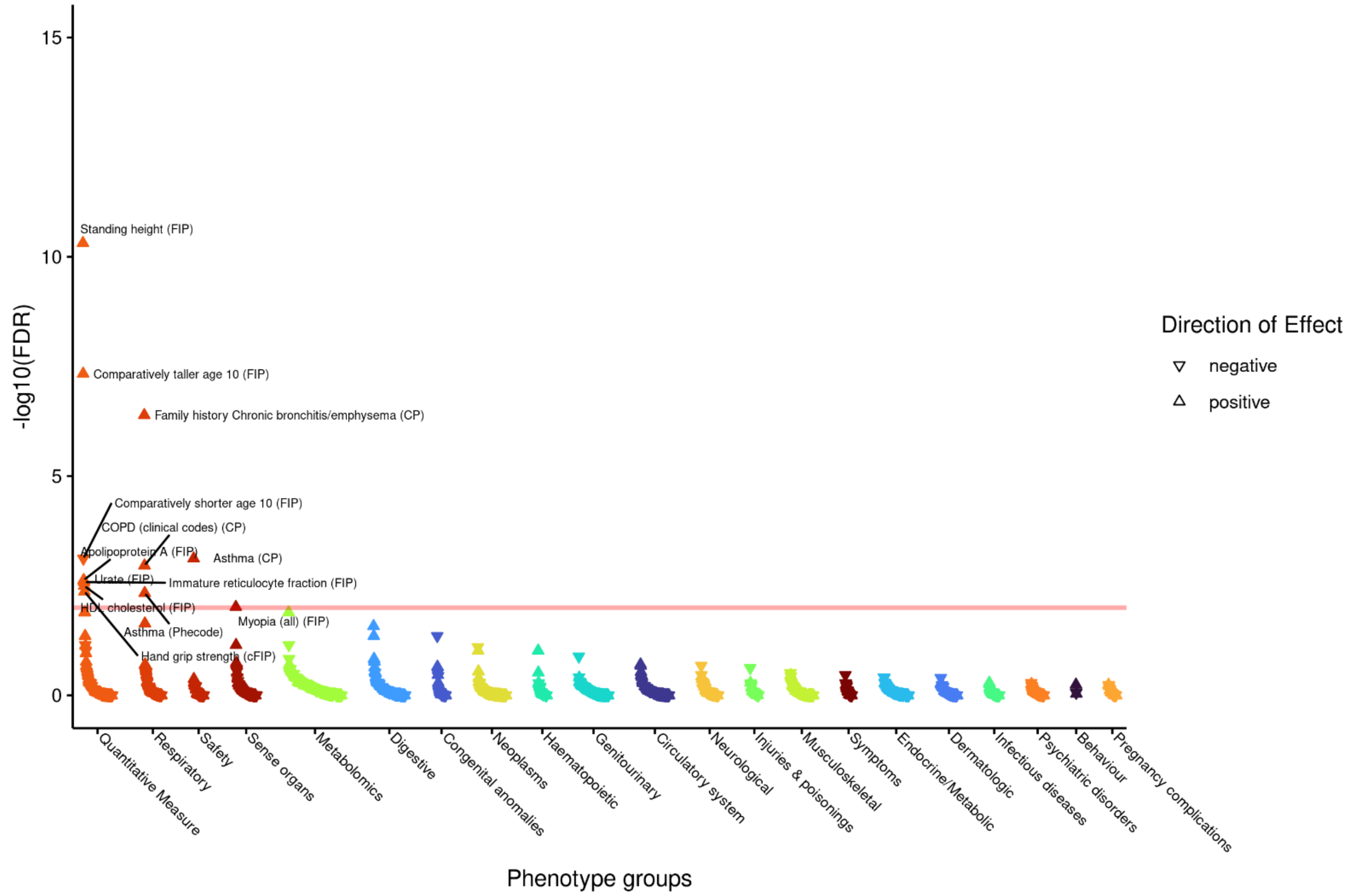

#### w) VEGFA-VEGFR2 Signalling Pathway (Wikipathways)

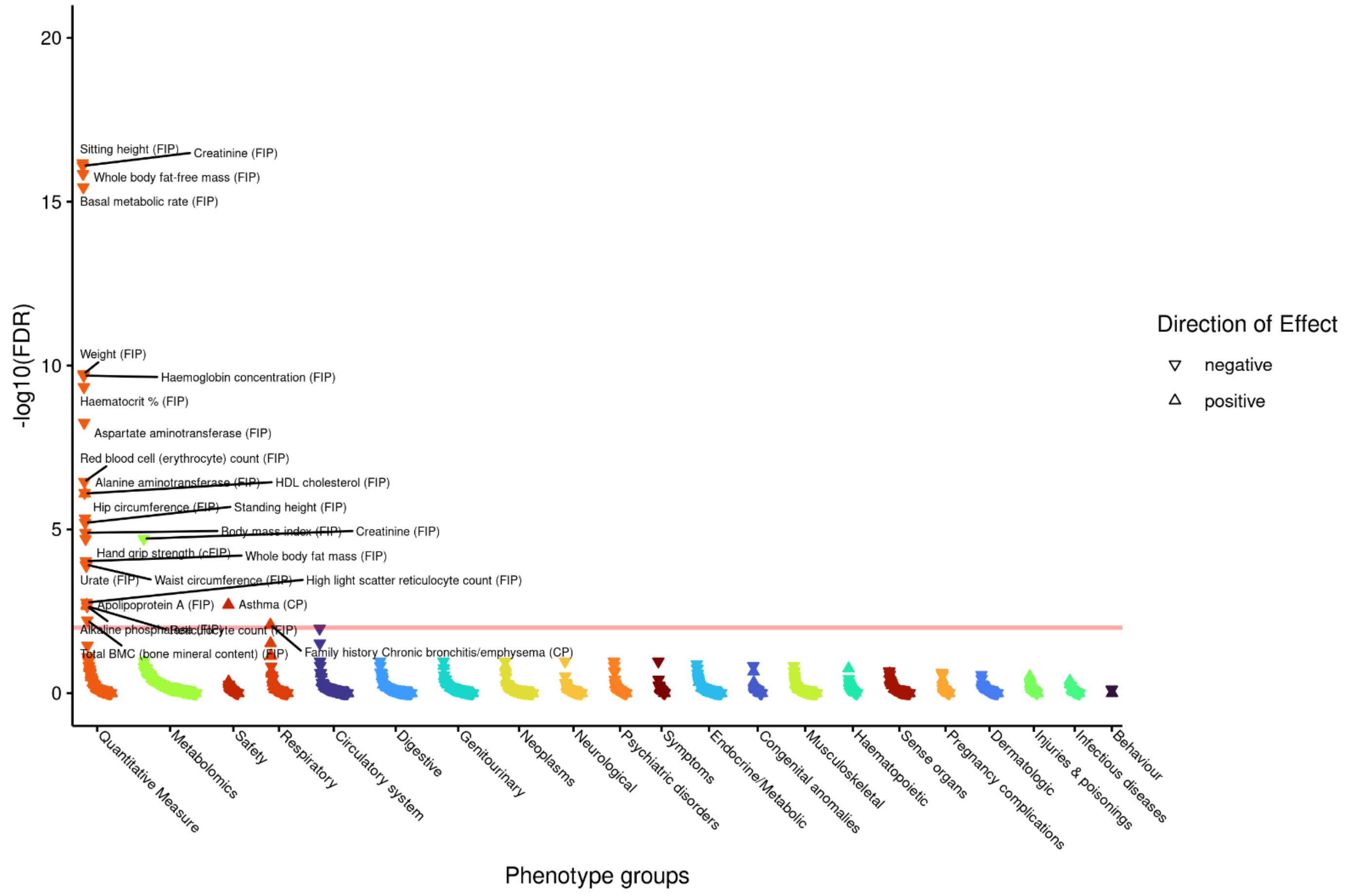

### Supplementary Tables

**Supplementary Table 1: (Excel – sheet "Sample size all cohorts") Sample size all cohorts:**
Sample size in each of 49 cohorts included in the multi-ancestry meta-analysis. The 33 "Super cohorts" groups cohorts into the studies from which multiple cohorts were obtained e.g. 5 ancestries from UK Biobank. Ancestry indicates which of the 5 ancestries each cohort was assigned to: European (EUR), African (AFR), Hispanic/Admixed American (AMR), East Asian (EAS) or South Asian (SAS).

**Supplementary Table 2: Study descriptives**

B58C (B58C-T1DGC, British 1958 Birth Cohort–Type 1 Diabetes Genetics Consortium; B58C-GABRIEL British 1958 Birth Cohort–GABRIEL consortium; B58C-WTCCC, British 1958 Birth Cohort–Wellcome Trust Case Control Consortium); BHS1&2, Busselton Health Study 1 and 2; the CROATIA- Korcula study; the CROATIA-Split study; the CROATIA-Vis study; EPIC -Norfolk,UK population based, European Prospective Investigation into Cancer and Nutrition Cohort; GS:SFHS, Generation Scotland: Scottish Family Health Study; H2000, Finnish Health 2000 survey; KORA F4, Cooperative Health Research in the Region of Augsburg; KORA S3, Cooperative Health Research in the Region of Augsburg; LBC1936, Lothian Birth Cohort 1936; NFBC1966, Northern Finland Birth Cohort of 1966; NFBC1966, Northern Finland Birth Cohort of 1986; NSPHS, Northern Sweden Population Health Study; ORCADES, Orkney Complex Disease Study; PIVUS, Prospective Investigation of the Vasculature in Uppsala Seniors; SHIP, Study of Health in Pomerania; SHIP-TREND; UKHLS; VIKING; YFS, the Young Finish Study. The total size in this table is not exactly equal to the maximum sample size given in the main text, since some studies had different subsets of individuals entering each of the four lung function trait GWAS.

| Study name | N Total | N male | N female | Age range (y) at lung function measurement | Mean age, y (s.d.) | Mean height, cm (s.d.) | Mean $FEV_1$, L (s.d.) | Mean FVC, L (s.d.) | Mean $FEV_1$/FVC (s.d.) | Mean PEF, L/min (s.d.) | N never smokers | N ever smokers | Genotyping Platform | Imputation Panel |
|---|---|---|---|---|---|---|---|---|---|---|---|---|---|---|
| ALHS | 2848 | 1447 | 1401 | 33-98 | 63.09 (10.92) | 169.21 (9.57) | 2.57 (0.82) | 3.50 (1.01) | 0.73 (0.09) | -- | 1884 | 964 | Affymetrix Axiom Genotyping Services | HRC |
| ALSPAC | 4857 | 2444 | 2413 | 7-10 | 8.64 (0.62) | 132.62 (5.83) | 1.70 (0.65) | 1.93 (0.32) | 0.88 (0.07) | 217.00 (42.89) | 4857 | 0 | Illumina HumanHap550 quad chip genotyping platforms by 23andme | HRC |
| ARIC-AA | 2820 | 1044 | 1776 | 44-66 | 53.39 (5.75) | 168.03 (8.94) | 2.57 (0.82) | 3.51 (0.71) | 0.73 (0.09) | 438 (131.4) | 1317 | 1503 | Affymetrix Genome-Wide Human SNP Array 6.0 | TOPmed |
| ARIC-EA | 9275 | 4376 | 4899 | 44-66 | 54.29 (5.69) | 168.72 (9.4) | 2.94 (0.78) | 3.99 (0.98) | 0.73 (0.07) | 483 (142.2) | 3700 | 5575 | Affymetrix Genome-Wide Human SNP Array 6.0 | TOPmed |
| B58C | 5788 | 2862 | 2926 | 44-46 | 45.16 (0.38) | 169.38 (9.29) | 3.30 (0.75) | 4.19 (0.97) | 0.79 (0.08) | 486.21 (136.27) | 1663 | 4125 | Illumina 550k/610k | HRC |
| BHS | 4272 | 1884 | 2388 | 17-97 | 50.80 (17.00) | 168.50 (9.39) | 3.00 (0.96) | 3.90 (1.15) | 0.77 (0.07) | -- | 2414 | 1884 | Illumina 610-Quad (N=1,168 ) & Illumina 660W-Quad (N=3,428 ) | HRC |
| CHS-AA | 635 | 238 | 397 | 65-93 | 73.09 (5.42) | 164.92 (9.22) | 1.75 (0.58) | 2.49 (0.8) | 0.71 (0.12) | -- | 272 | 363 | Illumina HumanOmni1-Quad_v1 BeadChip system | HRC |
| CHS-EA | 3220 | 1259 | 1961 | 65-95 | 72.34 (5.35) | 164.64 (9.35) | 2.11 (0.66) | 3 (0.87) | 0.7 (0.1) | -- | 1541 | 1679 | Illumina 370CNV BeadChip system | HRC |

| Study name | N Total | N male | N female | Age range (y) at lung function measurement | Mean age, y (s.d.) | Mean height, cm (s.d.) | Mean $FEV_1$, L (s.d.) | Mean FVC, L (s.d.) | Mean $FEV_1$/FVC (s.d.) | Mean PEF, L/min (s.d.) | N never smokers | N ever smokers | Genotyping Platform | Imputation Panel |
|---|---|---|---|---|---|---|---|---|---|---|---|---|---|---|
| CKB | 83715 | 36399 | 47316 | 21-96 | 53.40 (10.94) | 158.47 (8.27) | 2.21 (0.69) | 2.65 (0.78) | 0.831 (0.09) | -- | 49029 | 34686 | Custom Affymetrix Axiom arrays | 1KGP phase3 (EAS) |
| CROATIA-Korcula | 2701 | 996 | 1705 | 18-98 | 54.24 (15.80) | 169.3 (9.37) | 2.90 (0.86) | 3.25 (0.971) | 0.859 (0.09) | 362 (140.16) | 1382 | 1319 | Illumina HumanHap370CNV duo chip | HRC |
| CROATIA-Split | 966 | 381 | 585 | 18-85 | 50.26 (14.41) | 172.30 (9.36) | 3.14 (0.86) | 3.75 (1.01) | 0.84 (0.07) | 419.6 (117.96) | 447 | 524 | Illumina HumanHap370CNV quad chip | HRC |
| CROATIA-Vis | 925 | 390 | 535 | 18-91 | 55.94 (15.51) | 167.80 (9.88) | 3.42 (1.21) | 4.41 (1.42) | 0.77 (0.09) | 379.62 (173.46) | 390 | 535 | Illumina Infinium HumanHap300 BeadChip | HRC |
| EPIC-Norfolk | 20290 | 9664 | 11107 | 39-79 | 59.1 (9.27) | 167.1 (9.08) | 2.51 (0.74) | 3.06 (0.93) | 0.83 (0.11) | 364.07 (123.16) | 9532 | 11239 | Affymetrix UKBioBank Axiom | 1000G |
| EXCEED | 1334 | 516 | 818 | 25-71 | 58.28 (8.68) | 166.76 (9.20) | 2.71 (0.69) | 3.59 (0.85) | 0.76 (0.08) | -- | 646 | 688 | Affymetrix | HRC |
| FHS | 7905 | 3656 | 4249 | 19-92 | 52.2 (14.6)) | 168.5 (9.67) | 3.03 (0.94) | 4.02 (1.114) | 0.75 (0.07) | -- | 3546 | 4132 | Affymetrix GeneChip Human Mapping 500K Array Set | TOPmed |
| FinnTwin | 539 | 91 | 448 | 22-61 | 45.21 (15) | 165.20 (9.43) | 2.77 (1.05) | 3.45 (1.21) | 0.78 (0.08) | 400.8 (145.8) | 378 | 161 | Batch1: IlluminaHuman610-Quad v1.0 B, Human670-QuadCustom v1.0 A<br>Batch2: Illumina HumanCoreExome<br>Batch3: Affymetrix FinnGen Axiom array | HRC |
| GS:SFHS | 16048 | 6633 | 10415 | 18-99 | 46.87 (14.6) | 168.4 (9.50) | 2.97 (0.88) | 3.88 (1.00) | 0.76 (0.11) | -- | 8581 | 7467 | Illumina OmniExpress+Exome | 1000G |
| H2000 | 3808 | 1665 | 2153 | 30-97 | 55.73 (15.70) | 167.5 (9.76) | 2.99 (0.96) | 3.77 (1.13) | 0.79 (0.07) | -- | 2080 | 1728 | Illumina HumanHap 610K | 1000G |
| HCHS | 10965 | 4558 | 6407 | 18-76 | 45.92 (13.88) | 162.18 (9.30) | 2.86 (0.8) | 3.56 (0.95) | 0.80 (0.07) | -- | 6648 | 4317 | Illumina Omni2.5M array (plus 150K custom SNP) and Illumina MEGA array | TOPmed |
| HUNT | 16354 | 7583 | 8771 | 19-99 | 49.5 (16.5) | 170.6 (9.17) | 3.15 (1.02) | 4.10 (1.18) | 0.76 (0.10) | 427.8 (147.6) | 6169 | 8801 | Illumina HumanCoreExome | HRC |
| KORA F4 | 1474 | 717 | 757 | 41-84 | 55.08 (9.90) | 169.15 (9.42) | 3.23 (0.85) | 4.19 (1.05) | 0.77 (0.07) | -- | 556 | 918 | Affymetrix Axiom | 1000G |
| KORA S3 | 1147 | 551 | 596 | 28-89 | 50.82 (15.23) | 169.22 (9.32) | 3.34 (0.90) | 4.10 (1.06) | 0.81 (0.08) | -- | 520 | 627 | Illumina Omni 2.5/<br>Illumina Omni Express | 1000G |
| LBC1936 | 1002 | 509 | 493 | 68-71 | 69.55 (0.84) | 166.44 (8.93) | 2.37 (0.69) | 3.05 (0.87) | 0.79 (0.10) | 352.73 (133.7) | 466 | 536 | Illumina 610-Quadv1 | HRC |

| Study name | N Total | N male | N female | Age range (y) at lung function measurement | Mean age, y (s.d.) | Mean height, cm (s.d.) | Mean $FEV_1$, L (s.d.) | Mean FVC, L (s.d.) | Mean $FEV_1$/FVC (s.d.) | Mean PEF, L/min (s.d.) | N never smokers | N ever smokers | Genotyping Platform | Imputation Panel |
|---|---|---|---|---|---|---|---|---|---|---|---|---|---|---|
| MESA-AFA | 908 | 439 | 469 | 48-90 | 65.62 (9.57) | 168.41 (9.0) | 2.19 (0.65) | 2.93 (0.85) | 0.75 (0.09) | -- | 405 | 503 | Affymetrix Human SNP array 6.0 | TOPmed |
| MESA-CAU | 1397 | 707 | 690 | 48-90 | 66.05 (9.71)) | 168.87 (9.61)) | 2.57 (0.76) | 3.52 (0.99) | 0.73 (0.08) | -- | 599 | 798 | Affymetrix Human SNP array 6.0 | TOPmed |
| MESA-HIS | 905 | 478 | 427 | 41-88 | 63.87 (9.76) | 161.88 (9.30) | 2.43 (0.72) | 3.17 (0.90) | 0.77 (0.07) | -- | 487 | 376 | Affymetrix Human SNP array 6.0 | TOPmed |
| NEO | 5705 | 2736 | 2969 | 44-66 | 55.98 (5.95) | 173.70 (9.0) | 3.26 (0.80) | 4.26 (1.02) | 0.77 (0.70) | -- | 1938 | 2843 | Illumina HumanCoreExome-24 BeadChip | TOPmed |
| NFBC1966 | 5078 | 2417 | 2661 | 30-32 | 31.15 (0.35) | 171.24 (9.09) | 3.95 (0.79) | 4.72 (0.99) | 0.84 (0.06) | -- | 2478 | 2600 | Illumina HumanCNV-370DUO Analysis BeadChip | 1000G |
| NFBC1986 | 3210 | 1516 | 1694 | 14-16 | 16.01 (0.37) | 169.34 (8.43) | 3.78 (0.70) | 4.31 (0.85) | 0.88 (0.08) | -- | 2476 | 734 | Illumina Human Omni Express Exome 8v1.2 | 1000G |
| ORCADES | 1821 | 730 | 1091 | 17-92 | 54.00 (15.07) | 167.00 (9.20) | 2.85 (0.83) | 3.55 (0.98) | 0.81 (0.07) | 411.30 (125.5) | 1116 | 705 | Illumina Hap300, Illumina Omni1 & Illumina OmniX | HRC |
| PIVUS | 806 | 395 | 411 | 69-72 | 70.20 (0.176) | 169.09 (9.208) | 2.45 (0.680) | 3.23 (0.869) | 0.764 (0.103) | -- | 393 | 413 | Illumina OmniExpress and Metabochip | 1000G |
| QBB | 5823 | 2541 | 3254 | 18-85 | 39.97 (12.43) | 163.97 (9.36) | 2.82 (0.78) | 3.44 (0.96) | 0.82 (0.08) | -- | 4022 | 1763 | HiSeq X Ten (Illumina, USA) | -- |
| Raine Study | 1213 | 637 | 576 | 13-15 | 14.09 (0.19) | 164.8 (1.9) | 3.08 (0.56) | 3.41 (0.70) | 0.91 (0.07) | 218.04 (51.12) | 1182 | 31 | Illumina Human660W Quad BeadChip | HRC |
| RS1 | 1240 | 526 | 714 | 72-96 | 79.24 (4.6) | 165.76 (9.2) | 2.19 (0.65) | 2.92 (0.83) | 0.76 (0.08) | -- | 411 | 829 | Illumina 500 (+duo) and Illumina Human 610-Quad BeadChips | TOPmed |
| RS2 | 1143 | 532 | 611 | 65-99 | 72.3 (4.96) | 168.65 (9.01) | 2.49 (0.69) | 3.28 (0.87) | 0.76 (0.08) | -- | 382 | 761 | Illumina 500 (+duo) and Illumina Human 610-Quad BeadChips | TOPmed |
| RS3 | 1431 | 624 | 807 | 60-93 | 65.41 (4.27) | 178.1 (9.06) | 2.81 (0.76) | 3.70 (0.97) | 0.76 (0.08) | -- | 474 | 957 | Illumina 500 (+duo) and Illumina Human 610-Quad BeadChips | TOPmed |
| SAPALDIA-Edinburgh | 2773 | 1438 | 1335 | 29-73 | 51.38 (11.0) | 169.80 (9.0) | 3.53 (0.86) | 3.28 (0.81) | 0.75 (0.08) | -- | 1247 | 1526 | Illumina 610k quad | HRC |
| SAPALDIA-Gabriel | 1355 | 656 | 699 | 29-72 | 52.29 (11.02) | 168.96 (9.28) | 3.14 (0.86) | 4.25 (1.04) | 0.73 (0.08) | -- | 582 | 773 | Human610-Quad BeadChip | HRC |
| SHIP | 1759 | 860 | 899 | 20-80 | 47.17 (13.67) | 169.7 (9.13) | 3.28 (0.89) | 3.87 (1.03) | 0.85 (0.06) | 437.58 (125.17) | 818 | 941 | Affymetrix SNP 6.0 | HRC |

| **Study name** | **N Total** | **N male** | **N female** | **Age range (y) at lung function measurement** | **Mean age, y (s.d.)** | **Mean height, cm (s.d.)** | **Mean $FEV_1$, L (s.d.)** | **Mean FVC, L (s.d.)** | **Mean $FEV_1$/FVC (s.d.)** | **Mean PEF, L/min (s.d.)** | **N never smokers** | **N ever smokers** | **Genotyping Platform** | **Imputation Panel** |
|---|---|---|---|---|---|---|---|---|---|---|---|---|---|---|
| SHIP-TREND | 804 | 363 | 441 | 21-81 | 51.24 (13.34) | 169.9 (9.00) | 3.29 (0.87) | 4.14 (1.06) | 0.80 (0.06) | 392.82 (125.91) | 342 | 462 | Illumina Human Omni 2.5 | HRC |
| Twins UK | 4227 | 380 | 3847 | 16-82 | 47.8 (12.46) | 163.69 (7.22) | 2.86 (0.67) | 3.54 (0.75) | 0.85 (0.08) | -- | 1806 | 2419 | Illumina HumanHap300 BeadChip and Illumina HumanHap610 QuadChip | HRC |
| UKB-EUR | 320656 | 142403 | 178253 | 39-72 | 56.45 (8.016) | 168.6 (9.12) | 2.84 (0.75) | 3.74 (0.95) | 0.76 (0.06) | 406.3 (117.2) | 173441 | 147215 | UK BiLEVE array (50K), UK Biobank Axiom Array (450K) | HRC |
| UKB-AFR | 4227 | 1711 | 2516 | 40-70 | 51.64 (7.8) | 166.9 (8.5) | 2.32 (0.65) | 2.98 (0.81) | 0.78 (0.06) | 370.8 (110.5) | 3046 | 1181 | UK Biobank Axiom Array | HRC |
| UKB-AMR | 2798 | 1350 | 1448 | 40-70 | 56.66 (9.32) | 166.2 (9.32) | 2.75 (0.71) | 3.61 (0.91) | 0.76 (0.91) | 406.7 (119.5) | 1500 | 1298 | UK Biobank Axiom Array | HRC |
| UKB-EAS | 1564 | 520 | 1044 | 40-70 | 52.12 (7.77) | 160.7 (8.02) | 2.40 (0.61) | 3.06 (0.77) | 0.78 (0.05) | 370.1 (102.1) | 1184 | 380 | UK Biobank Axiom Array | HRC |
| UKB-SAS | 4270 | 2263 | 2007 | 40-72 | 53.3 (8.32) | 163.5 (9.09) | 2.18 (0.64) | 2.79 (0.79) | 0.78 (0.06) | 357.4 (113.6) | 3272 | 998 | UK Biobank Axiom Array | HRC |
| UKHLS | 7442 | 3290 | 4152 | 16-99 | 53.11 (15.94) | 167.7 (9.45) | 2.84 (0.90) | 3.83 (1.09) | 0.75 (0.09) | -- | 2938 | 4504 | Illumina CoreExome v1.0 | HRC |
| VIKING | 1701 | 672 | 1029 | 18-91 | 50.72 (14.97) | 168 (0.09) | 3.07 (0.81) | 4.02 (0.96) | 0.76 (0.09) | 450.08 (130.14) | 943 | 757 | Illumina OmniExpress Exome | HRC |
| YFS | 419 | 198 | 221 | 30-47 | 38.88 (5.07) | 172.25 (8.90) | 3.73 (0.75) | 4.68 (0.99) | 0.8 (0.06) | -- | 233 | 186 | Illumina 670k custom | 1000G |

**Supplementary Table 3: Imputation QC**

| Study name | Individual call rate filter (applied before imputation) | SNP call rate filter (applied before imputation) | SNP HWE *P* filter (applied before imputation) | SNP MAF filter (applied before imputation) | Other filters | No of SNPs after filtering (before imputation) | Imputation software and version | Reference panel used for imputation | Genotype-phenotype association software |
|---|---|---|---|---|---|---|---|---|---|
| ALHS | ≥ 95% | ≥ 95% | p<1x10-6 (if MAF > 5%) | None | Individuals: Failed Affy QC/Affy Controls, non-Europeans, IBS distance>0.9, sex discrepancies. SNPs: Failed Affy QC/Affy Controls, Failed plate effect check. | 788,635 | Michigan imputation server | HRC (r. 1.1) | Rvtests |
| ALSPAC | ≥ 97% | ≥ 95% | 5.00E-07 | 0.01 | Evidence of cryptic relatedness (IBD > 0.1). | 500,527 | Michigan imputation server | HRC (r. 1.1) | SNPTEST.2.5.2 |
| ARIC AA | ≥ 95% | ≥ 95% | 1.00E-06 | 0.00 | first degree relatives, ancestry outliers, unexpected duplicates, and sex | 831,612 | Michigan Imputation Server (minimac4) | 1000G (phase 3 version 5) | rvtest v. 20190205 |
| ARIC EA | ≥ 95% | ≥ 95% | 1.00E-06 | 0.00 | first degree relatives, ancestry outliers, unexpected duplicates, and sex | 828230 | Michigan Imputation Server (minimac4) | HRC (hrc.r1.1.2016) | rvtest v. 20190205 |
| B58C | None | ≥ 95% | ≥0.0001 (tested on females only for chromosome X) | 0.01 | Consistent allele frequencies across data deposits (*P*≥0.0001 for pairwise comparisons) and for chrX SNPs, consistent allele frequencies between males and females (*P*≥0.0001). | 500,521 (including 11,696 chrX) | MACH 1.0.18 & Minimac 2012-11-16 | 1000 Genomes Phase 1 March 2012 | probABEL 0.1-9e |
| BHS1&2 | ≥ 95% | ≥ 95% | 1.00E-06 | 0.01 | Individuals were removed if they had sex inconsistencies, had heterozygosity >5 s.d. from the mean, were PCA outliers, were 1 individual from a pair of duplicates or had IBD inconsistencies. | 521,307 | Minimac and MACH1 v1.0.18 | b37; 1000 Genomes Phase 1 March 2012 | ProbABEL |
| CKB | ≥ 95% | ≥ 98% | manually exam of SNP clustering plots if p<1e-6 | None | None | 532,529 | shapeit3/impute4 | 1KGP | BOLT-LMM v2.3.4 |
| CROATIA-Korcula | ≥ 97% | ≥ 98% | 1.00E-06 | 0.01 | None | 316,879 | SHAPEIT2, IMPUTE2 | b37; ALL (1000 Genomes Phase 1 integrated release v3, April 2012) | ProbABEL |

| Study name | Individual call rate filter (applied before imputation) | SNP call rate filter (applied before imputation) | SNP HWE *P* filter (applied before imputation) | SNP MAF filter (applied before imputation) | Other filters | No of SNPs after filtering (before imputation) | Imputation software and version | Reference panel used for imputation | Genotype-phenotype association software |
|---|---|---|---|---|---|---|---|---|---|
| CROATIA-Split | ≥ 97% | ≥ 98% | 1.00E-06 | 0.01 | None | 321,727 | SHAPEIT2, IMPUTE2 | b37; ALL (1000 Genomes Phase 1 integrated release v3, April 2012) | ProbABEL |
| CROATIA-Vis | ≥ 97% | ≥ 98% | 1.00E-06 | 0.01 | None | 273,671 | SHAPEIT2, IMPUTE2 | b37; ALL (1000 Genomes Phase 1 integrated release v3, April 2012) | ProbABEL |
| CHS AA | ≥ 95% | ≥ 97% | 1.00E-06 | None | Participants: exluded if genotype discordant with known sex or prior genotyping<br>SNPs: Excluded for >1 duplicate error or Mendelian inconsistency, heterozygote frequency = 0. | 940,567 autosomal | Phasing: Eagle, Pre-phasing: ShapeIT, sever: Michigan server | HRC r1.1 2016 | R, SNPs were excluded for variance on the allele dosage ≤ 0.01 |
| CHS EA | ≥ 95% | ≥ 97% | 1.00E-06 | None | Participants: excluded due to presence at study baseline of coronary heart disease, congestive heart failure, peripheral vascular disease, valvular heart disease, stroke or transient ischemic attack or lack of available DNA or genotype discordant with known sex or prior genotyping<br>SNPs: Excluded for >2 duplicate error or Mendelian inconsistencies, heterozygote frequency = 0, SNP not found in HapMap. | 306,655 autosomal SNPs | Phasing: Eagle, Pre-phasing: ShapeIT, sever: Michigan server | HRC r1.1 2016 | R, SNPs were excluded for variance on the allele dosage ≤ 0.01 |
| EPIC-Norfolk * | None | ≥ 95% | 1.00E-08 | Per-plate basis | Monomorphic SNPs; chr 23-26; INDELS; monomorphic; call rate<95%; chr-pos-allels duplicates; delta-AF > 0.2; delta_AF>0.1 if MAF<0.01. Oxford QC:) exclude SNPs if not in HRC ref (no INDEL in HRC ref); 2) exclude if don't match on chr-pos-allele; 3) strand check and flip; 4) exclude if delta-AF>0.2; 5) exclude A/T and G/C SNPs with MAF>0.4 in ref; 6) exclude if chr-pos duplicates | 708,715 | SHAPEIT v2.r790, Oxford | HRC v1.0, 1000 Genomes p3 | BOLT-LMM v2.2 |

| Study name | Individual call rate filter (applied before imputation) | SNP call rate filter (applied before imputation) | SNP HWE *P* filter (applied before imputation) | SNP MAF filter (applied before imputation) | Other filters | No of SNPs after filtering (before imputation) | Imputation software and version | Reference panel used for imputation | Genotype-phenotype association software |
|---|---|---|---|---|---|---|---|---|---|
| EXCEED | ≥ 97% | ≥ 95% | 1.00E-06 | 0.01 | None | 5,216 | Michigan Imputation Server – Minimac3 v1.0.4 | HRC panel r1.1 2016 (EUR) | SAIGE v0.39.2 |
| FHS | ≥ 97% | ≥ 95% | 1.00E-06 | 0.01 | Remove if Mendelian errors>=1000, or at locations that did not map to GRCh37 | 412,049 | Michigan Imputation Server - Minimac3 | HRC panel release 1.1 | lmekin(), R |
| FinnTwin | ≥ 98% (BATCH1); ≥95% (BATCH2); ≥95% (BATCH3) | ≥ 97.5% (BATCH1); ≥97.5% (BATCH2); ≥95% (BATCH3) | 1.00E-06 | 0.01 | Exclusion of individuals with high heterozygosity rate (BATCH1: F< -0.03 or >0.05; BATCH2: F< -0.03 or >0.05; BATCH3: F ±4SD from the mean), mismatched sex, duplicates and ancestry outliers. | 497,956 (BATCH1); 251,589 (BATCH2); 423,229 (BATCH3) | Michigan Imputation Server - Eagle2 (SHAPEIT2 for chrX), | HRC (release 1.1) | RVTESTS v2.1.0 |
| GS:SFHS* | ≥ 97% | ≥ 98% | 1.00E-06 | 0.01 | Genetic ancestry outliers; monomorphic SNPs; high heterozygosity | 602,451 | SHAPEIT2 v2.r837, Sanger | HRC panel v1.1, European | REGSCAN |
| H2000 | ≥ 95% | ≥ 95% (≥ 99% for SNPs with MAF < 0.05) | 1.00E-06 | 0.01 | None | 553,722 | IMPUTE version 2.2.2 | 1,000 Genomes haplotypes -- Phase I integrated variant set release (v3) in NCBI build 37 (hg19) coordinates | SNPTest |
| HUNT | ≥ 99% | ≥ 99% | 1.00E-04 | >0 | Estimated contamination >2.5% (BAF regress), gonosomal constellations other than XX and XY, gender discordance, probe sequences mismatch, cluster separation < 0.3, Gentrain score < 0.15, assay with higher call rate genotyped the same variant, frequency differences > 15% between arrays dataset, monomorphic in one and had MAF > 1% in another array data set. | 499,377 | Minimac3 (v2.0.1) | HRC v1.1 reference European and HUNT WGS | BOLT-LMM |

| Study name | Individual call rate filter (applied before imputation) | SNP call rate filter (applied before imputation) | SNP HWE *P* filter (applied before imputation) | SNP MAF filter (applied before imputation) | Other filters | No of SNPs after filtering (before imputation) | Imputation software and version | Reference panel used for imputation | Genotype-phenotype association software |
|---|---|---|---|---|---|---|---|---|---|
| KORA F4* | ≥ 97% | ≥ 98% | 1.00E-06 | 0.01 | -mismatch of phenotypic and genetic gender<br>- 5s.d. from mean heterozygosity rate<br>- check for European ancestry<br>- check for population outlier | 523,260 (chr 1-26)<br>508,532 (chr 1-22)<br>14,096(chrX-nonPAR)<br>444(chrX-PAR1)<br>58(chrX-PAR2) | SHAPEIT v2, IMPUTE v2.3.0 | 1000g phase1 all (ALL_1000G_phase1integrated_v3_impute_mac1) | SNPTEST v2.4.1 |
| KORA S3* | ≥ 97% | ≥ 98% | 1.00E-06 | 0.01 | person wise: -mismatch of phenotypic and genetic gender<br>- 5s.d. from mean heterozygosity rate<br>- check for European ancestry<br>- check for population outlier<br>SNP wise: only SNPs that were genotyped with good quality on both chips | 600641 (chr 1-26)<br>588307 (chr 1-22)<br>14625 (chrX-nonPAR) | SHAPEIT v2, IMPUTE v2.3.0 | 1000g phase1 all (ALL_1000G_phase1integrated_v3_impute_mac1) | SNPTEST v2.4.1 |
| LBC1936 | ≥ 95% | ≥ 98% | ≥0.001 | 0.01 | None | 549,692 | Michigan Imputation Server | HRC v1.1, European | SNPTEST |
| MESA AFA | ≥ 95% | ≥ 95% | None | None | Monomophic variants, excess heterozygosity > 53%, genetic ancestry, first degree relatives. | 897,981 | Michigan Imputation Server | TOPMed Freeze 5b | SNPTEST v2.5 Plink v2 chrX |
| MESA CAU | ≥ 95% | ≥ 95% | None | None | Monomophic variants, excess heterozygosity > 53%, genetic ancestry, first degree relatives. | 897,981 | Michigan Imputation Server | HRC Release 1 | SNPTEST v2.5 Plink v2 chrX |
| MESA HIS | ≥ 95% | ≥ 95% | None | None | Monomophic variants, excess heterozygosity > 53%, genetic ancestry, first degree relatives. | 897,981 | Michigan Imputation Server | TOPMed Freeze 5b | SNPTEST v2.5 Plink v2 chrX |
| NEO | ≥ 98% | ≥ 98% | 1.00E-06 | 0.00 | Exclusion of first/second degree relatives* (measure, threshold if relevant)=pi^>0.25; Exclusion of genetic ancestry outliers= +/-3.5SD | 361,046 | HRC (r1.1) | HRC ALL | SNPTEST |
| NFBC1966* | ≥ 95% | ≥ 95% | 1.00E-04 | 0.01 | Genetic ancestry outliers; monomorphic SNPs; high heterozygosity; Gender mismatch; 0 genetic sex; high heterozygosity; high relatedness | 364,535 | Eagle v2.3, Michigan | HRC r1.1 2016, European | rvtests |

| Study name | Individual call rate filter (applied before imputation) | SNP call rate filter (applied before imputation) | SNP HWE *P* filter (applied before imputation) | SNP MAF filter (applied before imputation) | Other filters | No of SNPs after filtering (before imputation) | Imputation software and version | Reference panel used for imputation | Genotype-phenotype association software |
|---|---|---|---|---|---|---|---|---|---|
| NFBC1986* | ≥ 99% | ≥ 99% | 1.00E-04 | 0.01 | Genetic ancestry outliers; monomorphic SNPs; high heterozygosity; Gender mismatch; high heterozygosity; high relatedness | 889,119 | Eagle v2.3, Michigan | HRC r1.1 2016, European | rvtests |
| NSPHS | ≥ 99% | ≥ 95% | 3.2E-08 (Infinum) & 1.4E-08 (OmniExpress) | 0.01 | FDR level of heterozygosity 0.01 | 306,086 (Infinum) & 631503 (OmniExpress) | Impute2 (v 2.2.2) | hg19, 1000 Genomes | ProbABEL |
| ORCADES | ≥ 98% | ≥ 97% | 1.00E-06 | 0.01 (Hap300) & monomorphic (Omni & OmniX) | Subject Heterozygosity FDR<1% | 287,208 (Hap300), 843723 (Omni) & 654651 (OmniX) | shapeit.v2.r644.+impute_v2.2.2_x86_64_static/impute2 | 1000G Phase I Integrated Release Version 3 Haplotypes (2010-11 data freeze, 2012-03-14 haplotypes). | probABEL v. 0.4.3 |
| PIVUS* | ≥ 95% | ≥ 95% (≥ 99% if MAF<0.05) | 1.00E-06 | 0.01 | Genetic ancestry outliers; monomorphic SNPs; >3SD from mean for heterozygosity, pi-hat>0.125, gender discordance | 738,583 | SHAPEITv2, Oxford | HRC v1.1, all | SNPTEST v2.5 |
| Raine Study | ≥ 97% | ≥ 95% | 1.00E-06 | 0.01 | A/T and C/G ambiguous SNPs | 514,142 | Michigan Imputation Server – ShapeIT (CEU, Admixed) | HRC r1.1 2016 | ProbAbel v.0.4.1 |
| Rotterdam Study | ≥ 98% | ≥ 98% | 1.00E-06 | 0.01 | None | 512,849 (RSI), 537,405 (RSII) | Michigan Imputation Server | HRC v1.1 | rvtest |
| SAPALDIA | ≥ 95% | ≥ 95% | 1.00E-06 | 0.01 | None | 545,131 | Mach 1.0.16.a, minimac-omp RELEASE STAMP 2012-05-29 (autosomes) & MiniMac RELEASE STAMP 2012-11-16 (chr X) | build37, 1000 Genomes | probABEL |
| SHIP | ≥ 92% | ≥ 95% | 1.00E-04 | None | Genetic ancestry outliers; gender mismatch; pi-hat>0.25; monomorphic SNPS | 760,787 | Eagle v2.3, Michigan | HRC v1.1 reference, European | Rvtests |
| SHIP-TREND | ≥ 94% | ≥ 95% | 1.00E-04 | None | Genetic ancestry outliers; gender mismatch; pi-hat>0.25; monomorphic SNPS | 1,691,610 | Eagle v2.3, Michigan | HRC v1.1 reference, European | Rvtests |

| Study name | Individual call rate filter (applied before imputation) | SNP call rate filter (applied before imputation) | SNP HWE *P* filter (applied before imputation) | SNP MAF filter (applied before imputation) | Other filters | No of SNPs after filtering (before imputation) | Imputation software and version | Reference panel used for imputation | Genotype-phenotype association software |
|---|---|---|---|---|---|---|---|---|---|
| TwinsUK | ≥98% | ≥98% | 1.00E-06 | None | pi^>0.25. Ancestry outliers +/-3.5SD | 275,235 | Michigan Imputation Server | HRC r 1.1 | GEMMA v0.98 |
| UK Biobank | ≥ 95% | ≥ 90% | 1.00E-12 | 0.0001 | Batch effects, plate effects, gender discordance, heterozygosity outliers | 670,739 | SHAPEITv3, IMPUTEv4, Oxford | HRC v1.1 reference, European + merged UK10K and 1000 Genomes phase 3 | BOLT-LMM |
| UKHLS* | ≥ 98% | ≥ 98% | 1.00E-04 | None | Genetic ancestry, monomorphic SNPs, heterozygosity 3sd <>mean - visualised at 2 different MAF bins (≥1% and <1%); PI_HAT 0.2; Cluster separation score <0.4; sex check, ethnicity duplicates, withdrawn consent. Pre-imputation variants excluded that were: monomorphic, indels, differed to HRC in terms of strand, alleles, allele frequency (>0.2), A/T & G/C SNPs if MAF >0.4 and not in reference panel. | 357,230 | Autosomes: Eagle v2.3; ChrX: Shapeit v2.r790, Michigan | Autosomes: HRC r1.1 2016; ChrX: HRC r1.1 2017, European | SNPTEST v2.5 |
| VIKING* | ≥ 97% | ≥ 98% | 1.00E-06 | MAF>0.01 for OMNI markers; MAF>0.0001 for Exome Chip markers | Genetic ancestry outliers; monomorphic SNPs; Duplicates and siblings | 668,762 | shapeit2r837 + duohmm; PBWT Sanger | HRC v1.1, European | REGSCAN 0.4 |
| YFS* | ≥ 95% | ≥ 95% | 1.00E-06 | 0.01 | heterozygosity, relatedness | 546,674 | SHAPEIT v1 and IMPUTE v2.2.2 | 1000 Genomes Phase 1, release v3, March 2012 haplotypes | SNPTEST v.2.4.1 |

*indicates 10 studies where association results from Shrine et al. (2019) were used as opposed to an updated imputation and association testing.

**Supplementary Table 4: UK Biobank ancestry composition**

Numbers of samples assigned to each ancestry group using different thresholds for the ADMIXTURE dominant ancestry group. The total number of samples varies as an increasing number of samples are assigned as missing as the threshold becomes more stringent. EUR for European, AFR for African, EAS for East Asian, SAS for South Asian and AMR for American/Hispanic.

| | | EUR | AFR | EAS | SAS | AMR | Total |
|---|---|---|---|---|---|---|---|
| ADMIXTURE dominant ancestry threshold used | 70% | 133,982 | 7,938 | 2,536 | 8,810 | 5,010 | 158,276 |
| | 75% | 132,923 | 7,687 | 2,487 | 6,975 | 4,308 | 154,380 |
| | 80% | 131,643 | 7,341 | 2,417 | 4,569 | 3,851 | 149,821 |
| | 90% | 104,474 | 5,948 | 2,248 | 2,057 | 3,121 | 117,848 |
| Final sample numbers using 75% threshold, after quality control and with lung function data | | 320,656* | 4,227 | 1,564 | 4,270 | 2,798 | 333,515 |

*321,047 European samples as defined previously were not included in the ADMIXTURE run but were taken forward as our European samples for the GWAS. We did not add the potential extra European samples identified by the ADMIXTURE analysis to the GWAS.

**Supplementary Table 5: (Excel sheet "LD score intercept")**
LD score regression intercepts from GWAS in each of 49 contributing studies.

**Supplementary Table 6: (Excel – sheet "Reported signals") Summary of all reported signals**
Results from the signal selection of the trans-ethnic meta-analysis for all lung function signals for each of the lung function quantitative traits FEV1, FVC, $FEV_1$/FVC and PEF. The definition of each column in the table is given as below: *Reported*: indicating if the signal is novel (FALSE) or previously reported (TRUE); *trait*: the lung function quantitative traits; *sentinel, chr, pos, effect, other, eaf, MAF*: the sentinel variant of the signal and the corresponding position on the chromosome (GRCh37), effect/other allele, effect allele frequency, minor allele frequency; *Zscore/cZscore, P/cP*: the Z score (P value) or the conditional Z score (conditional P value) for the signal; *Studies, Direction*: the list of studies contributing to the signal selection in the corresponding round and their corresponding estimated effect direction (+:positive effect; -: negative effect; ?:missing data); *beta*: effect size; *se*: standard error; *P*: P value; EUR: European; AFR: African; EAS: East Asian; SAS: South Asian; AMR: American/Hispanic.

**Supplementary Table 7: (Excel – sheet "smoking look up") Summary of all reported signals**
Look up of lung function signals in GWAS and Sequencing Consortium of Alcohol and Nicotine use (GSCAN) consortium[79] for association with smoking behaviour traits Cigarettes per day (quantitative), Smoking initiation and Smoking cessation (binary). Column Z "BETA.aln" is the smoking effect aligned to the lung function decreasing allele. Columns AA to AN give the results for a look up of the effect is UK Biobank smokers and non-smokers separately.

**Supplementary Table 8: (Excel – sheet "MRMEGA") MR-MEGA ancestry-adjusted meta-regression results**
MR-MEGA results for meta-regression including 4 axes of genetic ancestry as covariates. 960 variants represented in at least 7 cohorts, of which 554 (57.71%) attained at least nominal evidence of association ($p < 0.05$) in two or more ancestry-specific meta-analysis. Heterogeneity in allelic effects at an association signal can be partitioned into two components: the first captures heterogeneity that is correlated with ancestry (column Pvalue_ancestry_het) and residual heterogeneity due to differences in study design (column Pvalue_residual_het). We observed 93 (9.69%) signals with nominal evidence ($p < 0.05$) of residual heterogeneity compared to that expected by chance (binominal test p = 2.97x10-9). In contrast, there was nominal evidence of ancestry-correlated heterogeneity at 109 (11.35%) signals (binomial test $p = 5.17x10^{-15}$), suggesting that heterogeneity in allelic effect sizes between GWAS are more likely due to factors related to ancestry than to study design. The definition of each column in the table is given as below: *trait*: the lung function quantitative traits; *sentinel, chr, pos, effect, other, eaf*: the sentinel variant of the signal and the corresponding position on the chromosome (GRCh37), effect/other allele, effect allele frequency; *Studies, Direction*: the list of studies contributing to the heterogeneity analysis and their corresponding estimated effect direction (+:positive effect; -: negative effect; ?:missing data); *beta_0* (*se_0*): the intercept (standard error) estimated in the meta-regression; *beta_1 (se_1), beta_2 (se_2), beta_3 (se_3), beta_4 (se_4)*: effect size (standard error) of the first to the fourth PC of meta-regression; *chisq_association*: chi square value of the association; *ndf_association*: degree of freedom of the association; *P-value_association*: p-value of the association; *chisq_ancestry_het*: chi square value of the heterogeneity due to different ancestry; *ndf_ancestry_het*: degree of freedom of the heterogeneity due to different ancestry; *P-value_ancestry_het*: p-value of the heterogeneity due to different ancestry; *chisq_residual_het*: chi square value of the residual heterogeneity; *ndf_residual_het*: degree of

freedom of the residual heterogeneity; P-value_residual_het: p-value of the residual heterogeneity; *lnBF*: log of Bayes factor.

**Supplementary Table 9: Variants showing a significantly different* effect in adults and children**

| Gene | Rsid | variant | trait | effect allele | Children | | Adults | | P.diff |
|---|---|---|---|---|---|---|---|---|---|
| | | | | | beta | se | beta | se | |
| MECOM | rs6806825 | 3_168789705_C_T | FVC | T | 0.0106 | 0.0046 | -0.0094 | 0.0013 | $2.86 \times 10^{-5}$ |
| CYTL1 | rs11722554 | 4_5016883_A_G | PEF | A | 0.0624 | 0.0327 | -0.0921 | 0.0087 | $4.97 \times 10^{-6}$ |
| CCDC91 | rs7977418 | 12_28588242_C_T | FVC | T | -0.0002 | 0.0043 | 0.0208 | 0.0013 | $2.94 \times 10^{-6}$ |
| MAPT, MGC57346-CRHR1 | rs11079718 | 17_43839951_A_T | $FEV_1$ | A | 0.0026 | 0.0044 | 0.0216 | 0.0013 | $3.45 \times 10^{-5}$ |

*P < $5.14 \times 10^{-5}$ (5% Bonferroni corrected for 972 signals tested)

**Supplementary Table 10: (Excel – sheet "Smoking interaction")**
Smoking interaction effect at lung function signals tested across ever and never smokers in UK Biobank.

**Supplementary Table 11: (Excel – sheet "Rare disease genes") Genes near our lung function signals associated with rare mendelian respiratory diseases**
199 genes associated with a rare mendelian respiratory disease were implicated.

**Supplementary Table 12: (Excel – sheet "Mouse knockout genes") Mouse ortholog genes near our lung function signals associated with a respiratory disease**
59 genes were implicated

**Supplementary Table 13: Enriched annotations in lung function associated loci used to calculate annotation-informed credible sets with fGWAS.**

| Phenotype | Enriched annotation | Log fold enrichment (95% C.I.) | Enrichment P |
|---|---|---|---:|
| $FEV_1$ | Matrix fibroblast 2 open chromatin | 1.43 (0.9, 1.9) | 0.0025 |
| | Exonic variant | 1.7 (0.69, 2.43) | 0.0031 |
| | UTR3' variant | 1.85 (0.75, 2.59) | 0.0041 |
| | FOXF1 TFBS* in lung | 3.39 (1.34, 4.61) | 0.0066 |
| | SOX2 TFBS in lung | 3.54 (1.2, 4.8) | 0.0115 |
| | Myofibroblast open chromatin | 1.1 (0.52, 1.59) | 0.0140 |
| | CEBPZ TFBS in lung | 3.96 (1.4, 5.17) | 0.0195 |
| FVC | Exonic variant | 2.11 (1.37, 2.71) | 6.28E-06 |
| | MEF2C TFBS in lung | 3.75 (2.2, 4.8) | 5.10E-04 |
| | Matrix fibroblast 1 open chromatin | 1.4 (0.72, 1.95) | 9.51E-04 |
| | ISX TFBS in lung | 4.77 (1.6, 6.56) | 0.0131 |
| | UTR3' variant | 1.41 (0.29, 2.17) | 0.0178 |
| | LMX1A TFBS in lung | 2.68 (0.75, 3.95) | 0.0223 |
| $FEV_1$/FVC | OLIG1 TFBS in lung | 5.25 (3.77, 6.3) | 4.14E-06 |
| | Alveolar type 1 open chromatin | 1.12 (0.73, 1.47) | 1.18E-05 |
| | Myofibroblast open chromatin | 1.2 (0.79, 1.57) | 8.70E-05 |
| | TF7L1 TFBS in lung | 2.57 (1.45, 3.39) | 1.83E-04 |
| | ZEP1 TFBS in bronchus | 3.55 (1.7, 4.79) | 0.0019 |
| | TCF7L2 TFBS in bronchus | 4.96 (2.89, 6.14) | 0.0030 |
| | Exonic variant | 1.23 (0.38, 1.89) | 0.0065 |
| | DUX4 TFBS in lung | 4.2 (1.81, 5.45) | 0.0073 |
| | MEIS1 TFBS in lung | 3.47 (1.28, 4.71) | 0.0102 |
| | YBOX1 TFBS in lung | 2.85 (0.9, 4.02) | 0.0160 |
| | IRF4 TFBS in lung | 1.63 (0.35, 2.47) | 0.0201 |
| | XBP1 TFBS in lung | 3.32 (0.85, 4.59) | 0.0201 |
| | MZF1 TFBS in lung | 1.45 (0.21, 2.32) | 0.0285 |
| PEF | Myofibroblast open chromatin | 2.31 (1.69, 2.86) | 9.27E-08 |
| | MYB TFBS in lung | 4.55 (2.57, 5.87) | 6.32E-04 |
| | PO3F1 TFBS in lung | 4.6 (2.23, 5.91) | 0.0037 |
| | ZN423 TFBS in lung | 3.22 (1.28, 4.42) | 0.0053 |
| | TAL1 TFBS in lung | 3.21 (0.93, 4.45) | 0.0160 |

*TFBS – transcription factor binding site.

**Supplementary Table 14: (Excel – sheet "Prioritised genes")**
List of prioritised genes according to the number of lines of variant-to-gene evidence implicates the gene (n_evidence column). The "Novel" column indicates whether the gene was previously prioritised in our previous paper[52].

**Supplementary Table 15: (Excel – sheet "Annotated credible sets")**
List of missense variants that are "putatively causal" i.e. account for >50% posterior probability in their 99% credible set. SIFT, PolyPhen and CADD measures of deleterious effect are shown with deleterious/damaging for SIFT/Polyphen of CADD PHRED >20 flagged in red as deleterious.

**Supplementary Table 16: (Excel – sheet "Druggability").**

Drug Gene Interaction Database (DGIDB), we surveyed 559 genes supported by ≥2 criteria. We found 292 drugs indicated by ChEMBL interactions mapping to 55 genes

Columns:

- Drug; drug/compound name,
- ChEMBL_ID; drug/compound identification number from ChEMBL,
- Gene; mapped gene(s),
- Gene_source; line(s) of evidence for each mapped gene, and signal implicated (including the associated lung function trait),
- Indication(Phase); drug indication phase. Phase 1: Testing of drug on healthy volunteers for dose-ranging; Phase 2: Testing of drug on patients to assess efficacy and safety; Phase 3: Testing of drug on patients to assess efficacy, effectiveness and safety; and Phase 4: Approval of drug and post-marketing surveillance,
- MAB; whether the drug is monoclonal antibody,
- Cancer; whether the drug is used to for the treatment of some form of cancer,
- AsthmaCOPD; whether the drug is indicated for the treatment of asthma or COPD

**Supplementary Table 17: Stratified LD score regression analysis of four lung function traits heritability enrichment at lung and smooth-muscle specific histone marks**

The four lung function traits were significantly enriched in nearly all histone marks specific to lung and smooth muscle containing cell lines. The most significant cell-type-specific annotation enrichment was found in H3K4me1 of fetal lung, with 6.99% input SNPs explaining 57.81% ($P=4.31\times10^{-26}$), 49.60% ($P=1.58\times10^{-19}$), 42.45% ($P=2.01\times10^{-22}$) and 36.20% ($P=4.81\times10^{-22}$) of the SNP-chip heritability for $FEV_1/FVC$, PEF, $FEV_1$ and FVC, respectively. SNPs associated with $FEV_1/FVC$ were concordantly found to be most significantly enriched in all the tested cell-type-specific annotations.

| Cell type | Chromatin mark | Proportion of overlapping SNPs | Trait | Proportion of heritability | Proportion of heritability standard error | Fold Enrichment | Enrichment standard error | Enrichment P-value |
|---|---|---|---|---|---|---|---|---|
| **Fetal Lung** | H3K4me1 | 6.99% | FVC | 36.20% | 0.0262 | 5.1766 | 0.3750 | 4.81E-22** |
| | | | FEV1 | 42.45% | 0.0301 | 6.0712 | 0.4300 | 2.01E-22** |
| | | | FEV1/FVC | 57.81% | 0.0391 | 8.2674 | 0.5594 | 4.31E-26** |
| | | | PEF | 49.60% | 0.0380 | 7.0928 | 0.5439 | 1.58E-19** |
| | H3K4me3 | 1.07% | FVC | 9.50% | 0.0169 | 8.8480 | 1.5716 | 1.81E-06** |
| | | | FEV1 | 9.02% | 0.0176 | 8.3946 | 1.6356 | 1.15E-05** |
| | | | FEV1/FVC | 15.14% | 0.0245 | 14.0977 | 2.2778 | 3.02E-08** |
| | | | PEF | 13.55% | 0.0260 | 12.6198 | 2.4228 | 2.14E-06** |
| | H3K9ac | 1.30% | FVC | 11.56% | 0.0196 | 8.8912 | 1.5073 | 5.83E-07** |
| | | | FEV1 | 12.60% | 0.0212 | 9.6900 | 1.6340 | 2.51E-07** |
| | | | FEV1/FVC | 19.17% | 0.0279 | 14.7445 | 2.1465 | 1.66E-10** |
| | | | PEF | 15.08% | 0.0244 | 11.5986 | 1.8752 | 2.07E-08** |
| **Lung** | H3K4me1 | 1.75% | FVC | 7.25% | 0.0155 | 4.1330 | 0.8850 | 0.000428** |
| | | | FEV1 | 8.60% | 0.0192 | 4.9047 | 1.0965 | 0.000409** |
| | | | FEV1/FVC | 13.37% | 0.0248 | 7.6261 | 1.4158 | 6.13E-06** |
| | | | PEF | 7.61% | 0.0198 | 4.3404 | 1.1314 | 0.003694* |
| | H3K4me3 | 0.56% | FVC | 4.78% | 0.0157 | 8.4965 | 2.7853 | 0.008152* |
| | | | FEV1 | 4.02% | 0.0153 | 7.1541 | 2.7150 | 0.025478* |
| | | | FEV1/FVC | 4.85% | 0.0169 | 8.6157 | 2.9972 | 0.010893* |
| | | | PEF | 2.82% | 0.0182 | 5.0094 | 3.2346 | 0.214195 |
| **Colon Smooth Muscle** | H3K4me1 | 3.52% | FVC | 18.12% | 0.0204 | 5.1460 | 0.5790 | 1.12E-11** |
| | | | FEV1 | 24.02% | 0.0258 | 6.8229 | 0.7320 | 7.32E-14** |
| | | | FEV1/FVC | 27.99% | 0.0278 | 7.9507 | 0.7906 | 1.18E-16** |
| | | | PEF | 22.94% | 0.0269 | 6.5156 | 0.7644 | 2.86E-12** |
| | H3K4me3 | 1.44% | FVC | 10.54% | 0.0159 | 7.3147 | 1.1070 | 5.47E-08** |
| | | | FEV1 | 10.12% | 0.0165 | 7.0238 | 1.1454 | 3.31E-07** |
| | | | FEV1/FVC | 15.12% | 0.0217 | 10.4936 | 1.5069 | 1.27E-09** |
| | | | PEF | 11.43% | 0.0213 | 7.9327 | 1.4764 | 1.97E-06** |
| | H3K9ac | 0.57% | FVC | 3.09% | 0.0114 | 5.4418 | 2.0154 | 0.029651* |
| | | | FEV1 | 4.43% | 0.0112 | 7.8155 | 1.9826 | 0.000735** |
| | | | FEV1/FVC | 5.71% | 0.0139 | 10.0616 | 2.4434 | 0.000191** |
| | | | PEF | 3.43% | 0.0107 | 6.0446 | 1.8892 | 0.006954* |
| **Stomach Smooth Muscle** | H3K4me1 | 2.39% | FVC | 14.59% | 0.0193 | 6.0962 | 0.8086 | 1.53E-09** |
| | | | FEV1 | 19.70% | 0.0255 | 8.2355 | 1.0653 | 1.44E-10** |
| | | | FEV1/FVC | 22.20% | 0.0231 | 9.2769 | 0.9635 | 4.26E-15** |
| | | | PEF | 17.81% | 0.0208 | 7.4433 | 0.8709 | 4.09E-12** |
| | H3K4me3 | 2.01% | FVC | 13.46% | 0.0170 | 6.6985 | 0.8453 | 1.51E-10** |
| | | | FEV1 | 15.76% | 0.0196 | 7.8438 | 0.9775 | 4.25E-11** |
| | | | FEV1/FVC | 21.06% | 0.0227 | 10.4853 | 1.1285 | 5.82E-15** |
| | | | PEF | 17.14% | 0.0233 | 8.5317 | 1.1608 | 2.16E-10** |
| | H3K9ac | 1.40% | FVC | 9.19% | 0.0169 | 6.5598 | 1.2047 | 7.38E-06** |
| | | | FEV1 | 12.63% | 0.0200 | 9.0171 | 1.4282 | 7.75E-08** |
| | | | FEV1/FVC | 15.03% | 0.0221 | 10.7298 | 1.5756 | 2.53E-09** |
| | | | PEF | 11.55% | 0.0200 | 8.2451 | 1.4281 | 3.25E-07** |
| | H3K27ac | 2.67% | FVC | 10.09% | 0.0148 | 3.7858 | 0.5562 | 1.21E-06** |
| | | | FEV1 | 15.74% | 0.0215 | 5.9046 | 0.8053 | 7.67E-09** |

| Cell type | Chromatin mark | Proportion of overlapping SNPs | Trait | Proportion of heritability | Proportion of heritability standard error | Fold Enrichment | Enrichment standard error | Enrichment P-value |
|---|---|---|---|---|---|---|---|---|
| | | | FEV1/FVC | 18.20% | 0.0239 | 6.8249 | 0.8962 | 4.96E-10** |
| | | | PEF | 14.36% | 0.0209 | 5.3854 | 0.7845 | 2.46E-08** |

*denotes FDR < 0.05

**denotes significant at P<0.05 after Bonferroni correction for multiple hypotheses.

**Supplementary Table 18: Association results of COPD and $FEV_1$/FVC with multi-ancestry or ancestry-specific GRS in ancestry groups in UK Biobank**

UK Biobank individuals were divided into ancestry groups as described in Supplementary Table 4 and Supplementary Figure 1 and were included for association analysis. The multi-ancestry/ancestry-specific genetic risk score was tested with COPD and $FEV_1$/FVC. COPD case was defined as $FEV_1$/FVC < 0.7 and $FEV_1$ < 80% predicted (GOLD 2-4 standards). For the $FEV_1$/FVC model, linear regression was used with covariates adjusted as described in the Online Methods. The COPD model was only fitted in ancestry groups with >100 COPD cases using logistic regression. Abbreviations: OR=odds ratio; 95LCI/UCI=lower/upper bound of 95% confidence intervals; P=p-value; N=sample size.

| Ancestry groups | Multi-ancestry GRS | | | | Ancestry-specific GRS | | | | Total N | N Control | N Case | Number of SNPs constructing GRS |
|---|---|---|---|---|---|---|---|---|---|---|---|---|
| | Effect size* (OR/Beta) | 95LCI | 95UCI | P | Effect size* (OR/Beta) | 95LCI | 95UCI | P | | | | |
| **$FEV_1$/FVC** | | | | | | | | | | | | |
| UK Biobank AFR | 0.1534 | 0.1106 | 0.1961 | 8.19E-19 | 0.1465 | 0.1030 | 0.1899 | 7.59E-17 | 4227 | | | 437 |
| UK Biobank AMR | 0.2540 | 0.2163 | 0.2917 | 8.12E-36 | 0.2318 | 0.1939 | 0.2698 | 7.53E-29 | 2798 | | | 434 |
| UK Biobank EAS | 0.1960 | 0.1203 | 0.2716 | 2.65E-12 | 0.1937 | 0.1162 | 0.2711 | 5.54E-12 | 1564 | | | 406 |
| UK Biobank EUR | 0.2787 | 0.2753 | 0.2820 | <4.94E-324 | 0.2698 | 0.2665 | 0.2731 | <4.94E-324 | 320656 | | | 442 |
| **COPD** | | | | | | | | | | | | |
| UK Biobank AFR | 1.5123 | 1.2540 | 1.8237 | 1.50E-05 | 1.4886 | 1.2306 | 1.8007 | 4.19E-05 | 4227 | 3977 | 250 | 437 |
| UK Biobank AMR | 1.5591 | 1.3031 | 1.8654 | 1.21E-06 | 1.5481 | 1.2925 | 1.8542 | 2.06E-06 | 2798 | 2647 | 151 | 434 |
| UK Biobank EUR | 1.6340 | 1.6110 | 1.6574 | <4.94E-324 | 1.6078 | 1.5852 | 1.6307 | <4.94E-324 | 320656 | 296594 | 24062 | 442 |

*Effect sizes are change in Z-score units for $FEV_1$/FVC results and odds ratios for COPD results per SD change in GRS

**Supplementary Table 19: Demographics of COPD case-control cohorts included in genetic risk score analysis**

Descriptive statistics for each cohort are given separately for cases and controls, for 5 cohorts: the COPDGene study, ECLIPSE study (Evaluation of COPD Longitudinally to Identify Predictive Surrogate End-points), GenKOLS (the Bergen, Norway COPD cohort), NETT/NAS (the National Emphysema Treatment Trial [NETT] and the Normative Aging Study [NAS]) and the SPIROMICS study. Abbreviation: SD=standard deviation; age is given in years, height in centimetres, FEV1 and FVC in litres.

| **Cohort** | Case-control status | Total N | **Female (%)** | Age range | Mean age (SD) | Height range (cm) | Mean height (SD) | N with spirometry data available | Mean FEV1 (SD) | Mean FEV1/FVC (SD) | Mean FVC (SD) | % ever smokers (N with ever smoking data available) | Pack-years range (N with pack-years data available) | Mean pack-years (SD) |
|---|---|---|---|---|---|---|---|---|---|---|---|---|---|---|
| **SPIROMICS (Non-Hispanic White)** | Cases | 931 | 41.46 | 41-80 | 65.90 (7.79) | 141-197 | 170.18 (9.75) | 931 | 1.42 (0.62) | 0.48 (0.13) | 2.94 (0.92) | 930 | 0-450 (930) | 55.75 (29.71) |
| | Controls | 373 | 54.52 | 40-80 | 61.27 (9.60) | 147-205 | 169.41 (9.66) | 373 | 2.87 (0.70) | 0.76 (0.04) | 3.77 (0.92) | 308 | 0-150 (372) | 36.05 (25.92) |
| **SPIROMICS (African-American)** | Cases | 174 | 52.3 | 43-79 | 60.49 (8.13) | 145-193 | 169.46 (9.19) | 174 | 1.23 (0.57) | 0.49 (0.13) | 2.45 (0.82) | 173 | 0-110 (174) | 43.34 (19.37) |
| | Controls | 142 | 57.04 | 40-72 | 53.44 (8.12) | 151-192 | 169.56 (8.79) | 142 | 2.62 (0.61) | 0.78 (0.05) | 3.34 (0.76) | 119 | 0-105 (142) | 31.79 (19.83) |
| **COPDGene (Non-Hispanic White)** | Cases | 2811 | 44.33 | 59-71 | 64.67 (8.19) | 162.6-176.5 | 169.69 (9.40) | 2811 | 1.45 (0.64) | 0.49 (0.13) | 2.95 (0.91) | 2811 | 38-70.5 (2811) | 56.28 (27.98) |
| | Controls | 2534 | 50.67 | 52-66 | 59.50 (8.73) | 162.6-176.9 | 169.67 (9.44) | 2534 | 2.96 (0.69) | 0.78 (0.05) | 3.81 (0.90) | 2534 | 23.3-46.8 (2534) | 37.84 (20.30) |
| **COPDGene (African-American)** | Cases | 821 | 44.82 | 52-65 | 58.98 (8.17) | 163-178 | 170.55 (9.84) | 821 | 1.40 (0.60) | 0.53 (0.12) | 2.61 (0.85) | 821 | 25.4-52 (821) | 42.38 (23.03) |
| | Controls | 1749 | 41.85 | 48-56 | 52.81 (6.00) | 165-178 | 171.28 (9.41) | 1749 | 2.80 (0.65) | 0.80 (0.05) | 3.52 (0.84) | 1749 | 32.7-43.9 (1749) | 36.37 (20.14) |
| **ECLIPSE** | Cases | 1764 | 32.99 | 59-69 | 63.63 (7.10) | 163-176 | 169.48 (9.01) | 1764 | 1.33 (0.51) | 0.45 (0.12) | 3.00 (0.90) | 1764 | 32-60 (1764) | 50.29 (27.42) |
| | Controls | 178 | 42.13 | 50-65 | 57.48 (9.44) | 164.3-178.8 | 171.69 (9.68) | 178 | 3.26 (0.81) | 0.79 (0.05) | 4.14 (1.04) | 178 | 18-38.8 (178) | 32.11 (24.84) |
| **GenKOLS** | Cases | 864 | 60.19 | 58-74 | 65.55 (10.04) | 163-169.9 | 169.85 (9.00) | 864 | 1.57 (0.71) | 0.51 (0.13) | 2.99 (0.95) | 864 | 19.6-41.3 (864) | 32.01 (18.55) |
| | Controls | 808 | 50.12 | 48-62 | 55.62 (9.71) | 165-178 | 171.79 (8.79) | 808 | 3.24 (0.73) | 0.79 (0.04) | 4.11 (0.94) | 808 | 9.3-26.7 (808) | 19.66 (13.58) |
| **NETT/NAS** | Cases | 376 | 35.9 | 64-71 | 67.49 (5.77) | 162.4-176 | 168.85 (9.58) | 376 | 0.82 (0.26) | 0.32 (0.06) | 2.62 (0.83) | 376 | 44-84 (376) | 66.43 (30.68) |
| | Controls | 435 | 0 | 65-75 | 69.80 (7.49) | 169.7-179.1 | 174.43 (6.77) | 435 | 3.03 (0.51) | 0.79 (0.05) | 3.83 (0.62) | 435 | 20-52.5 (435) | 40.66 (27.85) |

**Supplementary Table 20**: **Association results of genetic risk score and COPD in external case-control studies:**
Association results between both weighted and unweighted genetic risk scores and COPD are given for five case-control studies: the COPDGene study, the ECLIPSE study (Evaluation of COPD Longitudinally to Identify Predictive Surrogate End-points), GenKOLS (the Bergen, Norway COPD cohort), NETT/NAS (the National Emphysema Treatment Trial [NETT] and the Normative Aging Study [NAS]) and the SPIROMICS study. COPDGene and SPIROMICS are stratified into African-American and Non-hispanic white subgroups. Odds ratios and 95% confidence intervals are given on two scales: a per-Allele scale and a per standard deviation (SD) scale. Abbreviations: AA=African-American; NHW=Non-hispanic white; OR=odds ratio; 95LCI/UCI=lower/upper bound of 95% confidence intervals; P=p-value; N=sample size.

| Ancestry | Study group | per Allele | | | per Standard Deviation | | | P | N | | | Mean risk score | SD risk score |
|---|---|---|---|---|---|---|---|---|---|---|---|---|---|
| | | OR | 95LCI | 95UCI | OR | 95LCI | 95UCI | | Total | Cases | Controls | | |
| **Weighted** | | | | | | | | | | | | | |
| African | COPDGene (AA) | 1.0179 | 1.0089 | 1.0269 | 1.2032 | 1.0972 | 1.3193 | 8.69E-05 | 2570 | 821 | 1749 | *274.70 | *10.45 |
| | SPIROMICS (AA) | 1.0353 | 1.0112 | 1.0599 | 1.4670 | 1.1314 | 1.9020 | 0.0037 | 316 | 174 | 142 | 497.87 | 11.04 |
| Meta-analysis | | 1.0200 | 1.0116 | 1.0285 | 1.2301 | 1.1278 | 1.3418 | 2.96E-06 | 2886 | 995 | 1891 | 299.14 | **10.52 |
| European | COPDGene (NHW) | 1.0509 | 1.0388 | 1.0631 | 1.8158 | 1.58 | 2.0868 | 2.87E-17 | 1304 | 931 | 373 | 439.71 | 12.03 |
| | ECLIPSE | 1.0379 | 1.0328 | 1.0430 | 1.5870 | 1.4928 | 1.6871 | 1.43E-48 | 5345 | 2811 | 2534 | 464.19 | 12.42 |
| | GenKOLS | 1.0447 | 1.0293 | 1.0604 | 1.6603 | 1.3969 | 1.9733 | 8.75E-09 | 1942 | 1764 | 178 | 432.91 | 11.59 |
| | NETT/NAS | 1.0416 | 1.0251 | 1.0583 | 1.6331 | 1.3479 | 1.9786 | 5.47E-07 | 811 | 376 | 435 | 464.32 | 12.04 |
| | SPIROMICS (NHW) | 1.0445 | 1.0339 | 1.0551 | 1.6657 | 1.4790 | 1.8758 | 8.44E-17 | 1672 | 864 | 808 | 431.41 | 11.73 |
| Meta-analysis | | 1.0409 | 1.0368 | 1.0450 | 1.6322 | 1.5574 | 1.7108 | 7.07E-93 | 11074 | 6746 | 4328 | 450.88 | 12.10 |
| **Unweighted** | | | | | | | | | | | | | |
| African | COPDGene (AA) | 1.0079 | 0.9999 | 1.0159 | 1.0967 | 0.9987 | 1.2043 | 0.0529 | 2570 | 821 | 1749 | 355.18 | 11.79 |
| | SPIROMICS (AA) | 1.0232 | 1.0021 | 1.0446 | 1.3186 | 1.0260 | 1.6948 | 0.0304 | 316 | 174 | 142 | 445.41 | 12.08 |
| Meta-analysis | | 1.0097 | 1.0023 | 1.0172 | 1.1213 | 1.0105 | 1.0273 | 0.0104 | 2886 | 995 | 1891 | 365.09 | 11.82 |
| European | COPDGene (NHW) | 1.0479 | 1.0365 | 1.0595 | 1.7933 | 1.5637 | 2.05653 | 5.84E-17 | 1304 | 931 | 373 | 454.35 | 12.48 |
| | ECLIPSE | 1.0325 | 1.0276 | 1.0373 | 1.5112 | 1.4225 | 1.6065 | 3.45E-40 | 5345 | 2811 | 2534 | 451.37 | 12.93 |
| | GenKOLS | 1.0386 | 1.0246 | 1.0528 | 1.5516 | 1.3254 | 1.8164 | 4.67E-08 | 1942 | 1764 | 178 | 432.91 | 11.59 |
| | NETT/NAS | 1.0355 | 1.0202 | 1.0511 | 1.555 | 1.2873 | 1.8784 | 4.64E-06 | 811 | 376 | 435 | 451.50 | 12.64 |
| | SPIROMICS (NHW) | 1.0391 | 1.0294 | 1.0489 | 1.6184 | 1.4393 | 1.8198 | 8.56E-16 | 1672 | 864 | 808 | 450.51 | 12.55 |
| Meta-analysis | | 1.0357 | 1.0319 | 1.0396 | 1.5619 | 1.4913 | 1.6358 | 3.15E-79 | 11074 | 6746 | 4328 | 448.36 | 11.67 |

*Genetic variants with MAF < 0.05 and imputation quality less than 0.7 were excluded in constructing GRS

**Approximated in R as sqrt(sum(SD^2*(N-1))/sum(N-1)), where N is a vector of sample sizes, and SD is a vector of standard deviations.

**Supplementary Table 21: Association results of genetic risk score decile and COPD in external case-control studies**: Within each study group, individuals were divided according to their value of the weighted genetic risk score. Logistic regressions were fitted for each decile, comparing odds of COPD between members of each decile (2-10) and the reference decile (1, lowest risk decile). Fixed-effects meta-analysis results were given across COPDGene(Non-hispanic white), ECLIPSE, GenKOLS, NETT/NAS, SPIROMICS(Non-hispanic white) for European ancestry and across COPDGene(African-American), SPIROMICS(African-American) for African ancestry. Abbreviations: OR=Odds Ration; 95LCI/UCI=lower/upper bound of 95% confidence intervals; P=p-value.

| Decile | Meta-analysis of 5 European Cohorts | | | | Meta-analysis of 2 African Cohorts | | | |
|---|---|---|---|---|---|---|---|---|
| | OR | 95LCI | 95UCI | P | OR | 95LCI | 95UCI | P |
| 1 | 1 | | | | 1 | | | |
| 2 | 1.26 | 1.03 | 1.55 | 2.39E-02 | 1.12 | 0.75 | 1.66 | 5.78E-01 |
| 3 | 1.62 | 1.33 | 1.98 | 1.50E-06 | 1.29 | 0.86 | 1.93 | 2.11E-01 |
| 4 | 1.90 | 1.55 | 2.34 | 7.70E-10 | 1.20 | 0.80 | 1.80 | 3.75E-01 |
| 5 | 2.18 | 1.79 | 2.67 | 2.00E-14 | 1.35 | 0.88 | 2.05 | 1.66E-01 |
| 6 | 2.56 | 2.09 | 3.13 | 8.05E-20 | 1.41 | 0.96 | 2.08 | 8.17E-02 |
| 7 | 2.49 | 2.03 | 3.05 | 1.30E-18 | 1.35 | 0.89 | 2.06 | 1.56E-01 |
| 8 | 3.27 | 2.64 | 4.06 | 6.71E-27 | 1.58 | 1.06 | 2.36 | 2.45E-02 |
| 9 | 3.72 | 3.02 | 4.58 | 9.07E-35 | 1.53 | 1.01 | 2.31 | 4.28E-02 |
| 10 | 5.16 | 4.14 | 6.42 | 1.03E-48 | 2.18 | 1.45 | 3.30 | 2.05E-04 |

**Supplementary Table 22: (Excel – sheet "PheWAS_phenotypes") Details of 1909 phenotypes used in PheWAS:** The category column refers to how the phenotype was made, phenotype_group is used to group results for plots, phenotype_group_narrow is a more specific grouping. Short description is used for PheWAS plots.

**Supplementary Table 23: (Excel – sheet "single_variant_PheWAS") Results of 27 single variant PheWAS**
For each variant the top 3 associations are shown plus any associations with FDR <5%. (N_ID = Number of participants in the analysis, FDR = false discovery rate, P = p value, OR = odds ratio, L95 = lower boundary of the 95% confidence interval, L95 = upper boundary of the 95% confidence interval, MAF = minor allele frequency, MAC = minor allele count, MAC_cases = minor allele count in cases, MAC_controls = minor allelecount in controls. Z_T_STAT = the Z or T STAT output from Plink2, SE = standard error)

**Supplementary Table 24: (Excel – sheet "trait_PheWAS") Results of 4 trait-specific PheWAS for FDR <5%:**
(FDR = false discovery rate, P = p value, OR = odds ratio, L95 = lower boundary of the 95% confidence interval, L95 = upper boundary of the 95% confidence interval)

**Supplementary Table 25: (Excel – sheet "ConsensuspathDB")**
ConsensuspathDB pathways enriched (FDR <5%) for genes from our 559 with at least 2 lines of evidence for being causal.

**Supplementary Table 26: (Excel – sheet "IPA pathways")**
Ingenuity pathways enriched (P <0.05) for genes from our 559 with at least 2 lines of evidence for being causal.

**Supplementary Table 27: (Excel – sheet "pathway_PheWAS") Results of 29 pathway-specific PheWAS for FDR <5%:**
(FDR = false discovery rate, P = p value, OR = odds ratio, L95 = lower boundary of the 95% confidence interval, L95 = upper boundary of the 95% confidence interval)

**Supplementary Table 28: (Excel – sheet "eQTL pQTL")**
eQTL/pQTL colocalisation results.

**Supplementary Table 29: (Excel – sheet "PoPS_500KB_window")**
Polygenic Priority Score (PoPS) results using a +/-250KB window.

**Supplementary Table 30: (Excel – sheet "PoPS_1MB_window")**
Polygenic Priority Score (PoPS) results using a +/-500KB window.

**Supplementary Table 31: (Excel – sheet "Rare variants UKB WES")**
Look up of UK Biobank WES variants within +/-500kb of lung function sentinels.

**Supplementary Table 32: Literature review of key genes.**

| Gene | Description |
|---|---|
| ***ABCA3*** | ATP-binding cassette class A3 is a highly conserved multi-membrane-spanning protein that is critical in the regulation of pulmonary surfactant homeostasis [80]. *ABCA3* belongs to the ABC superfamily of transporters with several membrane domains that hydrolyse ATP to transport multiple substrates through membranes. *ABCA3* is mapped to chromosome 6p13.3 and highly expressed in alveolar type II cells [81] [82]. Among its related pathways and Gene Ontology include; CDK-mediated phosphorylation, removal of Cdc6 and metabolism of protein, and transporter activity and ATPase activity, coupled to transmembrane movement of substances [80].<br>It has been difficult to predict the processes of disruption encoded by ABCA3 variants on the principle of location in the gene or the mature protein. Nonetheless, over 200 diseases-associated to *ABCA3* variants have been identified (the majority being missense variants), of which less than 10% have been functionally restricted as disrupting *ABCA3* protein trafficking (type I) or ATPase-mediated phospholipid transport (type II) in cell-based systems. *ABCA3* variants have been classified comparatively to CFTR variants, variants can aftermath in the lack of a mature protein (ie., nonsense, frameshift, ~CFTR class I), impairment of intracellular trafficking (~CFTR class II), or faulty phospholipid transport into the lamellar bodies (~CFTR class III). *ABCA3* pathogenic variants, along with other surfactant-associated genes have been identified in patients (from newborn to adulthood) with assorted pulmonary diseases, suggesting that *ABCA3* variants may be consequence not only from the impairment of surfactant metabolism, but also from the triggering other cellular pathways that disrupts alveolar type II cells and alveolar epithelial-cell homeostasis [81].<br>Mutations in *ABCA3* are linked to respiratory failure in neonates, interstitial lung disease in children (chILD), pulmonary fibrosis (IPF) and diffuse parenchymal lung disease (DPLD) in adults [82]. The majority of which have been reported in patients that were compound heterozygous in both adults [82] and children [83]. Despite the fact that *ABCA3* variants have been identified in diverse pulmonary diseases, a study from the Copenhagen City Heart Study (n=64,000) found no evidence that the *ABCA3* missense common mutation E292V can increase the risk of developing COPD [84].<br>Although no specific therapy exists for diseases consequence of ABCA3 mutations, correctors that bind to CFTR mutations (ie., lumacaftor, tezacaftor, and elexacaftor, potentiators of CFTR transport function (ie., ivacaftor), may indicate that can be used as pharmacologic strategies when clinical trials fails, or tissue accessibility is not feasible [81]. Direct approaches that are aimed to decreasing aberrant cellular responses to mutant protein expression, inflammatory signalling/cytokine formulation, and/or cell death may be adequate in the design of new therapies [82]. Drugs that have served as successful treatment include, prostacyclin analogue, warfarin, and inhaled oxygen[80]. |
| ***ACAN*** | This gene is a member of the aggrecan/versican proteoglycan family. *ACAN* codes for aggrecan, an integral part of the extracellular matrix in cartilaginous tissue. Alterations in the cartilage matrix content affect its functional role [85]. Mutations in this gene may be involved in skeletal dysplasia and spinal degeneration. Multiple alternatively spliced transcript variants that encode different protein isoforms have been observed in this gene[80].<br>*ACAN* was significantly associated with patient survival in a methylation study that examined CIMP (CpG island methylator phenotype) in patients with pulmonary adenocarcinoma after a surgical resection in 230 pulmonary adenocarcinoma cases [85].<br>Mutations on *ACAN* were found to be to be correlated with NSCLC (Non-small cell lung cancer) metastasis [86]. |
| ***ADAMTS10*** | *ADAMTS10* is a member of ADAMTS (disintegrin and [zinc] metalloproteinase domain with thrombospondin type-1 motifs), a multidomain extracellular protease enzymes superfamily. Evolutionary studies of ADAMTS proteins indicate key embryologic and physiological roles in humans with participation in multiple pathways, including connective tissue organization, coagulation, inflammation, arthritis, angiogenesis, and cell migration [87] [88]. Moreover, ADAMTS proteases are anti-cancer or pro-tumorigenic molecules. These enzymes are secreted by tumour or stromal cells, which then can alter the primary tumour microenvironment by proteolytic-dependent or independent mechanisms, thus suggesting that ADAMTS are not the primary oncogenes [87]. |

| Gene | Description |
|---|---|
| | While ADAMTS family have nonredundant functions in organogenesis (fetal lung, liver and kidney), *ADAMTS10* has been found to also be expressed in skin fibroblasts, fibrillin microfibrils in ocular zonule, chondrocytes, and the adult heart. Playing important roles in connective tissue organization, coagulation, inflammation, arthritis, angiogenesis and cell migration. Related pathways and gene ontology encompass diseases of glycosylation and O-linked glycosylation of mucins, as well as peptidase activity and metalloendopeptidase activity [88 89 80]. Have important roles in connective tissue organization, coagulation, inflammation, arthritis, angiogenesis and cell migration. The product of this gene plays a major role in growth and in skin, lens, and heart development.<br>Mendelian disorders linking *ADAMTS10* mutations, include acromelic dysplasias and syndromes distinguished by short stature and disproportionate distal limb shortening, each accompanied by other characteristic anomalies, such as the Weill-Marchesani syndrome. Weill-Marchesani syndrome is a rare-disease of the connective tissue that is distinguished by symptoms implicated by multiple structures, including the skeleton (proportionate short stature, joint stiffness, brachydactyly, scoliosis, lumbar lordosis, and maxillary hypoplasia), the eye (microspherophakia, ectopia lentis, severe myopia, glaucoma, shallow anterior chamber, cataract, and blindness), and the cardiovascular system (aortic valve stenosis, pulmonary valve stenosis, mitral valve insufficiency, persistent ductus arteriosus, and ventricular septal defect). This syndrome is mostly inherited as an autosomal recessive due to mutations of the *ADAMTS10* gene, in-frame deletion of the fibrillin 1 gene, as well as three missense variants, two non-sense variants and two splice site variants [88 89 90 91]. |
| ***ADGRG6*** | Encodes a G protein-coupled receptor. Is upregulated in human umbilical vein endothelial cells. Essential for normal differentiation of promyelinating Schwann cells and for normal myelination of axons [92]. *ADGRG6* is also important in lung development and micro-angiopathy [93]. Regulates neural, cardiac and ear development via G-protein- and/or N-terminus-dependent signaling (By similarity). May act as a receptor for PRNP which may promote myelin homeostasis (By similarity) [80].<br>Analysis of differential gene expression revealed an increased on *ADGRG6* /GPR126 in IPAH-PASMCs (idiopathic pulmonary arterial hypertension - disorder in pulmonary artery smooth muscle cells), compared to control-PASMCs, thus suggesting *ADGRG6* as a novel therapeutic target for IPAH [94]. Gene expression analysis of this gene in human lung tissue has also shown a decreased expression with COPD and patients with decreased carbon monoxide uptake per alveolar volume [93]. GWAS of lung function have identified missense variant predicted to be deleterious associated with $FEV_1$/FVC. The ADGRG6 locus is described in association with lung function, COPD, and height [95]. |
| ***ADRB2*** | This gene encodes beta-2-adrenergic receptor which is a member of the G protein-coupled receptor superfamily. This receptor is directly associated with one of its ultimate effectors, the class C L-type calcium channel Ca(V)1.2. This receptor-channel complex also contains a G protein, an adenylyl cyclase, cAMP-dependent kinase, and the counterbalancing phosphatase, PP2A. The assembly of the signalling complex provides a mechanism that ensures specific and rapid signalling by this G protein-coupled receptor. This receptor is also a transcription regulator of the alpha-synuclein gene, and together, both genes are believed to be associated with risk of Parkinson's Disease. This gene is intronless. Different polymorphic forms, point mutations, and/or downregulation of this gene are associated with nocturnal asthma, obesity, type 2 diabetes and cardiovascular disease [80].<br>*ADRB2* is an important regulator of airway smooth muscle tone. *ADRB2* acts on smooth muscle functions to dilate and antagonise constriction of the airways, thereby protecting the lungs from long-term bronchoconstriction. Variants that impair the function of the *ADRB2* receptor could result in narrowing of the airways—including small airways—and increase susceptibility to COPD High levels of *ADRB2* are also associated with reduced lung function and asthma [96]. *ADRB2* is also an anti-inflammatory gene and has been reported to be involved in hyperinflammation [97] and autoimmune disorders such as rheumatoid arthritis [98]. In addition, *ADRB2* has been demonstrated to be downregulated in prostate and breast cancer, and functional gain of *ADRB2* decreases cancer progression [99 100].<br>A study revealed that increased levels of *ADRB2* were associated with a longer survival time compared with the respective low expression groups in patients with squamous cell lung |

| Gene | Description |
|---|---|
| | carcinoma(SCC). It is suggested that *ADRB2* might play a key role in COPD-associated SCC and may provide novel therapeutic targets for the treatment of lung cancer [101]. |
| ***AP3B1*** | This gene encodes a protein that may play a role in organelle biogenesis associated with melanosomes, platelet dense granules, and lysosomes. The encoded protein is part of the heterotetrameric AP-3 protein complex which interacts with the scaffolding protein clathrin. Mutations in this gene are associated with Hermansky-Pudlak syndrome type 2 [80]. HPS2 is an autosomal recessive inherited disease caused by mutations in *AP3B1* , resulting in pulmonary fibrosis (PF) and immunodeficiency. However, the total number of HPS2 cases reported was less than 40 as of March 2021, making it difficult to accurately assess [102].<br>A study conducted whole-exome sequencing in 233 hospitalized COVID-19 patients, identified AP3B1 variants significantly enriched in COVID-19 patients experiencing severe cytokine storms and fatal outcomes in COVID-19 [103 104]. |
| ***CACNA1S*** | This gene encodes one of the five subunits of the slowly inactivating L-type voltage-dependent calcium channel in skeletal muscle cells. Mutations in this gene have been associated with hypokalemic periodic paralysis, thyrotoxic periodic paralysis and malignant hyperthermia susceptibility [80].<br>Mutations in this gene have been reported to be mainly responsible for Hypokalemic periodic paralysis (HOKPP), which paralytic attacks generally spare the respiratory muscles and the heart causing severe respiratory phenotype and a reduced susceptibility to cold exposure [105 106]. A study in UK Biobank identified a functionally deleterious missense variant of *CACNA1S* (rs3850625) associated with FVC [107]. Furthermore, an analysis found that *CACNA1S* was abundantly expressed in normal tissue but not in cancer tissue; thus, this gene could serve as tumour suppressor gene markers for specific subtypes of cancer [108]. |
| ***CFH*** | Member of the Regulator of Complement Activation (RCA) gene cluster. Gene codes for Complement Factor H, a glycoprotein secreted into the bloodstream and inhibits complement, thereby restricting the defence mechanism against microbial infections [80 109 110]. Also increases the decay of the complement alternative pathway (AP) C3 convertase C3bBb, which inhibits production of C3b, a key regulator of the complement amplification loop [111].<br>Diseases associated with this gene include Complement Factor H Deficiency, hemolytic-uremic syndrome (HUS) and chronic hypocomplementemic nephropathy (GeneCards).<br>Expression of Complement factor H is increased in bronchoalveolar lavage fluid and sputum from patients with lung cancer [112], and high levels of *CFH* have been shown to decrease the risk of death in patients with small-cell lung cancer (HR 0.23, 95% CI 0.10 to 0.57, p<0.001), indicating its potential use as a risk of death biomarker [113]. |
| ***CLDN18*** | This gene belongs to the claudin family, key elements of tight junctions which regulate paracellular barrier functions. Tight junction functions as a physical barrier to block solutes and water from passing freely through the paracellular space between epithelial or endothelial cell sheets, and also are implicated in maintaining cell polarity and signal transductions. *CLDN18* is highly expressed in the stomach and is strictly confined to differentiated epithelial cells of the gastric mucosa. Related pathways and gene ontology annotations include, cytoskeleton remodelling, regulation of actin cytoskeleton by Rho GTPases and Sertoli-Sertoli Cell Junction Dynamics, as well as identical protein binding and structural molecule activity [80 114].<br>Diseases linked to *CLDN18* are Bile Duct Cancer, Urachus Cancer, as well as Diffuse-type Gastric Cancer. In particular, studies have found an inter-chromosomal translocation between *CLDN18* and ARHGAP (Rho GTPase-activating protein that contributes to the organization of actin and microtubule cytoskeletons), leading to a RhoGAP domain-containing fusion protein with impaired function of *CLDN18* and RhoGAP (*CLDN18 -ARHGAP26*/6 fusion). Although this fusion has been associated to poor prognosis of Gastric Cancer, it could be considered as target for drug screening and for therapeutic strategies [115 116].<br>Cldn18.1 is a lung-specific isoform of the *CLDN18* gene, and it is highly expressed in alveolar epithelial cells. While, its role in the human airway epithelium has not been well investigated, a study in KO mice showed that the lack of Cldn18 suggested an important role in the regulation of airway progenitor cell homeostasis and cell composition, and implicate Cldn18 in regulation of downstream signalling pathways that inhibit goblet cell differentiation. Cldn18 is downregulated in asthma and chronic obstructive pulmonary disease (COPD), therefore, diseases characterized |

| Gene | Description |
|---|---|
| | by goblet cell hyperplasia, modulation of Cldn18 expression may be beneficial in the treatment of airway diseases, particularly Cldn18 is significantly induced in pulmonary epithelial cells following corticosteroid therapy [117 118]. |
| ***CYTL1*** | Cytokine-like protein 1 was first discovered from cells derived from bone marrow and cord blood mononuclear cells that function as haematopoietic stem/progenitor cells and bear the CD34 surface marker. Even though the Cytl1 gene has been mapped to chromosome 4p15–p16 in humans, the precise molecular structure of the Cytl1 protein is still to be determined, and there is doubt as to the true classification of Cytl1 due to its cytokine-like properties and chemokine activity [119].<br>KO mice studies have found that the lack of expression had an increased susceptibility to cartilage destruction consequence of osteoarthritis. Moreover, *CYTL1* may significantly inhibit osteoarthritis and downregulate the expression of inflammatory cytokines. This gene has also been reported associated to cardiac fibrosis, benign prostatic hypertrophy, neuroblastoma, lung squamous cell carcinoma, and familial colorectal cancer. However, although *CYTL1* play a key role in sepsis, the effects of this gene on neutrophil function is not well understood [120]. |
| ***FGFR1*** | Fibroblast Growth Factor Receptor 1 is a member of the fibroblast growth factor receptor family (*FGFR*). The protein is a tyrosine-protein kinase that acts as cell-surface receptor for fibroblast growth factors and plays an essential role in the regulation of embryonic development, cell proliferation, differentiation and migration. Mutations in this gene have been associated with Pfeiffer syndrome, Jackson-Weiss syndrome, Antley-Bixler syndrome, osteoglophonic dysplasia, and autosomal dominant Kallmann syndrome 2. *FGFR1* signals primarily via the PI3K and MAPK pathways and is involved in cancer processes such as auto- and paracrine activation, amplification and overexpression.<br>*FGFR1* play important roles in lung development and regeneration, mediating signalling between the epithelium and mesenchyme [121 122].<br>Epithelial *FGFR1* expression is minimal throughout lung development, homeostasis and regeneration but is strongly expressed in cartilage progenitors and airway smooth muscle cells during lung development [121]. Moreover, *FGFR1* is expressed in lipofibroblasts and vascular smooth muscle cells. In adult lung FGFR1 is downregulated in smooth muscle cells with upregulation after injury. *FGFR1* expression is maintained in mesenchymal alveolar cells with lower levels in alveolar myofibroblasts during alveologenesis [121].<br>*FGFR1* expression is increased in IPF lung homogenate lysates compared to donor control [123].<br>*FGFR1* localises in the bronchial epithelium, airway smooth muscle, submucosal glandular epithelium and vascular smooth muscle. Elevated levels of *FGFR1* is observed in airway smooth muscle and bronchial epithelium in COPD tissues versus control [124].<br>FGFRs have been shown to be overexpressed in numerous cancer types. *FGFR1* is frequently overexpressed in breast cancer. Amplification of *FGFR1* locus at chromosome 8p has been described in several cancer types, occurring in 20% of pulmonary squamous cell carcinomas, and is a potential therapeutic target in small-cell lung cancer [125 125 126]. Moreover, activation of *FGFR1* promotes epithelial-mesenchymal transition (EMT) [126]. |
| ***GATA5*** | Encodes *GATA5* (GATA Binding Protein 5), a transcription factor containing two GATA-type zinc fingers. Diseases associated with *GATA5* include Congenital Heart Defects and Aortic Valve Disease [80]. *GATA5* is required during cardiovascular development [126] and plays an important role in smooth muscle cell diversity [80].<br>*GATA5* has a unique spatial and temporal function in the embryonic heart and lung. Only *GATA5* is expressed in the pulmonary mesenchyme. *GATA5* is expressed in tissue-restricted subsets of smooth muscle cells (SMCs), including bronchial SMCs. It is thought that GATA5 is important in the transcriptional pathway(s) that underlie SMC diversity [127]. |
| ***GLI3*** | GLI Family Zinc Finger 3 belongs to the C2H2 type zinc finger proteins of the Gli family. These are DNA-binding transcription factors and mediate sonic hedgehog (shh) signalling. Mutations in *GLI3* are associated with Greig cephalopolysyndactyly syndrome, Pallister-Hall syndrome, preaxial polydactyly type IV, and postaxial polydactyly types A1 and B [80].<br>Gli3 can exist as a full length (Gli3-FL/*GLI3* -FL) or repressor (Gli3-R/*GLI3* -R) form. In response to HH activation, *GLI3*-FL regulates HH genes by targeting the GLI1 promoter. In the absence of HH signaling, *GLI3*is phosphorylated leading to its partial degradation and the generation of *GLI3*-R |

| Gene | Description |
|---|---|
| | which represses HH functions. *GLI3*is involved in tissue development, immune cell development and cancer. The absence of Gli3 in mice impaired brain and lung development and *GLI3* mutations in humans are the cause of Greig cephalopolysyndactyly (GCPS) and Pallister Hall syndromes (PHS) [128].<br>*GLI3* mRNA and protein expression is decreased in NRF2-null mouse embryonic fibroblasts. Moreover, NRF2-null cells grow fewer and shorter cilia and display impaired Hedgehog signalling, a cilia-dependent pathway [129].<br>*GLI3* mRNA levels increase concomitantly with the differentiation of the epithelium at ALI, highlighting a role for HH signalling in airway epithelial cell differentiation. Preventing HH activation leads to establishment of remodelled epithelium [130].<br>Gli3 processing has important roles in mouse lung organogenesis via shh signalling which also controls mesenchymal proliferation and differentiation. Lack of Shh signaling, mediated by the Gli proteins, leads to severe pulmonary hypoplasia. Shh signaling prevents Gli3 proteolysis to generate its repressor forms (Gli3R) in the developing murine lung. In Shh(-/-) lung, Gli3R is elevated, which appears to contribute to defects in proliferation and differentiation observed in the Shh(-/-) mesenchyme, where Gli3 is normally expressed. Moreover, Shh(-/-);Gli3(-/-) lungs exhibit enhanced growth potential [131]. |
| ***HIST1H2BE*** | Gene encodes a variant of the histone H2B family, one of the four core histone proteins [80]. No previous evidence to show a link between *HIST1H2BE* and lung. H2B protein expression is dysregulated in breast cancers [132], and altered *HIST1H2BE* expression is altered in numerous breast cancer cell lines, which results in decreased proliferation [133]. *HIST1H2BE* is also downregulated in goblet cell adenocarcinoma [134]. |
| ***HMCN1*** | Encodes a large extracellular member of the immunoglobulin family. Mutations in the gene may be associated with age-related macular degeneration. The protein may have a role in the formation of epithelial cell junctions and have effects on cardiac fibroblast migration [80].<br>Rare damaging variants in *HMCN1* have been implicated in the pathogenesis of pulmonary atresia. *HMCN1* affects the formation of cell contacts required for tissue organisation, migration and invasion and the formation of cell-cell and cell-ECM contacts via TGFb signalling [135] [136].<br>*HMCN1* has also been shown to regulate invasiveness of ovarian cancer via RhoA signalling pathway in fibroblasts and is upregulated in cancer associated fibroblasts [137]. |
| ***IGF2BP2*** | Insulin-like growth factor 2 mRNA-binding protein 2 (*IGF2BP2*) is a protein which is encoded by *IGF2BP2* gene. IGF2BPs are increasingly implicated in modulating varies of biological processes, including development, tumorigenesis, and stemness. It is also required for their recognition of m6A modifications and is critical for mRNA stability and translation. Dysregulation of *IGF2BP2* is implicated in certain diseases such as diabetes and cancer [80] [137].<br>Studies have found an overexpression of *IGF2BP2* in both ovarian cancer and ovarian low malignant potential tumour samples compared to either normal ovary or ovarian adenomas samples. *IGF2BP2*/IGF-1/IGF-1 receptor signaling pathways has also been found involved in cancer-mediated endothelial recruitment, which is an important feature of metastatic cancer in the tumour microenvironment. In addition it has been found that an lncRNA (*IGF2BP2*-AS1) was associated with better overall survival in lung squamous cell carcinoma [138].<br>*IGF2BP2* is highly expressed in both of classically activated (M1) and alternatively activated (M2) macrophages. The *IGF2BP2*−/− macrophages are refractory to interleukin-4 (IL-4) induced activation and alleviate cockroach extract induced pulmonary allergic inflammation. Thus, this gene is a potential target to modulate macrophages activation for treating pulmonary inflammation [138]. |
| ***IGHMBP2*** | Immunoglobulin Mu DNA Binding Protein 2. Helicase superfamily member that binds a specific DNA sequence from the immunoglobulin mu chain switch region. Mutations in the gene lead to spinal muscle atrophy with respiratory distress type 1 [80]. The most critical component of *SMARD1* is respiratory failure due to paralysis of the diaphragm, this typically manifests within the first few months of life.<br>The gene codes for an ATP-dependent helicase for DNA and RNA, belonging to superfamily1 of the helicases, specifically to the Upf1-like family. However, the role of *IGHMBP2* in cells is unclear [139]. |

| Gene | Description |
|---|---|
| ***LRBA*** | Member of the WDL-BEACH-WD (WBW) gene family. Gene codes for LPS Responsive Beige-Like Anchor Protein (*LRBA*). LPS induces expression of *LRBA* by B cells and macrophages, and *LRBA* acts via protein kinase A to increase vesicle trafficking and subsequent release of immune effectors [80].<br>The disease common variable immunodeficiency-8 with autoimmunity is associated with defects in *LRBA* [80].<br>Pulmonary/respiratory symptoms are common with *LRBA* deficiency, including cough [140] infections, bronchiectasis, interstitial lung disease, thoracic lymphadenopathy, and clubbing [141]. |
| ***LTBP4*** | Gene encodes Latent Transforming Growth Factor-Beta Binding Protein 4, which binds and maintains TGFβ in a latent state [80]. *LTBP4* is critical for normal lung development, as *LTBP4* deficiency leads to abnormal alveolarization, angiogenesis and lung fibrosis [142], as well as abnormal elastogenesis and TGFβ signalling in the lung [143]. Patients with *LTBP4* mutations have very severe phenotypes affecting multiple organs, including the lungs. Respiratory symptoms include emphysema [144], severe respiratory distress and respiratory failure [145]. Similarly, *LTBP4* knockout mice develop severe pulmonary emphysema, cardiomyopathy, and colorectal cancer, with severe defects in extracellular matrix structure, particularly elastic fibres [146]. |
| ***NPNT*** | Gene encodes Nephronectin, a highly conserved extracellular matrix protein involved in integrin binding/signalling and cell adhesion, differentiation, spreading and survival [80] [147]. Nephronectin is involved in numerous processes, including pulmonary function [147], and several GWAS studies have shown NPNT to be associated with COPD, reduced lung spirometry measures (particularly $FEV_1$) and smoking behaviours [148] [149] [147] [150] [151]. Serum nephronectin levels have also been shown to be higher in patients with silicosis, a disease characterised by lung fibrosis, and nephronectin is believed to be involved fibrosis progression in this disease [152]. |
| ***SCARF2*** | Gene encodes Scavenger Receptor Class F Member 2, a protein involved in the binding and degradation of acetylated low-density lipoprotein [80]. No previous evidence to show a link between *SCARF2* and lung. Mutations in *SCARF2* are associated with van den Ende Gupta syndrome, a rare genetic disorder associated with craniofacial and skeletal abnormalities [153] [154] [155] [156] [157]. |
| ***SCMH1*** | Gene encodes Scm Polycomb Group Protein Homolog 1, a protein thought to be involved in negative regulation of transcription by modulating the activity of chromatin binding and histone binding [80]. No previous evidence to show a link between *SCMH1* and lung. *SCMH1* is thought to be involved in chromatin modification during spermatogenesis [158], and in epigenetic regulation to improve neuroprotection in ischemic tolerance [159]. |
| ***SMAD3*** | The *SMAD3* protein functions in the transforming growth factor-beta signalling pathway, and transmits signals from the cell surface to the nucleus, regulating gene activity and cell proliferation. It also functions as a tumour suppressor. Mutations in this gene are associated with aneurysms-osteoarthritis syndrome and Loeys-Dietz Syndrome 3, a connective tissue disorder [80]. The role of TGFB signalling in lung development, injury and repair is well established. Smad3 transduces TGF-beta signals from the cell membrane to the nucleus. Loss of Smad3 in mice greatly attenuated morphological fibrotic responses to bleomycin in the mouse lungs, suggesting that Smad3 is implicated in the pathogenesis of pulmonary fibrosis [160]. Smad3 deficinet mice do not have developmentally normal lungs but the phenotype is not neonatal lethal. These changes include disorganisation of elastin fibres and correct complete alveolar septation [161].Abnormal alveolarization in Smad3-deficient mice is followed by progressive emphysema-like alveolar wall destruction initiated by MMP9. MMP9 knockout with Smad3 knockout rescues the phenotype suggesting Smad3 is involved in MMP9 suppression via epigenetic regulation [161]. |
| ***SOS2*** | *SOS2* encodes a regulatory protein that is involved in the regulation of ras proteins. Mutations in the gene are associated with Noonan Syndrome-9 [80]. The SOS family of Ras-GEFs encompasses two highly homologous and widely expressed members, SOS1 and *SOS2*. *SOS1* is a RAS-specific guanine nucleotide exchanger factor (GEF) that facilitates the conversion of RAS from the inactive GDP-bound to the active GTP-bound form. *SOS1*-KO mouse mutants were embryonic lethal while *SOS2* -KO mice were viable led to initially viewing *SOS2* as the main Ras-GEF linking external stimuli to downstream RAS signalling. However, specific *SOS2* functions include a critical role in regulation of the RAS–PI3K/AKT signaling axis in keratinocytes and KRAS-driven tumor lines and control of epidermal stem cell homeostasis [162]. |

| Gene | Description |
|---|---|
| ***STIM1*** | Gene encodes stromal interaction molecule 1 (*STIM1*) protein. Plays a role in mediating store-operated Ca(2+) entry (SOCE). Acts as a Ca(2+) sensor in the endoplasmic reticulum via its EF-hand domain [80]. Upon depletion of intracellular Ca(2+) stores, *STIM1* translocates from the endoplasmic reticulum to the plasma membrane where it activates the Ca(2+) release-activated Ca(2+) (CRAC) channel subunit ORAI1 [163] [164].<br>Alterations in this region have been associated with the Beckwith-Wiedemann syndrome, Wilms tumor, rhabdomyosarcoma, adrenocrotical carcinoma, and lung, ovarian, and breast cancer. Mutations in this gene are associated with fatal classic Kaposi sarcoma, immunodeficiency due to defects in store-operated calcium entry (SOCE) in fibroblasts, ectodermal dysplasia and tubular aggregate myopathy [80].<br>*STIM1* is upregulated in the COPD lung tissue. *STIM1* expression is positively associated with ROS levels and negatively correlated with pulmonary function. *STIM1* expression is increased in the bronchoalveolar lavage fluid (BALF) macrophages of COPD patients and PMA-differentiated THP-1 macrophages stimulated by cigarette smoke extract (CSE). CSE-induced ROS production may increase *STIM1* expression in macrophages, further promoting IL-8 release by regulating Ca2+ entry. *STIM1* may participate in the pathogenesis of COPD [165].<br>*STIM1* mediates Influenza A virus (IAV)-induced inflammation of lung epithelial cells by regulating NLRP3 and inflammasome activation via targeting miR-223. Silencing *STIM1* alleviated IAV-induced inflammation injury of lung epithelial cells by inactivating NLRP3 and inflammasome via promoting miR-223 expression [166].<br>Expression of *STIM1* decreased with age. ASM senescence may enhance fibrosis in a feed forward loop promoting remodeling and altered calcium storage and buffering [167]. 17β-Estradiol (E2) non-genomically inhibits basal phosphorylation of *STIM1*, leading to reduced SOCE. This has an implication for chronic lung diseases [168]. *STIM1* controls T cell-mediated immune regulation and inflammation in chronic infection [169]. |
| ***TGFB2*** | TGFB encodes a secreted ligand of the TGF-beta (transforming growth factor-beta) superfamily of proteins. Ligands of this family bind various TGF-beta receptors leading to recruitment and activation of SMAD family transcription factors that regulate gene expression. Disruption of the TGF-beta/SMAD pathway has been implicated in a variety of human cancers. A chromosomal translocation that includes this gene is associated with Peters' anomaly, a congenital defect of the anterior chamber of the eye. Mutations in this gene may be associated with Loeys-Dietz syndrome. This gene encodes multiple isoforms that may undergo similar proteolytic processing [80].TGF-β activation has been implicated in the fibrosis of both IPF and airway remodelling [170]. Upregulation of TGF-β ligands is observed in major pulmonary diseases, including pulmonary fibrosis and emphysema [171]. TGF-β regulates multiple cellular processes such as growth suppression of epithelial cells, alveolar epithelial cell differentiation, fibroblast activation, and extracellular matrix organization. These effects are closely associated with tissue remodeling in pulmonary fibrosis and emphysema [171].*TGFB2* and *TGFB3* are independently involved in mouse fibrosis models in vivo and selective inhibition does not result in increased inflammation observed with total TGBF isoform inhibition [171].A region containing SNP rs1690789, identified within an association peak from a GWAS of emphysema patterns, contacts the *TGFB2* promoter in fibroblasts. Deletion of the region (100bp) containing this SNP results in decreased *TGFB2* expression in primary human lung fibroblasts [172] |
| ***TNS1*** | Encodes a protein that localises to focal adhesions (regions of the plasma membrane where the cell attaches to the extracellular matrix). Diseases associated with *TNS1* include Cowden Syndrome 1 and Proteus Syndrome. Related pathways for this gene include integrin mediated cell adhesion and integrin pathway [80]. *TNS1* expression is increased in fibroblastic foci from lungs with IPF.*TNS1* is upregulated by *TGFB*, which is dependent on *TGFB1* signalling. *TNS1* plays an essential role in TGF-β-induced myofibroblast differentiation and myofibroblast-mediated formation of extracellular fibronectin and collagen matrix [173]. Tensin1 is expressed in human airway smooth muscle, with higher expression in COPD tissue versus controls. Tensin1 is also expressed in human airway smooth muscle cells, with expression upregulated by *TGFB1*. Tensin1 and a-smooth muscle actin are strongly co-localised. Depletion of Tensin1 in human airway smooth muscle cells attenuated a-smooth muscle actin expression and contraction of collagen cells [174]. |
